# Multi-organ MRI digitizes biological aging clocks across proteomics, metabolomics, and genetics

**DOI:** 10.1101/2025.07.10.25331263

**Authors:** The MULTI Consortium, Huizi Cao, Zhiyuan Song, Michael R. Duggan, Guray Erus, Dhivya Srinivasan, Ye Ella Tian, Wenjia Bai, Michael S. Rafii, Paul Aisen, Daniel W. Belsky, Keenan A. Walker, Andrew Zalesky, Luigi Ferrucci, Christos Davatzikos, Junhao Wen

## Abstract

Leveraging clinical phenotypes^1,2^, neuroimaging^3^, proteomics^4^, metabolomics^5^, and epigenetics^6^, biological aging clocks across organ systems and tissues have advanced our understanding of human aging and disease. In this study, we expand this biological aging clock framework to multi-organ magnetic resonance imaging (MRI) by developing 7 organ-specific MRI-based biological age gaps (MRIBAGs), including the brain, heart, liver, adipose tissue, spleen, kidney, and pancreas. Leveraging imaging, genetic, proteomic, and metabolomic data from 313,645 individuals curated by the MULTI consortium, we link the 7 MRIBAGs to 2,923 plasma proteins, 327 metabolites, and 6,477,810 common genetic variants. These associations reveal organ-specific and cross-organ interconnection landscapes, identifying distinct molecular signatures related to organ aging. Genome-wide associations identify 53 MRIBAG-locus pairs (P<5×10^D8^). Genetic correlation and Mendelian randomization analyses further support organ-specific and cross-organ interconnections with 9 phenotype-based^1,2^, 11 proteome-based^7^, and 5 metabolome-based aging clocks^5^, as well as 525 disease endpoints. Through functional gene mapping and Bayesian colocalization analysis linking evidence from genetics, proteomics, and metabolomics, we prioritize 9 druggable genes as targets for future anti-aging treatments. Finally, we demonstrate the clinical relevance of the 7 MRIBAGs in predicting disease endpoints (e.g., diabetes mellitus), all-cause mortality, and capturing differential and heterogeneous cognitive decline trajectories over 240 weeks of treatment with the Alzheimer’s disease drug (Solanezumab). Sex differences are evident across multiple organ systems, manifesting at structural, molecular, and genetic levels. In summary, we developed 7 MRI-based aging clocks that enhance the existing multi-organ biological aging framework, offer multi-scale insights into aging biology, and demonstrate clinical potential to advance future aging research.

## Main

Magnetic resonance imaging (MRI^8^) provides a non-invasive window into structural and functional changes occurring with age, enabling the development of personalized disease biomarkers. Brain MRI-based aging clocks, often referred to as “brain age” and derived from artificial intelligence and machine learning (AI/ML) applied to MRI data, have been widely used as clinical biomarkers of neurological aging, cognitive decline, and neurodegenerative disease risk^9^. However, while brain imaging has been extensively utilized for aging research, no studies have systematically extended this concept to other organ systems. With the advent of large-scale, multi-organ MRI datasets like those from the UK Biobank (UKBB), we now have an unprecedented opportunity to conduct a more holistic, system-wide investigation of biological aging across multiple organs.

Beyond imaging-based aging clocks, researchers have also developed various omics-based aging clocks to capture different biological dimensions of aging. For example, plasma proteomics-based aging clocks^10,4^ have been introduced to enhance the granularity and coverage of multi-organ aging clocks. Plasma proteomics-based aging clocks capture the biological aging process by analyzing the abundance of proteins in circulation. Since proteins are the functional units of the cell, these clocks provide insights into the downstream effects of genetic regulation and environmental influences on aging. Metabolomics-based aging clocks^5,11^ capture biological aging by profiling small-molecule metabolites that represent the biochemical activity of cellular and systemic metabolism. Metabolomics examines the end products of metabolic pathways, making it highly dynamic and sensitive to external factors such as diet, microbiome composition, lifestyle, and environmental exposures. Finally, epigenetics-based aging clocks^6^ focus on the analysis of DNA methylation patterns, particularly at specific CpG sites across the genome, to estimate biological age. Integrating these aging clocks into the multi-organ^12,713^ and multi-omics concept is essential for gaining a comprehensive understanding of aging biology, age-related diseases, and longevity.

In our previous efforts, we developed 9 phenome-based biological age gaps (BAGs)^1,2^ (PhenoBAGs), 11 proteome-based BAGs (ProtBAGs^7^), and 5 metabolome-based BAGs (MetBAGs^5^). In this study, we built upon our previous work, which developed multi-modal brain MRI-based biological age gaps (MRIBAGs^3^), to derive 6 additional MRIBAGs from the heart, liver, spleen, adipose, kidney, and pancreas. This was accomplished using multi-organ MRI data from the UK Biobank (UKBB^14^), Baltimore Longitudinal Study of Aging (BLSA^15,16^), and Anti-Amyloid Treatment in Asymptomatic Alzheimer’s (A4^17,18^), integrated through the MULTI consortium, along with other omics data (e.g., genetics and proteomics) and GWAS summary statistics from FinnGen^19^ and the Psychiatric Genomics Consortium (PGC^20^) (**Method 1**). We first benchmarked the age prediction performance of the 6 additional MRIBAGs and identified potential limitations and challenges in developing these aging clocks (**Method 2**). Next, we associated the 7 MRIBAGs with 2923 plasma proteins for proteome-wide associations (ProWAS) and 327 metabolites for metabolome-wide associations (MetWAS; small-molecule metabolites or lipid complexes and subclasses) to depict their proteomic and metabolomic landscapes (**Method 3**). Through genome-wide association studies (GWAS) and subsequent analyses, including genetic correlation, polygenic risk scores (PRS), Bayesian colocalization, causal inference, and potential drug repurposing opportunities (**Method 4**), we comprehensively assessed the genetic architecture of the 7 MRIBAGs. Finally, we thoroughly assessed the clinical applicability and predictability of the MRIBAGs and their PRSs in several prediction tasks (**Method 5**). All results and pre-trained AI/ML models are publicly available at the MEDICINE portal: https://labs-laboratory.com/medicine/.

## Results

### Age prediction performance of the 7 MRIBAGs

The brain MRIBAG was initially developed in our previous study^3^ using 119 gray matter volumes, and we re-trained the brain MRIBAG here using a consistent nested cross-validation procedure, along with an independent test dataset (**Extended Data Fig. 1**). Specifically, to rigorously evaluate the performance (i.e., overfitting and generalizability) of biological age prediction models, we partitioned the healthy control (CN, without any pathologies) participants into the CN training/validation/test (3573<*N*<6327 as the sample sizes vary across organs) and independent test (ind. test; *N*=500) datasets (**Method 2c** and **Supplementary eTable 1**).

When fitting the organ-specific MRI features (**Fig. 1a**, **Method 2a** and **Supplementary eNote 1**), the two AI/ML models (Lasso regression and support vecor regressor) exhibited slight variability in performance, with no single model consistently outperforming the others (**Fig. 1b**). The optimal model for each MRIBAG was chosen based on greater generalizability, indicated by a smaller Cohen’s D, as marked by the # symbol, comparing the training/validation/test datasets to the independent test dataset. The selected optimal models for the 7 MRIBAGs demonstrated moderate Pearson’s *r* coefficients (ranging from 0.23 to 0.77) and the mean absolute errors (MAEs) of approximately 5 years in the independent test dataset (**Fig. 1c**) before applying the age bias correction^21,22^. Notably, the lower Pearson’s *r* coefficients observed for several abdominal MRIBAGs may be attributed to the limited number of imaging features available (e.g., only 3 features for the spleen MRIBAG); correspondently, the MRIBAGs derived from abdominal MRI are more affected by age bias (refer to Wen’s comment^7^ for detailed illustrations). **Supplementary eTable 2** presents detailed statistics for the age prediction tasks before the age bias correction, as well as results for elastic net and neural network (**Supplementary eFigure 1**). **Supplementary eNote 2** discusses whether an age prediction model (e.g., spleen MRIBAG) that predicts poorly for chronological age can still offer meaningful insights into biological aging beyond chronological age (**Supplementary eFigure 2**).

**Figure 1:**
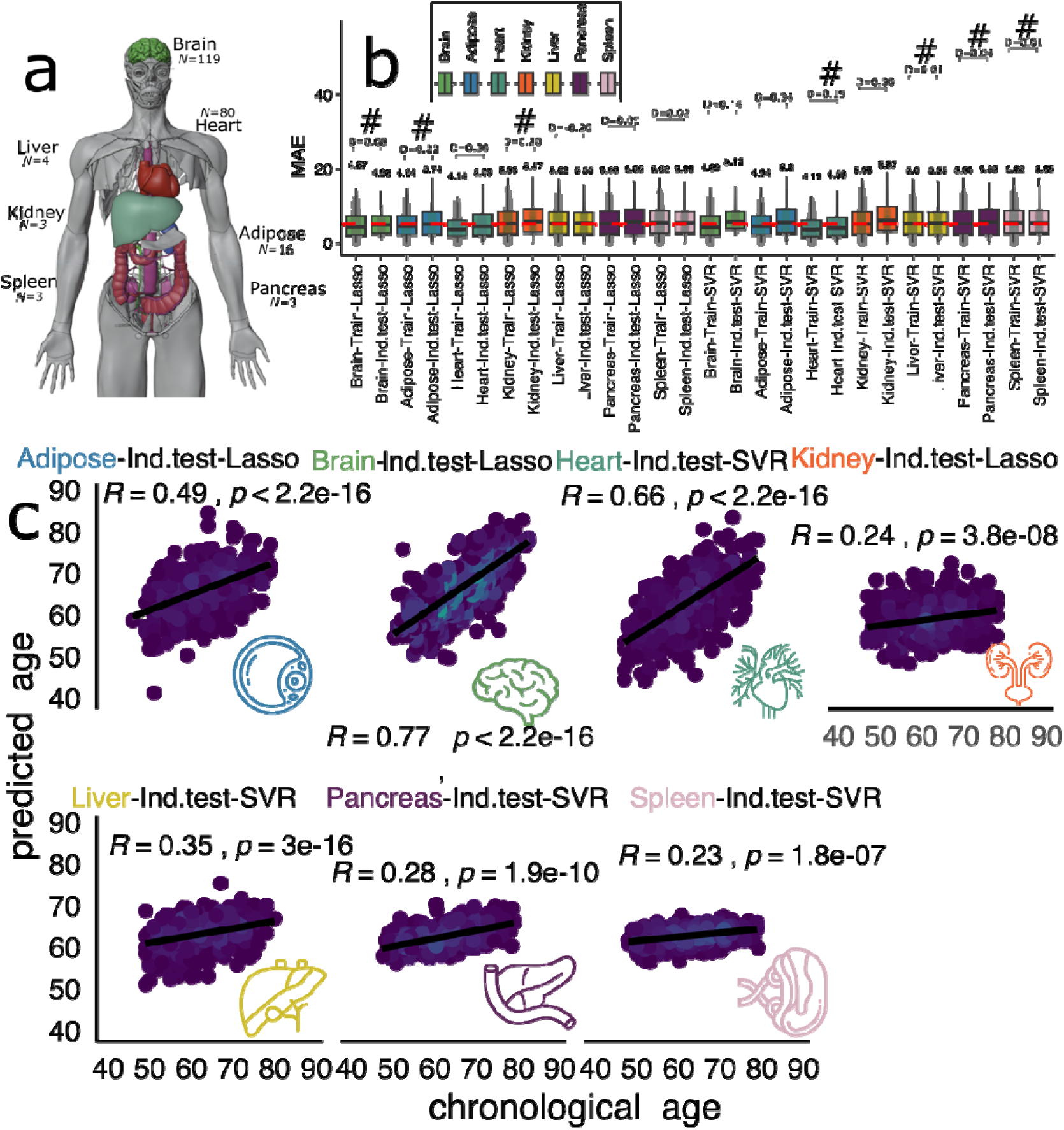
Model performance benchmarking for 7 multi-organ MRI-based biological aging clocks. **a**) The 7 MRIBAGs are derived from various imaging modalities available in the UK Biobank. The number (*N*) of imaging-derived phenotypes (IDPs) used to derive the 7 MRIBAGs is displayed. **b**) For the 7 MRIBAGs, the MAE of age prediction models using Lasso regression and linear support vector regressor is shown for both the training dataset (cross-validated training/validation/test) and the independent test dataset (Ind. test). Cohen’s D quantifies the effect size of the difference between these datasets, reflecting potential model generalizability, assuming comparable age and sex distributions. The optimal model (#) for each organ and tissue was selected based on the lower Cohen’s D value, and these models were used for all subsequent analyses. All MAEs are reported without age bias correction. **c**) Scatter plots display the optimal model for each organ/tissue in the Ind. test dataset, with Pearson’s *r* and P-values indicating the association between chronological age and predicted age. All results are shown before the correction for age bias was applied^22^.

Additionally, we found that high collinearity among imaging features from abdominal MRI led to poor generalizability in independent test datasets (**Supplementary eNote 3 and eFigure 3**). **Supplementary eNote 4 and eFigure 4-5** discuss potential domain shift while applying the pre-trained model of the brain MRIBAGs to external datasets, exemplified by the A4 study^17,18^. **Supplementary eNote 5 and eFigure 6** discuss sex differences in the 7 MRIBAGs. **Supplementary eNote 6 and eFigure 7-8** discuss the feature importance of deriving these MRIBAGs and their biological interpretation. For all subsequent analyses, we used the age bias-corrected^22^ MRIBAGs.

### The plasma proteomics and metabolomics landscape of the 7 MRIBAGs

In our ProWAS analyses (**Method 3a**), we identified 603 protein-MRIBAG significant associations (P-value<0.05/2923/7). Among these, the kidney MRIBAG exhibited the highest number of significant protein associations (*N*=301, e.g., NPDC1, IGFBP6, and TAFA5), followed by the spleen MRIBAG for 136 associations (e.g., VCAM1, PTPRH, and C1QA), liver MRIBAG for 62 associations (e.g., NCAN, SEZ6L, and LEP), adipose MRIBAG for 57 signals (e.g., GDF15, CHI3L1, and CA14), pancreas MRIBAG for 21 associations (e.g., PLA2G1B, CTRC, and CELA2A), brain MRIBAG for 16 associations (e.g., BCAN, NCAN, and GDF15), and heart MRIBAG for only REN (**Fig. 2a**).

**Figure 2:**
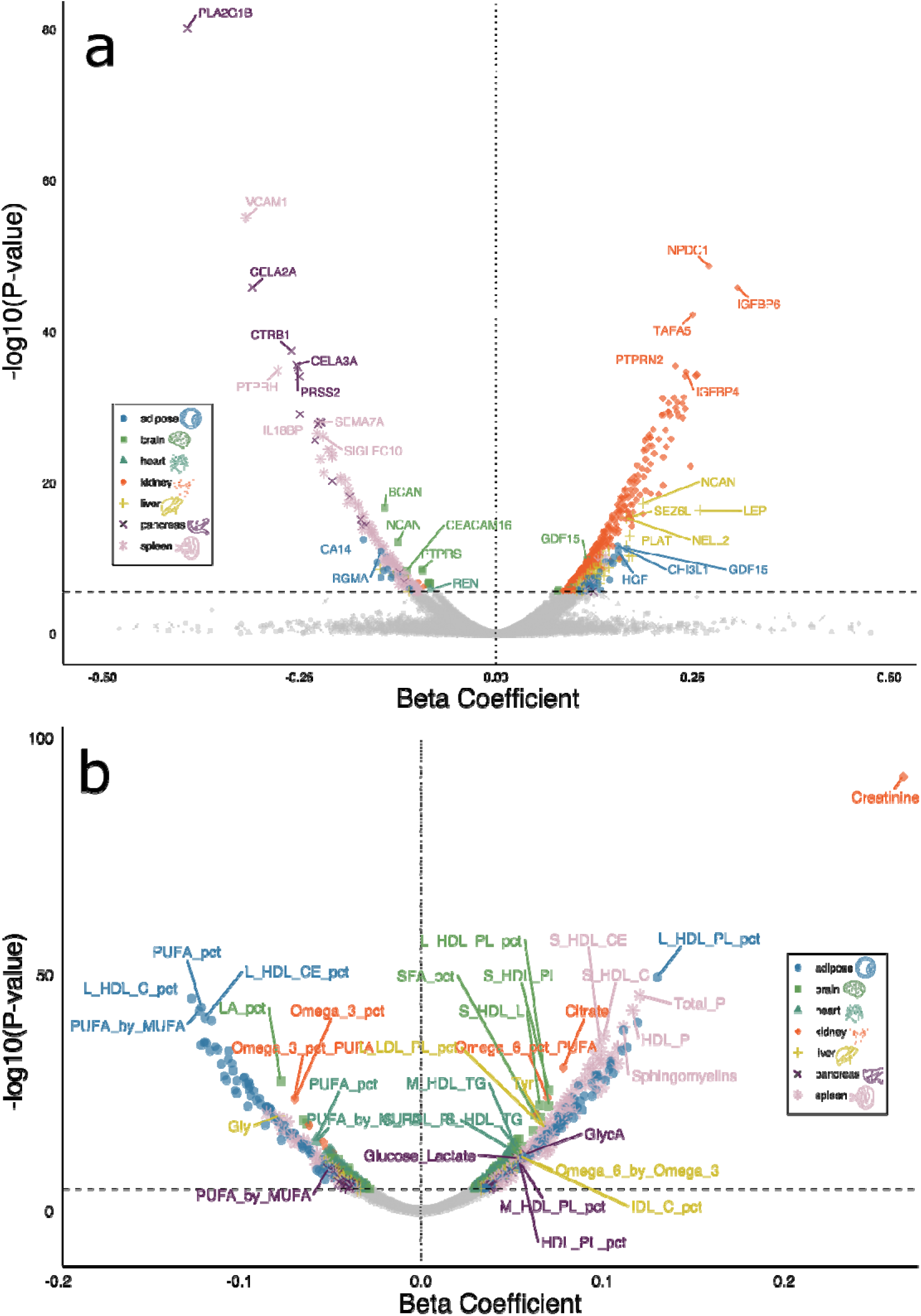
Proteome and metabolome-wide associations with the 7 MRIBAG. **a**) ProWAS between the 7 MRIBAGs and 2923 plasma proteins via a linear regression model, accounting for a full set of covariates (P-value<0.05/2923/7). An online interactive webpage is available at https://labs-laboratory.com/medicine/mribag_prowas.html to ease visualization. Plots of individual organs are shown in **Extended Data Fig. 2. b**) MetWAS between the 7 MRIBAGs and 327 plasma metabolites using a linear regression model while adjusting for a comprehensive set of covariates (P-value<0.05/327/7). Plots of individual organs are shown in **Extended Data Fig. 4**. An online interactive webpage is available at https://labs-laboratory.com/medicine/mribag_metwas.html to ease visualization. The colors and shapes of the icons indicate the organ systems involved in the associations. We used the standardized *β* value to represent the effect size.

VCAM1 was associated with the spleen MRIBAG (*β*=-0.89±0.04; P-value=7.05×10^−81^; *r*=-0.38); PLA2G1B was associated with the pancreas MRIBAG (*β*=-1.14±0.07; P-value=8.26×10^−56^; *r*=-0.30), adipose, and spleen MRIBAGs. The two proteins are organ-enriched in respective organ and tissue, as identified by the Human Protein Atlas (HPA: https://www.proteinatlas.org/), emphasizing their relevance to organ-specific aging processes, which has been widely used to define organ-enriched proteins in previous proteome-based aging clock papers^4,7^. For example, the PLA2G1B protein showed enriched expression in the pancreas in both RNA and protein data compared to other organs and tissues in HPA. The BCAN protein (nTPM=245.5 in the cerebral cortex) is a proteoglycan predominantly expressed in the central nervous system, particularly in the extracellular matrix of the brain. **Supplementary eFile 1a** presents detailed statistics. **Supplementary eNote 7, eFigure 9**, and **eFile 1b** explore the potential replication of UKBB Olink ProWAS signals in BLSA SomaScan data. **Extended Data Fig. 2-3** presents individual volcano plots and protein-set enrichment results via the STRING platform (v12.0^23^) for each MRIBAG (**Supplementary eFigure 10**). **Supplementary eNote 8 and eFigure 11** present the sex-specific ProWAS results.

In our MetWAS (**Method 3b**), we identified 758 metabolite-MRIBAG significant associations (P-value<0.05/327/7) between the 7 MRIBAGs and metabolites, showing relatively weak associations (Pearson’s *r*<0.3). The spleen MRIBAG showed the highest number of significant metabolite associations (*N*=199, e.g., Acetate, Acetoacetate, and Albumin), followed by the adipose MRIBAG (*N*=196, e.g., L_HDL_PL_pct, L_HDL_C_pct, and PUFA_pct), heart MRIBAG (*N*=139, e.g., PUFA_pct, M_HDL_TG, and HDL_TG), brain MRIBAG (*N*=97, e.g., LA_pct, Omega_6_pct, and ApoA1), pancreas MRIBAG (*N*=57, e.g., GlycA, Glucose_Lactate, and HDL_PL_pct), liver MRIBAG (*N*=51, e.g., Gly, Tyr, and L_LDL_PL_pct), and kidney MRIBAG (*N*=19, e.g., Creatinine, Citrate, and DHA) (**Fig. 2b**). Descriptions of these metabolites are provided in **Supplementary eTable 3**.

Creatinine was linked to the kidney (*β*=21.77±0.65; P-value=1.58×10^−258^; *r*=0.28) and adipose (*β*=-4.37±0.83; P-value=1.48×10^−7^; *r*=-0.18) MRIBAGs, which may suggest distinct roles of the kidney and adipose tissue in creatinine regulation. In addition to these small-molecule metabolites, other lipid complexes and subclasses also showed significant associations. Total concentration of phospholipids (Total_P) was negatively associated with liver (*β*=51.57±3.59; P-value=2.52×10^−46^; *r*=0.18). Phospholipids in small HDL (S_HDL_PL) were linked to the brain, heart, spleen, and adipose MRIBAGs. Similarly, cholesteryl esters in very large HDL (XL_HDL_CE), associated with the adipose and heart MRIBAGs. **Supplementary eFile 2** presents detailed statistics. **Extended Data Fig. 4** presents individual volcano plots for each MRIBAG. Metabolite-set enrichment analysis is presented in **Supplementary eNote 9 and Extended Data Fig. 5**. **Supplementary eFigure 12** shows the genetic correlations between the 7 MRIBAGs, 2923 proteins, and 327 metabolites. **Extended Data Fig. 6** presents example associations for the ProWAS and MetWAS. **Supplementary eNote 10 and eFigure 13** present the sex-specific MetWAS results.

### The genetic architecture of the 7 MRIBAGs

We conducted GWAS (**Method 4a**) for the 7 MRIBAGs (19,686<*N*<31,557 participants with European ancestries) and identified 53 (P-value<5×10^−8^) genomic locus-BAG pairs. We denoted the genomic loci using their top lead SNPs defined by FUMA (**Supplementary eNote 11**), considering linkage disequilibrium (LD); the genomic loci are presented in **Supplementary eTable 4**. We visually present the shared genomic loci annotated by cytogenetic regions based on the GRCh37 cytoband (**Fig. 3a**). **Supplementary eNote 12** and **Extended Data Fig. 6** detail the robustness of our GWASs. **Supplementary eNote 13 and eFigure 14** present the sex-specific GWAS analyses.

**Figure 3:**
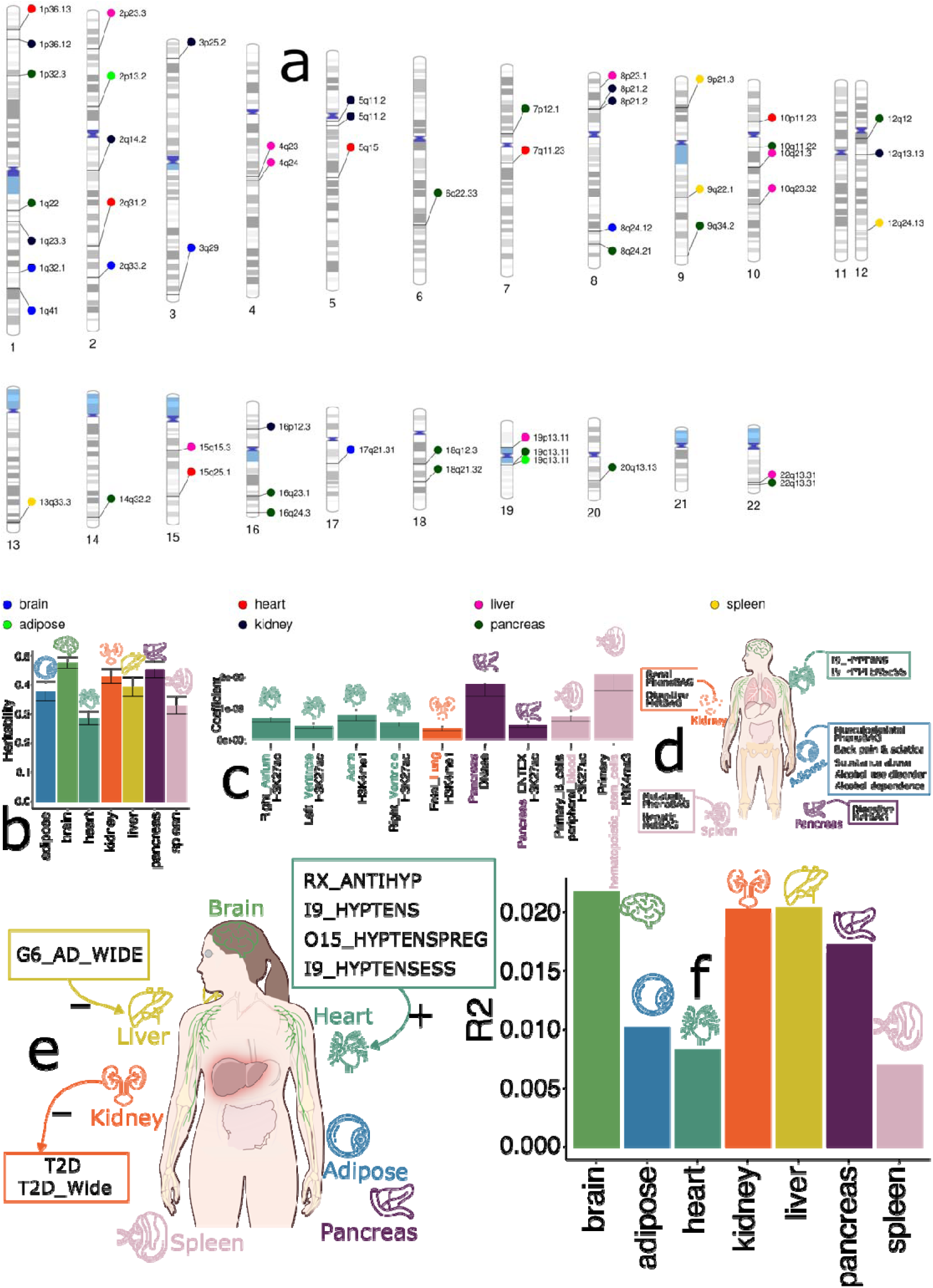
The genetics of the 7 multi-organ MRI-based biological aging clocks. **a**) Cytogenetic regions associated with the 7 MRIBAGs, with significant genomic loci identified using the genome-wide significance threshold (P-value<5×10^−8^). **b**) SNP-based heritability estimates of the 7 MRIBAGs. **c**) Partitioned heritability enrichment analysis using chromatin-specific multi-tissue chromatin data, multi-tissue gene expression profiles, and cell-type-specific datasets. Only significant results (P-value<0.05/697) are displayed. **d**) Genetic correlation estimates via LDSC between the 7 MRIBAGs and 8 phenotype-based aging clocks (PhenoBAGs, excluding the multi-modal brain PhenoBAG^1^), 11 proteome-based aging clocks (ProtBAGs^7^), and 5 metabolome-based aging clocks (MetBAGs^5^) (P-value<0.05/11), as well as 525 disease endpoints (DEs) from FinnGen and PGC (P-value<0.05/525). **e**) Potential causal relationships between the 7 MRIBAGs (i.e., number of instrumental variables > 7) and 525 DEs were examined through two networks: *BAG2DE*, where the 7 MRIBAGs serve as exposure variables and the 525 DEs as outcome variables, with P < 0.05/525, and *DE2BAG*, where the 214DEs serve as effective exposure variables (i.e., number of instrumental variables > 7), with P < 0.05/214. While multiple sensitivity checks were performed to evaluate potential violations of underlying assumptions, these findings should be interpreted with caution. The direction array represents the causal relationship from the exposure to the outcome variables, where “+” denotes an odds ratio (OR) > 1, and “–” indicates an OR < 1. **f**) The bar plot shows the incremental *R*^2^ (i.e., the *R*^2^ of the alternative model minus that of the null model) for the PRS of each MRIBAG. The PRS was calculated using the split2 target GWAS data, with split1 GWAS data serving as the training set for the PRScs model.

We estimated the SNP-based heritability 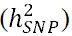 for the 7 MRIBAGs using the GCTA^24^ software (**Method 4b**), with values ranging from 0.29 to 0.47 (mean = 0.40) (**Fig. 3b**). **Supplementary eTable 5** presents detailed statistics. We validated the GWAS signals through partitioned heritability analyses (**Method 4c**) using the LDSC software, revealing strong organ-specific enrichment. Notably, the heart MRIBAG exhibited chromatin state-specific enrichment across four distinct heart tissues (**Fig. 3c**). For example, significant heritability enrichment was shown in the right atrium in the H3K4me3 region (P-value=1.00□×□10^−6^) and the left ventricle in the H3K27ac region (P-value=2.00□×□10^−5^) for the heart MRIBAG. **Supplementary eFile 3** presents detailed statistics.

We then assessed the genetic correlations (**Method 4d**) between the 7 MRIBAGs and 24 previously developed multi-organ, multi-omics aging clocks, including 8 PhenoBAGs^1^ (excluding the brain PhenoBAG based on multi-modal imaging), 11 ProtBAGs^7^, and 5 MetBAGs^5^. After Bonferroni correction, we found 7 within-organ and inter-organ significant genetic correlations (P-value<0.05/11), exemplified between the kidney MRIBAG and the renal PhenoBAG (*g_c_*=0.23±0.05; P-value<2.00×10^−5^), and between the spleen MRIBAG and the hepatic MetBAG (*g_c_*=0.23±0.08; P-value<7.00×10^−4^). We also performed genetic correlation between the 7 MRIBAGs and 525 disease endpoints (DE), and found 6 significant signals (P-value<0.05/525). For example, within-organ connections were demonstrated between the heart MRIBAG and two forms of hypertension, which is a major risk factor for cardiovascular diseases (e.g., FinnGen code: I9_HYPTENS: *g_c_*=0.28±0.06; P-value<1.00×10^−5^); cross-organ interactions were also identified between the adipose MRIBAG and substance abuse (*g_c_*=0.27±0.06; P-value<5.00×10^−5^) (**Fig. 3d**). **Supplementary eFigure 15** shows the phenotypic and genetic correlations within the 7 MRIBAGs. **Supplementary eFile 4** presents detailed statistics for all the genetic correlation analyses.

We conducted bi-directional Mendelian randomization analyses (**Method 4e**) between the 7 MRIBAGs and 525 DEs, ensuring at least 7 valid LD-considered SNPs as instrumental variables (IVs) after quality control. While the MRIBAGs were generally underpowered to meet this threshold, we identified 6 potential causal relationships. The within-organ interactions between the heart MRIBAG and hypertension were reaffirmed by a potential causal relationship from various forms of hypertension to the heart MRIBAG [e.g., I9_HYPTENS: P-value= 2.12×10^−7^; OR (95% CI)=1.13 (1.08, 1.18); number of IVs=110]. Additionally, we identified a potential causal relationship from Alzheimer’s disease (AD) to the liver MRIBAG [G6_AD_WIDE: P-value=3.02×10^−5^; OR (95% CI)=0.91 (0.88, 0.96); number of IVs=8], consistent with our previous findings linking AD to the hepatic PhenoBAG^1^. Finally, we found that the kidney MRIBAG was negatively linked to two forms of type II diabetes mellitus [e.g., T2D: P-value=8.40×10^−5^; OR (95% CI)=0.78 (0.70, 0.88); number of IVs=9] (**Fig. 3e**). **Supplementary eFile 5** presents detailed statistics for our causal analyses. **Supplementary eFigure 16** shows detailed sensitivity check analyses of potential causal signals for all 6 significant signals.

Using split-sample GWAS analyses, we also developed PRSs for the 7 MRIBAGs, with the first split serving as the training/source dataset and the second split as the test/target dataset (**Method 4f**). Overall, these PRSs exhibit limited predictive power. In the second split population, the brain PRS exhibited the highest incremental *R*^2^ (2.18%; P-value<1.00×10^D10^), while the spleen PRS accounted for an additional 0.70% (P-value<1.00×10^−10^) of the variance in the spleen MRIBAG (**Fig. 3f**). **Supplementary eTable 6** presents detailed statistics for our causal analyses.

### Genetic evidence to prioritize 9 druggable genes

We conducted multiple post-GWAS analyses (**Method 4g**) to identify potential druggable genes associated with the 7 MRIBAGs. Our overarching hypothesis posits that these imaging-derived aging clocks, as AI-driven endophenotypes^25^ (**Supplementary eFigure 17** for a conceptual visualization), reside along the causal pathway of human aging and disease, linking genetic predisposition to multi-omics data, such as gene expression, proteomics, and metabolomics, leading to disease outcomes and cognitive decline.

Initially, we mapped independent GWAS signals to genes using three approaches: *i*) positional mapping, *ii*) eQTL mapping, and *iii*) chromatin interaction mapping, identifying a total of 1164 unique genes. To further refine these genes, we prioritized genes that demonstrated both genetic evidence from eQTL mapping and chromatin interactions, resulting in a final set of 246 unique genes linked to the 7 MRIBAGs. For instance, *MAPT* was prioritized in the brain MRIBAG through both eQTL and chromatin interaction mapping and has previously been identified as a genetically supported druggable gene^26^. Subsequently, to further extend genetic evidence to other omics data types within the endophenotype hypothesis framework, we conducted Bayesian colocalization analysis on genomic loci associated with the 7 MRIBAGs and aging clocks derived from plasma proteomics^7^ (ProtBAGs) and metabolomics (MetBAGs^5^). The key hypothesis is that a causal variant positionally or functionally linked to a gene influencing both MRIBAGs and other omics-based aging clocks is more likely to play a causal role in human aging and disease (**Supplementary eFigure 17**), making the corresponding gene a strong candidate for drug targeting. Our colocalization analysis identified a total of 40 MRIBAG-MetBAG or MRIBAG-ProtBAG colocalization signals (PP.H4.ABF > 0.8) (**Fig. 4a**). By integrating these findings with the 246 prioritized genes, we identified 62 MRIBAG–gene pairs (comprising 62 unique genes) that share at least one causal SNP with either MetBAGs or ProtBAGs and exhibit significant signals in both eQTL and chromatin interaction–based gene mapping analyses (**Fig. 4b**). **Supplementary eTable 7 and eFile 6** present detailed results for our colocalization analysis and SNP-gene mapping results, respectively.

**Figure 4:**
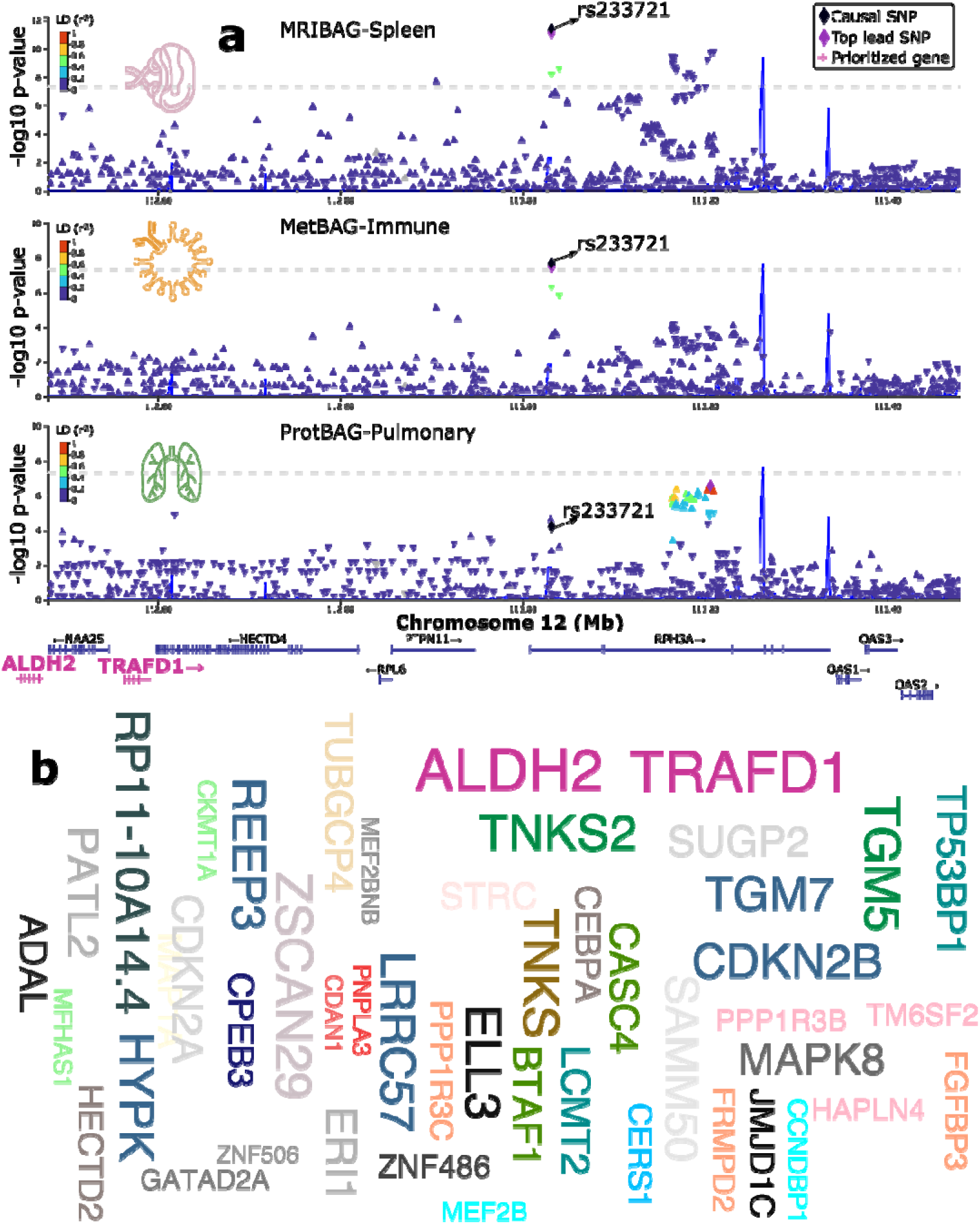
Multi-level genetic evidence prioritizes 62 genes as potential drug repurposing genes. **a**) To prioritize potential druggable genes, leveraging aging clocks derived from plasma proteomics (ProtBAGs) and metabolomics (MetBAGs), we filtered mapped genes to retain only those showing significant colocalization with any ProtBAGs or MetBAGs. As an example, we highlighted a specific genomic locus demonstrating colocalization between the spleen MRIBAG, the immune MetBAG, and the pulmonary ProtBAG. The shared causal variant (rs233721 on chromosome 12) exhibited a strong colocalization signal, with a posterior probability (PP.H4.ABF) of 0.99 between the spleen MRIBAG and the immune MetBAG, implying a single shared causal variant influencing both traits within this locus. This variant was mapped to the *TRAFD1* and *ALDH2* genes through both eQTL mapping (e.g., *ALDH2* in the aorta from GTEx) and chromatin interaction mapping (e.g., *TRAFD1* in the left ventricle and liver). **b**) By integrating this multi-level genetic evidence, we identified 62 unique genes associated with the liver, spleen, and pancreas MRIBAGs.

Among the 62 genes with multi-level genetic evidence supporting their potential causal role in human aging and disease, we further explored existing drug-gene interaction data to identify potential drug repurposing opportunities and evidence. To achieve this, we queried the DGIdb database (https://dgidb.org/; query date: 21 March 2025) and identified 9 genes (linked to the liver, spleen, and pancreas MRIBAGs) with known interactions involving 122 unique drugs (**Fig. 5a** and **Supplementary eFile 7**), either approved or unapproved drugs. Among the 122 unique drugs, many indications are manifested, including immunosuppressant agents (e.g., Everolimus; DrugBank ID: DB01590), antipsychotic agents (e.g., Loxapine for the management of the manifestations of psychotic disorders such as schizophrenia; DrugBank ID: DB00408), antidiabetic agents (e.g., Bromocriptine for treating Type 2 diabetes mellitus; DrugBank ID: DB01200), and antiparkinson agents (Lisuride for the management of Parkinson’s Disease; DrugBank ID: DB00589), as well as many antineoplastic agents.

**Figure 5:**
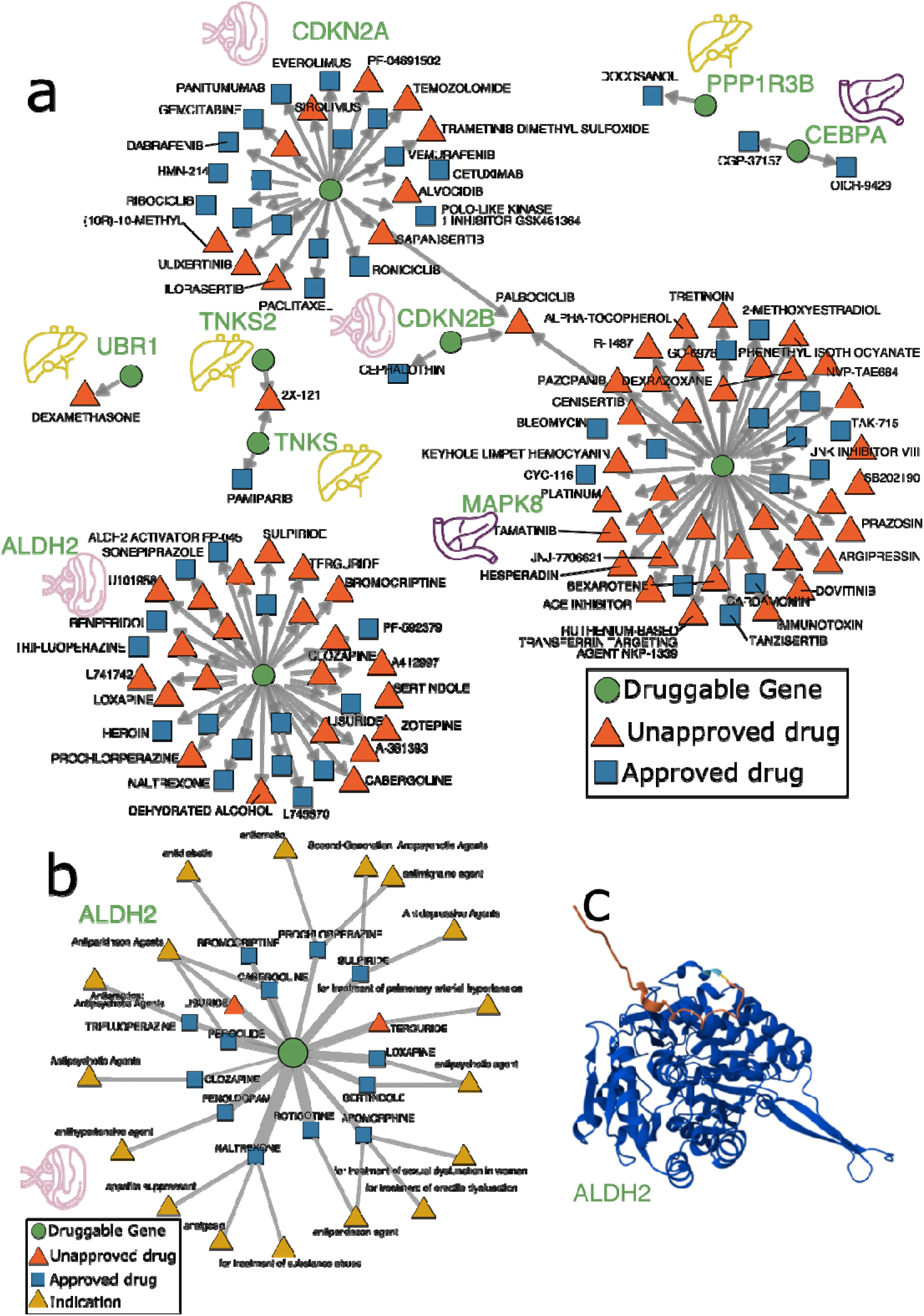
Drug-gene interactions for 9 potential drug repurposing genes. **a**) We queried the 62 prioritized MRIBAG-genes in the DGIdb platform (https://dgidb.org/) to identify drug-gene interactions and potential drug repurposing opportunities. We showed only the 9 MRIBAG-genes linked to existing drugs, whether approved or unapproved, using the curated data via DGIdb (**Supplementary eFile 7**). For visualization, we annotated representative drug names; the complete results can be found at an online interactive webpage at: https://labs-laboratory.com/medicine/mribag_gdi.html. **b**) Drug indications for existing drug-gene interactions related to the spleen MRIBAG-linked *ALDH2* gene. **c**) The 3D structure of the ALDH2 protein predicted by the AlphaFold model.

We highlighted the spleen MRIBAG-linked gene, *ALDH2*, as exemplified in our colocalizaiton analyses (**Fig. 4a**). Multi-level genetic evidence links the spleen BAG, *ALDH2*, and 15 drugs (Loxapine, Clozapine, Rotigotine, Sulpiride, Prochlorperazine, Terguride, Sertindole, Fenoldopam, Pergolide, Trifluoperazine, Naltrexone, Cabergoline, Apomorphine, Lisuride, Bromocriptine), with multiple clinical indications. These indications include antipsychotic (e.g., Clozapine for treating severe forms of psychiatric conditions like schizophrenia and for preventing suicide in patients who have this condition), antiparkinson (e.g., Rotigotine), antidepressive (e.g., Sulpiride, used to treat acute and chronic schizophrenia and major depressive disorder, but potentially toxic with a high dose, with side effects for depression), antimigraine (e.g., Prochlorperazine, used to treat psychotic symptoms such as schizophrenia and nonpsychotic anxiety, but has off-label uses including emergency use to treat migraine in adults and children), antihypertensive (e.g, Fenoldopam, a medication used to temporarily treat high blood pressure), antiemetics (e.g., Trifluoperazine, a medication used as an antipsychotic and an antiemetic), appetite suppressant, treatment of erectile dysfunction, antidiabetic, treatment of pulmonary arterial hypertension, treatment of substance abuse, analgesic, and treatment of sexual dysfunction in women (**Fig. 5b**). The 15 drugs linked to *ALDH2* encompass a broad range of therapeutic areas, many of which are relevant to aging-related diseases such as neurodegeneration, metabolic disorders, and immune system aging. As a central immune organ, the spleen’s aging may indicate immune dysfunction that contributes to neuroinflammation. *ALDH2* could influence both immune cell activity in the spleen and neuroinflammation in the brain, especially concerning drugs for neurodegenerative and psychiatric disorders. Drugs targeting *ALDH2* (or related pathways) for neurodegenerative diseases may also affect systemic aging, positioning them as potential candidates for modulating spleen or immune aging as well. **Supplementary eFigure 18** shows the full results of the drug implications on these priority genes.

### Potential clinical relevance of the 7 MRIBAGs

We highlight the clinical potential of the 7 MRIBAGs and their PRSs in predicting: *i*) the incidence of 53 individual disease conditions (with at least 50 patients) using ICD-10 codes, *ii*) all-cause mortality risk through survival analyses, and *iii*) differentiation in response to the anti-amyloid drug Solanezumab and cognitive decline trajectories in AD (**Method 5a-c**).

We found that the brain, adipose, pancreas MRIBAG, and the pancreas PRS could significantly (P-value<0.05/7) predict non-insulin-dependent diabetes mellitus (E119; 1.44<hazard ratio (HR)<1.70). The brain MRIBAG also predicted personal history of psychoactive substance abuse [Z864; HR=1.27 (1.11, 1.46); P-value=6.70×10^−4^], anxiety disorders [F419; HR=1.60 (1.20, 2.11); P-value=1.31×10^−3^], and gastrointestinal haemorrhage [K922; HR=1.46 (1.11, 1.91); P-value=6.63×10^−3^]. The Heart MRIBAG was associated with an increased risk of hypertension [I10; HR=1.16 (1.05, 1.28); P-value=4.80×10^−3^], reinforcing findings from genetic correlation and Mendelian randomization analyses (**Fig. 6a**). Crucially, in our survival analysis, non-case participants were selected by excluding all disease diagnoses. **Supplementary eFile 8** provides detailed statistics. In addition, certain diseases, such as AD, were excluded due to limited sample size, although they have been linked to aging clocks derived from other omics data, such as ProtBAG^4^.

**Figure 6:**
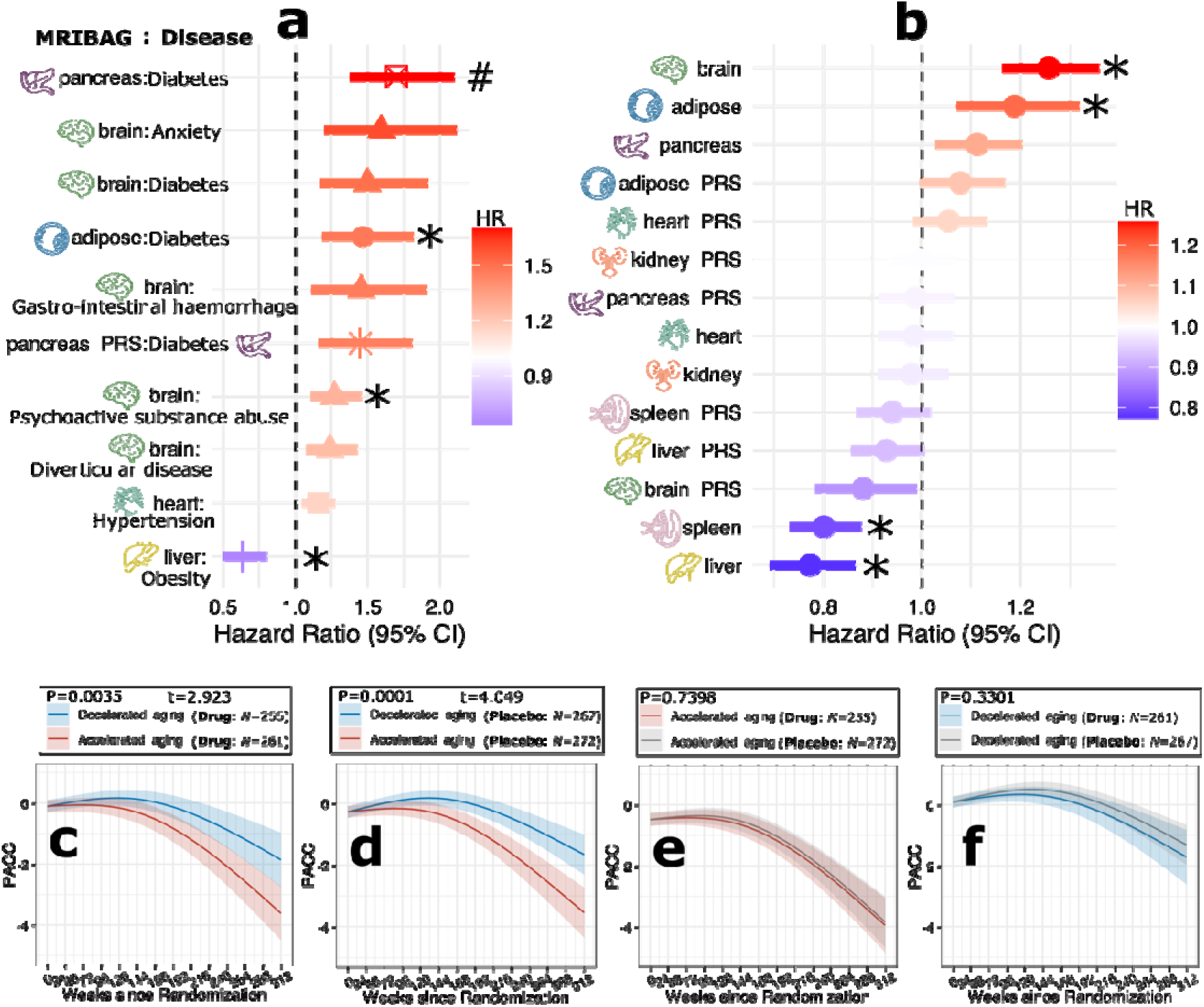
Clinical utility of the 7 MRIBAGs and their PRS. **a**) The 7 MRIBAGs and their PRS show significant associations with the incidence of single disease entities (e.g., I10 for hypertension). We only included disease entities > 50 cases (P-value<0.05/7 corrected for the number of organ systems), and non-cases are participants that do not show any disease diagnosis (disease-free) after enrolling in UKBB, so that all non-case populations remained the same reference population across all tasks. Age and sex were included as covariates in the Cox proportional hazard model. An asterisk (*) denotes results that remain significant after applying the Bonferroni correction for multiple comparisons (P-value < 0.05/53, accounting for the number of DEs), while a hash symbol (#) indicates significance under a more stringent threshold (P-value < 0.05/53/7). **b**) The brain, adipose, spleen, and liver MRIBAGs showed significant associations with all-cause mortality risk. To further validate the negative associations of the spleen and liver MRIBAGs with mortality, we conducted a disease-free analysis, including only participants without any prior disease diagnoses, to rule out potential confounders. **c**) At week 240, Preclinical Alzheimer Cognitive Composite (PACC) scores differed between the accelerated and decelerated aging groups – stratified by brain MRIBAG expression – among participants originally assigned to the drug group in the Solanezumab trial. **d**) At week 240, PACC scores differed between the accelerated and decelerated aging groups among participants originally assigned to the placebo group in the Solanezumab trial. **e**) At week 240 of the Solanezumab trial, PACC scores in the accelerated aging group showed no difference between participants receiving the drug and those receiving placebo. **f**) At week 240 of the Solanezumab trial, PACC scores in the decelerated aging group showed no difference between participants receiving the drug and those receiving placebo.

We further found that several MRIBAGs significantly (P-value<0.05/7) predicted all-cause mortality (**Fig. 6b**). Among these, the brain and adipose MRIBAGs were identified as risk biomarkers (HR>1) for all-cause mortality, while the liver and spleen MRIBAGs (HR<1) appeared to have a protective effect. We conducted additional sensitivity analyses by excluding participants with any disease diagnoses to assess disease-free survival. Despite the reduced sample size, the protective effect remained evident. For instance, the spleen MRIBAG retained nominal significance under this approach [P-value = 0.04; HR = 0.62 (0.40, 0.99)]. Detailed statistics are provided in **Supplementary eTable 8a-b**. **Supplementary eNote 14 and eFigure 19–20** presents a detailed discussion on additional sensitivity check results.

Finally, we assessed the use of the brain MRIBAG for stratifying clinical trial participants into subgroups to: *i*) demonstrate differential cognitive decline trajectories across subgroups, and *ii*) provide insights for future clinical trial design, using data from the A4. As shown in **Fig. 6c-d**, participants exhibiting decelerated aging signatures (i.e., decelerated aging group: MRIBAG below the median) showed a higher Preclinical Alzheimer Cognitive Composite (PACC) score than those within the accelerated aging group both in the drug group (P-value=0.003 and t-value=2.923) and the placebo group (P-value=0.001 and t-value=4.049). A permutation test showed that the cognitive differences between accelerated and decelerated agers, quantified by t-values, were significantly more pronounced in the placebo group compared to the drug group (one-sided P-value=0.03 based on 1000 permutations). A one-sided P-value was used, as our hypothesis was directional; we specifically aimed to test whether the placebo group exhibited stronger cognitive separation between aging subgroups, consistent with the observed pattern in cognitive trajectories (**Fig. 6c–d**). Furthermore, neither the decelerated nor the accelerated aging group exhibited significant drug effects (**Fig. 6e–f**) in preventing cognitive decline.

## Discussion

This study introduces 7 MRI-based aging clocks by leveraging large-scale multi-organ and multi-omics data from 313,645 participants. We evaluated model performance and underscored the need for advanced feature engineering in abdominal MRI data to enhance predictive accuracy. Additionally, we systematically linked the 7 MRIBAGs to 2,923 plasma proteins, 327 metabolites, and 6,477,810 common genetic variants. Notably, we prioritized 9 druggable genes with multi-level genetic evidence across different biological layers. The MRIBAGs are significantly associated with individual disease endpoints and all-cause mortality while also demonstrating differential cognitive decline trajectories in the Solanezumab AD drug trial, demonstrating future potential for AI-guided drug development. Collectively, this work expands existing biological aging clocks by introducing 7 MRIBAGs and highlights their clinical potential for future research.

### In-depth feature engineering is needed for abdominal MRI

Overall, the age prediction models achieved an MAE of approximately 5 years across all 7 organs and tissues. However, Pearson’s correlation metrics revealed limitations in the abdominal MRI features (*r*=0.23 for the spleen MRIBAG), both in terms of feature quantity and the presence of multicollinearity, which hindered model performance and generalizability to unseen data. For instance, the kidney MRIBAGs incorporated only three imaging features, as several features, including left kidney parenchyma (field ID=21161), right kidney volume (field ID=21082), and total kidney parenchyma (field ID=21160), exhibited high correlations (Pearson’s *r*>0.9) with other variables, leading to severe overfitting. Compared to the brain imaging community^27^, the development of imaging protocols and standardized imaging-derived phenotypes (IDPs) for other organs may be relatively underdeveloped. This disparity is reflected in large-scale biobanks such as UKBB, where the majority of well-characterized IDPs are derived from brain MRI (*N*>4000) and heart MRI, while imaging-derived features, returned and made available to the community, for other organs are limited in both number and diversity. Several factors contribute to this imbalance, including the historical focus of neuroimaging research, the availability of standardized brain imaging pipelines^28^, and the widespread adoption of advanced computational tools tailored for brain MRI analysis. Imaging data from other organs should also be incorporated into this framework. For instance, integrating retina imaging^29^ with brain MRI, genetic data, and systemic biomarkers could reveal novel mechanisms underlying aging and disease. As retina imaging becomes more widely available in biobanks, the development of standardized and automated pipelines will be crucial for enabling large-scale analysis. Additional methodological improvements to improve model performance for abdominal MRIBAG are discussed in **Supplementary eNote 15**.

### Proteomic and metabolomic associations across and within organs and tissues

The ProWAS results reveal a complex landscape of plasma protein associations with MRI-based aging clocks across 7 organs. These findings highlight both organ-specific aging mechanisms and systemic inter-organ communication, providing a foundation for understanding multi-tissue aging dynamics. For example, NCAN (neurocan; P-value=6.76×10^−13^, *β*=-0.28), BCAN (Brevican; P-value=1.91×10^−17^, *β*=-0.38), and MOG (myelin oligodendrocyte glycoprotein; P-value=2.45×10^−7^, *β*=-0.24) are all brain tissue-enriched proteins (i.e., at least four-fold higher mRNA level in brain compared to any other tissues: https://www.proteinatlas.org/humanproteome/brain/human+brain). They may imply that extracellular matrix and myelin-related pathways (e.g., neuronal adhesion and neurite growth during development) play key roles in maintaining neural structures during brain aging. Such organ-specificity of the brain MRIBAG was also reinforced by two previous studies that used plasma proteomics data to develop brain tissue-enriched aging clocks (i.e., the brain ProtBAG) using both SomaScan and Olink platforms. Oh et al.^4^ identified three brain tissue-enriched proteins based solely on GTEx data to construct the brain ProtBAG, whereas Wen et al.^7^ leveraged the HPA platform, which integrates more extensive curation than GTEx at both RNA and protein levels, to define brain tissue-specific proteins. Other proteins include CHI3L1 (P-value=2.04×10^−6^, *β*=0.10), a well-established neuroinflammatory marker elevated in AD and multiple sclerosis^30^, and GDF15 (P-value=1.55×10^−10^, *β*=0.25), a stress-induced cytokine linked to cognitive decline, AD, and aging^31,32^.

The significant metabolites identified in the MetWAS analysis further reinforce both cross-organ and within-organ associations. Several metabolites exhibited consistent links with MRI-BAGs across multiple organs, suggesting shared metabolic pathways contributing to aging in different tissues. HDL-related metabolites^33^ (e.g., HDL cholesterol esters, phospholipids) were significantly associated with BAGs in the brain, adipose tissue, heart, and spleen, while VLDL and LDL metabolites^34^ (e.g., VLDL_TG, LDL_CE) were predominant in the liver, pancreas, and spleen, indicating lipid metabolism disruptions in these organs. Overall, metabolites such as GlycA, HDL subclasses, and PUFA ratios may serve as broad aging biomarkers across tissues, whereas others (e.g., creatinine, sphingomyelins) appear to be more organ-specific. In summary, these findings emphasize that organ aging follows distinct pathways, such as myelination in the brain and fibrosis in the liver and kidneys. While certain proteins or metabolites, like GDF15 and FABP4, serve as systemic aging markers, others, such as NCAN and IGFBP6, exhibit tissue-specific roles. Shared underlying biological pathways may include inflammation^35^, extracellular matrix dysregulation^36^, and metabolic dysfunction^37^.

### Genetic evidence to support cross-organ and within-organ interactions of the 7 organs and tissues

The results of our genetic analyses provide compelling evidence for both cross-organ and within-organ genetic interactions, illustrating the intricate relationship^38^ between the 7 organs and tissues. These findings reveal that while the MRIBAGs share certain genetic underpinnings, each organ also demonstrates a distinct genetic architecture shaped by its unique biological processes. Our partitioned heritability analysis further supports this notion by identifying strong organ-specific genetic enrichments, with the heart MRIBAG, for instance, showing chromatin state-specific enrichment across distinct heart tissues, such as the right atrium and left ventricle. These enrichments emphasize the tissue-specific regulation of genetic risk factors and suggest that each organ’s genetic architecture is not merely a reflection of shared genetic variation but is also shaped by local, organ-specific processes that impact disease susceptibility and biological functions^39^.

Moreover, genetic correlations between the MRIBAGs and other multi-omics aging clocks, as well as the MRIBAGs’ associations with various disease endpoints, strengthen the notion of both shared and organ-specific genetic influences. For example, the correlation between the kidney MRIBAG and the renal PhenoBAG highlights how kidney-specific genetic factors can interact with broader multi-organ aging mechanisms at multiple omics levels^40^.

Similarly, the strong genetic association of heart MRIBAG with hypertension, a risk factor for cardiovascular disease, also suggests a clear example of shared genetic influences. Importantly, our causal analysis further confirmed this association. These interactions between organ-specific genetic factors and broader, more systemic disease risk factors^41^ further emphasize the complexity of genetic regulation across different tissues. Importantly, while our analyses show that these organs share common genetic influences that link them to broader disease pathways, they also exhibit unique, organ-specific contributions that reflect their individual physiological roles and functions. Together, these findings provide strong support for the hypothesis that the MRIBAGs share genetic influences across organs, while also demonstrating distinct genetic signatures that contribute to the specificity of each organ’s role in aging and disease. Another key question is whether MRIBAGs are linked to molecular-level aging clocks, such as ProtBAGs, in the context of human diseases. Using structural equation modeling, we demonstrated a proof-of-concept suggesting that brain MRIBAG may mediate the pathway from ProtBAG to 2 AI– and MRI-defined subtypes of late-life depression^42^, while the reverse direction, with ProtBAG as the mediator, was not supported (**Supplementary eFigure 21 and eTable 9**).

### Potential drug repurposing opportunities for anti-aging and age-related diseases

The genetic, metabolomic, and proteomic evidence supporting cross-organ and within-organ specificity in the 7 MRIBAGs further strengthens our findings in the potential drug repurposing opportunities. These existing drugs cover a wide range of therapeutic indications, including immunosuppressants, antipsychotics, antidiabetic agents, antiparkinsonian drugs, and various anticancer agents, indicating their potential utility across multiple organs and conditions. The broad spectrum of drug indications linked to these genes highlights the opportunity to repurpose existing therapies for targeting aging and age-related diseases in different organs, providing a promising direction for future therapeutic advancements.

For example, Everolimus (spleen MRIBAG mapped gene: *CDKN2A*), a rapamycin analog, has shown promise in extending lifespan and improving lifespan in humans^43,44^, particularly in delaying age-related diseases and improving immune function. It is known for its ability to suppress immune system activity, which can reduce chronic inflammation, a hallmark of aging. Studies have suggested that chronic low-grade inflammation, often referred to as “inflammaging^45^,” contributes to various age-related diseases, such as cardiovascular disease^46^ and AD^47^. Immunosuppressants may mitigate this process by modulating immune responses and promoting tissue repair and regeneration. Notably, the mTOR pathway^43^, targeted by drugs like Everolimus, has been shown to regulate cellular processes like growth, survival, and metabolism, and its inhibition has been linked to lifespan extension in model organisms, including yeast, worms, and mice^48^. Although Everolimus is not typically a first-line agent for treating splenomegaly, its effects on modulating immune responses and inhibiting cell proliferation suggest its potential use in treating spleen-related diseases, particularly those associated with hematological malignancies.

### The 7 MRIBAGs show promise as aging biomarkers in clinical applications

The 7 MRIBAGs show potential as aging biomarkers in clinical applications, demonstrating their association with disease incidence, all-cause mortality, and differential cognitive decline trajectories in AD patients taking Solanezumab. Our analyses revealed that MRIBAGs were associated with conditions such as non-insulin-dependent diabetes mellitus, anxiety disorders, hypertension, and gastrointestinal hemorrhages, with some MRIBAGs showing increased risks and others, like the liver and spleen MRIBAGs, indicating a protective effect (i.e., with a higher BAG score).

The counterintuitive finding that higher values of the liver and spleen MRIBAGs are associated with a lower risk of mortality could reflect their role in preserving organ function and overall physiological balance as people age. Typically, one might expect a lower risk of mortality to be associated with younger biological age or lower MRIBAG values. However, in this case, the higher MRIBAG values likely reflect healthier, more resilient organs that maintain their metabolic and immune functions more effectively over time. Specifically, a higher liver MRIBAG may signal better metabolic health, efficient detoxification, and regulation of systemic inflammation, which are crucial for maintaining overall homeostasis. Similarly, a higher spleen MRIBAG could indicate better immune function, including more effective immune surveillance and response to infections and other stressors. In the context of aging, these organs might act as “reserve systems^49^”, buffering the body against environmental insults and chronic diseases, which would reduce the risk of mortality. Essentially, higher MRIBAG values may reflect an enhanced capacity of the liver and spleen to perform their vital functions, suggesting that their efficient operation is protective in terms of longevity and lifespan. This may explain why individuals with higher liver and spleen MRIBAG values experience lower mortality risk despite the seemingly counterintuitive association. Additional sensitivity check approaches are presented in **Supplementary eNote 14**.

These findings suggest that MRIBAGs capture organ-specific factors influencing aging and disease risk, with potential applications in clinical decision-making. Their ability to predict mortality risk underscores their value in assessing biological aging. MRIBAG may also help stratify patients in aging-related drug trials, although no significant benefit was found between the drug and placebo groups after stratifying participants into aging subgroups (**Fig. 6e-f**).

Overall, these results support the integration of the 7 MRIBAGs into clinical practice for improved risk prediction and personalized treatment strategies in aging-related diseases.

### Limitations

This study has several limitations. First, the analysis is based on cross-sectional data, which limits the ability to infer and understand the temporal progression of aging and disease. Longitudinal studies will be crucial to better investigate the dynamic trajectories of MRIBAGs over time and their temporal relationships with health outcomes. Such studies could help disentangle causal pathways, assess the stability or reversibility of organ-specific aging patterns, and provide deeper insight into whether changes in MRIBAGs precede or follow the onset of age-related diseases. Second, while the analysis primarily focused on genetic and imaging data, environmental factors such as lifestyle, diet, and pollution were not considered, yet these may play significant roles in aging and disease^50^. Incorporating these factors in future studies would provide a more comprehensive understanding. Third, the imaging features for the 5 abdominal organs and tissues were relatively underdeveloped, underscoring the need for future efforts to implement more advanced feature engineering techniques and methodologies, such as deep neural networks. Furthermore, the genetic analysis was focused primarily on individuals of European ancestry, and future studies should extend these findings to underrepresented populations to enhance their generalizability. Relatedly, the relatively low predictive power of the MRIBAG-derived PRSs, which explained less than 2% of the variance, indicates that these genetic scores are not yet clinically actionable. Future refinement through larger, multiethnic GWAS and the inclusion of rare variants may improve their utility and biological interpretability. In addition, a key limitation of our study is the lack of an external biobank with similarly comprehensive imaging and multi-omics data to independently replicate and validate our findings. The current results derived from the UKBB study would benefit from confirmation in other large-scale cohorts to ensure their generalizability across populations and study designs. Finally, as a future avenue, studies incorporating more AD-oriented and AI-derived imaging biomarkers (e.g., subtypes informed by tau or amyloid PET data) may enhance statistical power and clinical relevance to benefit future clinical trials.

## Methods

### Method 1: The MULTI consortium

The MULTI consortium is an ongoing initiative to integrate and consolidate multi-organ data and multi-omics data, including imaging, genetics, and proteomics. Building on existing consortia and studies, such as those listed below, MULTI aims to curate and harmonize the data to model human aging and disease across the lifespan. In this study, in total, individual-level data for 313,645 participants were analyzed, including multi-organ MRI data across 7 organs and tissues (Category 100003), genetics, plasma proteomics data, and metabolomics data. GWAS summary statistics from FinnGen and PGC were downloaded and harmonized for our post-GWAS analyses. Refer to **Supplementary eTable 1** for comprehensive information, including the complete list of data analyzed and their respective sample characteristics. Participants provided written informed consent to the corresponding studies. The MULTI consortium is approved by the Institutional Review Board at Columbia University (AAAV6751).

### UK Biobank

UKBB^14^ is a population-based research initiative comprising around 500,000 individuals from the United Kingdom between 2006 and 2010. Ethical approval for the UKBB study has been secured, and information about the ethics committee can be found here: https://www.ukbiobank.ac.uk/learn-more-about-uk-biobank/governance/ethics-advisory-committee.

This study collectively analyzed multi-organ MRI data from 7 organ systems and tissues, including the brain, heart, liver, pancreas, spleen, adipose, and kidney. For the genetic data, we conducted a quality check on the imputed genotype data^14^ for the entire UKBB population (approximately ∼500k individuals). Subsequently, we merged the processed data with the organ-specific populations for all genetic analyses. Finally, we also included Olink plasma proteomics data released by the UK Biobank Pharma Proteomics Project (UKB-PPP) and metabolomics data from UKBB, which were detailed in the following sections.

### Baltimore Longitudinal Study of Aging

The main goal of the BLSA is to understand the normal aging process. Tracking physiological and cognitive changes over time aims to identify risk factors for age-related diseases, study patterns of decline, and discover predictors of healthy aging. BLSA^15,16^ brain MRI, and SomaScan proteomics data (https://www.blsa.nih.gov/) were used to compare and replicate the ProWAS results from the UKBB Olink data. After quality checks in this study, we included 1114 brain MRI scans at baseline and measurements of 7268 plasma proteins from 909 participants quantified with the SomaScan v4.1 platform. Age (years), sex (male/female), race (white/non-white), and education level (years) were defined based on participant self-reports.

### Anti-Amyloid Treatment in Asymptomatic Alzheimer’s

The A4 study^17,18^ (https://atri.usc.edu/study/a4-study/) is a clinical trial study to test a specific way to prevent memory loss associated with AD (clinical trial number: NCT02008357). The A4 study focused on symptom-free adults at higher risk for AD to assess whether an investigational drug (i.e., Solanezumab) could slow memory decline linked to amyloid plaques in the brain. It also examined whether Solanezumab could delay AD progression, measuring related brain changes using imaging, blood biomarkers, and baseline PET scans to assess amyloid levels. This study analyzed 1,055 participants at baseline with brain MRI scans to derive the brain MRIBAG. Longitudinal outcomes from the clinical trial, with the Preclinical Alzheimer’s Cognitive Composite (PACC) score as the primary measure over 312 weeks, were included. The PACC scores between groups were evaluated at week 240.

### FinnGen

The FinnGen^19^ study is a large-scale genomics initiative that has analyzed over 500,000 Finnish biobank samples and correlated genetic variation with health data to understand disease mechanisms and predispositions. The project is a collaboration between research organizations and biobanks within Finland and international industry partners. For the benefit of research, FinnGen generously made their GWAS findings accessible to the wider scientific community (https://www.finngen.fi/en/access_results). This research utilized the publicly released GWAS summary statistics (version R9), which became available on May 11, 2022, after harmonization by the consortium. No individual data were used in the current study.

FinnGen published the R9 version of GWAS summary statistics via REGENIE software (v2.2.4)^51^, covering 2272 DEs, including 2269 binary traits and 3 quantitative traits. The GWAS model encompassed covariates like age, sex, the initial 10 genetic principal components, and the genotyping batch. Genotype imputation was referenced on the population-specific SISu v4.0 panel. We included GWAS summary statistics for 521 FinnGen DEs in our analyses.

### Psychiatric Genomics Consortium

PGC^20^ is an international collaboration of researchers studying the genetic basis of psychiatric disorders. PGC aims to identify and understand the genetic factors contributing to various psychiatric disorders such as schizophrenia, bipolar disorder, major depressive disorder, and others. The GWAS summary statistics were acquired from the PGC website (https://pgc.unc.edu/for-researchers/download-results/), underwent quality checks, and were harmonized to ensure seamless integration into our analysis. No individual data were used from PGC. Each study detailed its specific GWAS models and methodologies, and the consortium consolidated the release of GWAS summary statistics derived from individual studies. In the current study, we included summary data for 4 brain diseases for which allele frequencies were present.

### Method 2: Multi-organ MRI analyses to derive the 7 MRIBAGs

In our previous analysis of raw brain MRI data from the UKBB and other studies included in this work, we extracted 119 gray matter (GM) regions of interest (ROIs) from T1-weighted MRI to generate brain MRIBAGs through the iSTAGING consortium^52^. For the heart MRIBAG, we used 80 heart MRI traits from Bai et al^53^ and used these imaging features in a previous study^13^. To develop the remaining 5 MRIBAGs based on abdominal MRI (Category ID=105^54,55,56,57,58,59^), we incorporated 16 imaging features for adipose MRIBAG, 4 for liver MRIBAG, 3 for kidney MRIBAG, 3 for spleen MRIBAG, and 3 for pancreas MRIBAG. Notably, in the development of adipose and kidney MRIBAGs, we observed a high degree of collinearity among certain imaging features, which led to overfitting. To mitigate this issue, we removed highly correlated features to improve model robustness.

**(a)** **Additional quality checks for the abdominal MRI:** The abdominal MRI data underwent initial quality control procedures prior to public release, including the removal of low-quality images and biologically implausible values. As noted on the UKBB website (https://biobank.ndph.ox.ac.uk/ukb/label.cgi?id=158), different pipelines were used to generate imaging features from abdominal MRI, and data should not be combined without careful consideration. To account for this, we ensured that each abdominal feature was derived from a consistent pipeline, selecting the one with the largest available sample size across different pipelines. For liver MRI metrics specifically, we included only images acquired using the IDEAL protocol (Data-Field 40063, acquisition protocol 2). Following these additional QC steps, our analyses incorporated abdominal MRI biomarkers from all available UKBB participants for the liver, kidney, spleen, pancreas, and adipose MRIBAGs.
**(b)** **AI/ML models**: Our previous study systematically evaluated age prediction performance across various AI/ML models using multi-modal brain MRI features^3^ for the brain MRIBAG. Applying the same framework, we assessed the performance of models in deriving the 6 additional MRIBAGs using several ML methods. Hyperparameter tuning was performed through nested, repeated hold-out cross-validation^60^ with 50 repetitions (80% training/validation and 20% testing). Specifically, the hyperparameters of each model were tuned using grid search: Lasso regression’s alpha (α), linear support vector regressor’s *C*, and both alpha and L1-ratio for the elastic net. Due to the flexibility and large hyperparameter space of neural networks (e.g., number of layers and neurons), they were not included in the nested cross-validation pipeline. Instead, the neural network was manually tuned and evaluated on the cross-validation test dataset, and the final model was subsequently assessed on the independent test set. An independent test dataset was held out to unbiasedly evaluate model performance (**Supplementary eFigure 22**).
**(c)** **Dataset split**: To rigorously train the AI/ML models, we first defined participants without any pathologies based on ICD code and clinical history as CN (*N*=5291 for the heart MRIBAG, varying across the 7 organs/tissues). We further split the CN into the following datasets:
  - **CN independent test dataset**: 500 participants were randomly drawn from the CN population. Independent test datasets are ideal for objectively evaluating ML model performance, especially in studies with large sample sizes like the current one.
  - **CN training/validation dataset**: 80% of the remaining 4791 CN (taking the heart MRIBAG for illustration) were used for the inner loop 10-fold CV for hyperparameter selection;
  - **CN cross-validation test dataset**: 20% of the remaining 4791 CN were used for the outer loop 50 repetitions;
  - **PT dataset**: 29,785 patients who have at least one ICD-10-based diagnosis.

The CN training/validation/test datasets were used for model development and were employed with a nested cross-validation procedure for 4 AI/ML models (Lasso regression and support vector regressor, elastic net, and neural network), while the independent test set provided an unbiased assessment of model performance. Model evaluation metrics included mean absolute error (MAE) and Pearson’s *r*. Importantly, consistent with our prior studies, only CN participants were included in the training/validation dataset for modeling and clinical interpretation considerations^7^. The BAG was determined by subtracting the participant’s chronological age from the AI/ML-predicted age. Age bias correction was applied using the approach outlined by Beheshti et al.^22^, and has been systematically discussed in our previous comment^7^. To summarize, we applied an age bias correction method commonly used in neuroimaging studies, where a linear regression of the BAG on chronological age is used to estimate and remove systematic age-related bias via the model’s slope and intercept. Importantly, to address potential domain shift in the PT data compared to the CN training set, we computed the regression parameters directly from the PT set to more accurately correct for this bias (refer to our comment^7^ for further details).

### Method 3: Proteome and metabolome-wide associations with the 7 MRIBAGs

**(a)** **ProWAS**: We utilized the original dataset (Category code: 1838), which was analyzed and shared with the research community by the UKB-PPP^61^. The initial quality control procedures were described in the original study^62^, and we implemented additional quality control steps as outlined below. Our analysis focused on the first instance of the proteomics data (“instance” = 0). We then integrated Olink files containing coding information, batch numbers, assay details, and limit of detection (LOD) data (Category ID: 1839) by matching them to the proteomics dataset ID. Finally, we excluded Normalized Protein eXpression (NPX) values that fell below the protein-specific LOD. We conducted ProWASs by linking the 7 MRIBAGs to 2923 unique plasma proteins measured in 53,016 participants (with sample sizes ranging from 10,018 to 39,489 per protein after quality control) using the Olink platform. The linear regression model adjusted for common covariates, including age (Field ID: 21003), sex (Field ID: 31), body weight (Field ID: 21002), height (Field ID: 50), waist circumference (Field ID: 48), BMI (Field ID: 23104), assessment center (Field ID: 54), protein batch number (Category ID: 1839), limit of detection (LOD; Category ID: 1839), and the first 40 genetic principal components. Additionally, organ-specific covariates were incorporated, such as brain positioning in the scanner (lateral, transverse, and longitudinal; Field ID: 25756-25758), head motion (Field ID: 25741), and intracranial volume for the brain MRIBAG, as well as diastolic (Field ID: 4079) and systolic (Field ID: 4080) blood pressure for the heart MRIBAG, among others. Multiple testing corrections were applied using Bonferroni adjustment (P-value < 0.05/2923/7). To identify and exclude extreme outliers, we defined an upper threshold as the mean plus six times the standard deviation for each protein/metabolite. Finally, ProWAS signals identified in the UKBB Olink dataset were compared with SomaScan data from BLSA.
**(b)** **MetWAS**: We used the original data (Category ID: 220), which were analyzed and made available to the community by Nightingale Health Plc. The original data were *i*) calibrated absolute concentrations (or ratios) and not raw NMR spectra, and *ii*) before release, had already been subject to quality control procedures by Nightingale Health Plc^63^. Following the additional procedures described in Ritchie et al.^64^, we performed additional quality check steps to remove a range of unwanted technical variations, including shipping batch, 96-well plate, well position, aliquoting robot, and aliquot tip. We focused our analysis on the first instance of the metabolomics data (“instance”=0). The analysis included 327 metabolites (comprising both small molecules and lipoprotein measures), of which 107 were non-derived metabolites and the remainder were composite metabolites, across 274,247 participants. Descriptions of these metabolites are provided in **Supplementary eTable 3**. We conducted MetWASs by linking the 7 MRIBAGs to 327 plasma metabolites. The linear regression model adjusted for common covariates, including age (Field ID: 21003), sex (Field ID: 31), body weight (Field ID: 21002), height (Field ID: 50), waist circumference (Field ID: 48), BMI (Field ID: 23104), assessment center (Field ID: 54), and the first 40 genetic principal components. Additionally, organ-specific covariates were incorporated, such as brain positioning in the scanner (lateral, transverse, and longitudinal; Field ID: 25756-25758), head motion (Field ID: 25741), and intracranial volume for the brain MRIBAG, as well as diastolic (Field ID: 4079) and systolic (Field ID: 4080) blood pressure for the heart MRIBAG, among others. Multiple testing corrections were applied using Bonferroni adjustment (P-value < 0.05/327/7).

### Method 4: Genetic analyses

We used the imputed genotype data from UKBB for all genetic analyses. Our quality check pipeline focused on European ancestry in UKBB (6,477,810 SNPs passing quality checks), and the quality-checked genetic data were merged with respective organ-specific populations for GWAS. We summarize our genetic quality check steps. First, we skipped the step for family relationship inference^65^ because the linear mixed model via fastGWA^66^ inherently addresses population stratification, encompassing additional cryptic population stratification factors. We then removed duplicated variants from all 22 autosomal chromosomes. Individuals whose genetically identified sex did not match their self-acknowledged sex were removed. Other excluding criteria were: *i*) individuals with more than 3% of missing genotypes; *ii*) variants with minor allele frequency (MAF; dosage mode) of less than 1%; *iii*) variants with larger than 3% missing genotyping rate; *iv*) variants that failed the Hardy-Weinberg test at 1×10^−10^. To further adjust for population stratification,^67^ we derived the first 40 genetic principal components using the FlashPCA software^68^. Details of the genetic quality check protocol are described elsewhere^52,3,1,42,69^.

**(a):** **GWAS** We applied a linear mixed model regression to the European ancestry populations using fastGWA^66^ implemented in GCTA^24^. We used fastGWA to perform the 7 MRIBAG GWASs, adjusting common variates, including age, dataset status (training/validation/test or independent test), age-squared, sex, interactions of age with sex, BMI, waist circumference, standing height, weight, and the first 40 genetic principal components, as well as organ-specific covariates, including the brain scan positions for the brain MRIBAG, and systolic/diastolic blood pressure for the heart MRIBAG, etc. We applied a genome-wide significance threshold (5×10^−8^) to annotate the significant independent genomic loci. We previously conducted GWAS of 2,923 plasma proteins and 327 metabolites using fastGWA^70,5^. **Annotation of genomic loci**: For all GWASs, genomic loci were annotated using FUMA^71^. For genomic loci annotation, FUMA initially identified lead SNPs (correlation *r*^2^ ≤ 0.1, distance < 250 kilobases) and assigned them to non-overlapping genomic loci. The lead SNP with the lowest P-value (i.e., the top lead SNP) represented the genomic locus. Further details on the definitions of top lead SNP, lead SNP, independent significant SNP, and candidate SNP can be found in **Supplementary eNote 5**. For visualization purposes in **Fig. 3a**, we have mapped the top lead SNP of each locus to the cytogenetic regions based on the GRCh37 cytoband.
**(b):** **SNP-based heritability**: We estimated the SNP-based heritability (*h*^2^) using GCTA^24^ with the same covariates as in GWAS. GCTA estimates the SNP-based heritability using a method called restricted maximum likelihood (REML) to quantify the proportion of phenotypic variance in a trait that the additive effects of all common SNPs can explain. The main steps involved in GCTA include constructing the genetic relationship matrix, modeling phenotypic variance, and using REML to estimate the *h*^2^.
**(c):** **Partitioned heritability estimate**: The partitioned heritability analysis via stratified LD score regression calculates the extent to which heritability enrichment can be attributed to predefined and annotated genome regions and categories^72^. Three sets of functional categories and cell and tissue-specific types were considered. First, the partitioned heritability was calculated for 53 general functional categories (one including the entire set of SNPs). The 53 functional categories are not specific to any cell type and include coding, UTR, promoter, and intronic regions, etc. The details of the 53 categories are described elsewhere^72^. Subsequently, cell and tissue type-specific partitioned heritability was estimated using gene sets from Cahoy et al.^73^ for three main cell types (i.e., astrocyte, neuron, and oligodendrocyte), multi-tissue chromatin states-specific data (ROADMAP^74^ and ENTEx^75^), and multi-tissue gene expression data (GTEx V8^76^). Bonferroni correction was performed for all tested annotations and categories. The detailed methodologies for the stratified LD score regression are presented in the original work^72^. The LD scores and allele frequencies for the European ancestry were obtained from a predefined version based on data from the 1000 Genomes project.
**(d):** **Genetic correlation**: We estimated the genetic correlation (*g_c_*) using the LDSC^77^ software. We employed precomputed LD scores from the 1000 Genomes of European ancestry, maintaining default settings for other parameters in LDSC. It’s worth noting that LDSC corrects for sample overlap, ensuring an unbiased genetic correlation estimate^78^. Statistical significance was determined using Bonferroni correction.
**(e):** **Two-sample bidirectional Mendelian randomization**: We constructed two-sample bidirectional Mendelian randomization by linking the 7 MRIBAGs and 525 DEs from FinnGen^19^ and PGC^20^. In total, 2 networks were established: i) *MRIBAG2DE* and ii) *DE2MRIBAG*. The systematic quality-checking procedures to ensure unbiased exposure/outcome variable and instrumental variable (IVs) selection are detailed below. We used a two-sample Mendelian randomization approach implemented in the *TwoSampleMR* package^79^ to infer the causal relationships within these networks. We employed five distinct Mendelian randomization methods, including the inverse variance weighted (IVW) method, Egger, weighted median, simple mode, and weighted mode estimators. The STROBE-MR Statement^80^ guided our analyses to increase transparency and reproducibility, encompassing the selection of exposure and outcome variables, reporting statistics, and implementing sensitivity checks to identify potential violations of underlying assumptions. First, we performed an unbiased quality check on the GWAS summary statistics. Notably, the absence of population overlapping bias^81^ was confirmed, given that FinnGen and UKBB participants largely represent populations of European ancestry without explicit overlap with UKBB. PGC GWAS summary data were ensured to exclude UKBB participants. Furthermore, all consortia’s GWAS summary statistics were based on or lifted to GRCh37. Subsequently, we selected the effective exposure variables by assessing the statistical power of the exposure GWAS summary statistics in terms of instrumental variables (IVs), ensuring that the number of IVs exceeded 7 before harmonizing the data. Crucially, the function “*clump_data*” was applied to the exposure GWAS data, considering LD. The function “*harmonise_data*” was then used to harmonize the GWAS summary statistics of the exposure and outcome variables. Bonferroni correction was applied to all tested traits based on the number of effective DEs. Finally, we conducted multiple sensitivity analyses. First, we conducted a heterogeneity test to scrutinize potential violations of the IV’s assumptions. To assess horizontal pleiotropy, which indicates the IV’s exclusivity assumption^82^, we utilized a funnel plot, single-SNP Mendelian randomization methods, and the Egger estimator. Furthermore, we performed a leave-one-out analysis, systematically excluding one instrument (SNP/IV) at a time, to gauge the sensitivity of the results to individual SNPs.
**(f)** **PRS calculation**: PRS was computed using split-sample GWASs (split1 and split2) for the 7 MRIBAG GWASs. The PRS weights were established using split1/discovery GWAS data as the base/training set, while the split2/replication GWAS summary statistics served as the target/testing data. Both base and target data underwent rigorous quality control procedures involving several steps: *i*) excluding duplicated and ambiguous SNPs in the base data; *ii*) excluding high heterozygosity samples in the target data; and *iii*) eliminating duplicated, mismatching, and ambiguous SNPs in the target data. After completing the QC procedures, PRS for the split2 and split1 groups was calculated using the PRS-CS^83^ method. PRS-CS applies a continuous shrinkage prior, which adjusts the SNP effect sizes based on their LD structure. SNPs with weaker evidence are “shrunk” toward zero, while those with stronger evidence retain larger effect sizes. This avoids overfitting and improves prediction performance. The shrinkage parameter was not set, and the algorithm learned it via a fully Bayesian approach.
**(g)** **Genetic evidence for prioritizing potential druggable genes**: We conducted multiple analyses to establish genetic evidence for prioritizing potential druggable genes in future drug repurposing. Our central hypothesis is that genes with functional implications and causal roles validated across multiple omics layers, such as genomics, transcriptomics, and proteomics (**Supplementary eFigure 10**), are more likely to be actionable for drug repurposing, as they offer a stronger and more robust foundation for identifying therapeutic targets^84,85^.

First, we performed SNP-to-gene mapping via three approaches: *i*) positional mapping, *ii*) eQTL mapping, and *iii*) chromatin interaction mapping. For eQTL and chromatin interaction mapping, we utilized data from various resources, including GTEx, PsychENCODE, EyeGEx, TIGER, DICE, eQTLGen, Blood eQTL, MuTHER, CommonMind Consortium, BRAINEAC,

FANTOM5, the Brain Open Chromatin Atlas, and the Roadmap 111 epigenomes, all consolidated through the FUMA platform. By requiring both eQTL and chromatin interaction support, we enhance confidence in selecting functionally relevant genes because eQTL mapping identifies genetic variants that regulate gene expression. Chromatin interaction mapping links distal regulatory elements (e.g., enhancers) to target genes, offering spatial genomic context.

Together, these criteria ensure that the prioritized genes are not only associated with GWAS loci but also have strong regulatory and functional evidence, making them more biologically plausible as druggable targets.

Subsequently, we connected MRIBAG, as an imaging-derived endophenotype, to aging clocks derived from plasma proteomics and metabolomics to further strengthen the genetic evidence for prioritizing these druggable genes. To achieve this, we used a Bayesian colocalization approach to assess whether the genomic loci associated with the 7 MRIBAGs share common causal variants with the 11 ProtBAGs or the 5 MetBAGs. We used the R package (*coloc*) to investigate the genetic colocalization signals between two traits (e.g., the brain MRIBAG vs. brain ProtBAG) at each genomic locus associated with the MRIBAG. This method (*coloc.abf*) examines the posterior probability (PP.H4.ABF: Approximate Bayes Factor) to evaluate hypothesis *H4*, which suggests the presence of a single shared causal variant associated with both traits within a specific genomic locus. To determine the significance of the *H4* hypothesis, we set a threshold of PP.H4.ABF>0.8^86^. All other parameters (e.g., the prior probability of p_12_) were set as default. For each pair of traits, the genomic locus was defined by default from FUMA for one trait, and then the *coloc* package extracted and harmonized the GWAS summary statistics within this locus for the other trait.

Finally, we searched the DGIdb platform (https://dgidb.org/) for genes that demonstrated both functional mapping evidence and causal associations to investigate their drug-gene interactions, revealing existing drugs and their therapeutic indications.

### Method 5: Prediction analyses for 14 systemic disease categories, the risk of mortality, and single disease endpoints

We investigated the clinical promise of the 7 MRIBAGs and their PRSs (MRIBAG-PRS) in three sets of prediction analyses: *i*) survival analysis for the incidence of single disease entities based on the ICD-10 code, *ii*) survival analysis for the risk of all-cause mortality, and *iii*) differential response to the AD drug (Solanezumab).

**(a)** **Survival analysis for ICD-based single disease endpoint**: we employed a Cox proportional hazard model while adjusting for covariates(i.e., age and sex) to test the associations of the 7 MRIBAGs with the incidence of ICD-based single disease entities. The covariates were included as additional right-side variables in the model. To train the model, the “time” variable was determined by calculating the difference between the date of death (Field ID: 40000) for cases (or the censoring date for non-cases) and the date attending the assessment center (Field ID: 53). Participants who were diagnosed for a specific disease of interest after enrolling in the study were classified as cases; non-cases were defined by participants without any disease diagnoses.
**(b)** **Survival analysis for mortality risk**: we employed a Cox proportional hazard model while adjusting for covariates(i.e., age and sex) to test the associations of the 7 MRIBAGs and 7 MRIBAG-PRSs with all-cause mortality. The covariates were included as additional right-side variables in the model. The hazard ratio (HR), exp(β*_R_*), was calculated and reported as the effect size measure that indicates the influence of each biomarker on the risk of mortality. To train the model, the “time” variable was determined by calculating the difference between the date of death (Field ID: 40000) for cases (or the censoring date for non-cases) and the date attending the assessment center (Field ID: 53). Participants who passed away after enrolling in the study were classified as cases. We also conducted a disease-free survival analysis, including only participants without any diagnosed conditions, to eliminate potential confounding effects from disease pathology.
**(c)** **Differential trajectories of cognitive decline based on brain MRIBAG-defined decelerated and accelerated aging groups from drug outcomes in the A4 study**: To evaluate our hypothesis that individuals with varying aging clock paces may progress differently to cognitive decline, we analyzed clinical trial data from the A4^17^ study. Specifically, we investigated whether brain MRIBAG could be used to stratify participants into decelerated and accelerated aging groups and demonstrated different cognitive profiles at 240 weeks by taking Solanezumab. The original trial did not demonstrate cognitive decline slowing compared to the placebo over 240 weeks, with the treatment group even showing a slight worsening in the PACC score. To evaluate this, we performed four comparisons using natural cubic spline modeling: *i*) decelerated aging (e.g., below the brain MRIBAG median) vs. accelerated aging (e.g., above the median) within the drug group, and *ii*) within the placebo group; *iii*) between drug and placebo groups among participants with accelerated aging, and *iv*) between drug and placebo groups among those with decelerated aging.

From a statistical perspective, this is to model repeated measures (i.e., PACC as the primary trial outcome for global cognition) as a continuous outcome in a mixed-effect model. We used the same method proposed by the original work from Donohue et al.^87^, in which the authors proposed a constrained longitudinal data analysis with natural cubic splines that treated time as continuous and used test version effects to model the mean over time for PACC. Fixed effects included the following terms: *i*) spline basis expansion terms (two terms), *ii*) interaction of the spline basis expansion terms with treatment (two terms), *iii*) the version of the PACC test implemented, *iv*) baseline age, *v*) education, *vi*) *APOE4* carrier status (yes/no), and *vii*) baseline florbetapir cortical SUVr value. At week 240, we compared the mean PACC values between each pair of the two groups.

## Data Availability

The GWAS summary statistics and pre-trained AI models from this study are publicly accessible via the MEDICINE Knowledge Portal (https://labs-laboratory.com/medicine/) and Synapse (https://www.synapse.org/Synapse:syn64923248/wiki/630992^88^). Our study used data generated by the Human Protein Atlas (https://www.proteinatlas.org). GWAS summary data for the DEs were downloaded from the official websites of FinnGen (R9: https://www.finngen.fi/en/access_results) and PGC (https://pgc.unc.edu/for-researchers/download-results/). Individual data from UKBB can be requested with proper registration at https://www.ukbiobank.ac.uk/. The A4 study data can be requested at: https://www.a4studydata.org/. The NIA-supported BLSA data can be requested at: https://www.nia.nih.gov/research/labs/blsa. Certain sensitive data (e.g., allele frequency information) supporting the findings are also available from the author upon request.

## Code Availability

The software and resources used in this study are all publicly available:

- MLNI^89^ (v0.1.2 and v0.1.5): https://github.com/anbai106/mlni, MRIBAG generation
- FUMA (v1.5.0): https://fuma.ctglab.nl/, Gene mapping, genomic locus annotation;
- GCTA (v1.94.1): https://yanglab.westlake.edu.cn/software/gcta/#Overview, fastGWA;
- LDSC (git version aa33296): https://github.com/bulik/ldsc, genetic correlation
- TwoSampleMR (v0.5.6): https://mrcieu.github.io/TwoSampleMR/index.html, Mendelian randomization;
- PRScs (release date: Aug 10, 2023): https://github.com/getian107/PRScs, PRS calculation;
- Lifelines (v0.27.8): https://lifelines.readthedocs.io/en/latest/, Survival analysis;
- String (V12.0): https://string-db.org/, protein analyses.
- coloc (v5): https://github.com/chr1swallace/coloc; Bayesian colocalization.

## Competing Interests

B.W.D. is an inventor of the DunedinPACE epigenetic clock, which is licensed by Duke University and the University of Otago to TruDiagnostic, from which he receives royalties (DunedinPACE is freely available to researchers). All other authors declare no competing interests.

## Authors’ contributions

Dr. Wen has full access to all the study data and is responsible for its integrity and accuracy.

*Study concept and design*: W.J.

*Drafting of the manuscript*: W.J.

*Critical revision of the manuscript for important intellectual content*: All authors

*Statistical analysis*: W.J. ran all main analyses; C.H. helped with analyses using GWAS summary data from FinnGen and PGC; S.Z. helped with the development of the MEDICINE portal; D.M. ran the ProWAS for the SomaScan proteomics data in the BLSA sample.

*Detailed contributions*: W.J. leads the MULTI consortium; D.C. leads the iSTAGING consortium; W.A.K. represents the BLSA study; R.S.M and A.P represent the A4 study; B.W. derived the original heart IDPs. Z.A. and T.Y. derived the 9 original PhenoBAGs, and we only used the derived GWAS summary statistics in this study.

## Acknowledgments

Dr. Junhao Wen and the Laboratory of AI and Biomedical Science (LABS) are supported by the start-up funding from Columbia University. The MULTI consortium (J.W; UK Biobank Application Number: 647044) aims to integrate multi-organ imaging and multi-omics data to advance our understanding of human aging and disease mechanisms. This study used the UK Biobank resource under Application Number 35148 (D.C.; NIA grant number: RF1 AG054409). We want to express our sincere gratitude to the UK Biobank team for their invaluable contribution to advancing clinical research in our field (https://www.ukbiobank.ac.uk/). We want to acknowledge the participants and investigators of the FinnGen study and the PGC consortium, and we thank FinnGen (https://www.finngen.fi/en) and PGC (https://pgc.unc.edu/) for their generosity in sharing the GWAS summary statistics with the scientific community. We thank the BLSA participants and staff for their participation and continued dedication. The BLSA protocol was approved by the Institutional Review Board of the National Institute of Environmental Health Science, National Institutes of Health (03AG0325). The A4 Study was a secondary prevention trial in preclinical Alzheimer’s disease, aiming to slow cognitive decline associated with brain amyloid accumulation in clinically normal older individuals. The A4 Study was funded by a public-private-philanthropic partnership, including funding from the National Institutes of Health-National Institute on Aging, Eli Lilly and Company, Alzheimer’s Association, Accelerating Medicines Partnership, GHR Foundation, an anonymous foundation, and additional private donors, with in-kind support from Avid Radiopharmaceuticals, Cogstate, Albert Einstein College of Medicine and the Foundation for Neurologic Diseases. The companion observational Longitudinal Evaluation of Amyloid Risk and Neurodegeneration (LEARN) Study was funded by the Alzheimer’s Association and GHR Foundation. The A4 and LEARN Studies were led by Dr. Reisa Sperling at Brigham and Women’s Hospital, Harvard Medical School, and Dr. Paul Aisen at the Alzheimer’s Therapeutic Research Institute (ATRI) at the University of Southern California. The A4 and LEARN Studies were coordinated by ATRI at the University of Southern California, and the data are made available under the auspices of the Alzheimer’s Clinical Trial Consortium through the Global Research & Imaging Platform (GRIP). The complete A4 Study Team list is available at: https://www.actcinfo.org/a4-study-team-lists/. We want to acknowledge the dedication of the study participants and their partners who made the A4 and LEARN Studies possible. D.R.M, W.A.K, and F.A were supported by the Intramural Research Program (IRP) of the NIH, National Institute on Aging (NIA). We acknowledge the leadership of the Brain Imaging Genetics (BIG) workgroup, led by Dr. Tavia Evans, Dr. Natalia Vilor-Tejedor, and Dr. Junhao Wen, within the International Society to Advance Alzheimer’s Research and Treatment (ISTAART) community, for advocating brain imaging genetics in Alzheimer’s and aging research.

## Extended figures

**Extended Data Figure 1:**
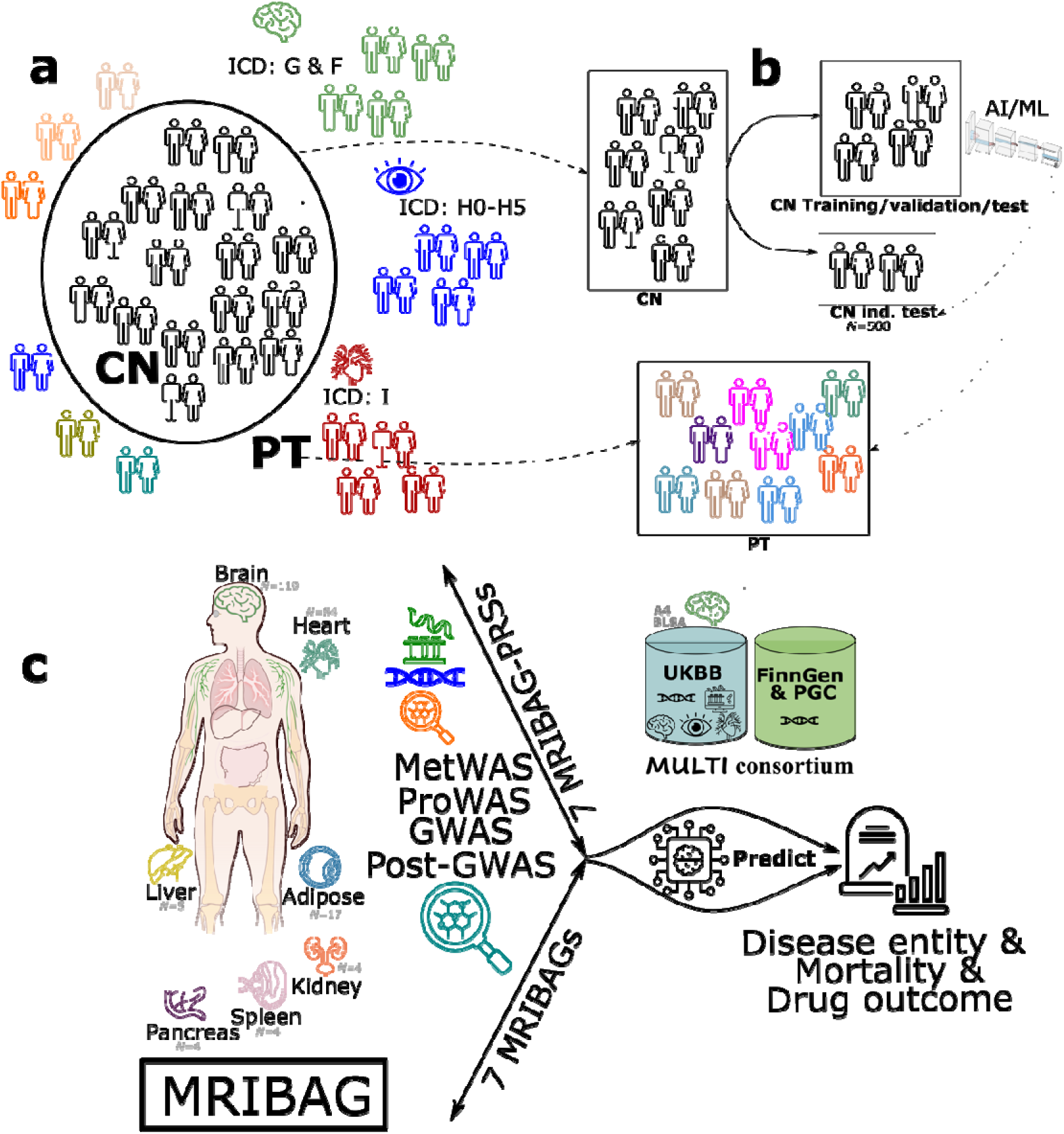
Schematic figure of the population definition and workflow used to derive MRIBAG. **a)** For each organ-specific MRIBAG, we split the UK Biobank populations into healthy controls (CN, without any pathologies) and patients (PT) based on ICD-10 codes and their clinical history. **b)** To develop the 7 MRIBAGs, we trained two AI/ML models exclusively on the CN training/validation/test population using a nested cross-validation procedure to identify the optimal model. Independent datasets included the CN independent test dataset (*N*=500) and the PT population. **c**) The study’s analytical workflow encompassed deriving the 7 MRIBAGs and their PRSs, performing ProWAS, MetWAS, GWAS, and post-GWAS analyses, and assessing the predictive performance of the MRIBAG and MRIBAG-PRSs for single disease entities, mortality, and drug outcomes. Data from the BLSA and A4 studies were used to generate the brain MRIBAG in independent cohorts.

**Extended Data Figure 2:**
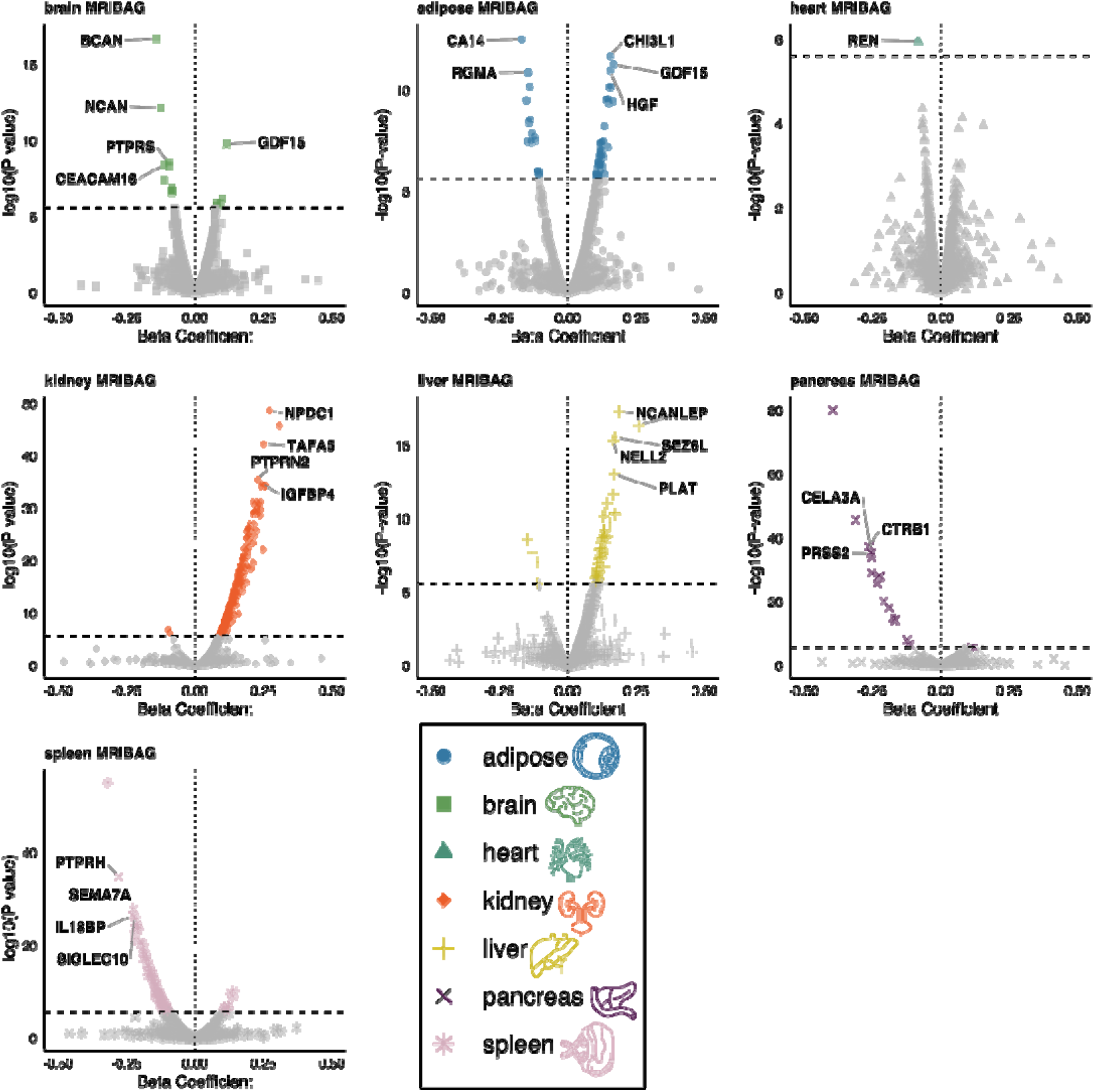
Individual volcano plots for the 7 MRIBAGs in ProWAS. Proteome-wide association studies (ProWAS) between the 7 MRIBAGs and 2923 plasma proteins via a linear regression model, accounting for a full set of covariates. Representative significant (P-value<0.05/2923/7, indicated by the dotted horizontal line) MRIBAG-protein associations are annotated by the respective gene symbol. We used the standardized *β* value as an effect size measurement.

**Extended Data Figure 3:**
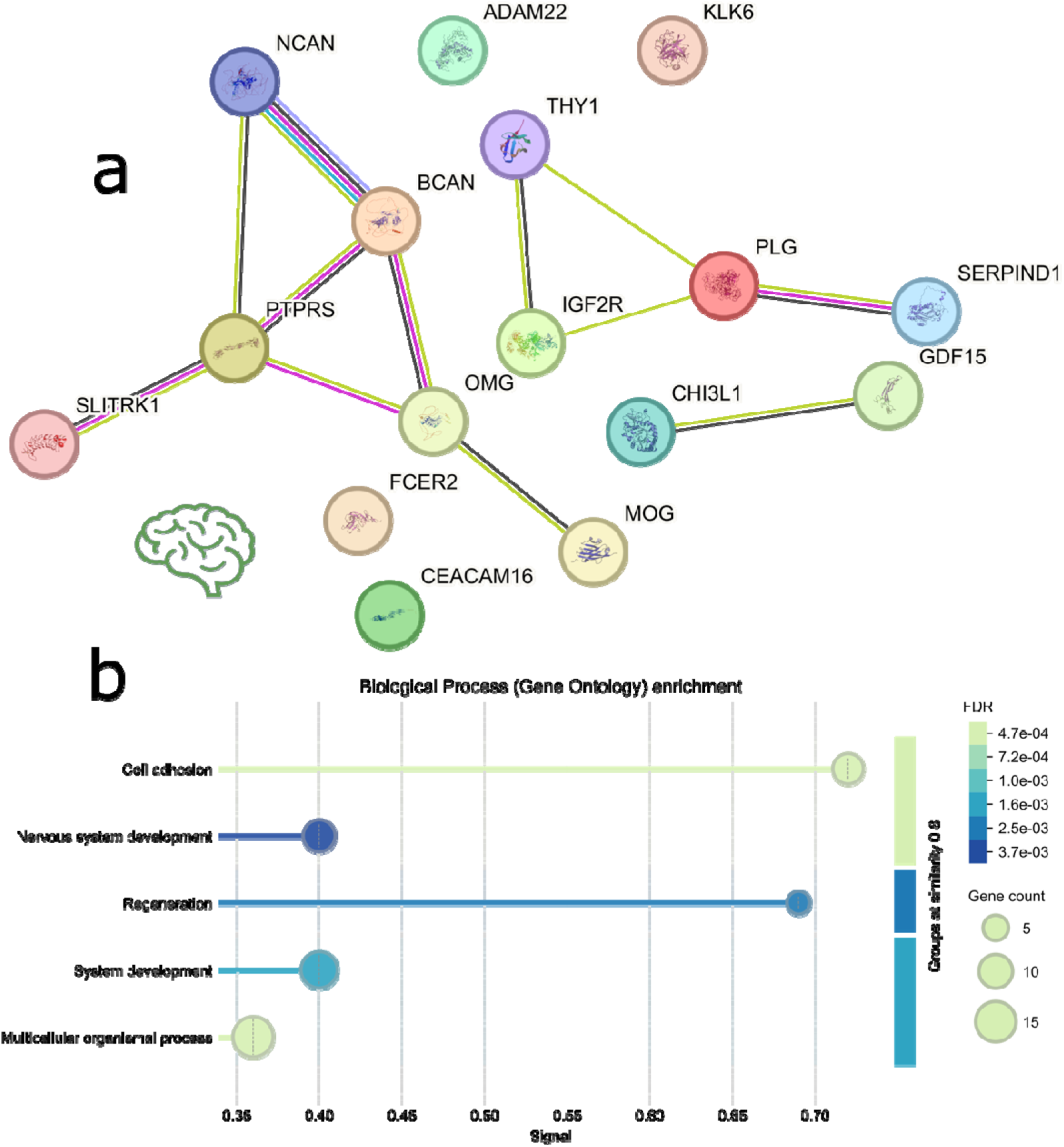
Functional protein-set enrichment analysis (PSEA) and protein-protein interaction (PPI) network for the brain MRIBAG-linked proteins. STRING conducts protein-protein interaction (PPI) analysis by combining various sources of interaction evidence, such as experimental data, computational predictions, and text mining. In addition to interactions, STRING can carry out functional enrichment analysis to identify Gene Ontology (GO) terms (e.g., biological processes, molecular functions), KEGG pathways (e.g., metabolic and signaling pathways), and protein domains (e.g., shared structural motifs). **a**) PPI network of the brain MRIBAG-related proteins. **b**) Functional PSEA of the brain MRIBAG-related proteins. Results and figures are generated using STRING v12.0 (https://string-db.org/).

**Extended Data Figure 4:**
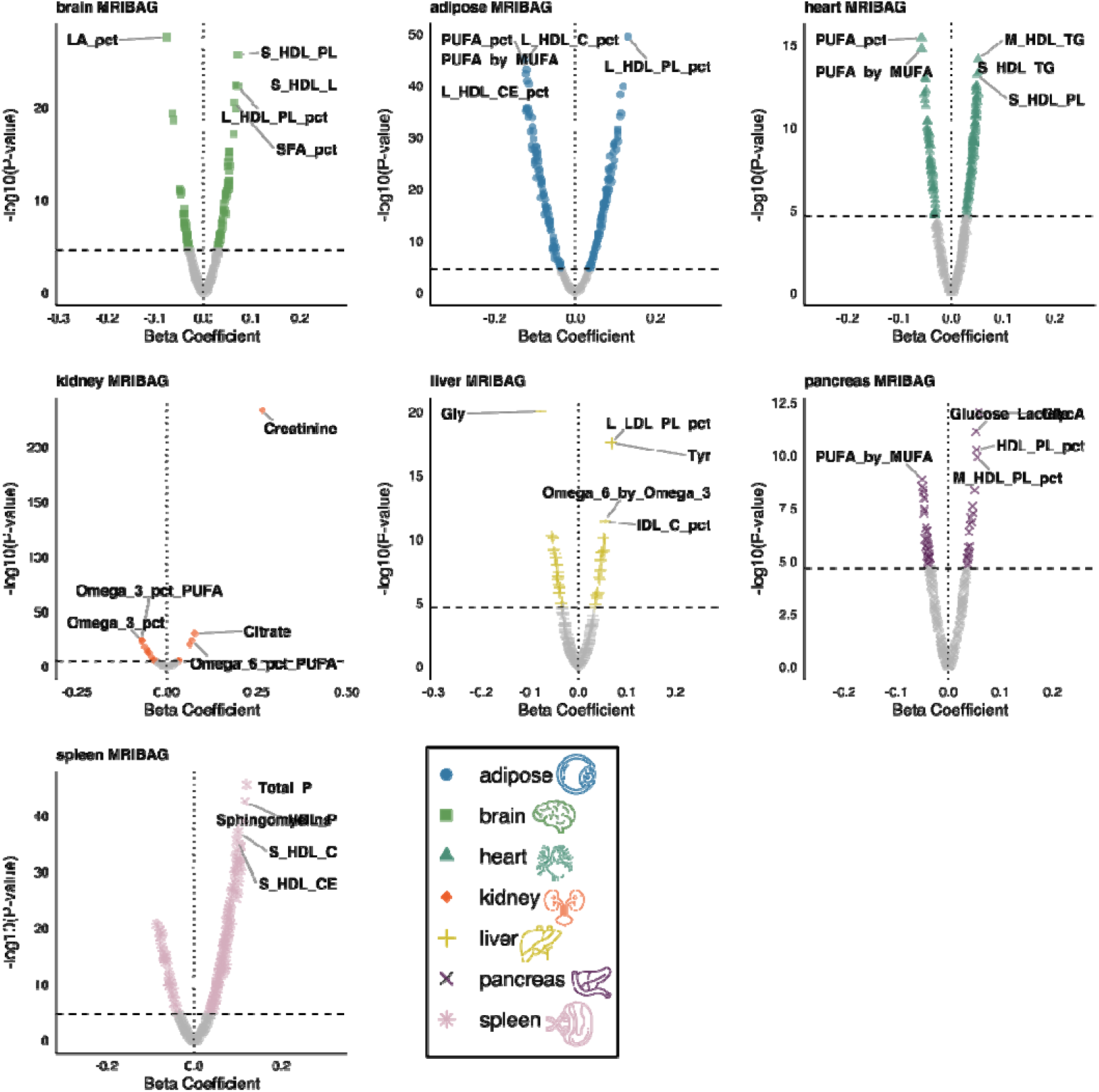
Individual volcano plots for the 7 MRIBAGs in MetWAS. Metabolome-wide association studies (MetWAS) between the 7 MRIBAGs and 327 plasma metabolites via a linear regression model, accounting for a full set of covariates. Representative significant (P-value<0.05/327/7, indicated by the dotted horizontal line) MRIBAG-metabolite associations are annotated. We used the standardized *β* value as an effect size measurement.

**Extended Data Figure 5:**
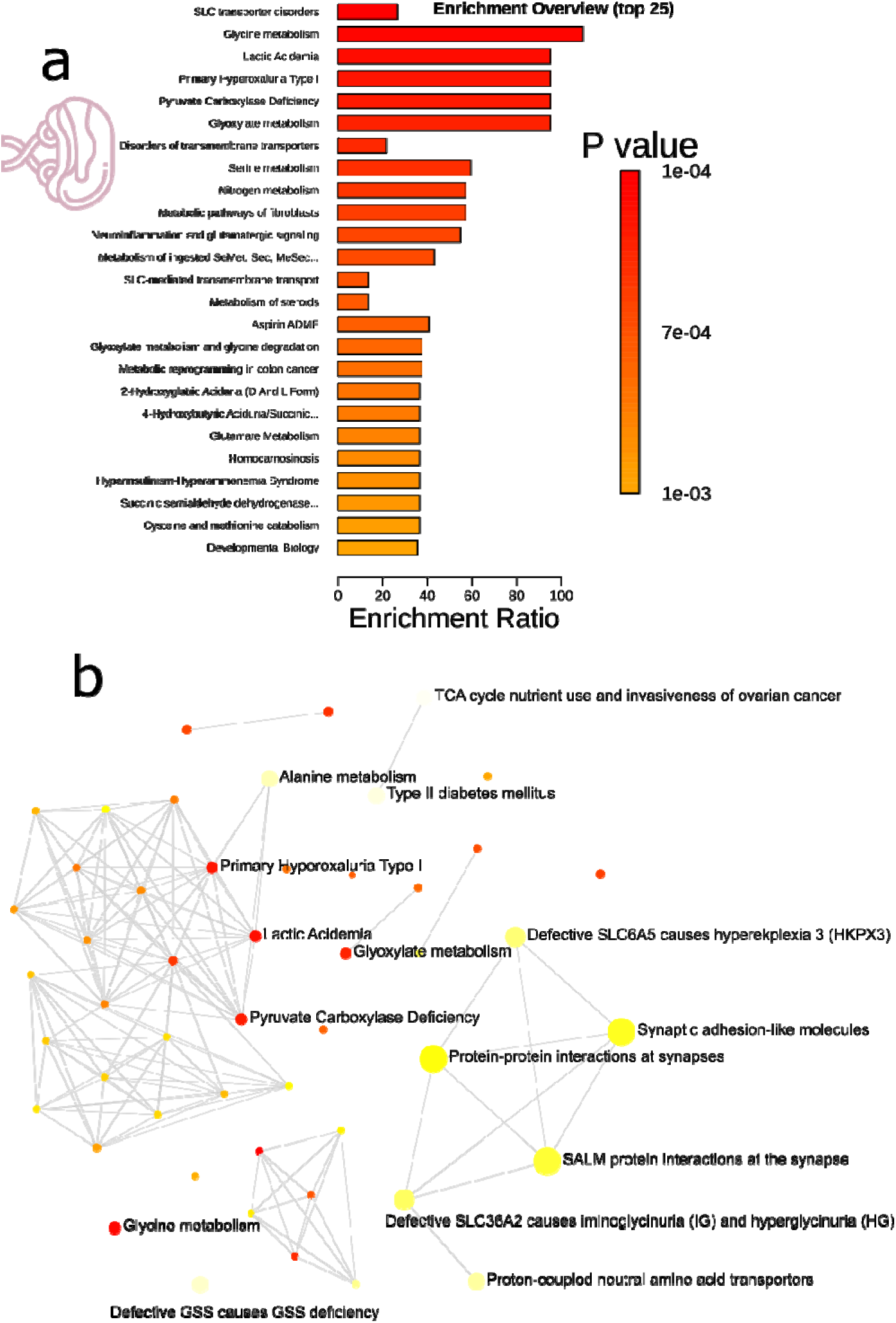
Functional metabolite-set enrichment analysis (MSEA) and metabolite-metabolite interaction (MMI) network for the 6 spleen MRIBAG-linked small-molecular metabolites. **a)** Top 25 MSEA results for the spleen MRIBAG-related small molecule metabolites. **b**) The MMI network results for these metabolite sets are displayed as follows: each node represents a metabolite set, with color indicating the enrichment P-value and size reflecting the fold enrichment ratio. Two metabolite sets are connected by an edge if the proportion of shared metabolites between them exceeds 25% of the total combined metabolites. Results and figures are generated using MetaboAnalyst v6.0 (https://www.metaboanalyst.ca/). We used 3694 metabolite and lipid pathways from RaMP-DB (which integrates KEGG via HMDB, Reactome, and WikiPathways, curated via MetaboAnalyst) as the background metabolite dataset. Only metabolite sets with at least two entries were included.

**Extended Data Figure 6:**
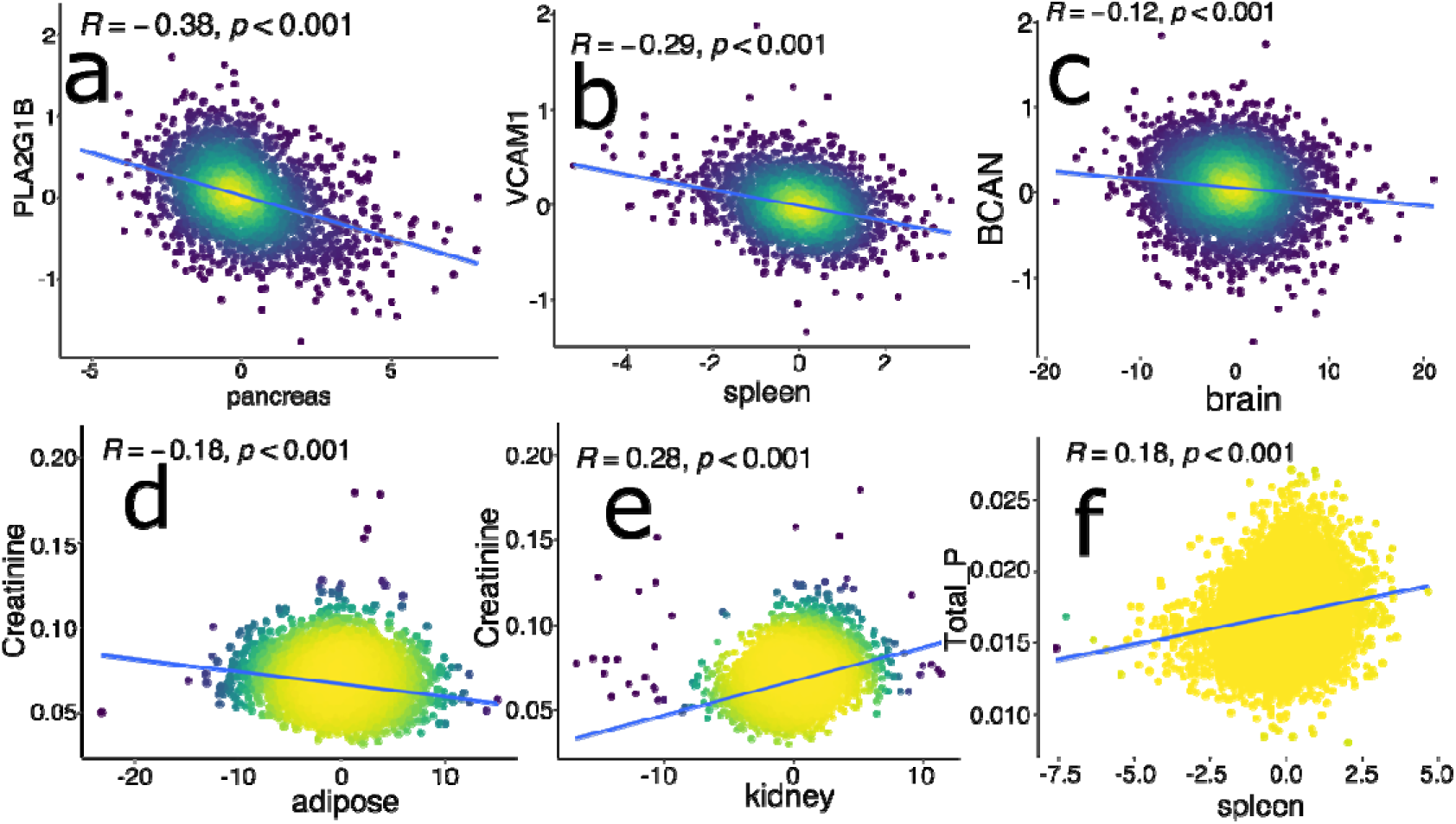
Example associations for the ProWAS and MetWAS results. **a)** ProWAS results for the PLA2G1B protein and the pancreas MRIBAG. **b**) ProWAS results for the VCAM1 protein and the spleen MRIBAG. **c**) ProWAS results for the BCAN protein and the brain MRIBAG. **d**) MetWAS results for the creatinine metabolite and the adipose MRIBAG. **e**) MetWAS results for the creatinine metabolite and the kidney MRIBAG. **f**) MetWAS results for total concentration of phospholipids (Total_P) and the spleen MRIBAG. A linear regression model was fitted, and the resulting Pearson’s *r* and P-value are reported.

**Extended Data Figure 7:**
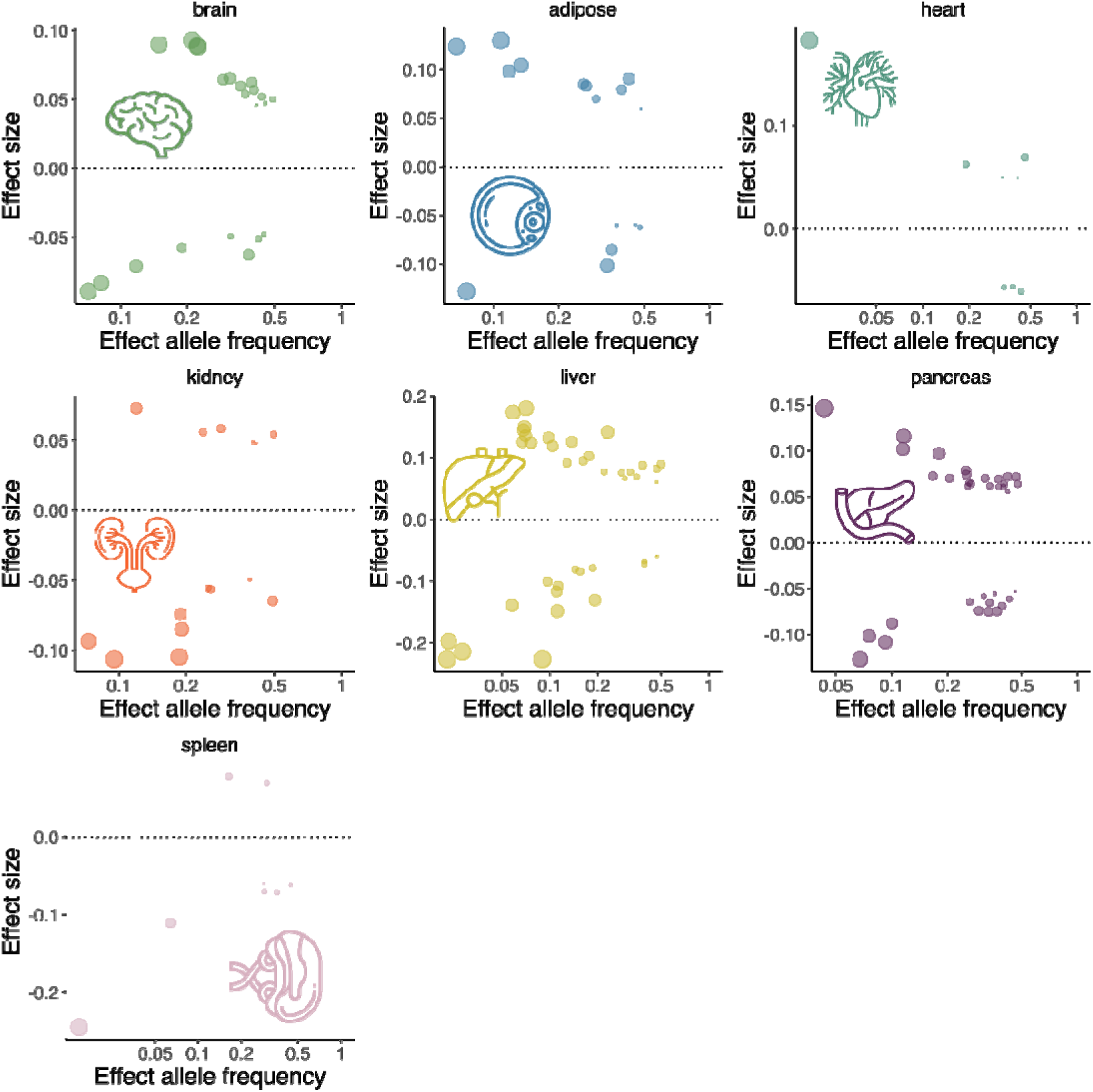
Trumpet plots of the effect allele frequency vs. the *β* coefficient of the 7 MRIBAG GWASs. The trumpet plots display the inverse relationship between the alternative (effect) allele frequency and the effect size (*β* coefficient) for the 7 MRIBAGs. We present the independent, significant SNPs defined in FUMA. The dot size corresponds to the effect size, while the transparency of the dot is proportional to its statistical significance.

## The MULTI Consortium

Junhao Wen^1,9,10,11,12,13,14^*, Christos Davatzikos^3^, Ye Ella Tian^4^, Wenjia Bai^5^, Michael S. Rafii^6^, Paul Aisen^6^, Daniel W. Belsky^7^, Keenan A. Walker^2^, Andrew Zalesky^4^, Luigi Ferrucci^8^, Jian Zeng^15^

^1^Laboratory of AI and Biomedical Science (LABS), Columbia University, New York, NY, USA

^2^Laboratory of Behavioral Neuroscience, National Institute on Aging, National Institutes of Health, Baltimore, MD, USA

^3^Artificial Intelligence in Biomedical Imaging Laboratory (AIBIL), Center for AI and Data Science for Integrated Diagnostics (AI^2^D), Perelman School of Medicine, University of Pennsylvania, Philadelphia, PA, USA

^4^Systems Lab, Department of Psychiatry, Melbourne Medical School, The University of Melbourne, Melbourne, Victoria, Australia

^5^Department of Brain Sciences and Department of Computing, Imperial College London, London, UK

^6^Alzheimer’s Therapeutic Research Institute, Keck School of Medicine of the University of Southern California, San Diego, CA 92121, USA

^7^Robert N Butler Columbia Aging Center and Department of Epidemiology, Columbia University Mailman School of Public Health, New York, NY, USA

^8^National Institute on Aging, National Institutes of Health, Baltimore, MD, USA

^9^Center for Innovation in Imaging Biomarkers and Integrated Diagnostics (CIMBID), Department of Radiology, Columbia University, New York, NY, USA

^10^Department of Radiology, Columbia University, New York, NY, USA ^11^Department of Biomedical Engineering, Columbia University, New York, NY, USA ^12^Zuckerman Institute, Columbia University, New York, NY, USA

^13^New York Genome Center (NYGC), New York, NY, USA

^14^Data Science Institute (DSI), Columbia University, New York, NY, USA

^15^The Institute for Molecular Bioscience, University of Queensland, Brisbane, QLD 4072, Australia

## Notes

### Author Declarations

The MULTI consortium is approved by the Institutional Review Board at Columbia University (AAAV6751).

## References

1. Wen, J. et al. The genetic architecture of biological age in nine human organ systems. Nat Aging 1–18 (2024) doi:10.1038/s43587-024-00662-8.

2. Tian, Y. E. et al. Heterogeneous aging across multiple organ systems and prediction of chronic disease and mortality. Nat Med 1–11 (2023) doi:10.1038/s41591-023-02296-6.

3. Wen, J. et al. The genetic architecture of multimodal human brain age. Nat Commun 15, 2604 (2024).

4. Oh, H. S.-H. et al. Organ aging signatures in the plasma proteome track health and disease. Nature 624, 164–172 (2023).

5. Anagnostakis, F. et al. Multi-organ metabolome biological age implicates cardiometabolic conditions and mortality risk. Nat Commun 16, 4871 (2025).

6. Teschendorff, A. E. & Horvath, S. Epigenetic ageing clocks: statistical methods and emerging computational challenges. Nat Rev Genet 1–19 (2025) doi:10.1038/s41576-024-00807-w.

7. Wen, J. Refining the generation, interpretation, and application of multi-organ, multi-omics biological aging clocks. Nature Aging (in press) 2025.02.06.25321803 (2025) doi:10.1101/2025.02.06.25321803.

8. Eisenstein, M. Pushing the limits of MRI brain imaging. Nat Methods 21, 1975–1979 (2024).

9. Cole, J. H., Marioni, R. E., Harris, S. E. & Deary, I. J. Brain age and other bodily ‘ages’: implications for neuropsychiatry. Mol Psychiatry 24, 266–281 (2019).

10. Argentieri, M. A. et al. Proteomic aging clock predicts mortality and risk of common age-related diseases in diverse populations. Nat Med 30, 2450–2460 (2024).

11. Mutz, J., Iniesta, R. & Lewis, C. M. Metabolomic age (MileAge) predicts health and life span: A comparison of multiple machine learning algorithms. Science Advances 10, eadp3743 (2024).

12. Boquet-Pujadas, A. et al. MUTATE: a human genetic atlas of multiorgan artificial intelligence endophenotypes using genome-wide association summary statistics. Briefings in Bioinformatics 26, bbaf125 (2025).

13. The MULTI Consortium et al. Brain-heart-eye axis revealed by multi-organ imaging genetics and proteomics. Nature BME (in press) 2025.01.04.25319995 (2025) doi:10.1101/2025.01.04.25319995.

14. Bycroft, C. et al. The UK Biobank resource with deep phenotyping and genomic data. Nature 562, 203–209 (2018).

15. Normal Human Aging: The Baltimore Longitudinal Study of Aging. Journal of Gerontology 40, 767 (1985).

16. Duggan, M. R. et al. Proteomic analyses reveal plasma EFEMP1 and CXCL12 as biomarkers and determinants of neurodegeneration. Alzheimers Dement (2024) doi:10.1002/alz.14142.

17. Sperling, R. A. et al. Trial of Solanezumab in Preclinical Alzheimer’s Disease. New England Journal of Medicine 389, 1096–1107 (2023).

18. Sperling, R. A. et al. Association of Factors With Elevated Amyloid Burden in Clinically Normal Older Individuals. JAMA Neurology 77, 735–745 (2020).

19. Kurki, M. I. et al. FinnGen provides genetic insights from a well-phenotyped isolated population. Nature 613, 508–518 (2023).

20. O’Donovan, M. C. What have we learned from the Psychiatric Genomics Consortium. World Psychiatry 14, 291–293 (2015).

21. de Lange, A.-M. G. & Cole, J. H. Commentary: Correction procedures in brain-age prediction. Neuroimage Clin 26, 102229 (2020).

22. Beheshti, I., Nugent, S., Potvin, O. & Duchesne, S. Bias-adjustment in neuroimaging-based brain age frameworks: A robust scheme. NeuroImage: Clinical 24, 102063 (2019).

23. Szklarczyk, D. et al. The STRING database in 2023: protein-protein association networks and functional enrichment analyses for any sequenced genome of interest. Nucleic Acids Res 51, D638–D646 (2023).

24. Yang, J., Lee, S. H., Goddard, M. E. & Visscher, P. M. GCTA: A Tool for Genome-wide Complex Trait Analysis. Am J Hum Genet 88, 76–82 (2011).

25. Kendler, K. & Neale, M. Endophenotype: a conceptual analysis. Mol Psychiatry 15, 789– 797 (2010).

26. Yi, F. et al. Genetically supported targets and drug repurposing for brain aging: A systematic study in the UK Biobank. Science Advances 11, eadr3757 (2025).

27. Markiewicz, C. J. et al. The OpenNeuro resource for sharing of neuroscience data. eLife 10, e71774 (2021).

28. Gorgolewski, K. et al. Nipype: a flexible, lightweight and extensible neuroimaging data processing framework in python. Front. Neuroinform. 5, 13 (2011).

29. MacGillivray, T. J. et al. Suitability of UK Biobank Retinal Images for Automatic Analysis of Morphometric Properties of the Vasculature. PLoS One 10, e0127914 (2015).

30. Connolly, K. et al. Potential Role of Chitinase-3-like Protein 1 (CHI3L1/YKL-40) in Neurodegeneration and Alzheimer’s Disease. Alzheimers Dement 19, 9–24 (2023).

31. Chiariello, A. et al. The expression pattern of GDF15 in human brain changes during aging and in Alzheimer’s disease. Front Aging Neurosci 14, 1058665 (2023).

32. Beydoun, M. A. et al. GDF15 and its association with cognitive performance over time in a longitudinal study of middle-aged urban adults. Brain, Behavior, and Immunity 108, 340– 349 (2023).

33. Walter, M. Interrelationships Among HDL Metabolism, Aging, and Atherosclerosis. *Arteriosclerosis*, Thrombosis, and Vascular Biology 29, 1244–1250 (2009).

34. Millar, J. S. et al. Impact of age on the metabolism of VLDL, IDL, and LDL apolipoprotein B-100 in men. J Lipid Res 36, 1155–1167 (1995).

35. Li, X. et al. Inflammation and aging: signaling pathways and intervention therapies. Sig Transduct Target Ther 8, 1–29 (2023).

36. Statzer, C., Park, J. Y. C. & Ewald, C. Y. Extracellular Matrix Dynamics as an Emerging yet Understudied Hallmark of Aging and Longevity. Aging Dis 14, 670–693 (2023).

37. Amorim, J. A. et al. Mitochondrial and metabolic dysfunction in ageing and age-related diseases. Nat Rev Endocrinol 18, 243–258 (2022).

38. Lappalainen, T., Li, Y. I., Ramachandran, S. & Gusev, A. Genetic and molecular architecture of complex traits. Cell 187, 1059–1075 (2024).

39. Finucane, H. K. et al. Heritability enrichment of specifically expressed genes identifies disease-relevant tissues and cell types. Nat Genet 50, 621–629 (2018).

40. Rowland, J. et al. Uncovering genetic mechanisms of kidney aging through transcriptomics, genomics, and epigenomics. Kidney Int 95, 624–635 (2019).

41. Allayee, H. et al. Systems genetics approaches for understanding complex traits with relevance for human disease. eLife 12, e91004 (2023).

42. Wen, J. et al. Characterizing Heterogeneity in Neuroimaging, Cognition, Clinical Symptoms, and Genetics Among Patients With Late-Life Depression. JAMA Psychiatry (2022) doi:10.1001/jamapsychiatry.2022.0020.

43. Mannick, J. B. et al. mTOR inhibition improves immune function in the elderly. Sci Transl Med 6, 268ra179 (2014).

44. Blagosklonny, M. V. Rejuvenating immunity: “anti-aging drug today” eight years later. Oncotarget 6, 19405–19412 (2015).

45. Franceschi, C., Garagnani, P., Parini, P., Giuliani, C. & Santoro, A. Inflammaging: a new immune–metabolic viewpoint for age-related diseases. Nat Rev Endocrinol 14, 576–590 (2018).

46. Barcena, M. L., Aslam, M., Pozdniakova, S., Norman, K. & Ladilov, Y. Cardiovascular Inflammaging: Mechanisms and Translational Aspects. Cells 11, 1010 (2022).

47. Kosyreva, A. M., Sentyabreva, A. V., Tsvetkov, I. S. & Makarova, O. V. Alzheimer’s Disease and Inflammaging. Brain Sci 12, 1237 (2022).

48. Harrison, D. E. et al. Rapamycin fed late in life extends lifespan in genetically heterogeneous mice. Nature 460, 392–395 (2009).

49. Atamna, H., Tenore, A., Lui, F. & Dhahbi, J. M. Organ Reserve, Excess Metabolic Capacity, and Aging. Biogerontology 19, 171–184 (2018).

50. Argentieri, M. A. et al. Integrating the environmental and genetic architectures of aging and mortality. Nat Med 1–10 (2025) doi:10.1038/s41591-024-03483-9.

51. Mbatchou, J. et al. Computationally efficient whole-genome regression for quantitative and binary traits. Nat Genet 53, 1097–1103 (2021).

52. Wen, J. et al. Genomic loci influence patterns of structural covariance in the human brain. Proceedings of the National Academy of Sciences 120, e2300842120 (2023).

53. Bai, W. et al. A population-based phenome-wide association study of cardiac and aortic structure and function. Nat Med 26, 1654–1662 (2020).

54. Mojtahed, A. et al. Reference range of liver corrected T1 values in a population at low risk for fatty liver disease—a UK Biobank sub-study, with an appendix of interesting cases. Abdom Radiol 44, 72–84 (2019).

55. Whitcher, B. et al. Precision MRI phenotyping enables detection of small changes in body composition for longitudinal cohorts. Sci Rep 12, 3748 (2022).

56. Parisinos, C. A. et al. Genome-wide and Mendelian randomisation studies of liver MRI yield insights into the pathogenesis of steatohepatitis. J Hepatol 73, 241–251 (2020).

57. Karlsson, A. et al. Automatic and quantitative assessment of regional muscle volume by multi-atlas segmentation using whole-body water-fat MRI. J Magn Reson Imaging 41, 1558–1569 (2015).

58. Borga, M. et al. Validation of a fast method for quantification of intra-abdominal and subcutaneous adipose tissue for large-scale human studies. NMR Biomed 28, 1747–1753 (2015).

59. Liu, Y. et al. Genetic architecture of 11 organ traits derived from abdominal MRI using deep learning. eLife 10, e65554 (2021).

60. Wen, J. et al. Convolutional neural networks for classification of Alzheimer’s disease: Overview and reproducible evaluation. Medical Image Analysis 63, 101694 (2020).

61. Li, W. UK Biobank pharma proteomics resource. Nat Genet 55, 1781–1781 (2023).

62. Sun, B. B. et al. Plasma proteomic associations with genetics and health in the UK Biobank. Nature 622, 329–338 (2023).

63. Würtz, P. et al. Quantitative Serum Nuclear Magnetic Resonance Metabolomics in Large-Scale Epidemiology: A Primer on-Omic Technologies. American Journal of Epidemiology 186, 1084–1096 (2017).

64. Ritchie, S. C. et al. Quality control and removal of technical variation of NMR metabolic biomarker data in ∼120,000 UK Biobank participants. Sci Data 10, 64 (2023).

65. Manichaikul, A. et al. Robust relationship inference in genome-wide association studies. Bioinformatics 26, 2867–2873 (2010).

66. Jiang, L. et al. A resource-efficient tool for mixed model association analysis of large-scale data. Nat Genet 51, 1749–1755 (2019).

67. Price, A. L., Zaitlen, N. A., Reich, D. & Patterson, N. New approaches to population stratification in genome-wide association studies. Nat Rev Genet 11, 459–463 (2010).

68. Abraham, G., Qiu, Y. & Inouye, M. FlashPCA2: principal component analysis of Biobank-scale genotype datasets. Bioinformatics 33, 2776–2778 (2017).

69. Wen, J. et al. Genetic and clinical correlates of two neuroanatomical AI dimensions in the Alzheimer’s disease continuum. Transl Psychiatry 14, 1–14 (2024).

70. Wen, J. Multiorgan biological age shows that no organ system is an island. Nat Aging 1–2 (2024) doi:10.1038/s43587-024-00690-4.

71. Watanabe, K., Taskesen, E., van Bochoven, A. & Posthuma, D. Functional mapping and annotation of genetic associations with FUMA. Nat Commun 8, 1826 (2017).

72. Finucane, H. K. et al. Partitioning heritability by functional annotation using genome-wide association summary statistics. Nat Genet 47, 1228–1235 (2015).

73. Cahoy, J. D. et al. A Transcriptome Database for Astrocytes, Neurons, and Oligodendrocytes: A New Resource for Understanding Brain Development and Function. J. Neurosci. 28, 264–278 (2008).

74. Bernstein, B. E. et al. The NIH Roadmap Epigenomics Mapping Consortium. Nat Biotechnol 28, 1045–1048 (2010).

75. Dunham, I. et al. An integrated encyclopedia of DNA elements in the human genome. Nature 489, 57–74 (2012).

76. The Genotype-Tissue Expression (GTEx) project. Nat Genet 45, 580–585 (2013).

77. Bulik-Sullivan, B. K. et al. LD Score regression distinguishes confounding from polygenicity in genome-wide association studies. Nat Genet 47, 291–295 (2015).

78. Bulik-Sullivan, B. et al. An atlas of genetic correlations across human diseases and traits. Nat Genet 47, 1236–1241 (2015).

79. Hemani, G. et al. The MR-Base platform supports systematic causal inference across the human phenome. eLife 7, e34408 (2018).

80. Skrivankova, V. W. et al. Strengthening the Reporting of Observational Studies in Epidemiology Using Mendelian Randomization: The STROBE-MR Statement. JAMA 326, 1614–1621 (2021).

81. Sanderson, E. et al. Mendelian randomization. Nat Rev Methods Primers 2, 1–21 (2022).

82. Bowden, J. et al. A framework for the investigation of pleiotropy in two-sample summary data Mendelian randomization. Stat Med 36, 1783–1802 (2017).

83. Ge, T., Chen, C.-Y., Ni, Y., Feng, Y.-C. A. & Smoller, J. W. Polygenic prediction via Bayesian regression and continuous shrinkage priors. Nat Commun 10, 1776 (2019).

84. Qi, T., Song, L., Guo, Y., Chen, C. & Yang, J. From genetic associations to genes: methods, applications, and challenges. Trends in Genetics 40, 642–667 (2024).

85. Allesøe, R. L. et al. Discovery of drug–omics associations in type 2 diabetes with generative deep-learning models. Nat Biotechnol 41, 399–408 (2023).

86. Giambartolomei, C. et al. Bayesian Test for Colocalisation between Pairs of Genetic Association Studies Using Summary Statistics. PLOS Genetics 10, e1004383 (2014).

87. Donohue, M. C. et al. Natural cubic splines for the analysis of Alzheimer’s clinical trials. Pharmaceutical Statistics 22, 508–519 (2023).

88. Wen, J. MEDICINE: AI-derived GWAS summary statistics sharing portal. (2025) doi: 10.7303/syn64923248.

89. Wen, J. MLNI (Version 0.1.4) [Computer software]. (2025) doi: https://github.com/anbai106/mlni.

