## Supplementary materials for "Multi-organ MRI digitizes biological aging clocks across proteomics, metabolomics, and genetics"

### Online Supplementary Materials

**eNote 1: MRI features to derive the 7 MRIBAGs**
**eNote 2: Additional prediction power of the 7 MRIBAGS on top of calendar age and sex**
**eNote 3: Multicollinearity among adipose and kidney imaging features resulted in poor generalization to independent test datasets**
**eNote 4: Generalizability of the brain MRIBAG to external datasets**
**eNote 5: Sex difference in the 7 MRIBAGs**
**eNote 6: Feature importance in deriving the 7 MRIBAGs and their biological interpretation with underlying imaging features**
**eNote 7: ProWAS results for the brain MRIBAG using UKBB Olink data and its comparison with BLSA SomaScan data**
**eNote 8: Sex difference in ProWAS**
**eNote 9: Metabolite-set enrichment analysis for small-molecule from the UK Biobank NMR metabolomics data**
**eNote 10: Sex difference in MetWAS**
**eNote 11: The definition of genomic loci, independent significant SNP, lead SNP, candidate SNP**
**eNote 12: Sensitivity check analyses for the GWAS robustness**
**eNote 13: Sex difference in GWAS**
**eNote 14: Additional sensitivity test analysis of spleen and liver MRIBAG's protective effect on all-cause mortality**
**eNote 15: Further methodological improvements in the age prediction performance of abdominal MRI BAGs**
**eFigure 1: Model performance using the elastic net regression and the neural network**
**eFigure 2: Incremental $R^2$ of the 7 MRIBAGs to predict 14 systematic disease categories and age at mortality on top of age and sex**
**eFigure 3: Evidence for model overfitting and poor generalizability to independent test dataset due to feature multicollinearity**
**eFigure 4: Generalizability of the brain MRIBAG from UKBB to A4**
**eFigure 5: Comparison between UKBB Olink ProWAS and BLSA SomaScan ProWAS for the common significant proteins**
**eFigure 6: Sex difference for the 7 MRIBAGs**
**eFigure 7: SHAP values to denote feature importance for generating the 7 MRIBAGs**
**eFigure 8: ProWAS for the left thalamus and protein-set enrichment analysis (PSEA) for the significant proteins**
**eFigure 9: AD drug differential responses using the brain MRI derived i) by directly applying the pretrained model to A4 vs. ii) by independently training the model in A4**
**eFigure 10: Functional protein set enrichment analysis (PSEA) and protein-protein interaction (PPI) network**
**eFigure 11: Sex-specific ProWAS results**
**eFigure 12: Genetic correlation between the 7 MRIBAGs and 2923 proteins and 327 metabolites**
**eFigure 13: Sex-specific MetWAS results**
**eFigure 14: sex-specific GWAS analyses**
**eFigure 15: Genetic correlation and phenotypic correlation between the 7 MRIBAGs**
**eFigure 16: Sensitivity check analyses for the potential causal signal in our MR analyses**

**eFigure 17: Conceptualization of the endophenotype hypothesis along the multi-scale causal pathway of human aging and disease**
**eFigure 18: Evidence of drug indications for 4 prioritized genes**
**eFigure 19: The relationships among the liver MRIBAG, age at mortality, and liver volume (ID: 21080)**
**eFigure 20: The relationships among the spleen MRIBAG, age at mortality, and spleen volume (ID: 21083)**
**eFigure 21: Mediation pathways between the brain ProtBAG, brain MRIBAG, and two subtypes of late-life depression**
**eFigure 22: Population selections and the nested cross-validation procedure to derive the 7 MRIBAGs**
**eFigure 23: Feature importance in deriving the brain MRIBAG in the UK Biobank, BLSA, and A4**
**eTable 1: The characteristics of participants consolidated via the MULTI consortium**
**eTable 2: Biological age prediction performance before age bias correction**
**eTable 3: The description of the 327 metabolites from the UKBB NMR platform**
**eTable 4: Genomic loci of the 7 MRIBAG GWASs**
**eTable 5: The detailed statistics of SNP-based heritability estimates via GCTA**
**eTable 6: The incremental R2 explained by the MRIBAG-PRS in split2 GWAS**
**eTable 7: Colocalization analyses between the 7 MRIBAGs and 11 ProtBAGs, and 5 MetBAGs**
**eTable 8: Results for the survival analyses for predicting all-cause mortality**
**eTable 9: Results for the mediation analysis between brain ProtBAG, brain MRIBAG, and LLD subtypes**
**eTable 10: Concordant hits for the sex-stratified ProWAS analyses**
**eTable 11: Discordant hits for the sex-stratified MetWAS analyses**

**eNote 1: MRI features to derive the 7 MRIBAGs**

**Brain MRIBAG features** derived from the MUSE pipeline[1]: Right Accumbens Area, Left Accumbens Area, Right Amygdala, Left Amygdala, Right Caudate, Left Caudate, Right Cerebellum Exterior, Left Cerebellum Exterior, Right Hippocampus, Left Hippocampus, Right Pallidum, Left Pallidum, Right Putamen, Left Putamen, Right Thalamus Proper, Left Thalamus Proper, Cerebellar Vermal Lobules I-V, Cerebellar Vermal Lobules VI-VII, Cerebellar Vermal Lobules VIII-X, Left Basal Forebrain, Right Basal Forebrain, Right ACgG (anterior cingulate gyrus), Left ACgG (anterior cingulate gyrus), Right AIns (anterior insula), Left AIns (anterior insula), Right AOrG (anterior orbital gyrus), Left AOrG (anterior orbital gyrus), Right AnG (angular gyrus), Left AnG (angular gyrus), Right Calc (calcarine cortex), Left Calc (calcarine cortex), Right CO (central operculum), Left CO (central operculum), Right Cun (cuneus), Left Cun (cuneus), Right Ent (entorhinal area), Left Ent (entorhinal area), Right FO (frontal operculum), Left FO (frontal operculum), Right FRP (frontal pole), Left FRP (frontal pole), Right FuG (fusiform gyrus), Left FuG (fusiform gyrus), Right GRe (gyrus rectus), Left GRe (gyrus rectus), Right IOG (inferior occipital gyrus), Left IOG (inferior occipital gyrus), Right ITG (inferior temporal gyrus), Left ITG (inferior temporal gyrus), Right LiG (lingual gyrus), Left LiG (lingual gyrus), Right LOrG (lateral orbital gyrus), Left LOrG (lateral orbital gyrus), Right MCgG (middle cingulate gyrus), Left MCgG (middle cingulate gyrus), Right MFC (medial frontal cortex), Left MFC (medial frontal cortex), Right MFG (middle frontal gyrus), Left MFG (middle frontal gyrus), Right MOG (middle occipital gyrus), Left MOG (middle occipital gyrus), Right MOrG (medial orbital gyrus), Left MOrG (medial orbital gyrus), Right MPoG (postcentral gyrus medial segment), Left MPoG (postcentral gyrus medial segment), Right MPrG (precentral gyrus medial segment), Left MPrG (precentral gyrus medial segment), Right MSFG (superior frontal gyrus medial segment), Left MSFG (superior frontal gyrus medial segment), Right MTG (middle temporal gyrus), Left MTG (middle temporal gyrus), Right OCP (occipital pole), Left OCP (occipital pole), Right OFuG (occipital fusiform gyrus), Left OFuG (occipital fusiform gyrus), Right OpIFG (opercular part of the inferior frontal gyrus), Left OpIFG (opercular part of the inferior frontal gyrus), Right OrIFG (orbital part of the inferior frontal gyrus), Left OrIFG (orbital part of the inferior frontal gyrus), Right PCgG (posterior cingulate gyrus), Left PCgG (posterior cingulate gyrus), Right PCu (precuneus), Left PCu (precuneus), Right PHG (parahippocampal gyrus), Left PHG (parahippocampal gyrus), Right PIns (posterior insula), Left PIns (posterior insula), Right PO (parietal operculum), Left PO (parietal operculum), Right PoG (postcentral gyrus), Left PoG (postcentral gyrus), Right POrG (posterior orbital gyrus), Left POrG (posterior orbital gyrus), Right PP (planum polare), Left PP (planum polare), Right PrG (precentral gyrus), Left PrG (precentral gyrus), Right PT (planum temporale), Left PT (planum temporale), Right SCA (subcallosal area), Left SCA (subcallosal area), Right SFG (superior frontal gyrus), Left SFG (superior frontal gyrus), Right SMC (supplementary motor cortex), Left SMC (supplementary motor cortex), Right SMG (supramarginal gyrus), Left SMG (supramarginal gyrus), Right SOG (superior occipital gyrus), Left SOG (superior occipital gyrus), Right SPL (superior parietal lobule), Left SPL (superior parietal lobule), Right STG (superior temporal gyrus), Left STG (superior temporal gyrus), Right TMP (temporal pole), Left TMP (temporal pole), Right TrIFG (triangular part of the inferior frontal gyrus), Left TrIFG (triangular part of the inferior frontal gyrus), Right TTG (transverse temporal gyrus), Left TTG (transverse temporal gyrus).

**Heart MRIBAG features**[2]: 'lv_end_diastolic_volume_f24100_2_0', 'lv_end_systolic_volume_f24101_2_0', 'lv_stroke_volume_f24102_2_0', 'lv_ejection_fraction_f24103_2_0', 'lv_cardiac_output_f24104_2_0', 'lv_myocardial_mass_f24105_2_0', 'rv_end_diastolic_volume_f24106_2_0', 'rv_end_systolic_volume_f24107_2_0', 'rv_stroke_volume_f24108_2_0', 'rv_ejection_fraction_f24109_2_0', 'la_maximum_volume_f24110_2_0', 'la_minimum_volume_f24111_2_0', 'la_stroke_volume_f24112_2_0', 'la_ejection_fraction_f24113_2_0', 'ra_maximum_volume_f24114_2_0', 'ra_minimum_volume_f24115_2_0', 'ra_stroke_volume_f24116_2_0', 'ra_ejection_fraction_f24117_2_0', 'ascending_aorta_maximum_area_f24118_2_0', 'ascending_aorta_minimum_area_f24119_2_0', 'descending_aorta_maximum_area_f24121_2_0', 'descending_aorta_minimum_area_f24122_2_0', 'lv_mean_myocardial_wall_thickness_aha_1_f24124_2_0', 'lv_mean_myocardial_wall_thickness_aha_2_f24125_2_0', 'lv_mean_myocardial_wall_thickness_aha_3_f24126_2_0', 'lv_mean_myocardial_wall_thickness_aha_4_f24127_2_0', 'lv_mean_myocardial_wall_thickness_aha_5_f24128_2_0', 'lv_mean_myocardial_wall_thickness_aha_6_f24129_2_0', 'lv_mean_myocardial_wall_thickness_aha_7_f24130_2_0', 'lv_mean_myocardial_wall_thickness_aha_8_f24131_2_0', 'lv_mean_myocardial_wall_thickness_aha_9_f24132_2_0', 'lv_mean_myocardial_wall_thickness_aha_10_f24133_2_0', 'lv_mean_myocardial_wall_thickness_aha_11_f24134_2_0', 'lv_mean_myocardial_wall_thickness_aha_12_f24135_2_0', 'lv_mean_myocardial_wall_thickness_aha_13_f24136_2_0', 'lv_mean_myocardial_wall_thickness_aha_14_f24137_2_0', 'lv_mean_myocardial_wall_thickness_aha_15_f24138_2_0', 'lv_mean_myocardial_wall_thickness_aha_16_f24139_2_0', 'lv_mean_myocardial_wall_thickness_global_f24140_2_0', 'lv_circumferential_strain_aha_1_f24141_2_0', 'lv_circumferential_strain_aha_2_f24142_2_0', 'lv_circumferential_strain_aha_3_f24143_2_0', 'lv_circumferential_strain_aha_4_f24144_2_0', 'lv_circumferential_strain_aha_5_f24145_2_0', 'lv_circumferential_strain_aha_6_f24146_2_0', 'lv_circumferential_strain_aha_7_f24147_2_0', 'lv_circumferential_strain_aha_8_f24148_2_0', 'lv_circumferential_strain_aha_9_f24149_2_0', 'lv_circumferential_strain_aha_10_f24150_2_0', 'lv_circumferential_strain_aha_11_f24151_2_0', 'lv_circumferential_strain_aha_12_f24152_2_0', 'lv_circumferential_strain_aha_13_f24153_2_0', 'lv_circumferential_strain_aha_14_f24154_2_0', 'lv_circumferential_strain_aha_15_f24155_2_0', 'lv_circumferential_strain_aha_16_f24156_2_0', 'lv_circumferential_strain_global_f24157_2_0', 'lv_radial_strain_aha_1_f24158_2_0', 'lv_radial_strain_aha_2_f24159_2_0', 'lv_radial_strain_aha_3_f24160_2_0', 'lv_radial_strain_aha_4_f24161_2_0', 'lv_radial_strain_aha_5_f24162_2_0', 'lv_radial_strain_aha_6_f24163_2_0', 'lv_radial_strain_aha_7_f24164_2_0', 'lv_radial_strain_aha_8_f24165_2_0', 'lv_radial_strain_aha_9_f24166_2_0', 'lv_radial_strain_aha_10_f24167_2_0', 'lv_radial_strain_aha_11_f24168_2_0', 'lv_radial_strain_aha_12_f24169_2_0', 'lv_radial_strain_aha_13_f24170_2_0', 'lv_radial_strain_aha_14_f24171_2_0', 'lv_radial_strain_aha_15_f24172_2_0', 'lv_radial_strain_aha_16_f24173_2_0', 'lv_radial_strain_global_f24174_2_0', 'lv_longitudinal_strain_segment_1_f24175_2_0', 'lv_longitudinal_strain_segment_2_f24176_2_0', 'lv_longitudinal_strain_segment_3_f24177_2_0', 'lv_longitudinal_strain_segment_4_f24178_2_0', 'lv_longitudinal_strain_segment_5_f24179_2_0', 'lv_longitudinal_strain_segment_6_f24180_2_0', 'lv_longitudinal_strain_global_f24181_2_0'.

**Adipose MRIBAG features**:
'Visceral_fat_volume_21085-2.0' (PMID: 34128465, 35568031),
'Pancreas_PDFF_(fat_fraction)_21090-2.0' (PMID: 34128465, 35568031),
'Anterior_thigh_fat-free_muscle_volume_(right)_22403-2.0' (PMID: 25111561, 26768490, 27662190, 29785727, 32519807),
'Posterior_thigh_fat-free_muscle_volume_(right)_22404-2.0' (PMID: 25111561, 26768490, 27662190, 29785727, 32519807),
'Anterior_thigh_fat-free_muscle_volume_(left)_22405-2.0' (PMID: 25111561, 26768490, 27662190, 29785727, 32519807),
'Posterior_thigh_fat-free_muscle_volume_(left)_22406-2.0' (PMID: 25111561, 26768490, 27662190, 29785727, 32519807),
'Abdominal_subcutaneous_adipose_tissue_volume_(ASAT)_22408-2.0' (PMID: 25111561, 26768490, 27662190, 29785727, 32519807),
'Total_trunk_fat_volume_22410-2.0' (PMID: 25111561, 26768490, 27662190, 29785727, 32519807),
'Total_abdominal_adipose_tissue_index_22432-2.0' (PMID: 25111561, 26768490, 27662190, 29785727, 32519807),
'Abdominal_fat_ratio_22434-2.0' (PMID: 25111561, 26768490, 27662190, 29785727, 32519807),
'Muscle_fat_infiltration_22435-2.0' (PMID: 25111561, 26768490, 27662190, 29785727, 32519807),
'Posterior_thigh_muscle_fat_infiltration_(MFI)_(left)_23355-2.0' (PMID: 25111561, 26768490, 27662190, 29785727, 32519807),
'Posterior_thigh_muscle_fat_infiltration_(MFI)_(right)_23356-2.0' (PMID: 25111561, 26768490, 27662190, 29785727, 32519807),
'Anterior_thigh_muscle_fat_infiltration_(MFI)_(left)_24353-2.0' (PMID: 25111561, 26768490, 27662190, 29785727, 32519807),
'Anterior_thigh_muscle_fat_infiltration_(MFI)_(right)_24354-2.0' (PMID: 25111561, 26768490, 27662190, 29785727, 32519807),
'Proton_density_fat_fraction_(PDFF)_40061-2.0' (PMID: 28241076, 30032383).

Removed features due to overfitting:
'Visceral_adipose_tissue_volume_(VAT)_22407-2.0' (PMID: 25111561, 26768490, 27662190, 29785727, 32519807),
'Subcutaneous_fat_volume_21086-2.0' (PMID: 34128465, 35568031),
'Liver_PDFF_(fat_fraction)_21088-2.0' (PMID: 34128465, 35568031),

'Total_thigh_fat-free_muscle_volume_22409-2.0' (PMID: 25111561, 26768490, 27662190, 29785727, 32519807).

**Liver MRIBAG features**:
'Liver_volume_21080-2.0' (PMID: 34128465, 35568031),
'Liver_PDFF_(fat_fraction)_21088-2.0' (PMID: 34128465, 35568031),
'Liver_iron_21089-2.0' (PMID: 34128465, 35568031),
'Liver_iron_corrected_T1_(ct1)_40062-2.0' (PMID: 28241076, 30032383).

Removed features due to high collinearity, although no severe overfitting was observed:
'Proton_density_fat_fraction_(PDFF)_40061-2.0' (PMID: 28241076, 30032383),
'Liver_iron_(Fe)_40060-2.0' (PMID: 28241076, 30032383),
'FR_liver_PDFF_mean_24352-2.0' (PMID: 25111561, 26768490, 27662190, 29785727, 32519807).

**Spleen MRIBAG features**:
'Spleen_volume_21083-2.0' (PMID: 34128465, 35568031),
'Spleen_iron_-_IDEAL_21170-2.0' (PMID: 34128465, 35568031),
'Spleen_iron_-_protocol_normalised_21173-2.0' (PMID: 34128465, 35568031).

**Kidney MRIBAG features**:
'Left_kidney_volume_21081-2.0' (PMID: 34128465, 35568031),
'Kidney_parenchyma_(right)_21162-2.0' (PMID: 33262432),
'Kidney_distance_21163-2.0' (PMID: 33262432).

Removed features due to overfitting:
'Kidney_parenchyma_(left)_21161-2.0' (PMID: 33262432),
'Right_kidney_volume_21082-2.0' (PMID: 34128465, 35568031),
'Kidney_parenchyma_total_21160-2.0' (PMID: 33262432).

**Pancreas MRIBAG features**:
'Pancreas_volume_21087-2.0' (PMID: 34128465, 35568031),
'Pancreas_PDFF_(fat_fraction)_21090-2.0' (PMID: 34128465, 35568031),
'Pancreas_iron_21091-2.0' (PMID: 34128465, 35568031).

**eNote 2: Additional prediction power of the 7 MRIBAGS on top of calendar age and sex**

A critical question that remains is whether an age prediction model (e.g., spleen MRIBAG) that predicts poorly for chronological age can still offer meaningful insights into biological aging beyond chronological age. **eFigure 2** illustrates the incremental $R^2$ contribution of the 7 MRIBAGs, showing a 1.43% increase for the category of all mental and behavioral disorders and a 0.39% increase for predicting the age at mortality in explaining additional variance beyond chronological age and sex. While these increments appeared small, they could still capture clinically related aspects of aging biology beyond chronological age (e.g., predicting age-related outcomes[3]).

**eNote 3: Multicollinearity among adipose and kidney imaging features resulted in poor generalization to independent test datasets**

We found that multicollinearity among imaging features, particularly in adipose and kidney MRIBAGs, led to severe overfitting and poor generalizability.

Correlation heatmaps revealed strong multicollinearity (Pearson's $r$ >0.9) in these organs, which were also the only ones exhibiting clear overfitting when the full set of imaging features was used. To address this, we systematically removed highly correlated features: when a feature was strongly correlated with multiple others, it was preferentially excluded; if only two features exhibited high collinearity, one was randomly removed.

After feature removal, overfitting was resolved in both organs using both Lasso and Linear SVR methods. For instance, in adipose tissue analyzed with Lasso, the independent dataset MAE decreased from 6.47 to 5.74 years, while training MAE remained stable (4.89–4.94). Similar improvements were observed with Linear SVR. **eFigure 3** shows correlation heatmaps for adipose tissue and kidney, highlighting severe multicollinearity before feature removal. Additionally, **eFigure 3** presents box plots comparing training and independent dataset MAEs for both organs before and after feature removal, demonstrating enhanced generalization and reduced overfitting following the elimination of highly collinear features.

**eNote 4: Generalizability of the brain MRIBAG to external datasets**

We evaluated the generalizability of the brain MRIBAG model trained on UKBB by testing it on an external dataset with differing demographics, specifically the A4 study – a preclinical Alzheimer's disease cohort comprising cognitively unimpaired individuals.

As illustrated in **eFigure 4**, applying the pre-trained UKBB model to A4 data revealed a noticeable domain shift, indicating poor generalizability. However, age bias correction mitigated this effect to some extent. Furthermore, we found that independently training an age prediction model within the A4 study resulted in brain MRIBAG estimates that exhibited significant correlations with those obtained by directly applying the UKBB-trained model to A4. More importantly, we demonstrated that whether the age prediction model was trained independently within the A4 study or the brain MRIBAG was derived by directly applying the pre-trained model, the conclusions from the AD drug analyses (**Fig. 6c**) remained consistent, despite slight differences in P-value significance (**eFigure 5**).

Considering all the results and the observed domain shift, we used the brain MRIBAG derived from the independently trained model in the external datasets for all subsequent analyses in the main text. **eFigure 23** demonstrates that the most influential imaging features contributing to brain MRIBAGs are largely consistent across the UK Biobank, A4, and BLSA cohorts, although some study-specific brain volume patterns are also evident.

**eNote 5: Sex difference in the 7 MRIBAGs**

We assessed sex differences across the 7 MRIBAGs and observed the most pronounced disparities in the adipose tissue, heart, liver, and kidney (**eFigure 6**). Specifically, females exhibited higher BAGs in the adipose, heart, and liver, while males showed a higher BAG in the kidney.

The observed sex differences in MRIBAGs likely reflect underlying biological distinctions in organ aging patterns. Females showed higher BAGs in adipose tissue, heart, and liver, which may be driven by hormonal influences such as estrogen that affect fat distribution, myocardial remodeling, and hepatic metabolism, especially post-menopause. In contrast, males exhibited higher BAG in the kidney, potentially due to faster age-related decline in renal function and greater baseline kidney mass. These organ-specific aging trajectories align with known sex-based differences in physiology, metabolism, and immune response.

Sex differences in the biological aging clock have been previously demonstrated. For instance, Moguilner et al.[4] investigated brain aging clocks derived from EEG data across diverse global populations, analyzing 5,306 participants from 15 countries, including both Latin American and Caribbean (LAC) and non-LAC regions. Regarding sex differences, the study found that females in LAC regions exhibited larger brain-age gaps than males in both the control and AD groups, using a model trained on data from both sexes. More recently, Argentieri et al.[5] examined sex effects on proteome-based aging clocks using UK Biobank data. They reported nearly identical age prediction accuracy between sex-specific models and a combined-sex model, with predicted ages from each approach being highly correlated. Interestingly, the female-specific model demonstrated slightly better performance (MAE = 2.25) compared to the male-specific model (MAE = 2.45). Both models leveraged over 2,000 proteins and were not organ-specific, with substantial overlap observed in the top predictive proteins across sexes.

**eNote 6: Feature importance in deriving the 7 MRIBAGs and their biological interpretation with underlying imaging features**

We used SHAP analysis to investigate which imaging features contained more derived MRIBAG content in a post hoc manner. Overall, SHAP values interpret both the direction of a feature's effect (positive or negative) and how feature magnitude (high or low) relates to predicted biological aging. A positive SHAP value indicates that the feature contributes to increasing the predicted MRIBAG, suggesting that it is associated with an older-appearing organ relative to chronological age. In contrast, a negative SHAP value indicates that the feature lowers the predicted MRIBAG, implying a younger-appearing organ. These values help interpret how individual features drive organ-specific biological aging predictions in the model (**eFigure 7**).

We interpreted these features at the level of raw imaging features. For example, for brain MRIBAG, we found that left thalamus, right caudate nucleus volume, among others, were the most influential features that negatively affected brain MRIBAG. The thalamus acts as a relay center, transmitting sensory and motor signals to the cerebral cortex and playing a critical role in cognition, consciousness, and sleep regulation. Thalamic volume decline has been linked to impaired cognitive processing speed and executive function in older adults, making it a sensitive marker of brain aging. For the heart MRIBAG, aorta volume is a key feature in heart MRIBAG because it reflects vascular aging, which involves aortic dilation, stiffening, and wall thickening with age. These changes are strongly associated with increased cardiovascular risk, reduced arterial compliance, and are well-documented markers of both chronological and biological aging of the cardiovascular system.

Another approach to validate these associations with imaging features is through a proteome-wide association study (ProWAS) followed by protein-set enrichment analysis (PSEA), which can help identify biological pathways linked to specific organ imaging traits. For instance, we applied this method to the left thalamus volume as a case study (**eFigure 8**). We identified 17 proteins most strongly associated with this brain imaging feature, among which four were classified as brain-enriched, showing at least 4-fold higher expression in brain tissue compared to other tissues, including MOG, BCAN, and NCAN. We then took the 17 significant proteins as an input protein set to denote implicated biological pathways using the SMART database via the STRING platform. The biological pathways associated with brain MRIBAG, as highlighted by protein domains, point to key mechanisms involved in structural and functional decline. Proteins with link domains are involved in hyaluronan and extracellular matrix (ECM) binding, contributing to ECM remodeling, gliosis, and neuroinflammation – hallmarks of aging and neurodegeneration. EGF-like domains mediate cell signaling processes critical for neurogenesis, myelination, and cognitive plasticity. Trypsin-like protease domains facilitate proteolysis, including amyloid precursor protein (APP) cleavage, which is linked to neuroinflammation and synaptic pruning. Finally, immunoglobulin domains play roles in cell adhesion and immune function, influencing neuron-glia interactions, microglial activity, and blood-brain barrier (BBB) integrity, all of which are central to the aging brain's vulnerability.

**eNote 7: ProWAS results for the brain MRIBAG using UKBB Olink data and its comparison with BLSA SomaScan data**

We compared ProWAS signals using UKBB Olink and BLSA SomaScan protein data. Given potential domain shifts observed in **eFigure 4**, we first retrained the brain MRIBAG independently in the BLSA study.

SomaScan proteins in BLSA demonstrated sensitivity in linking with brain MRIBAG, despite the smaller sample size ($N = 909$) compared to UKBB Olink proteins ($N$ ~4000). Using a significance threshold of P-value < 0.05/2139 for the common proteins present in both platforms, we identified 19 significant MRIBAG-protein associations in SomaScan and 36 in UKBB Olink. However, the $\beta$ coefficient estimates from the two platforms were weakly correlated (Pearson's $r$ = 0.10, P-value = $1.62 \times 10^{-6}$). This aligns with prior findings by Eldjarn et al.[6], who reported a median Spearman correlation of 0.33 between plasma levels of 1848 proteins measured with both Olink and SomaScan assays in an Icelandic cohort of 1514 individuals. They also noted a higher coefficient of variation (CV) in Olink compared to SomaScan.

As shown in **eFigure 9**, the effect directions of the most significant proteins remained consistent across platforms, except for UXS1. Several factors may explain these discrepancies. First, SomaScan and Olink rely on distinct technologies; SomaScan uses aptamer-based binding, whereas Olink employs proximity extension assays, potentially leading to differences in protein quantification and associations with disease endpoints. Second, SomaScan has a broader protein coverage and is known for its higher sensitivity, which may affect $\beta$ coefficient estimates depending on sample characteristics and disease stage. Notably, our $\beta$ coefficient correlation was derived from two distinct populations (UKBB vs. BLSA) with clear demographic differences. Third, proteins may exhibit context-specific roles in different diseases or phenotypes, which may be differentially captured by the platforms. Lastly, biological complexity and technical artifacts could also contribute to these variations. Further validation is necessary to determine whether the observed differences in β coefficients reflect true biological variation or platform-specific effects.

**eNote 8: Sex difference in ProWAS**

We conducted sex-stratified ProWAS analyses separately for males and females, which have similar sample sizes. Based on the results, we categorized proteins as “Discordant”, “Concordant”, or “Not Significant” using the following criteria:

**Discordant hit**:

- At least one sex shows a statistically significant association (P-value<0.05/2923/0.05);
- The direction of effect differs between sexes (sign(female) ≠ sign(male)) for the standardized beta values.

**Concordant hit**:

- At least one sex shows a statistically significant association (P-value<0.05/2923/0.05);
- The direction of effect is the same between sexes (sign(female) = sign(male)) for the standardized beta values.

**Non-significant hit**:

- Neither sex shows a statistically significant association (P-value>0.05/2923/0.05)

With this definition, we found all the 411 significant MRIBAG-protein pairs to be concordant hits (**eFigure 11**). Among the 7 MRIBAGs, we identified the following numbers of concordant proteins shared between males and females: adipose (16), brain (7), kidney (235), liver (38), pancreas (19), and spleen (96) (**eTable 10**). Among the 411 protein associations demonstrating concordant effect directions across sexes, a notable number of concordant hits were observed across all 7 MRIBAGs, with particularly strong representation in the kidney MRIBAG. These concordant associations indicate that, despite some sex-specific significance differences, the directionality of protein-BAG relationships remains consistent between males and females, suggesting a shared biological aging trajectory modulated by these proteins. For instance, proteins such as TGFBR2, TNFRSF1A (e.g., $\beta$ =0.23 for females and 0.22 for males), and CD59 showed robust positive associations with kidney MRIBAG in both sexes. This concordance reinforces the reliability of these proteomic markers as sex-agnostic indicators of kidney-related brain aging processes and supports their relevance as potential targets for interventions aimed at mitigating organ-specific biological aging. More broadly, the prevalence of concordant hits across multiple BAGs underscores the presence of core molecular signatures that transcend sex while still allowing room for sex-differentiated regulation or thresholds of statistical detection.

At the ProtBAG level, a recent study by Argentieri et al.[5] investigated the influence of sex on proteome-based aging clocks (ProtBAG) using data from the UK Biobank. They observed comparable age prediction performance between sex-specific models and a combined-sex model, with predicted ages from both approaches showing near-perfect correlation. Interestingly, the female-only model demonstrated a slightly better fit (MAE = 2.25) than the male-only model (MAE = 2.45). Both models leveraged over 2,000 proteins and were not tailored to specific organs. In our previous ProtBAG study[7], we further showed clear sex differences in brain ProtBAG, and male-specific models are more susceptible to overfitting than female-specific models. Although the age predicted by sex-specific models and models trained on combined data is highly consistent (i.e., correlated), it is critical to address issues such as model overfitting and domain shift rather than relying solely on Pearson's r coefficient for evaluation.

In sum, while the current analysis revealed largely concordant effects across sexes (likely due to limited sample sizes: $2121<N<2$ after sex-split), more nuanced sex differences may emerge in larger or longitudinal datasets, or with alternative modeling approaches (e.g., interaction terms or sex-by-protein effects), which should be explored in future work.

**eNote 9: Metabolite-set enrichment analysis for small-molecule from the UK Biobank NMR metabolomics data**

The UKBB metabolomics data primarily focuses on lipoprotein-related complexes and subclasses (e.g., high-density lipoproteins), with a smaller representation of small-molecule metabolites (e.g., creatinine and glycine). Therefore, we could not directly map these lipoprotein-related measures to commonly used small-molecule metabolite IDs (e.g., HMDB ID[8]). As a result, for the metabolite set enrichment analysis using the MetaboAnalyst[9], we included only the small-molecule metabolites and converted them to the common compound names and HMDB ID. This was exemplified by the spleen MRIBAG, which mapped 6 significant metabolites: pyruvic acid, sphingomyelins, unsaturated fatty acids, dehydroepiandrosterone, glycine, and linoleic acid (**Extended Data Fig. 5**). For instance, the biological pathway associated with glycine metabolism exhibited significant enrichment (enrichment ratio of 109.89; P-value=$1.27 \times 10^{-4}$), alongside other pathways such as SLC transporter disorders and lactic acidemia. These findings highlight that organ-specific and inter-organ protein and metabolic molecular profiles of the 7 organ-specific MRIBAGs may serve as valuable biomarkers for early detection of age-related diseases and targeted therapeutic interventions.

**eNote 10: Sex difference in MetWAS**

We conducted sex-stratified MetWAS analyses separately for males and females, which have similar sample sizes. Based on the results, we categorized proteins as "Discordant", "Concordant", or "Not Significant" using the following criteria:

**Discordant hit**:

- At least one sex shows a statistically significant association (P-value<0.05/327/0.05);
- The direction of effect differs between sexes (sign(female) ≠ sign(male)) for the standardized beta values.

**Concordant hit**:

- At least one sex shows a statistically significant association (P-value<0.05/327/0.05);
- The direction of effect is the same between sexes (sign(female) = sign(male)) for the standardized beta values.

**Non-significant hit**:

- Neither sex shows a statistically significant association (P-value>0.05/327/0.05)

Of note, one key distinction between the sex-stratified MetWAS and ProWAS analyses is that MetWAS benefited from a substantially larger sample size (ranging from 6,621 to 12,169 participants). With this definition, we found 768 concordant hits and 28 discordant hits for these MRIBAG-metabolite associations (**eTable 11** and **eFigure 13**). The observed discordant metabolite associations with organ-specific MRIBAG between females and males highlight pronounced sex-specific metabolic regulation, particularly in cardiovascular-related metabolites linked to the heart MRIBAG. For example, multiple HDL-related metabolites (e.g., HDL_size, L_HDL_CE, XL_HDL_P) show strong negative associations with heart MRIBAG in females but are largely oppositely (albeit non-significant) trending in males, reflecting sex differences in lipid metabolism and cardiovascular risk profiles[10]. Conversely, certain VLDL and LDL metabolite fractions exhibit significant positive associations with heart MRIBAG in males but not females, further emphasizing distinct lipoprotein remodeling influenced by sex hormones such as estrogen, which is known to modulate HDL and LDL particle distribution and function. Similarly, discordances in metabolites associated with kidney and liver BAGs, such as S_LDL_C_pct and LDL_FC_by_CE, suggest sex-specific variations in lipid and cholesterol handling in these organs. In the MetWAS results, several MRIBAGs displayed pronounced sex-specific association patterns, with significant metabolite correlations emerging predominantly in one sex. For example, the heart and pancreas MRIBAGs showed significant metabolite associations only in females, while the liver MRIBAG exhibited significant associations exclusively in males. These findings suggest that metabolic pathways linked to organ-specific aging may differ substantially between sexes, potentially reflecting sex-specific physiological mechanisms, hormonal influences, or differences in metabolic regulation. Such divergence underscores the importance of performing sex-stratified analyses to fully capture the biological complexity underlying organ aging.

Overall, these findings reinforce that metabolic pathways underlying aging and disease phenotypes manifest differently by sex, driven by hormonal, genetic, and environmental factors that influence metabolite profiles dynamically, making MetWAS particularly sensitive to uncovering such sex-specific aging signatures.

**eNote 11: The definition of genomic loci, independent significant SNP, lead SNP, candidate SNP**

FUMA defined the significant independent SNPs, lead SNPs, candidate SNPs, and genomic risk loci as follows (https://fuma.ctglab.nl/tutorial#snp2gene):

*Independent significant SNPs*

They are defined as SNPs with $P \leq 5\times10^{-8}$ that are independent of each other at the user-defined $r^2$ (set to 0.6 in the current study). We further describe *candidate SNPs* as those in linkage disequilibrium (LD) with independent significant SNPs. FUMA then queries each candidate SNP in the GWAS Catalog to check whether any clinical traits have been reported to be associated with previous GWAS studies.

*Lead SNPs*

Lead SNPs are defined as independent significant SNPs that are also independent of each other at $r^2<0.1$. If multiple independent significant SNPs are correlated at $r^2\geq0.1$, then the one with the lowest individual *P*-value becomes the lead SNP. If $r^2$ threshold is set to 0.1 for the independent significant SNPs, then they would constitute the identical set as the lead SNPs. FUMA thus advises setting $r^2$ to be 0.6 or higher.

*Genomic risk loci*

FUMA defines genomic risk loci to include all independent signals physically close or overlapping in a single locus. First, independent significant SNPs dependent on each other at $r^2\geq0.1$ are assigned to the same genomic risk locus. Then, independent significant SNPs with less than the user-defined distance (250 kilobases by default) away from one another are merged into the same genomic risk locus – the distance between two LD blocks of two independent significant SNPs is the distance between the closest points from each LD block. Each locus is represented by the SNP within the locus with the lowest *P*-value.

**eNote 12: Sensitivity check analyses for the GWAS robustness**

We applied several analyses to scrutinize the robustness of our primary GWAS. Manhattan and QQ plots, as well as the genomic inflation factor ($\lambda$) of the 7 MRIBAG GWASs, are presented in the MEDICINE portal (https://labs-laboratory.com/medicine/). The LDSC[11] intercept ($LDSC_b$=1.01 [1.0016, 1.0285]) of the 7 MRIBAG GWASs was close to 1, and the LDSC ratio (an indication of inflation vs. true polygenicity; $LDSC_r$=0.13 [0.0083, 0.2847]) was small, indicating that no severe inflation due to population stratification was observed. **Extended Data Fig. 7** presents the trumpet plots of the effective allele frequency vs. the $\beta$ coefficients of the 7 MRIBAG GWASs.

#### eNote 13: Sex difference in GWAS

We conducted sex-stratified GWAS separately for males and females (**eFigure 16a**). Overall, we observed sex differences (more likely regarding the magnitude and effect of the association) in the genomic loci associated with MRIBAGs. The brain MRIBAG showed the most consistent signals across sexes, particularly at a locus on chromosome 17, and demonstrated a very high genetic correlation between females and males ($g_c$=0.99 via LDSC). In contrast, other organ-specific MRIBAGs exhibited sex-specific differences in the strength of association signals. For instance, the heart MRIBAG showed significant associations only in males, even though the loci appeared at the same genomic positions in both sexes, but females did not achieve the significant threshold (P-value<$5x10^{-8}$). These findings align with our previous results from aging clocks[12,13], where we similarly observed that while the effect sizes might differ between sexes, the associated loci often occupy the same physical genomic locations.

To further explore potential sex differences, we estimated pairwise genetic correlations among the seven MRIBAGs separately for males and females (**eFigure 16b**). The results revealed sex-specific patterns of shared genetic architecture. In females, the liver MRIBAG showed distinct positive genetic correlations with the spleen and kidney MRIBAGs, suggesting coordinated aging or shared genetic regulation among these organs in women. In contrast, in males, the liver MRIBAG was positively correlated with the pancreas MRIBAG, indicating a different axis of organ co-aging or genetic interplay. These findings underscore that the genetic relationships among organ-specific aging processes may be structured differently by sex, potentially reflecting underlying physiological, hormonal, or metabolic differences.

**eNote 14: Additional sensitivity test analysis of spleen and liver MRIBAG's protective effect on all-cause mortality**

We further accounted for potential confounding due to underlying pathologies, such as metabolic disorders that could influence the expression of the liver MRIBAG, through our disease-free sensitivity analyses (**Supplementary eTable 8b**). However, population selection bias may also contribute to this counterintuitive finding, as well as the complex interactions (nonlinear) between the underlying organ imaging features, the aging clock, and age at death (**Supplementary eFigure 19–20**). Another possible explanation is the suboptimal model performance for these organs, which may lead to biologically nonspecific or misestimated BAG values. While we applied rigorous cross-validation and external testing, the relatively low predictive accuracy suggests these results should be interpreted as exploratory and hypothesis-generating. Additionally, one potential explanation for the protective association of spleen and liver MRIBAGs with mortality is reverse causation. That is, individuals already on a trajectory toward mortality (due to unmeasured or subclinical conditions) may show signs of accelerated aging in other organs but not necessarily in the liver or spleen, either because these organs are less affected or because imaging markers fail to capture subtle pathology, thus making them appear "younger" even in individuals closer to death.

Furthermore, we conducted survival analyses on the raw imaging features underlying the liver and spleen MRIBAGs. Interestingly, while certain individual features showed significant associations with increased mortality risk, the composite measures (i.e., the MRIBAG) demonstrated a potentially protective effect (**Supplementary eTable 8c-e**). For example, a "younger-looking" liver (lower BAG) may reflect better metabolic health, reducing mortality risk; whereas larger liver volume (field ID=21080) may reflect steatosis and increase mortality risk. This reversal suggests that the MRIBAG algorithm synthesizes these biomarkers in a non-linear integration way that captures net protective biological pathways. That is, the liver BAG may reflect metabolic resilience (e.g., mitochondrial function), while cT1/volume reflects damage. The relationships among the MRIBAG, age at mortality, and the underlying imaging features (e.g., liver and spleen volume) are further illustrated in **Supplementary eFigure 19-20**. Additionally, SHAP analysis revealed that liver and spleen volumes exhibited opposite directional associations, as well as feature importance, with the MRIBAG compared to other imaging features of the same organ, further supporting that the MRIBAG may capture complementary biological insights beyond individual imaging measures (**Supplementary eFigure 7**). Previous aging clock studies have reported similar findings. For instance, a recent proteome-based aging clock study[14] observed that a younger biological age in certain organs (e.g., arteries) can be associated with higher mortality risk. Similarly, in our prior analysis of 5 MetBAGs[15], we identified a protective effect of the hepatic MetBAG on mortality. To summarize, the reduced risk of all-cause mortality linked to the liver and spleen MRIBAGs likely reflects their roles in maintaining metabolic health, immune homeostasis, and systemic aging regulation. Alternatively, another explanation is the "healthy survivor effect", where participants with healthier liver and spleen function may be overrepresented in the study population, especially in older age groups. This has been noted in prior research and commentaries[16,17].

**eNote 15: Further methodological improvements in the age prediction performance of abdominal MRI BAGs**

Additional improvements could be achieved by adopting more advanced modeling approaches, such as deep neural networks that operate directly on voxel-wise MRI data. Prior studies have demonstrated the effectiveness of such methods in enhancing age prediction accuracy for the brain and abdominal organs. For instance, Lee and colleagues[18] developed a deep learning model using large-scale brain FDG-PET and structural MRI data, achieving an MAE of approximately 3 years. Similarly, another study[19] applied convolutional neural networks to abdominal MRI data (e.g., liver and spleen) and reached a comparable MAE. However, as we demonstrated in our two previous studies involving neural networks applied to brain voxel-wise MRI[20] and plasma proteins[7], achieving state-of-the-art age prediction accuracy (e.g., lower MAE) is not necessarily the ultimate quest. For example, in our proteome-based aging clock analysis[7], we observed an inverse U-shaped relationship between model fitting precision (MAE ranges from 7 years to 3 years) and the effect size ($\beta$ coefficient) of associations between the brain ProtBAG and cognition (digital symbol substitution test). This supports our claim that the primary objective is to construct aging clocks optimized for cross-domain prediction (e.g., disease risk, cognitive performance, and mortality) rather than minimizing prediction error alone. Finally, contrastive learning frameworks (e.g., SimCLR[21], MoCo[22], BYOL[23]) represent a promising future direction for capturing shared and distinct aging representations across multiple organs. These approaches may enable the discovery of latent cross-organ aging signatures and individual-level discordances without requiring direct supervision.

**eFigure 1: Model performance using the elastic net regression and the neural network**

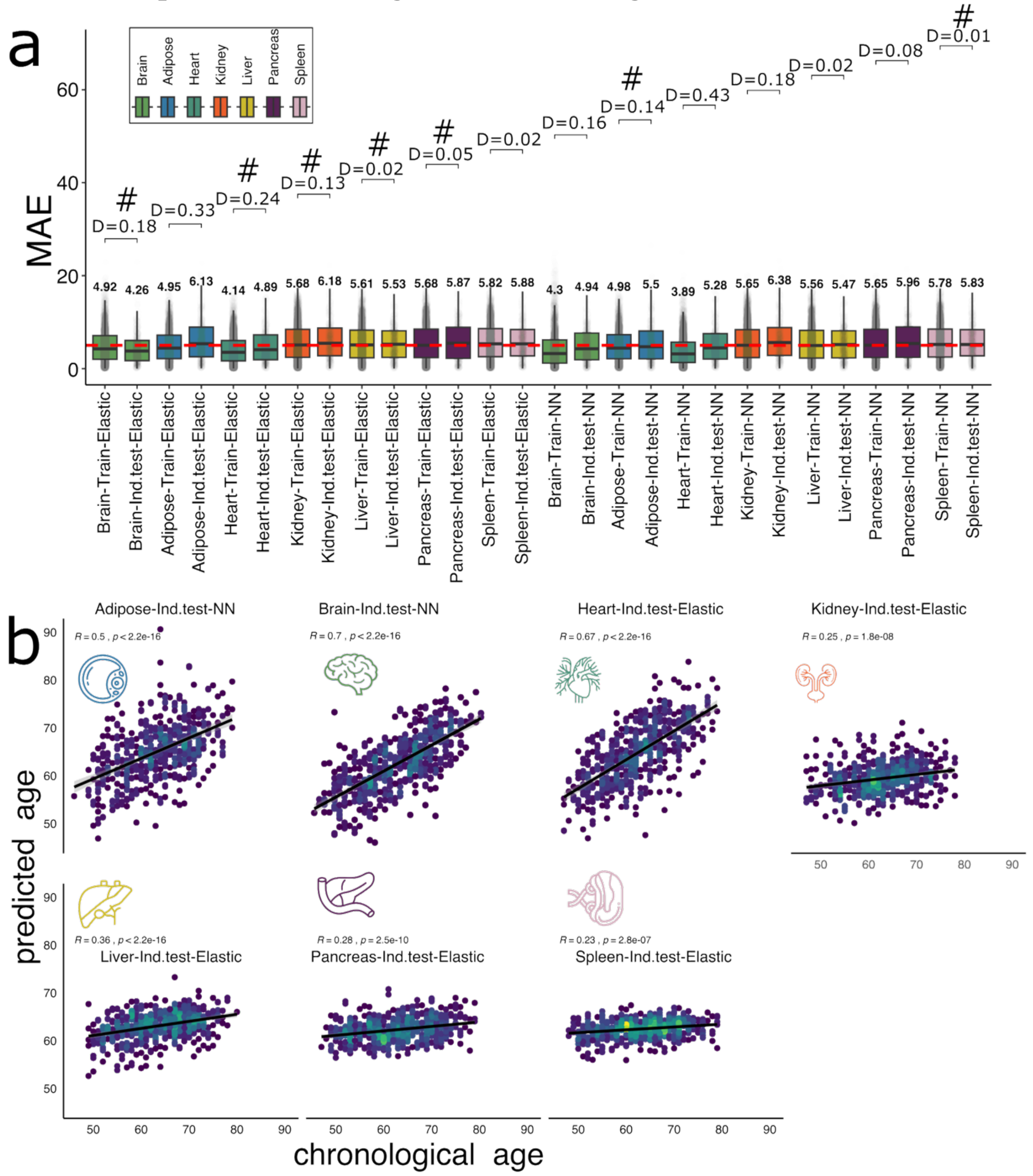


Compared to the two main models, including Lasso and SVR (**Fig. 1b)** in the main manuscript, Elastic Net and NN do not consistently outperform the two main models. **a**) For the 7 MRIBAGs, the mean absolute error (MAE) of age prediction models using Elastic Net and NN is shown for both the training dataset (cross-validated training/validation/test) and the independent test dataset (Ind. test). Cohen's D quantifies the effect size of the difference between these datasets, reflecting potential model generalizability, assuming comparable age and sex distributions. The optimal model (#) for each organ and tissue was selected based on the lower Cohen's D value between the two models. All MAEs are reported without age bias correction. **b**) Scatter plots display the optimal model for each organ/tissue in the Ind. test dataset, with Pearson's $r$ and P-values indicating the association between chronological age and predicted age. All results are shown before the correction for age bias was applied[24].

**eFigure 2: Incremental $R^2$ of the 7 MRIBAGs to predict 14 systematic disease categories and age at mortality on top of age and sex**

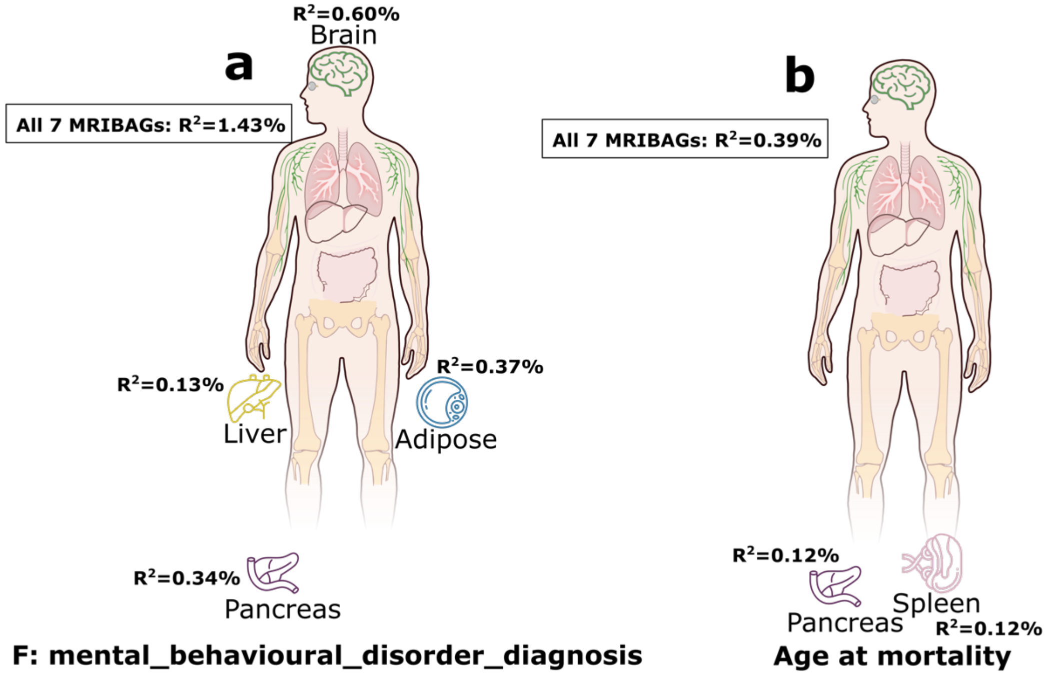


We evaluated the additional predictive power, quantified as incremental $R^2$, beyond calendar age and sex to predict 14 systemic disease categories for major human organ systems (e.g., CNS or circulatory system diseases). We showcase here for predicting **a**) the mental behavioral disease category (ICD=F, which showed the highest $R^2$ among the 14 disease categories) in 2652 healthy controls (without any ICD diagnosis) and 1111 patients, and **b**) all-cause mortality ($N$=175 participants). The null model included only calendar age, sex, and genetic principal components as predictors, while the alternative model incorporated additional MRIBAGs. Significant results (P-value< 0.05/7) are highlighted in bold for figure **a**) and a nominal P-value threshold (P-value < 0.05) for figure **b**) due to the smaller sample size. To calculate the incremental $R^2$, we first constructed a null model with age and sex as predictors and the disease outcome as the response variable. The alternative model included the same predictors plus one of the 7 MRIBAGs as an additional predictor; we also tested a full alternative model incorporating all 7 MRIBAGs simultaneously. The incremental $R^2$ was computed as the difference between the (pseudo) $R^2$ values of the alternative and null models, using the *DescTools* R package (v0.99.38). The choice of (pseudo) $R^2$ metric depended on whether the model was a logistic or linear regression.

**eFigure 3: Evidence for model overfitting and poor generalizability to independent test dataset due to feature multicollinearity**

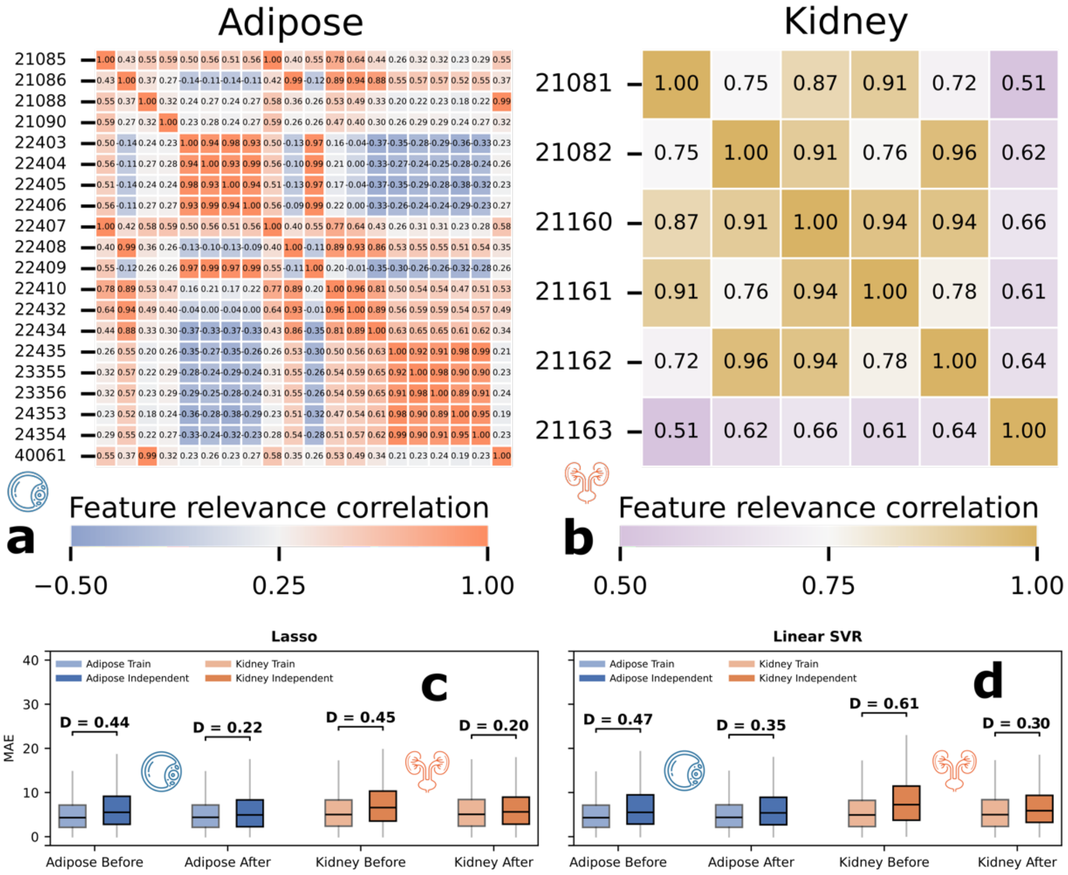


**a**) Pairwise Pearson's $r$ for the adipose features. **b**) Pair-wise Pearson's $r$ for the kidney features. The full name of the features is presented in **eNote 1**. **c**) Model generalizability to independent test data before and after removing the highly correlated features for the adipose MRIBAG. **d**) Model generalizability to independent test data before and after removing the highly correlated features for the kidney MRIBAG.

**eFigure 4: Generalizability of the brain MRIBAG from UKBB to A4**

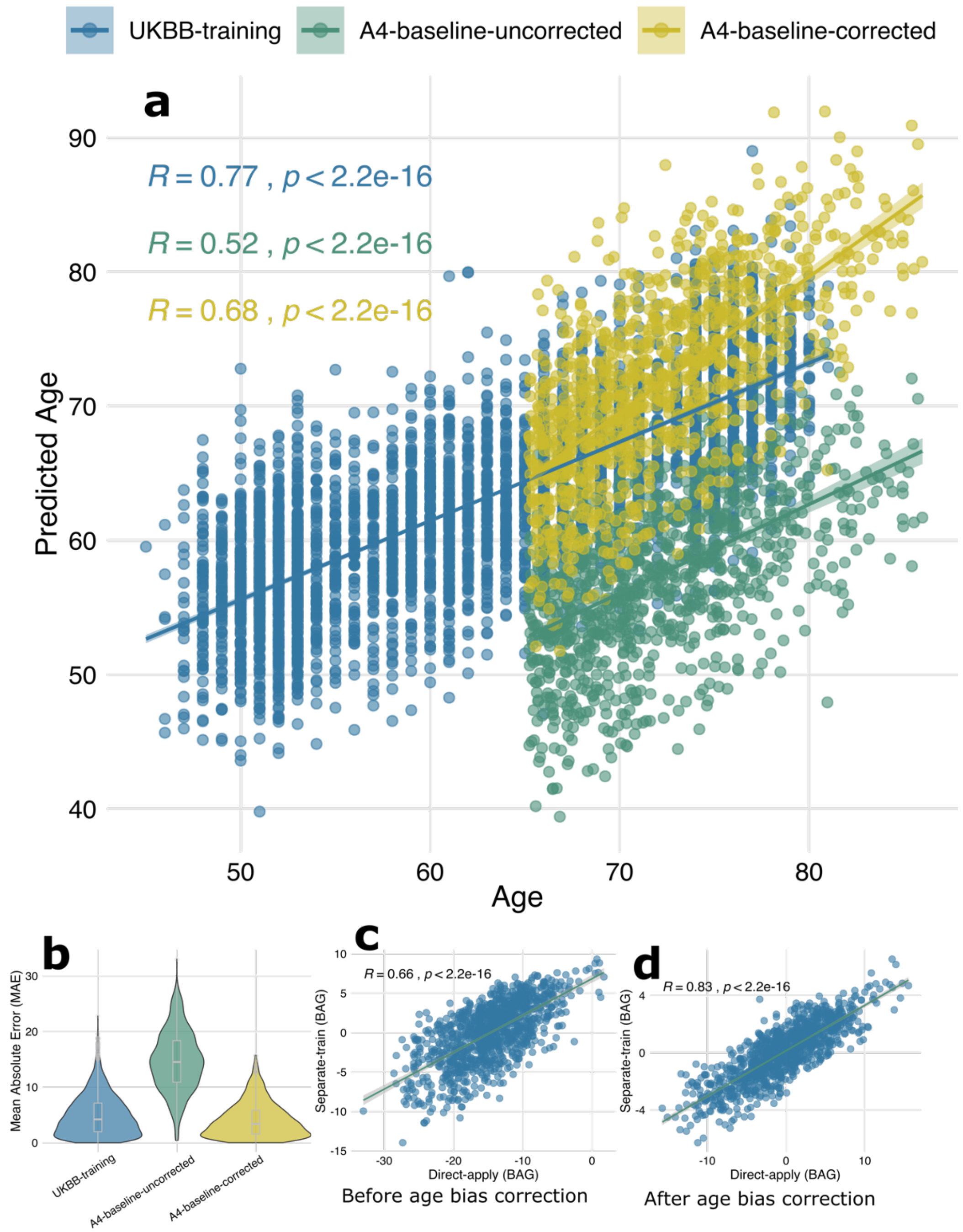


**a**) When applying the trained model to A4, after statistically harmonizing imaging features using our ComBat-GAM[25] model, we observed potential domain shifts between the UKBB training data and the independent A4 dataset. Implementing age bias correction partially mitigated these discrepancies. **b**) Mean absolute error (MAE) in the UKBB training dataset, and in the A4 baseline data before and after age bias correction. **c**) Scatter plot illustrating the correlation between BAG estimates obtained by directly applying the trained model to A4 and those derived from an independently trained age prediction model within the A4 study before age bias correction. **d**) Scatter plot illustrating the correlation between BAG estimates obtained by directly applying the trained model to A4 and those derived from an independently trained age prediction model within the A4 study after age bias correction.

**eFigure 5: AD drug differential responses using the brain MRI derived i) by directly applying the pretrained model to A4 vs. ii) by independently training the model in A4**

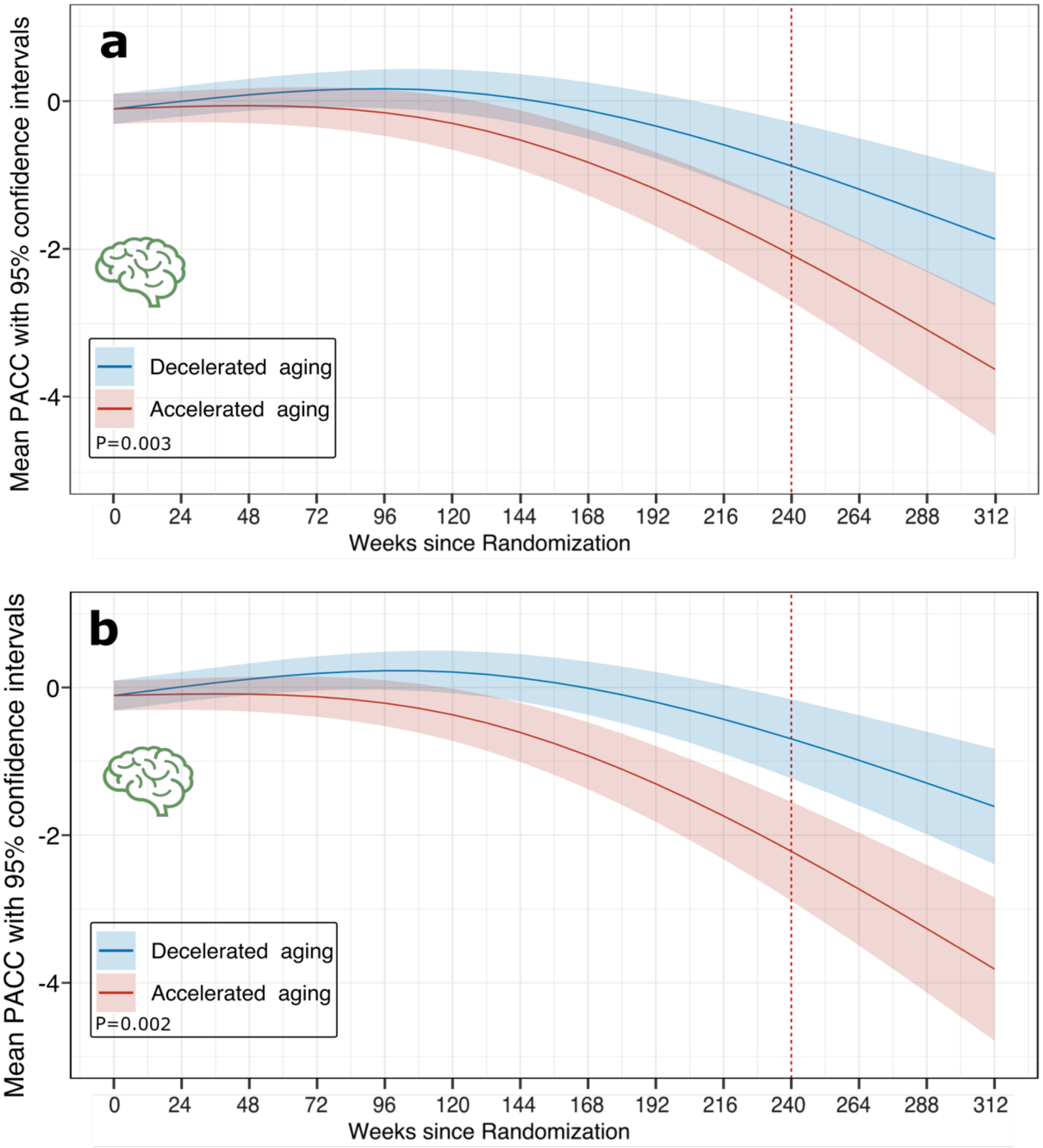


**a**) The same analysis was done as in **Fig. 6**. **b**) We directly applied the Lasso model trained in UKBB to brain MRI scan in A4 to derive the brain MRIBAG. The conclusions from brain MRIBAG using the two different approaches remain the same.

**eFigure 6: Sex difference for the 7 MRIBAGs**

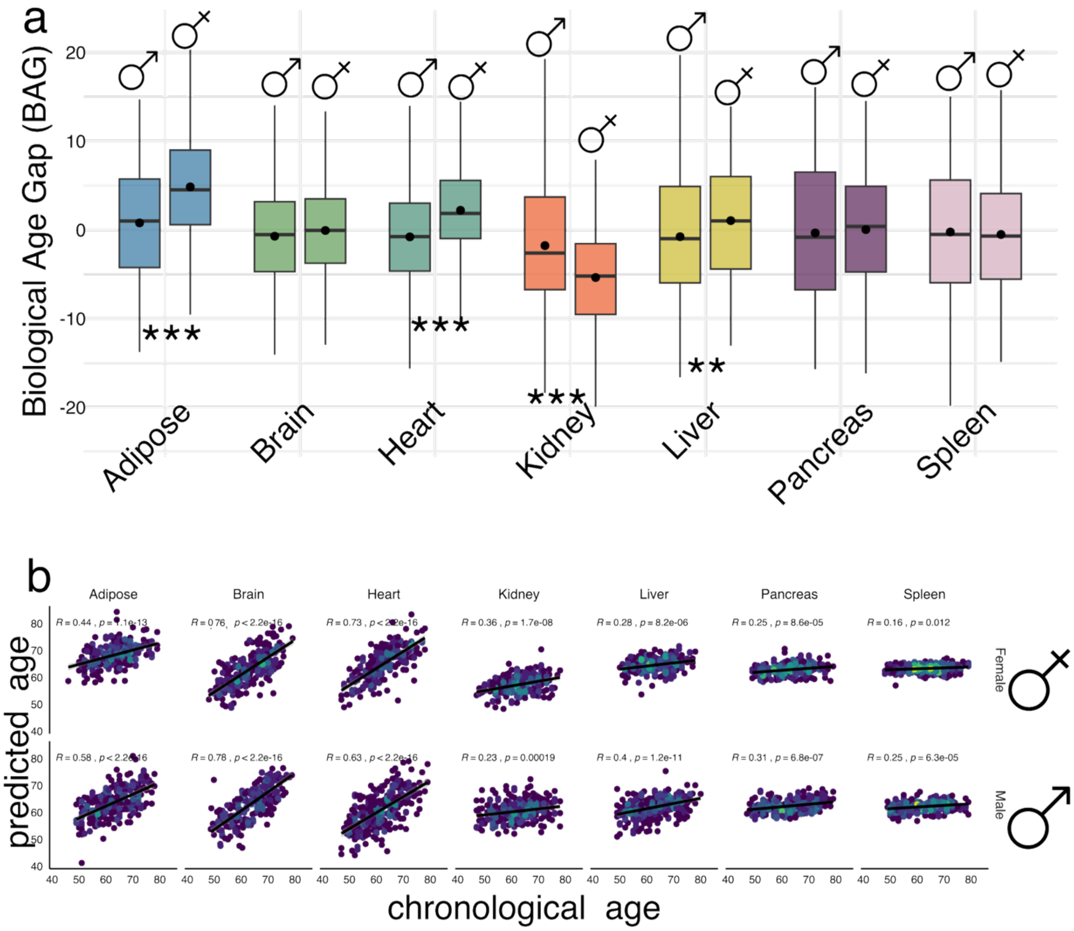


**a**) Sex differences in the 7 MRIBAGs, shown by the distribution of each BAG across organs for males and females. Statistical significance between sexes was assessed using Welch's t-test, with asterisks indicating the level of significance (P-value<0.05/7). **b**) Scatter plots of predicted age versus chronological age for each organ, stratified by sex. Pearson's correlation coefficient (*r*) and associated P-value are annotated separately for males and females.

**eFigure 7: SHAP values to denote feature importance for generating the 7 MRIBAGs**

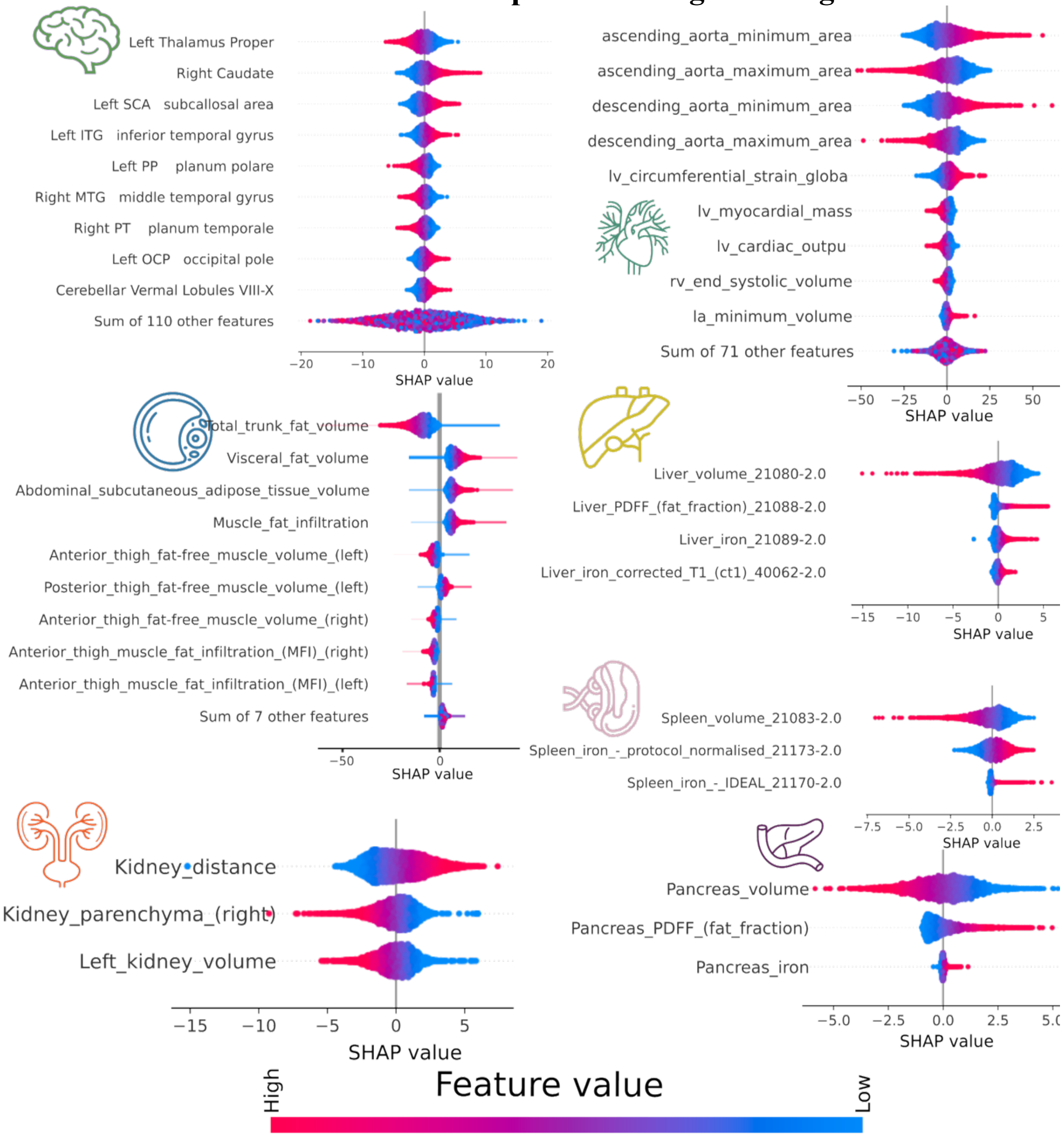


We applied SHAP to assess feature importance in deriving the 7 MRIBAGs, quantifying the contribution of each imaging feature to the predicted biological age. This approach also helps explain the differing directional associations observed for organ volumes (e.g., liver and spleen volumes) relative to other imaging-derived features. A positive SHAP value indicates that the feature contributes to increasing the predicted MRIBAG, suggesting that it is associated with an older-appearing organ relative to chronological age. In contrast, a negative SHAP value indicates that the feature lowers the predicted MRIBAG, implying a younger-appearing organ. These values help interpret how individual features drive organ-specific biological aging predictions in the model. Overall, SHAP values interpret both the direction of a feature's effect (positive or negative) and how feature magnitude (high or low) relates to predicted biological aging.

**eFigure 8: ProWAS for the left thalamus and protein-set enrichment analysis (PSEA) for the significant proteins**

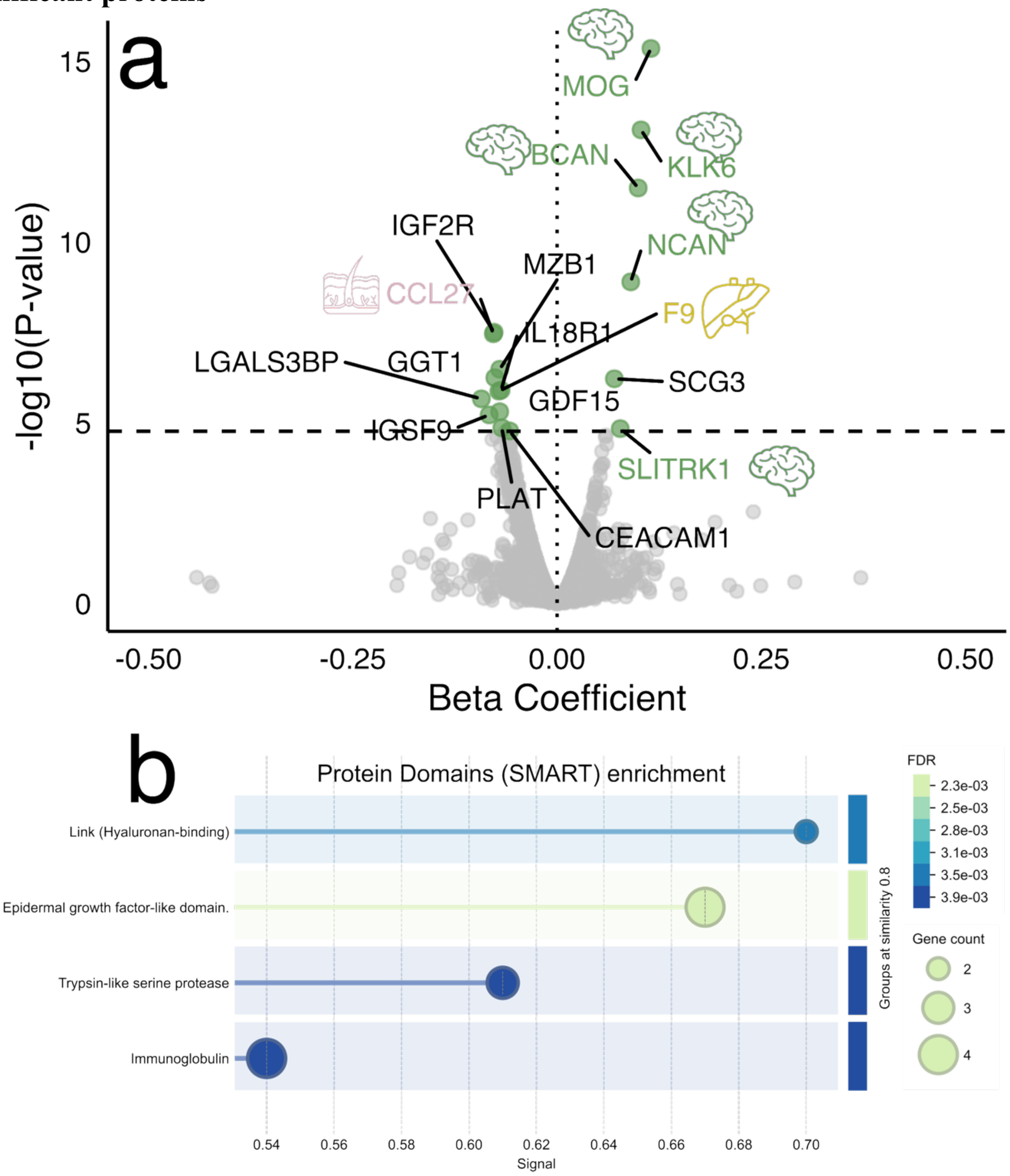


**a**) Protein-wide association study (ProWAS) results linking left thalamus volume with 2,923 plasma proteins. Seventeen significant proteins are highlighted with colored dots; organ-enriched proteins are marked with organ icons as defined in our previous ProtBAG study[7]. **b**) Protein-set enrichment analysis of the 17 significant proteins using the SMART database via the STRING platform.

**eFigure 9: Comparison between UKBB Olink ProWAS and BLSA SomaScan ProWAS for the common significant proteins**

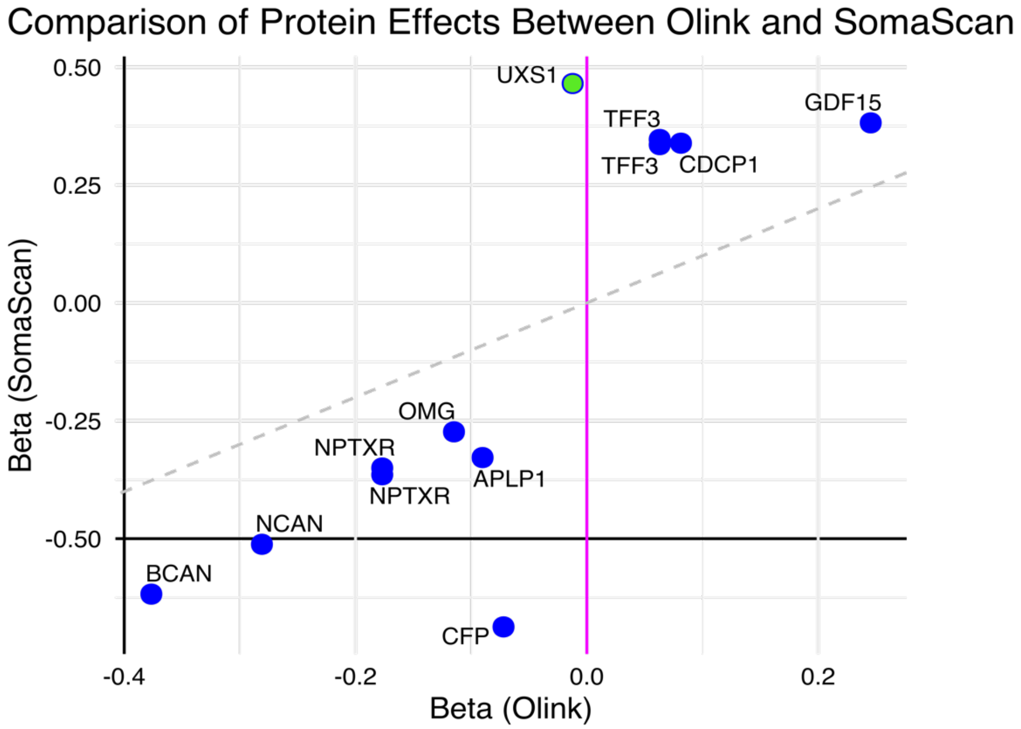


After applying Bonferroni correction for the 2,139 common proteins shared between the two platforms (P-value < 0.05/2139), we identified 12 significant proteins in both platforms. The majority exhibit similar directional effects, with UXS1 being the exception.

**eFigure 10: Functional protein set enrichment analysis (PSEA) and protein-protein interaction (PPI) network**

STRING conducts protein-protein interaction (PPI) analysis by combining various sources of interaction evidence, such as experimental data, computational predictions, and text mining. In addition to interactions, STRING can carry out functional enrichment analysis to identify Gene Ontology (GO) terms (e.g., biological processes, molecular functions), KEGG pathways (e.g., metabolic and signaling pathways), and protein domains (e.g., shared structural motifs). We found that the kidney MRIBAG exhibited predominantly positive associations with these plasma proteins, while the spleen MRIBAG displayed predominantly negative associations.

**Brain MRIBAG**: This has been shown in the main manuscript as an example.

**Heart MRIBAG**

We only found 1 heart MRIBAG-related protein, so we were not able to perform the same analyses as in other MRIBAGs.

**Adipose MRIBAG**

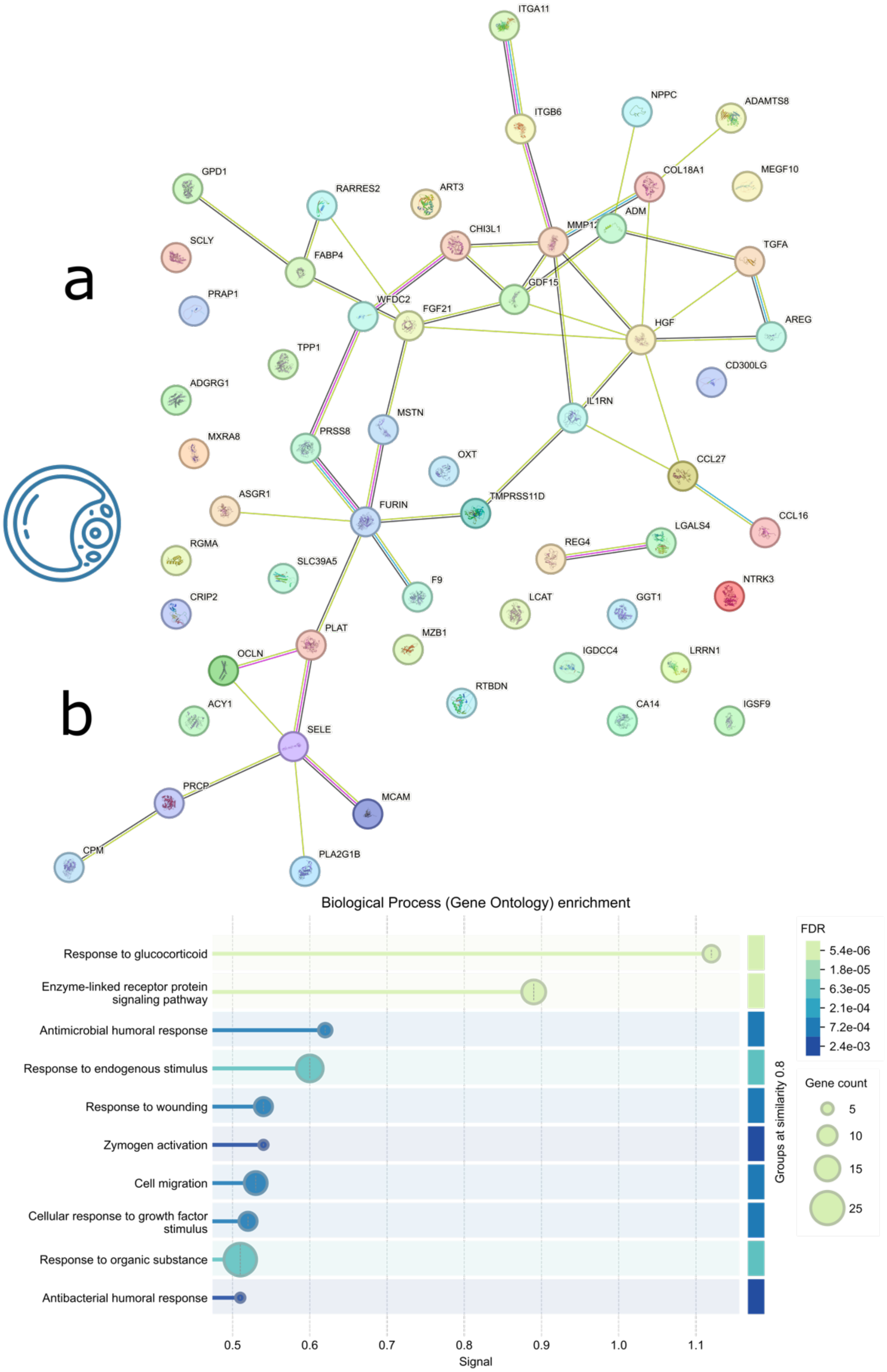


**a**) Protein-protein interaction network of the adipose MRIBAG-related proteins. **b**) Functional enrichment analyses of the adipose MRIBAG-related proteins. Results and figures are generated using STRING v12.0 (https://string-db.org/).

**Kidney MRIBAG**

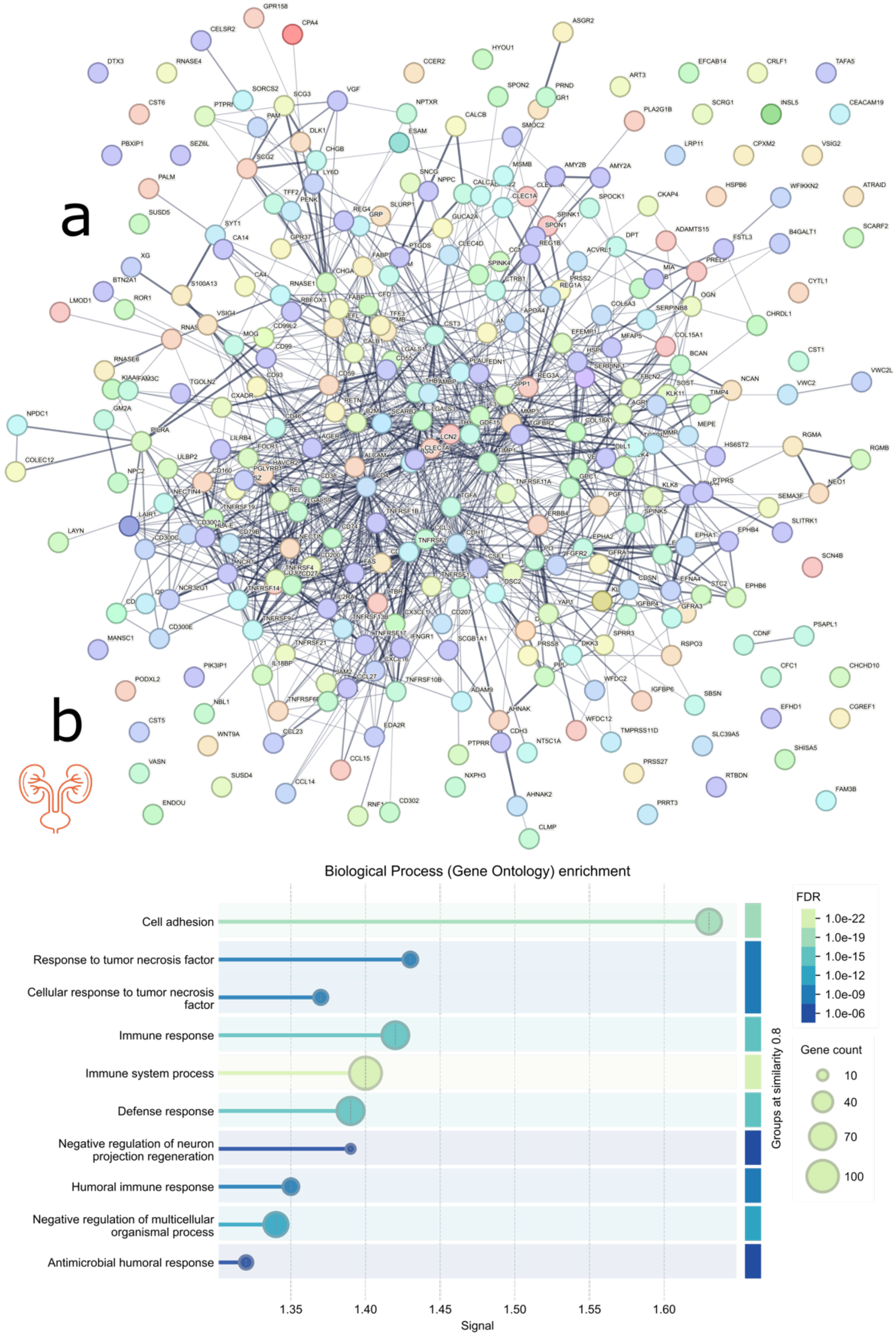


**a**) Protein-protein interaction network of the kidney MRIBAG-related proteins. **b**) Functional enrichment analyses of the kidney MRIBAG-related proteins. Results and figures are generated using STRING v12.0 (https://string-db.org/).

**Spleen MRIBAG**

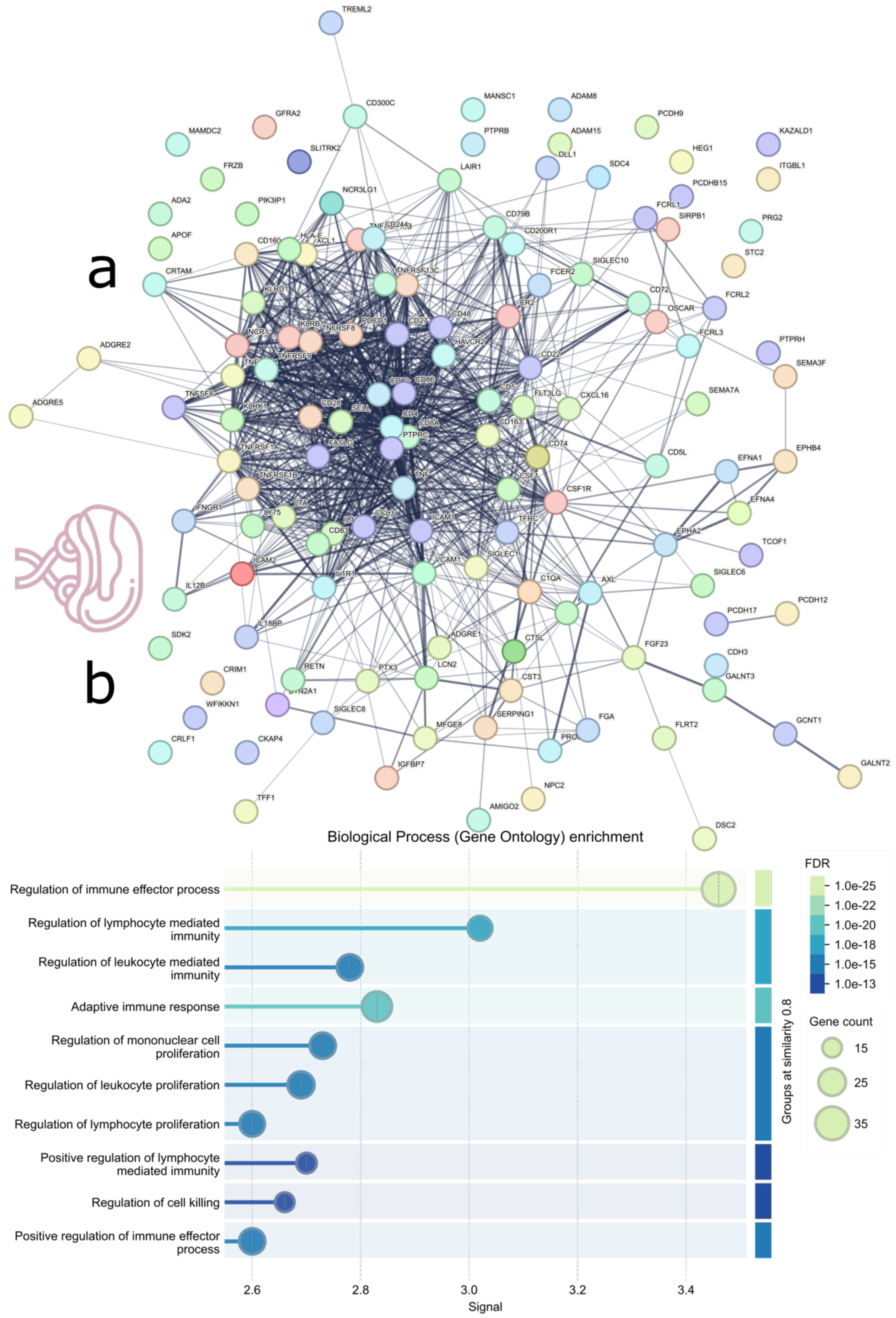


**a**) Protein-protein interaction network of the spleen MRIBAG-related proteins. **b**) Functional enrichment analyses of the spleen MRIBAG-related proteins. Results and figures are generated using STRING v12.0 (https://string-db.org/).

**Pancreas MRIBAG**

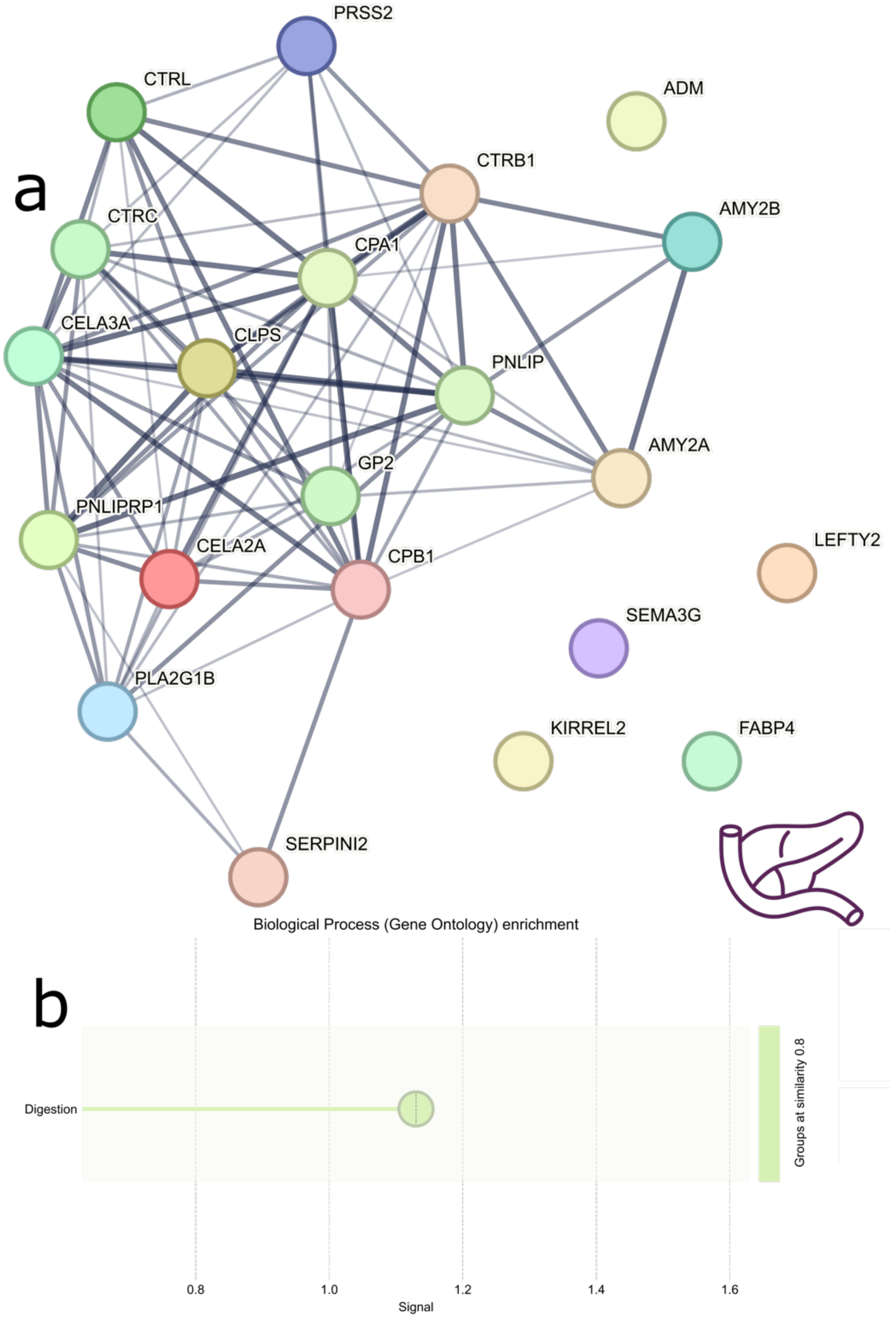


**a**) Protein-protein interaction network of the pancreas MRIBAG-related proteins. **b**) Functional enrichment analyses of the pancreas MRIBAG-related proteins. Results and figures are generated using STRING v12.0 (https://string-db.org/).

#### Liver MRIBAG

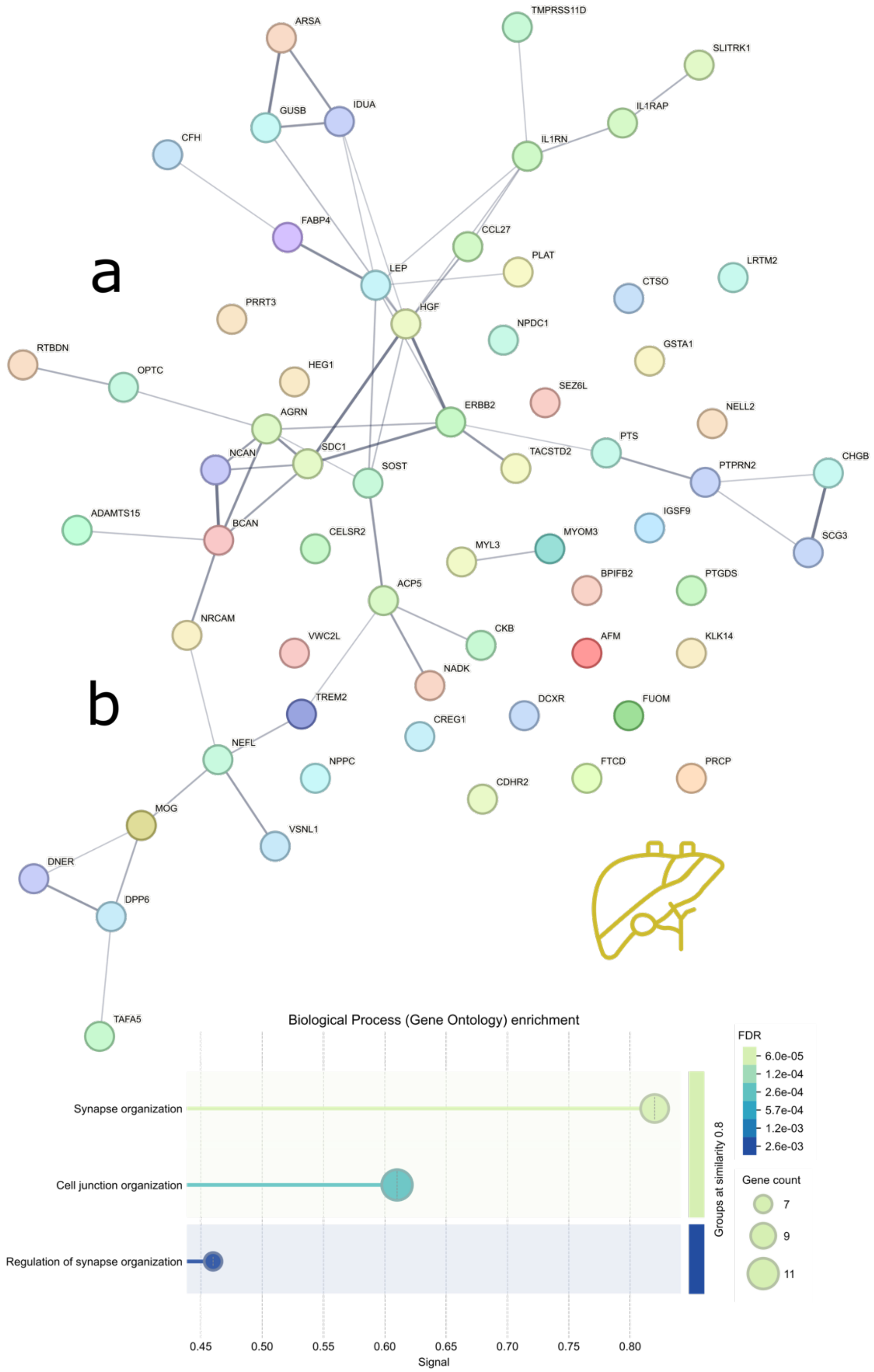


**a**) Protein-protein interaction network of the liver MRIBAG-related proteins. **b**) Functional enrichment analyses of the liver MRIBAG-related proteins. Results and figures are generated using STRING v12.0 (https://string-db.org/).

**eFigure 11: Sex-specific ProWAS results**

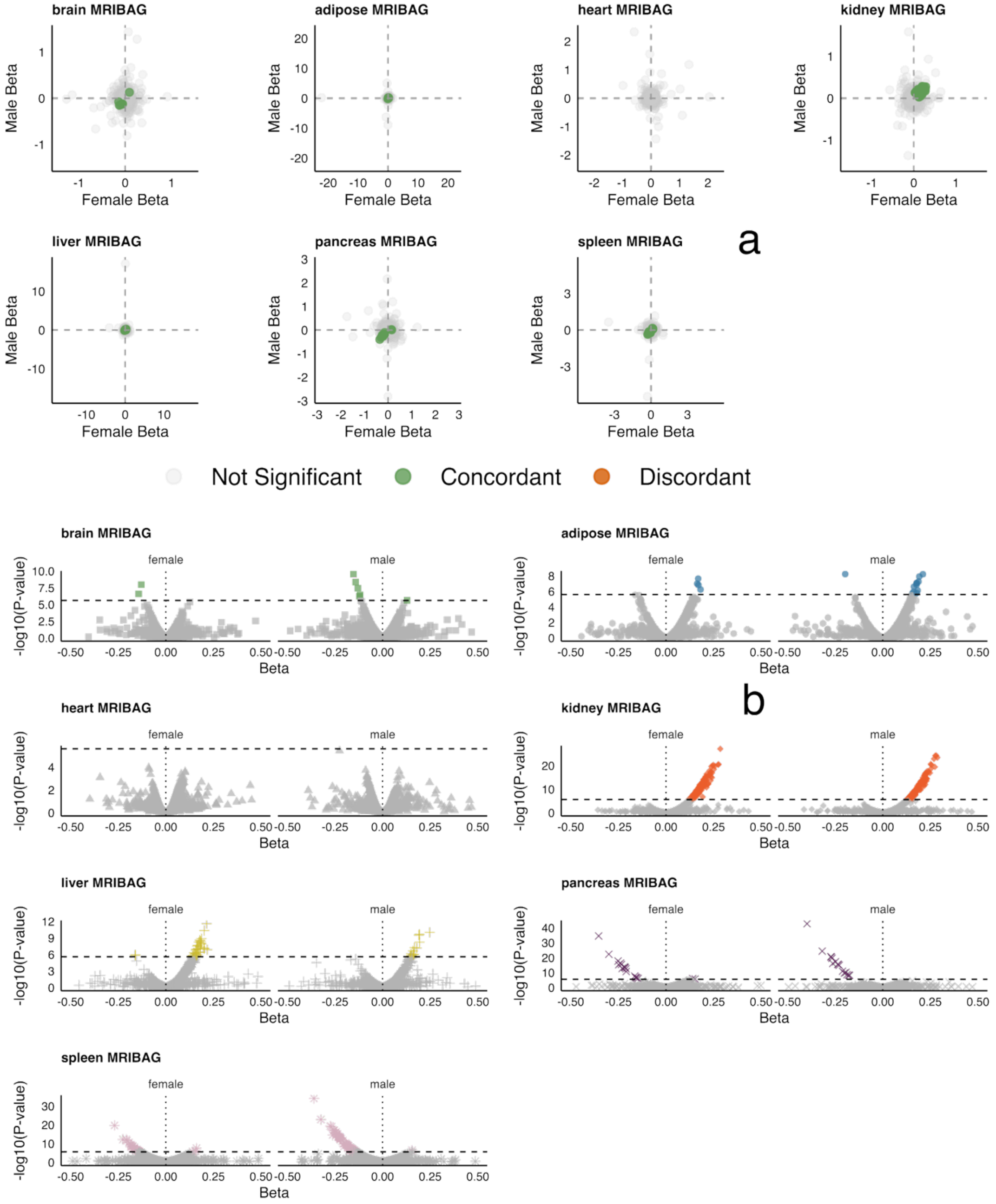


**a**) Scatter plot between the beta coefficient from the male and female only ProWAS analyses. Associations were classified as discordant if at least one sex showed significance, and the direction of effect (sign of $\beta$) differed between sexes. They were considered concordant if at least one sex was significant and both sexes had the same direction of effect, while associations with no significant result in either sex were labeled not significant (P-value<0.05/2923/7). **b**) Volcano plots for sex-stratified ProWAS results. Colored icons denote significant proteins.

**eFigure 12: Genetic correlation between the 7 MRIBAGs and 2923 proteins and 327 metabolites**

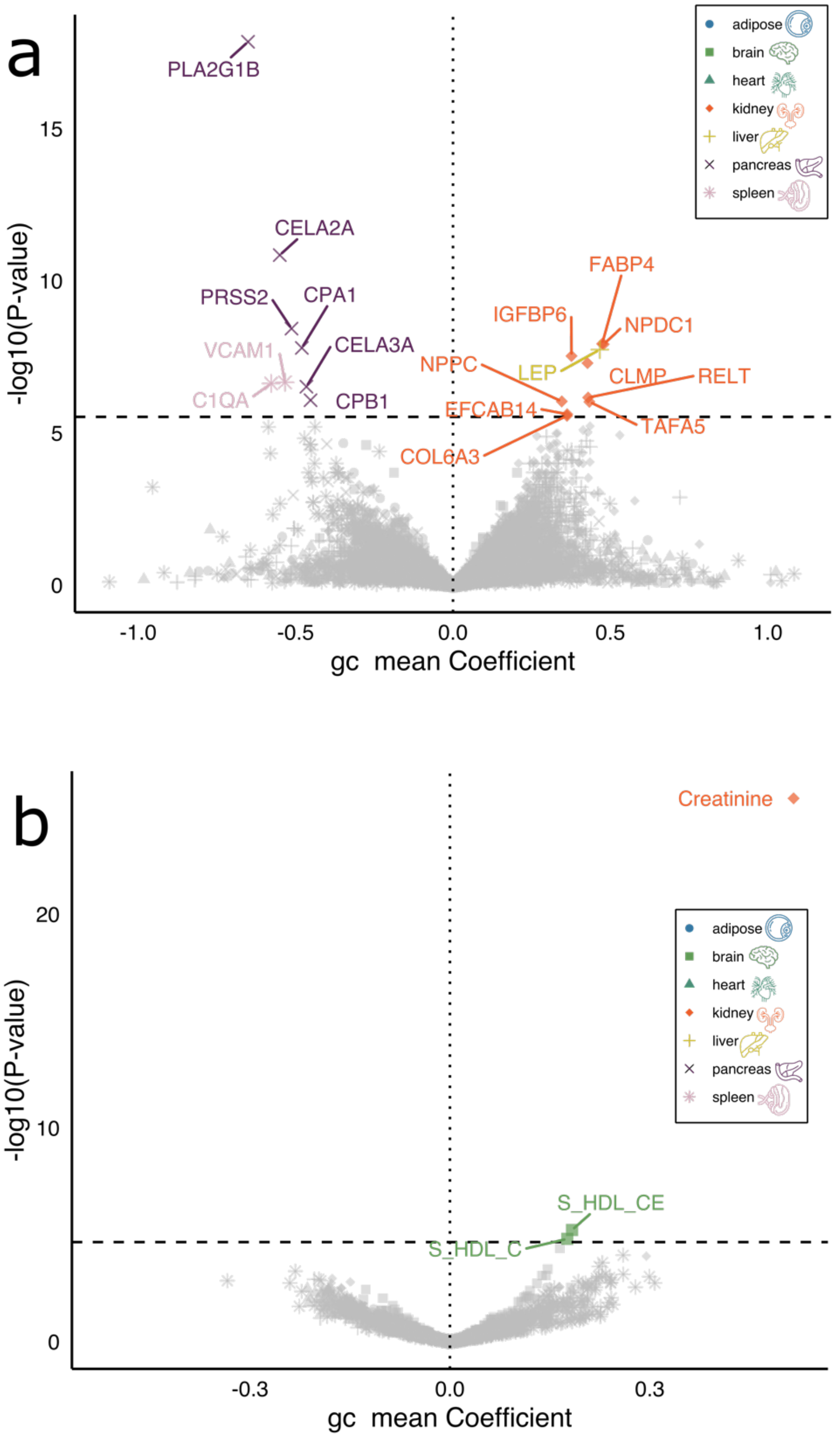


**a**) Genetic correlations between the 7 MRIBAGs and 2923 plasma proteins. Significant signals were denoted after Bonferroni correction (0.05/2923/7). **b**) Genetic correlations between the 7 MRIBAGs and 327 plasma metabolites. Significant signals were denoted after Bonferroni correction (0.05/327/7). It is expected for fewer significant associations as LDSC used GWAS summary statistics, and many of the MRIBAG GWASs were underpowered due to sample sizes.

**eFigure 13: Sex-specific MetWAS results**

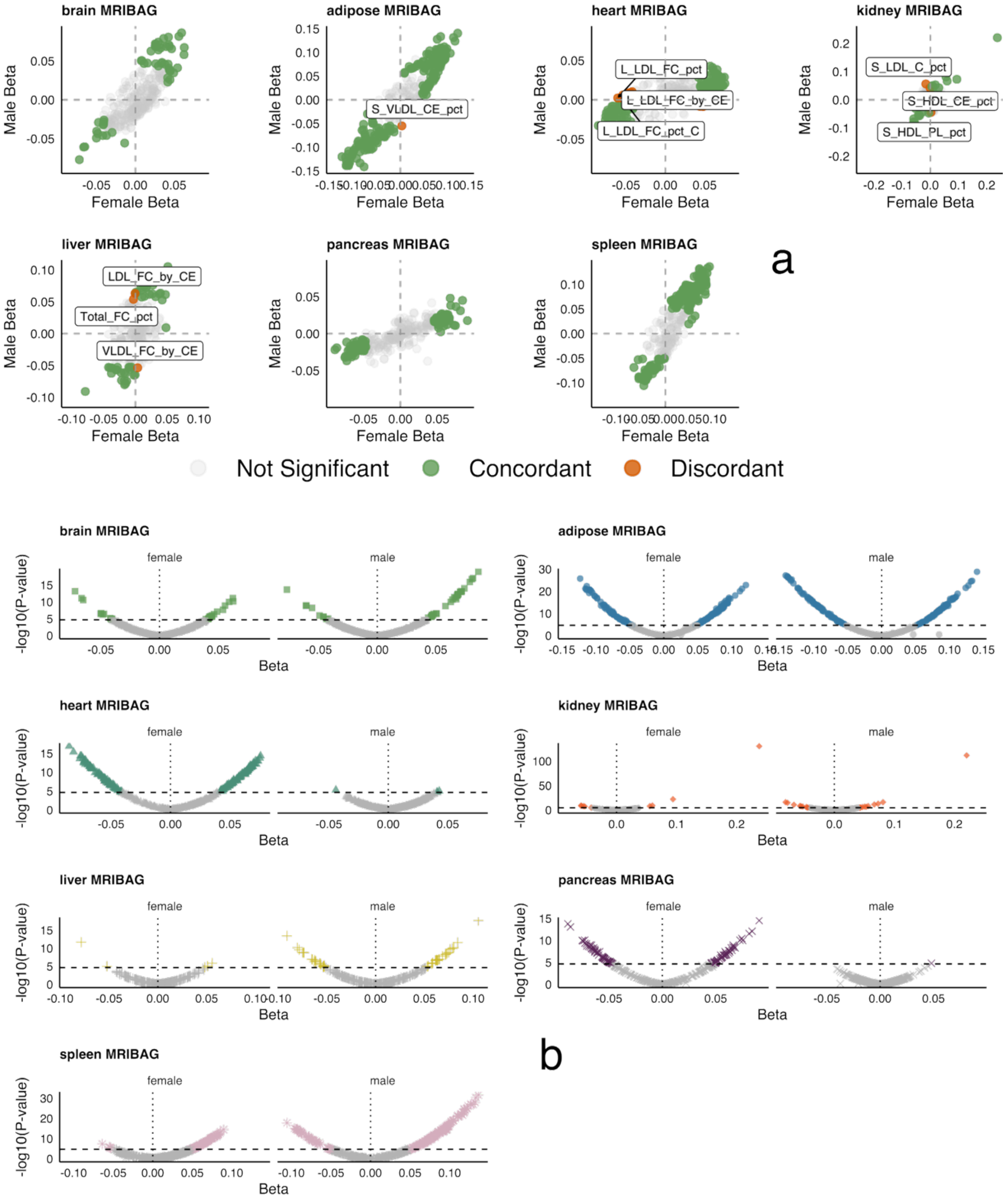


**a**) Scatter plot between the beta coefficient from the male and female only MetWAS analyses. Associations were classified as discordant if at least one sex showed significance, and the direction of effect (sign of $\beta$) differed between sexes. They were considered concordant if at least one sex was significant and both sexes had the same direction of effect, while associations with no significant result in either sex were labeled not significant (P-value<0.05/327/7). We annotated exemplary discordant hits for each MRIBAG. **b**) Volcano plots for sex-stratified MetWAS results. Colored icons denote significant metabolites.

**eFigure 14: sex-specific GWAS analyses**

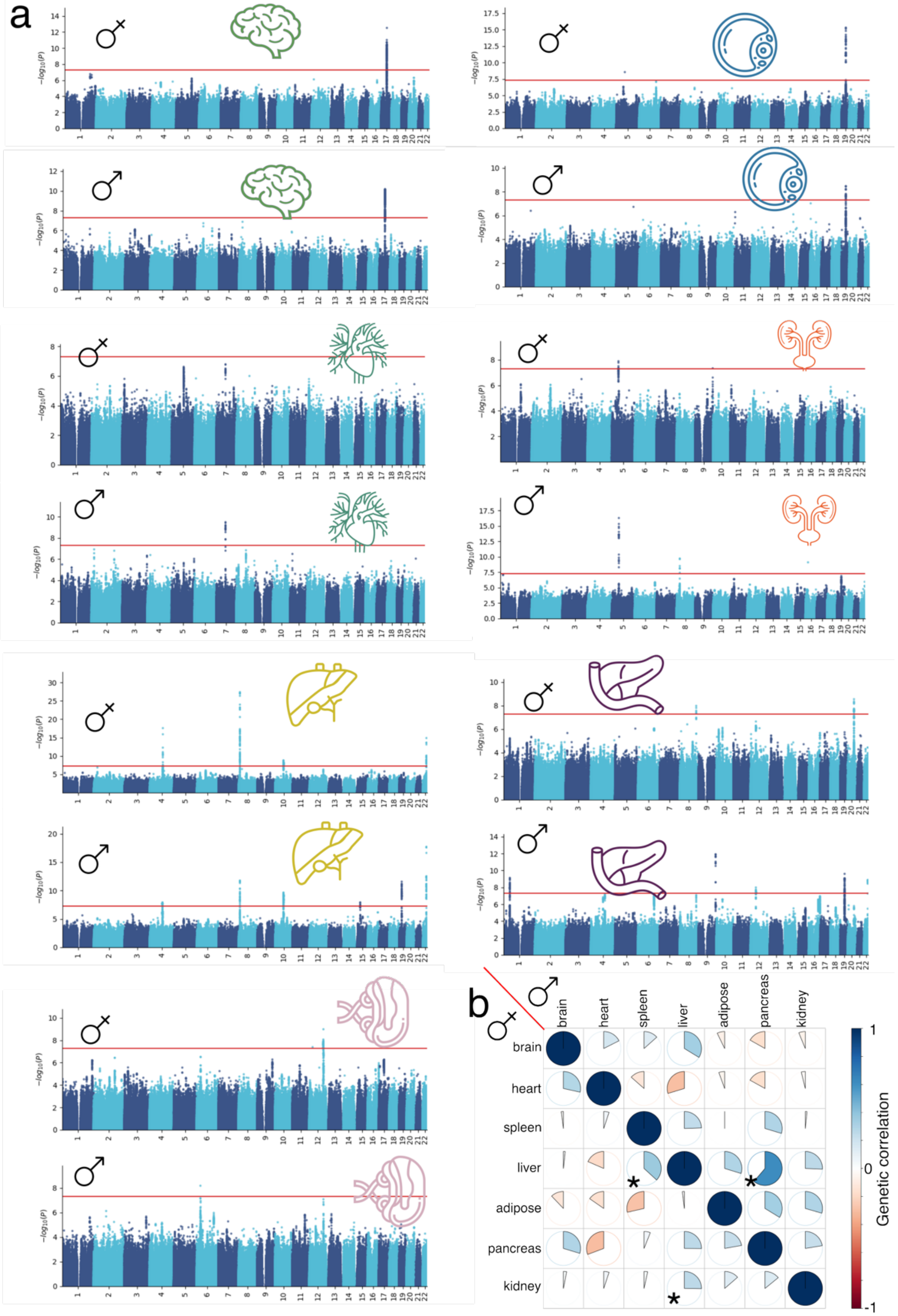


**a**) Sex-stratified GWAS using females and males only for each MRIBAG. **b**) Pair-wise genetic correlation between the 7 MRIBAG using the sex-stratified GWAS summary data from a. P-values were FDR-corrected to denote statistical significance.

**eFigure 15: Genetic correlation and phenotypic correlation between the 7 MRIBAGs**

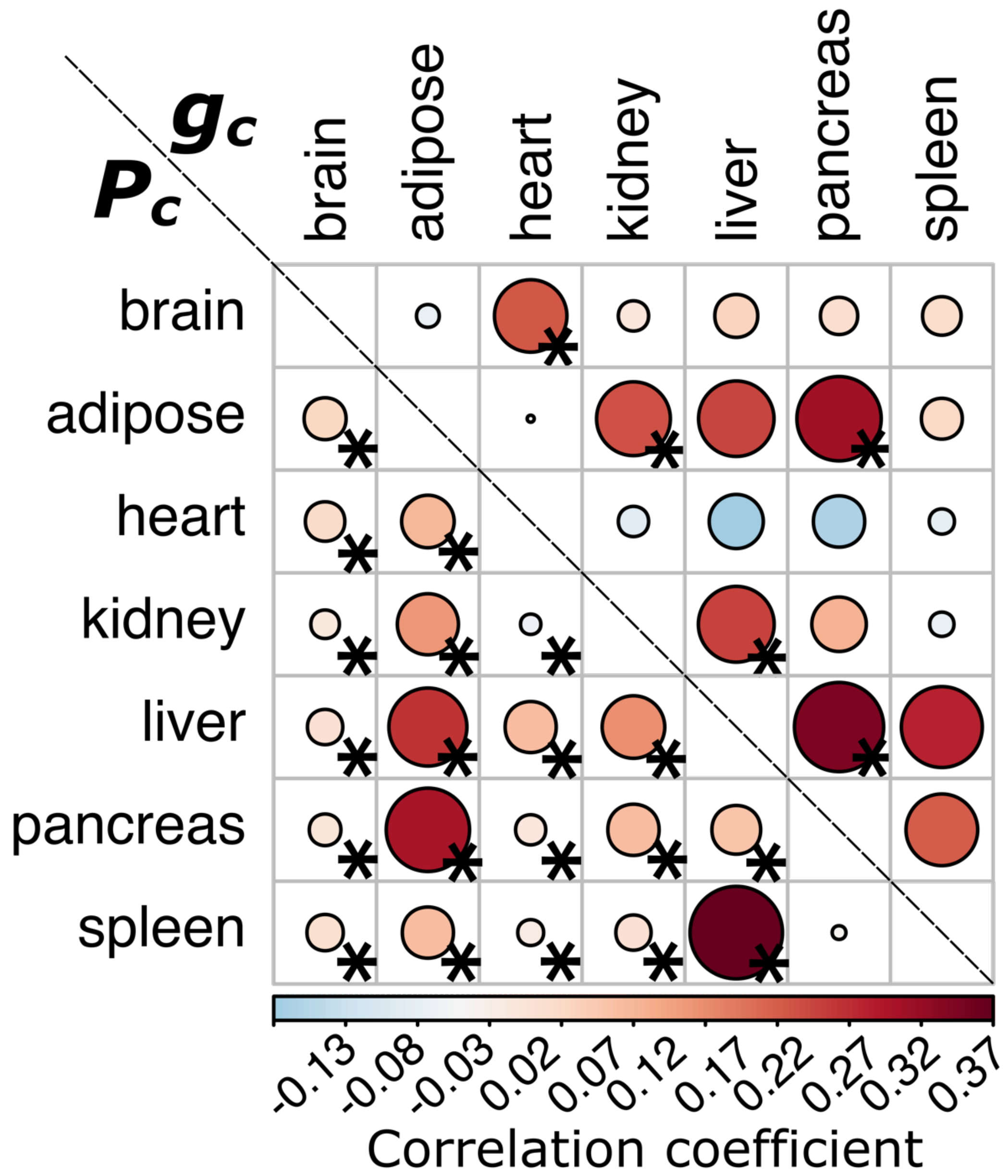


Genetic correlations (upper triangle, estimated using LDSC) and phenotypic correlations (lower triangle, Pearson's *r*) among pairwise MRIBAGs. Asterisks (*) indicate correlations that remain significant after Bonferroni correction (0.05/7). Overall, phenotypic correlations exhibit more significant signals due to the use of individual-level data, in contrast to genetic correlations derived solely from GWAS summary statistics. Notably, genetic correlations tend to reflect their corresponding significant phenotypic correlations.

**eFigure 16: Sensitivity check analyses for the potential causal signal in our MR analyses**

We list here the full set of sensitivity check analysis results for the significant causal signals we found in our two causal networks: *BAG2DE* and *DE2BAG*

**a): *DE2BAG*: Hypertension (I9_HYPTENS) to the heart MRIBAG**

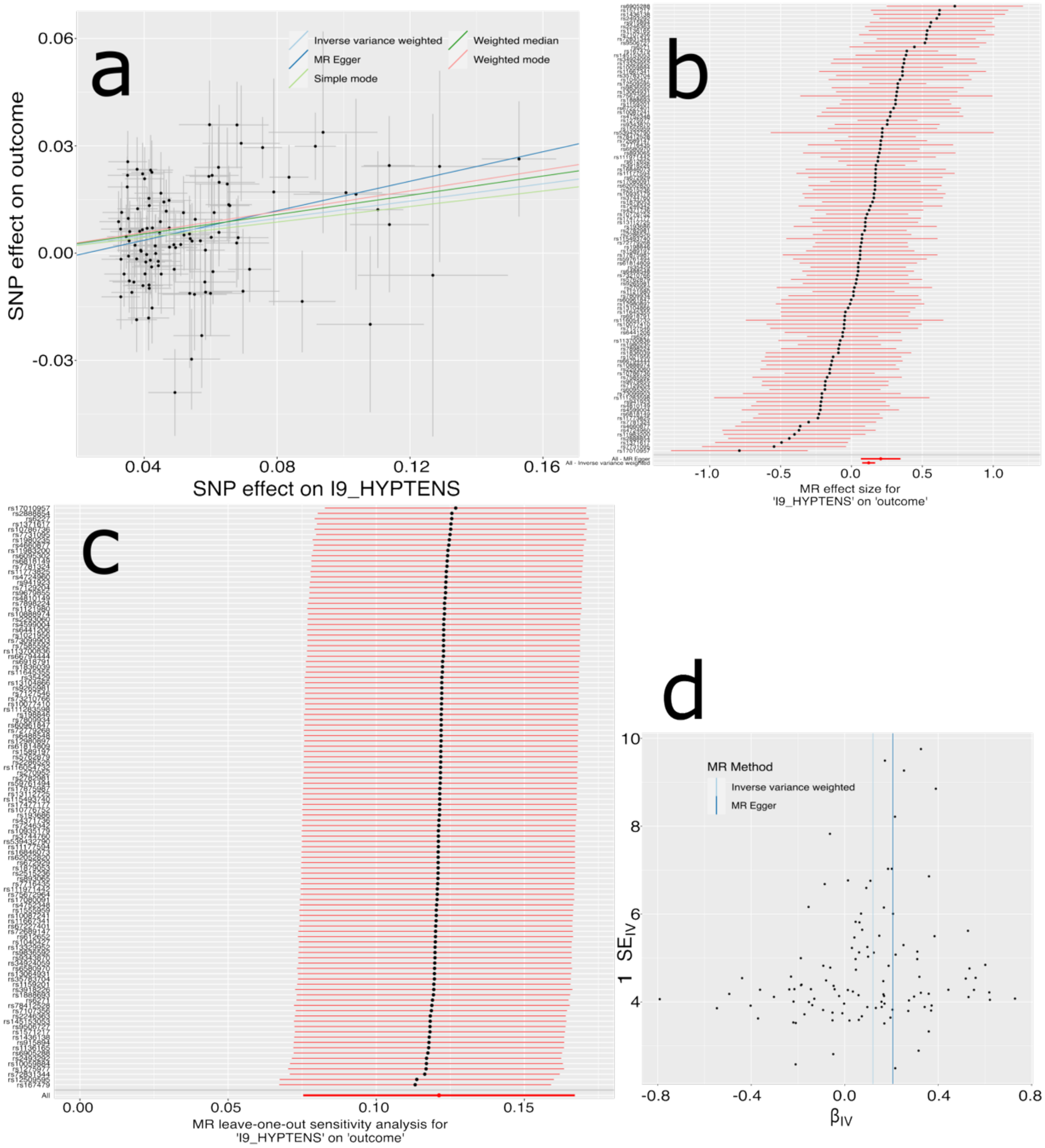


**a**) Scatter plot for the MR effect sizes of the SNP-I9_HYPTENS association (*x*-axis, log OR) and the SNP-heart MRIBAG associations (*y*-axis, log OR) with standard error bars. The slopes of the five lines correspond to the causal effect sizes estimated by the five MR estimators, respectively. There are no obvious outlier IVs. **b**) Forest plot for the single-SNP MR results. Each dot represents the MR effect (log OR), and the error bar displays the 95% CI for I9_HYPTENS on the heart MRIBAG using only one SNP; the red line shows the MR effect using all SNPs together for IVW and MR Egger estimators. **c**) Leave-one-SNP-out analysis of the I9_HYPTENS on the heart MRIBAG. Each dot represents the MR effect (log OR), and the error bar displays the 95% CI by excluding that SNP from the analysis. The red line depicts the IVW estimator using all SNPs. **d**) Funnel plot for the relationship between the causal effect of the

I9_HYPTENS on the heart MRIBAG. Each dot represents MR effect sizes estimated using each SNP as a separate instrument against the inverse of the standard error of the causal estimate. The Egger estimator yielded an intercept of -0.0046±0.0036 with a P-value of 0.21, suggesting no evidence of horizontal pleiotropy.

**b): *DE2BAG*: Hypertension (I9_HYPTENSESS) to the heart MRIBAG**

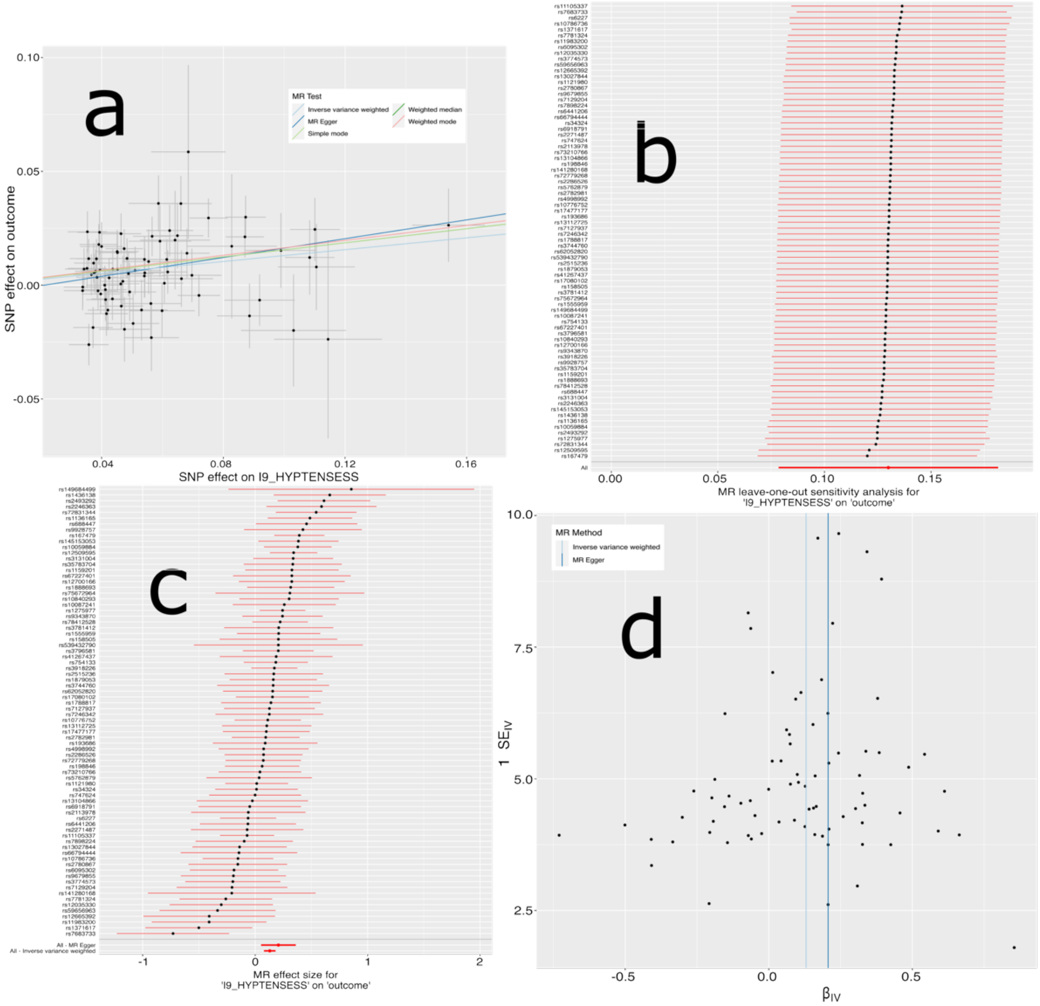


**a**) Scatter plot for the MR effect sizes of the SNP-I9_HYPTENSESS association (*x*-axis, log OR) and the SNP-heart MRIBAG associations (*y*-axis, log OR) with standard error bars. The slopes of the five lines correspond to the causal effect sizes estimated by the five MR estimators, respectively. There are no obvious outlier IVs. **b**) Leave-one-SNP-out analysis of the I9_HYPTENSESS on the heart MRIBAG. Each dot represents the MR effect (log OR), and the error bar displays the 95% CI by excluding that SNP from the analysis. The red line depicts the IVW estimator using all SNPs. **c**) Forest plot for the single-SNP MR results. Each dot represents the MR effect (log OR), and the error bar displays the 95% CI for I9_HYPTENSESS on the heart MRIBAG using only one SNP; the red line shows the MR effect using all SNPs together for IVW and MR Egger estimators. **d**) Funnel plot for the relationship between the causal effect of the I9_HYPTENSESS on the heart MRIBAG. Each dot represents MR effect sizes estimated using each SNP as a separate instrument against the inverse of the standard error of the causal estimate. The Egger estimator yielded an intercept of -0.004±0.004 with a P-value of 0.30, suggesting no evidence of horizontal pleiotropy.

**c): *DE2BAG*: Pregnancy hypertension (O15_HYPTENSPREG) to the heart MRIBAG**

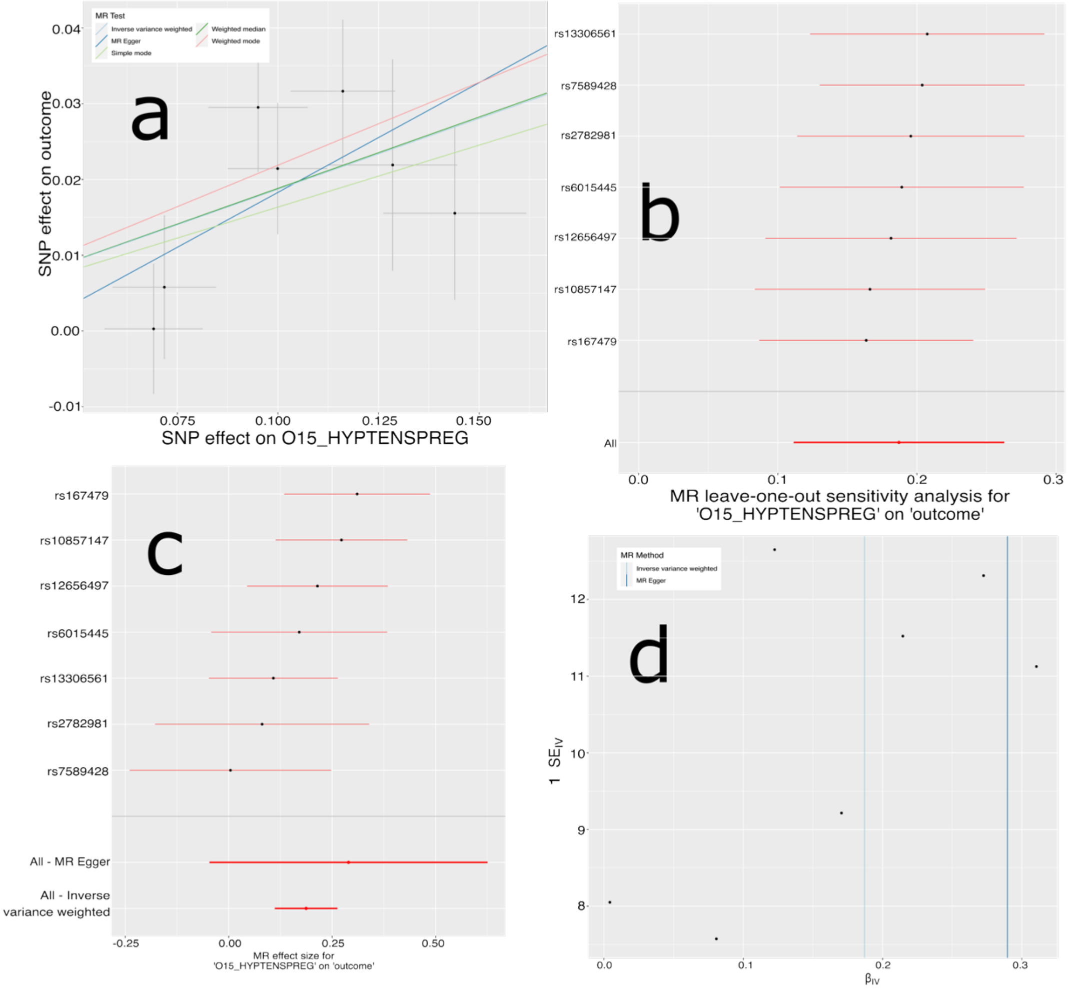


**a**) Scatter plot for the MR effect sizes of the SNP-O15_HYPTENSPREG association (*x*-axis, log OR) and the SNP-heart MRIBAG associations (*y*-axis, log OR) with standard error bars. The slopes of the five lines correspond to the causal effect sizes estimated by the five MR estimators, respectively. There are no obvious outlier IVs. **b**) Leave-one-SNP-out analysis of the O15_HYPTENSPREG on the heart MRIBAG. Each dot represents the MR effect (log OR), and the error bar displays the 95% CI by excluding that SNP from the analysis. The red line depicts the IVW estimator using all SNPs. **c**) Forest plot for the single-SNP MR results. Each dot represents the MR effect (log OR), and the error bar displays the 95% CI for O15_HYPTENSPREG on the heart MRIBAG using only one SNP; the red line shows the MR effect using all SNPs together for IVW and MR Egger estimators. **d**) Funnel plot for the relationship between the causal effect of the O15_HYPTENSPREG on the heart MRIBAG. Each dot represents MR effect sizes estimated using each SNP as a separate instrument against the inverse of the standard error of the causal estimate. The Egger estimator yielded an intercept of -0.01±0.01 with a P-value of 0.57, suggesting no evidence of horizontal pleiotropy.

**d): *DE2BAG*: Antihypertensive medication (RX_ANTIHYP) to the heart MRIBAG**

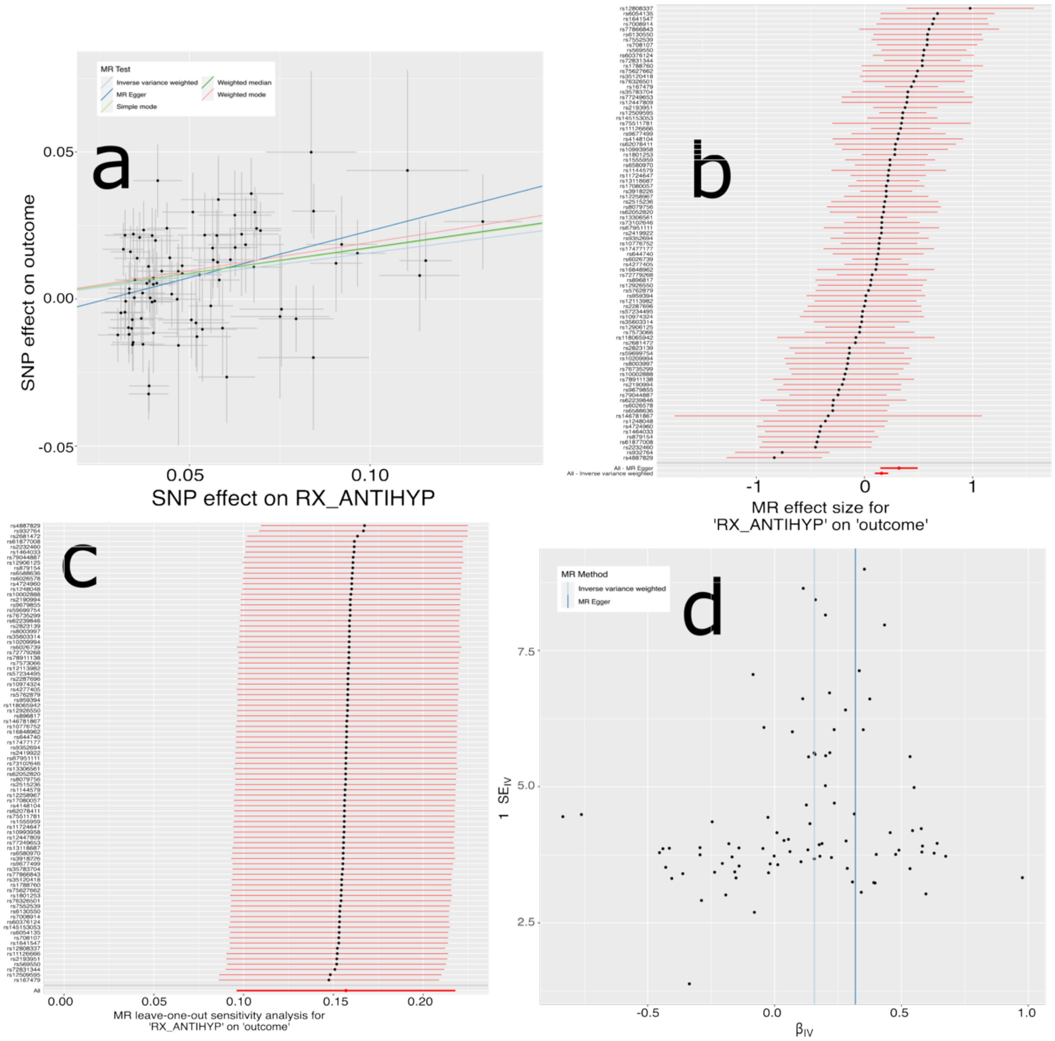


**a**) Scatter plot for the MR effect sizes of the SNP-RX_ANTIHYP association (*x*-axis, log OR) and the SNP-heart MRIBAG associations (*y*-axis, log OR) with standard error bars. The slopes of the five lines correspond to the causal effect sizes estimated by the five MR estimators, respectively. There are no obvious outlier IVs. **b**) Forest plot for the single-SNP MR results. Each dot represents the MR effect (log OR), and the error bar displays the 95% CI for RX_ANTIHYP on the heart MRIBAG using only one SNP; the red line shows the MR effect using all SNPs together for IVW and MR Egger estimators. **c**) Leave-one-SNP-out analysis of the RX_ANTIHYP on the heart MRIBAG. Each dot represents the MR effect (log OR), and the error bar displays the 95% CI by excluding that SNP from the analysis. The red line depicts the IVW estimator using all SNPs. **d**) Funnel plot for the relationship between the causal effect of the RX_ANTIHYP on the heart MRIBAG. Each dot represents MR effect sizes estimated using each SNP as a separate instrument against the inverse of the standard error of the causal estimate. The Egger estimator yielded an intercept of -0.008±0.004 with a P-value of 0.06, suggesting no evidence of horizontal pleiotropy.

**e): *DE2BAG*: AD 2 liver MRIBAG**

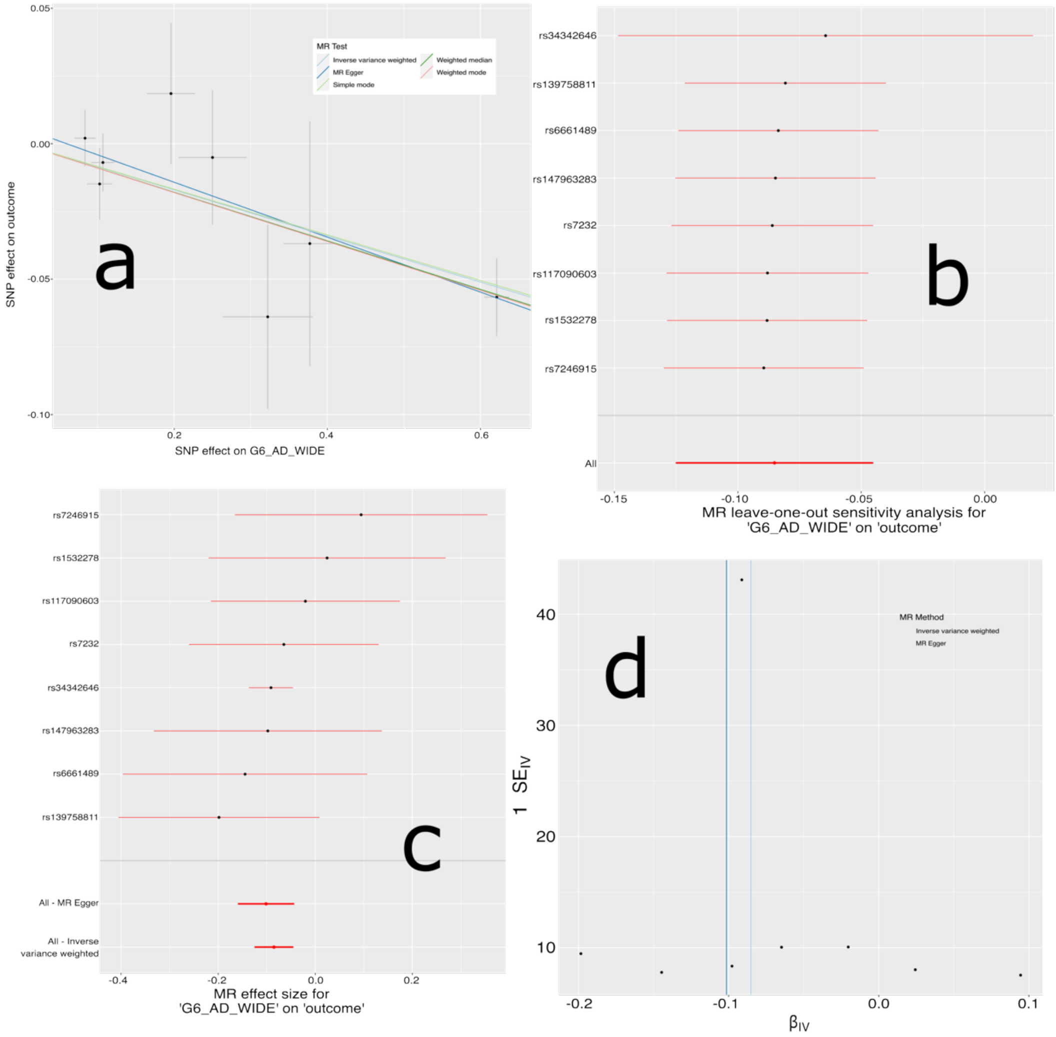


**a**) Scatter plot for the MR effect sizes of the AD association (*x*-axis, log OR) and the SNP-liver MRIBAG associations (*y*-axis, log OR) with standard error bars. The slopes of the five lines correspond to the causal effect sizes estimated by the five MR estimators, respectively. No obvious outlier was observed. **b**) Leave-one-SNP-out analysis of the AD on the liver MRIBAG. Each dot represents the MR effect (log OR), and the error bar displays the 95% CI by excluding that SNP from the analysis. **c**) Forest plot for the single-SNP MR results. Each dot represents the MR effect (log OR), and the error bar displays the 95% CI for AD on the liver MRIBAG using only one SNP; the red line shows the MR effect using all SNPs together for IVW and MR Egger estimators. **d**) Funnel plot for the relationship between the causal effect of the AD on the liver MRIBAG. Each dot represents MR effect sizes estimated using each SNP as a separate instrument against the inverse of the standard error of the causal estimate. The Egger estimator yielded an intercept of 0.006±0.0007 with a P-value of 0.47, suggesting no evidence of horizontal pleiotropy.

**f): *BAG2DE*: kidney MRIBAG to T2D (Wide definition)**

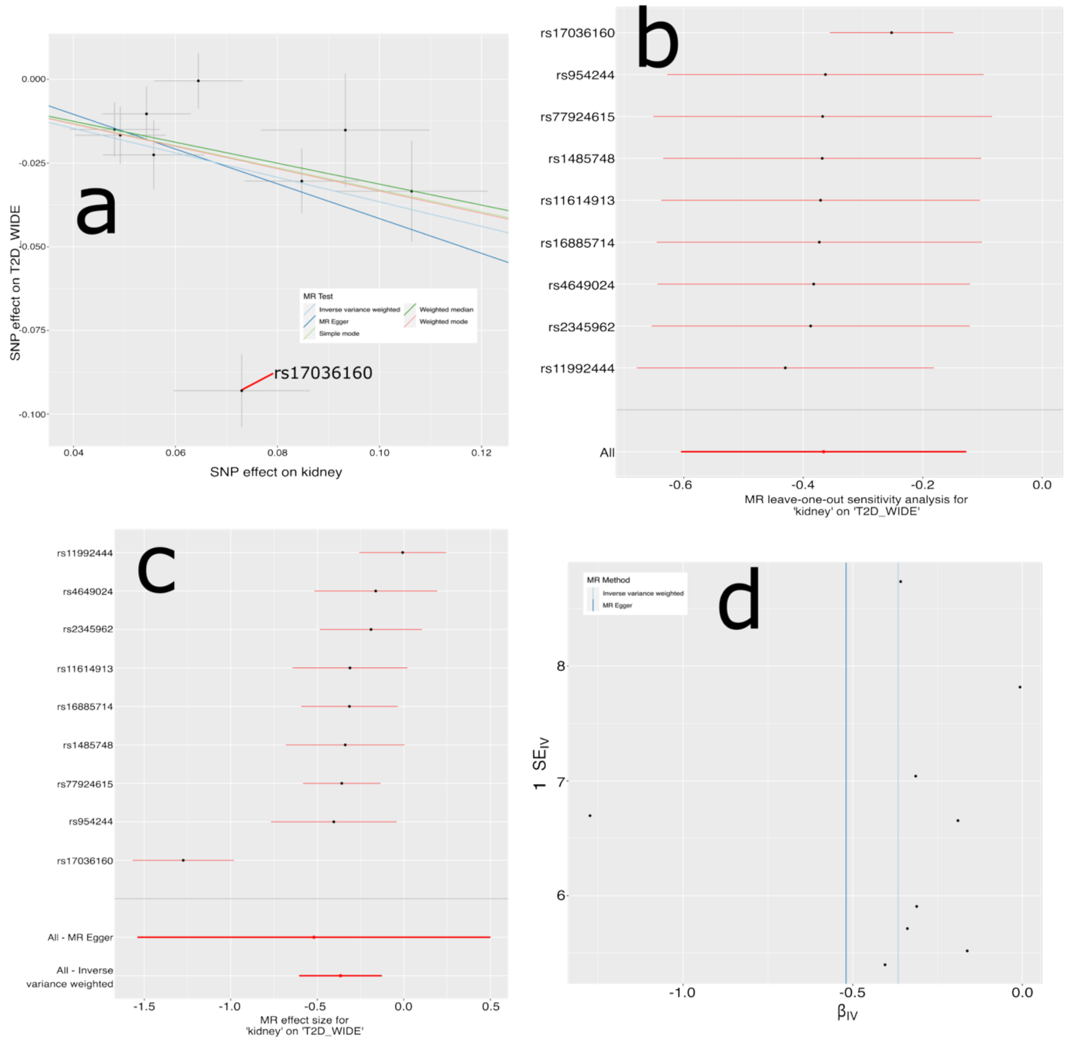


**a**) Scatter plot for the MR effect sizes of the T2D association (*x*-axis, log OR) and the SNP-kidney MRIBAG associations (*y*-axis, log OR) with standard error bars. The slopes of the five lines correspond to the causal effect sizes estimated by the five MR estimators, respectively. There exists one potential outlier IV (rs17036160); additional analysis excluding this IV resulted in a consistent signal (P-value=$1.79 \times 10^{-6}$). **b**) Leave-one-SNP-out analysis of the T2D on the kidney MRIBAG. Each dot represents the MR effect (log OR), and the error bar displays the 95% CI by excluding that SNP from the analysis. **c**) Forest plot for the single-SNP MR results. Each dot represents the MR effect (log OR), and the error bar displays the 95% CI for T2D on the kidney MRIBAG using only one SNP; the red line shows the MR effect using all SNPs together for IVW and MR Egger estimators. **d**) Funnel plot for the relationship between the causal effect of T2D on the kidney MRIBAG. Each dot represents MR effect sizes estimated using each SNP as a separate instrument against the inverse of the standard error of the causal estimate. The Egger estimator yielded an intercept of 0.01±0.04 with a P-value of 0.77, suggesting no evidence of horizontal pleiotropy. The MR results for kidney to T2D (narrow definition) are very similar to the T2D (wide).

**eFigure 17: Conceptualization of the endophenotype hypothesis along the multi-scale causal pathway of human aging and disease**

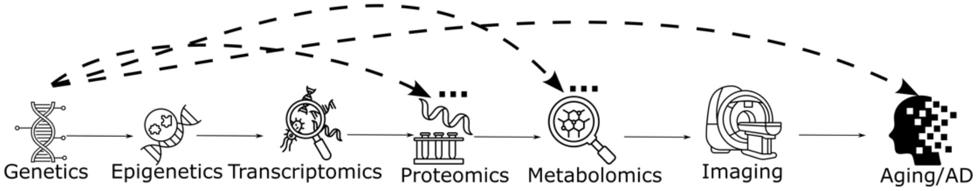


Multi-scale causal pathway for modeling human aging and disease, inspired by the endophenotype hypothesis[26–28], where the liability-index model (dotted arrow) and the mediation model (solid arrow) are illustrated.

**eFigure 18: Evidence of drug indications for 4 prioritized genes**

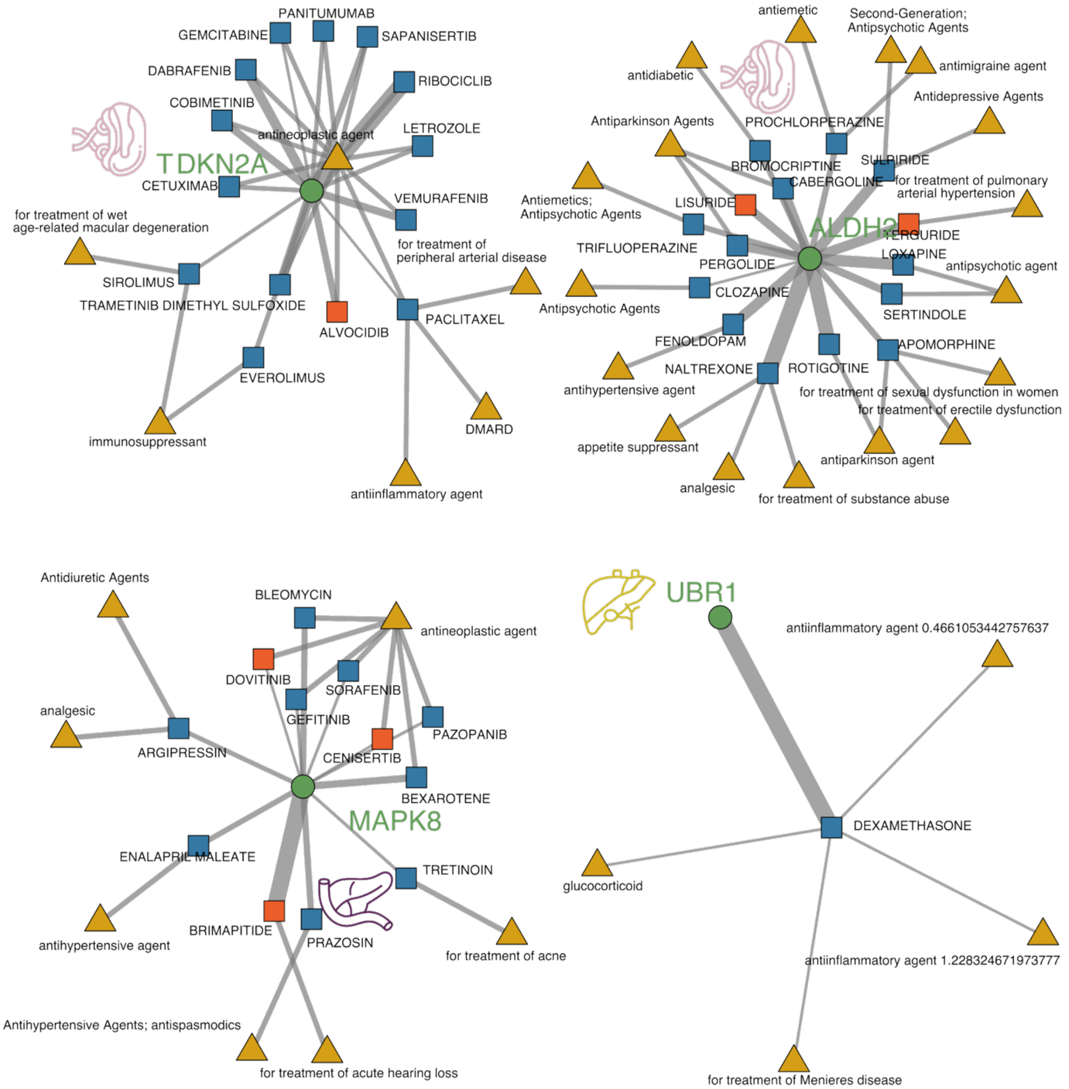


Evidence for drug indications for existing drug-gene interactions related to the 4 genes in the DGIdb platform (https://dgidb.org/).

**eFigure 19: The relationships among the liver MRIBAG, age at mortality, and liver volume (ID: 21080)**

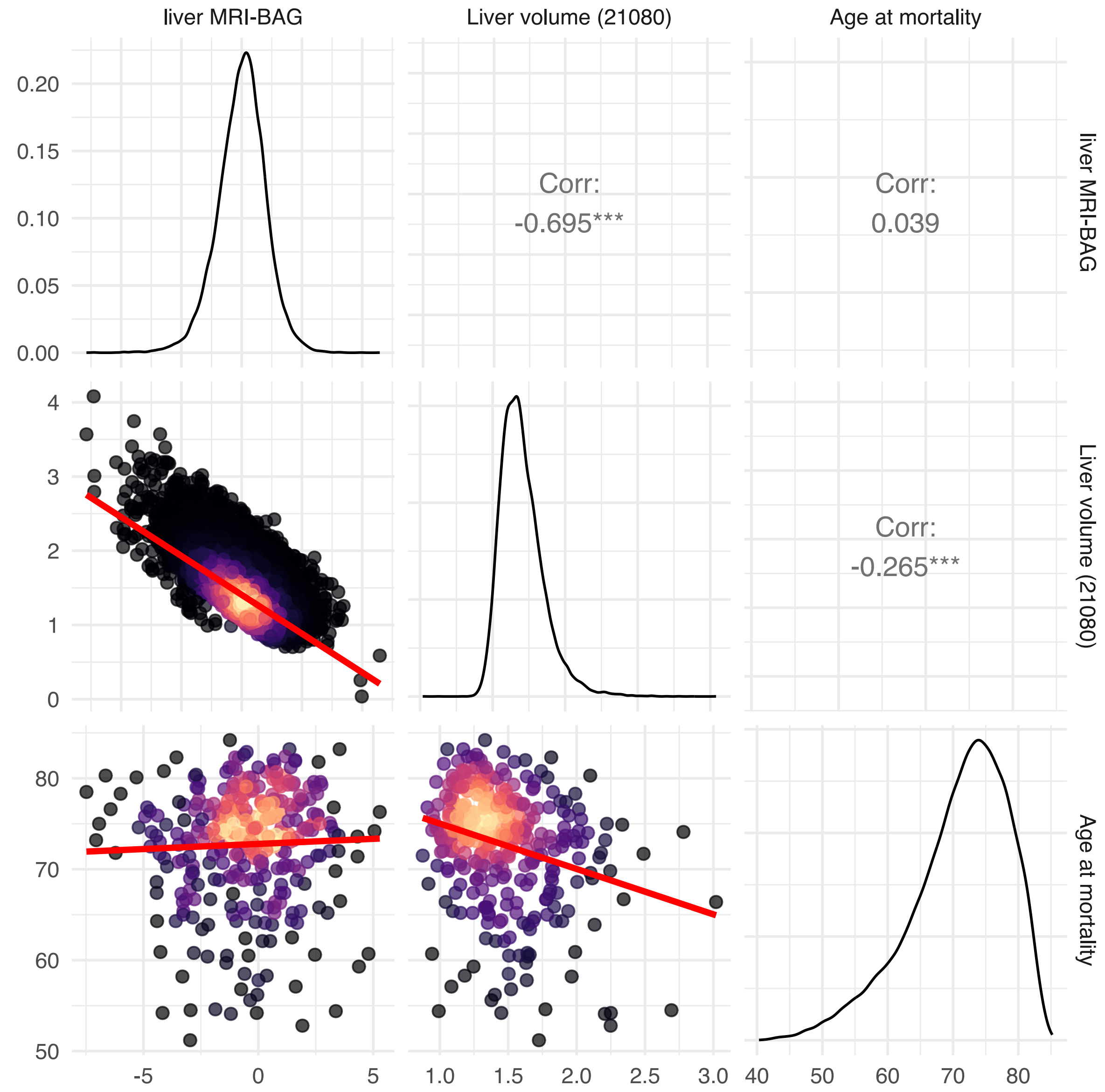


We show the pair-wise relationship between the liver MRIBAG, liver volume, and age at mortality. Of note, a simple linear regression between age at mortality and MRIBAG is different from a survival analysis using a Cox proportional hazards model in the main analyses. While linear regression examines the correlation between the MRIBAG and lifespan (age at mortality) directly, the Cox model provides insights into how the MRIBAG influences the risk of mortality over time (considering censoring).

**eFigure 20: The relationships among the spleen MRIBAG, age at mortality, and spleen volume (ID: 21083)**

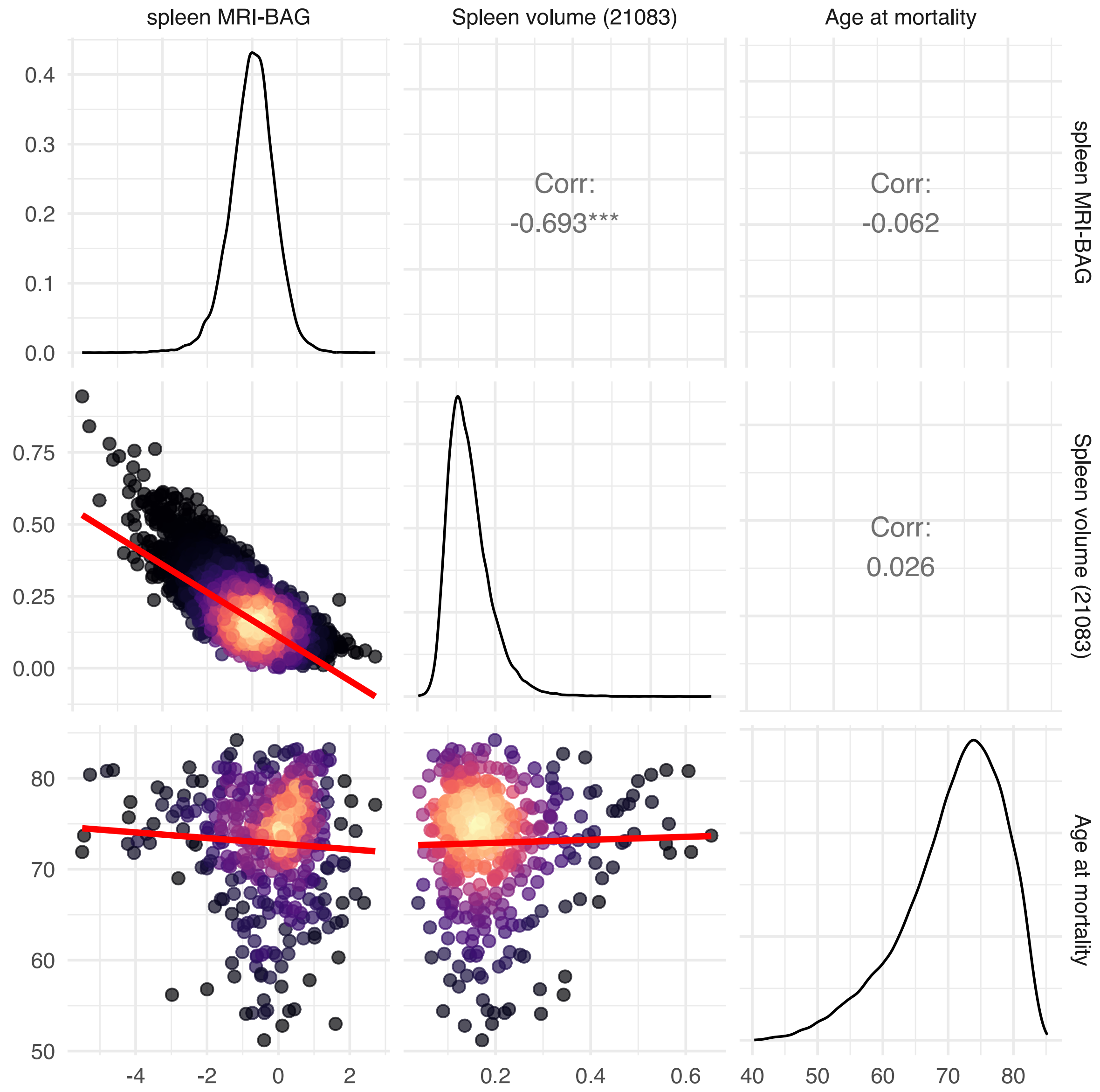


We show the pair-wise relationship between the spleen MRIBAG, spleen volume, and age at mortality. Of note, a simple linear regression between age at mortality and MRIBAG is different from a survival analysis using a Cox proportional hazards model in the main analyses. While linear regression examines the correlation between the MRIBAG and lifespan (age at mortality) directly, the Cox model provides insights into how the MRIBAG influences the risk of mortality over time (considering censoring).

**eFigure 21: Mediation pathways between the brain ProtBAG, brain MRIBAG, and two subtypes of late-life depression**

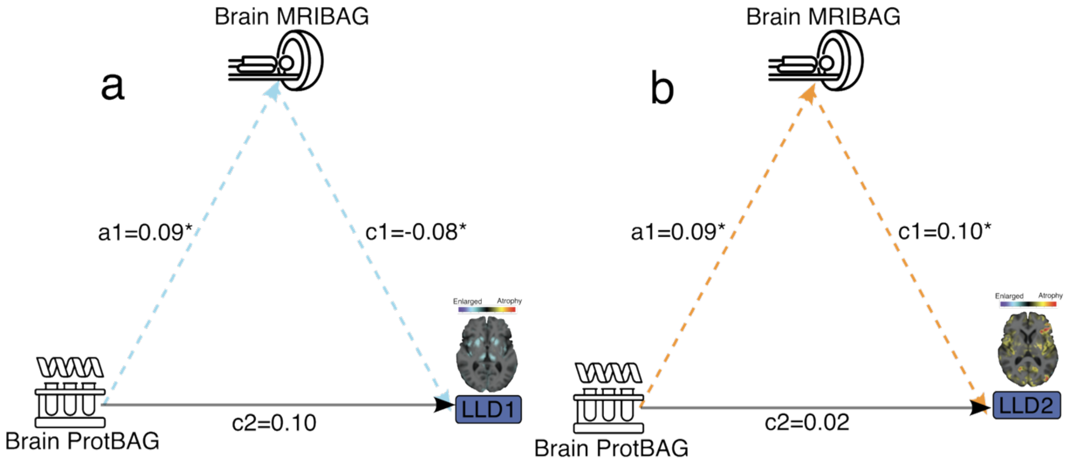


**a**) We first tested a mediation model where Brain MRIBAG served as a potential mediator linking molecular-level aging, represented by Brain ProtBAG, to late-life depression (LLD) subtypes (LLD1: reserved brain volume without atrophy). This model hypothesizes that proteomic aging influences neuroanatomical aging, which in turn affects LLD1. Significant indirect effects through MRIBAG would suggest that structural brain aging is a downstream pathway by which molecular aging contributes to depression risk. Covariates included age, sex, BMI, blood pressure (systolic and diastolic), and the first five genetic principal components. **b**) The same mediation model towards LLD2 as outcome (global brain atrophy).

**eFigure 22: Population selections and the nested cross-validation procedure to derive the 7 MRIBAGs**

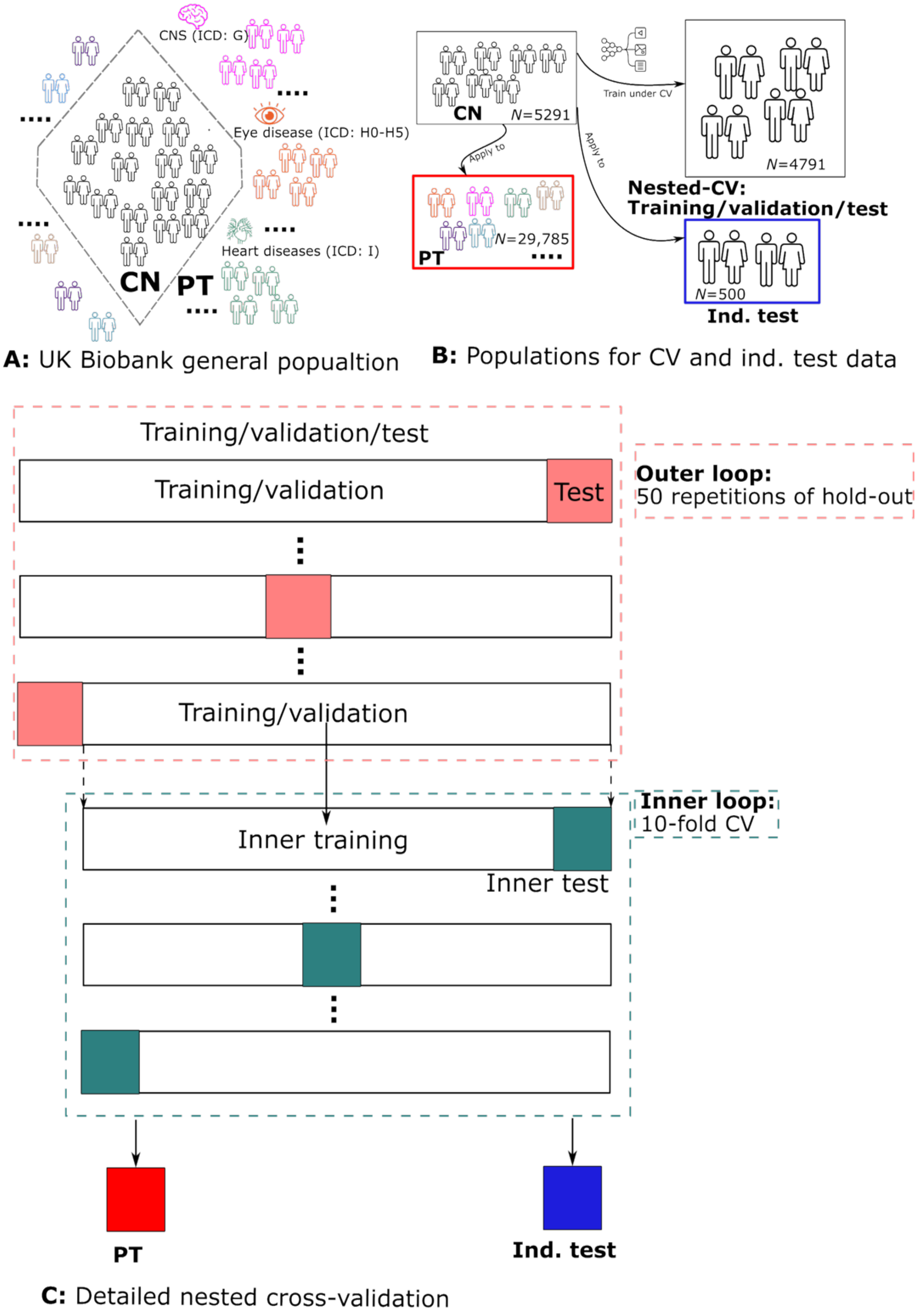


**a**) The UK Biobank is a UK-based general population. We define a healthy control population (CN) based on the ICD code and the in-patient medical history (i.e., participants that did not have any disease diagnosis records in the system), and a patient population (PT) (i.e., participants that had any single or multiple disease diagnosis records in the system). **b**) The CN population was further split into the CN independent test (ind. test; *N*=500) dataset and the CN nested CV dataset (*N*=4791 for training/validation/test datasets, using the heart MRIBAG as an example). The ML models were trained on the CN nested-CV dataset and then applied to the CN ind. test dataset and the PT dataset (*N*=29,785). **c**) Details on the nested CV procedure include a repeated hold-out cross-validation with 50 repetitions (80% training/validation and 20% testing) for the outer loop and 10-fold cross-validation for the inner loop for hyperparameter selection (e.g., *C* for SVR).

**eFigure 23: Feature importance in deriving the brain MRIBAG in the UK Biobank, BLSA, and A4**

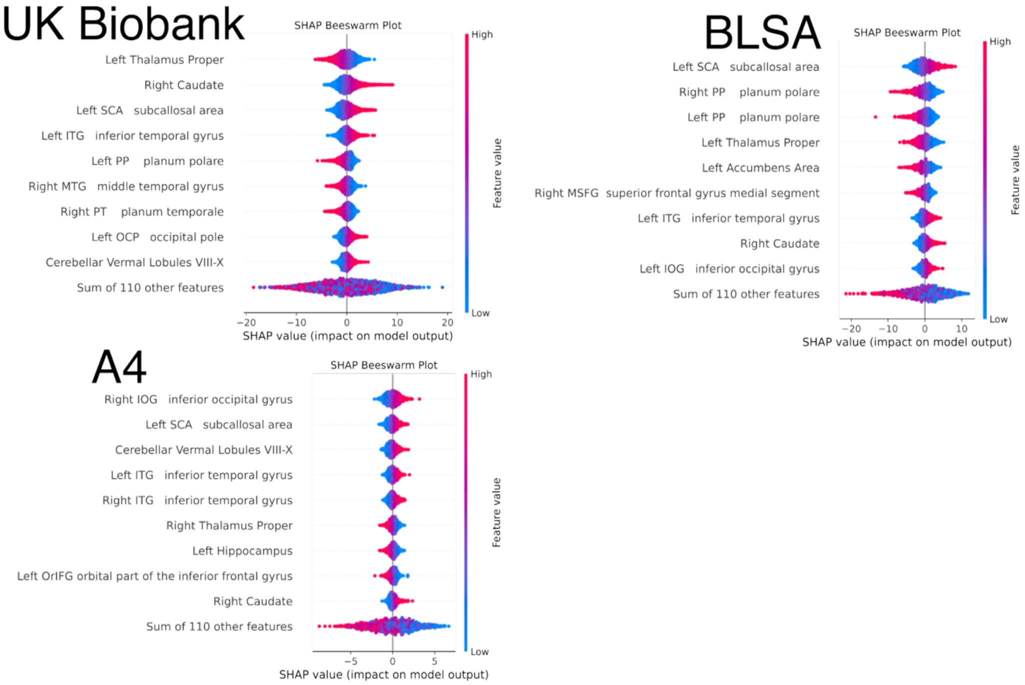


SHAP analysis revealed that the most influential features contributing to brain MRIBAGs were largely consistent across the three independent studies.

**eTable 1: The characteristics of participants consolidated via the MULTI consortium**

| Data type | Omics type | | Study | *N* (313,645) | Age [year (mean/std)] | | | Sex (% female) | | |
|---|---|---|---|---|---|---|---|---|---|---|
| | | | | | CN[b] training/validation/test [*N*] | CN ind. test [*N*] | PT [*N*] | CN training/validation/test | CN ind. test | PT |
| Individual-level | MRI | Brain[a] | UKBB | 37,226 | 63.06±8.69 [6327] | 63.13±8.32 [500] | 64.23±9.03 [30,399] | 3226/51% | 249/50% | 15120/50% |
| | | Adipose | | 24,494 | 62.87±7.16 [3573] | 63.18±7.13 [500] | 64.50±7.48 [20,421] | 1755/49% | 253/51% | 10,812/53% |
| | | Heart | | 35,576 | 62.13±7.21 [5291] | 62.28±7.16 [500] | 63.76±7.60 [29,785] | 2570/49% | 230/46% | 15,512/52% |
| | | Kidney | | 34,560 | 62.47±7.16 [5098] | 62.40±6.98 [500] | 64.11±7.54 [28,962] | 2554/50% | 238/48% | 15,293/53% |
| | | Liver | | 24,650 | 62.90±7.08 [3612] | 63.10±7.10 [500] | 64.58±7.49 [20,538] | 1808/50% | 240/48% | 10,925/53% |
| | | Pancreas | | 28,015 | 62.76±7.14 [4120] | 62.74±7.41 [500] | 64.38±7.50 [23,395] | 2014/49% | 247/49% | 12,167/52% |
| | | Spleen | | 27,017 | 62.94±7.13 [3958] | 63.08±7.24 [500] | 64.60±7.49 [22,559] | 1967/50% | 240/48% | 11,892/53% |
| | | Brain | A4 | 1055 | 72.10±4.75 | | | 798/45% | | |
| | | | BLSA | 1114 | 65.44±14.11 | | | 589/53% | | |
| | Proteomics | Olink | UKBB | 50,316 | 64.16±8.04 | | | 28,581/54% | | |
| | | SomaScan | BLSA | 909 | 65.30±14.76 | | | 500/55% | | |
| | Metabolomics | | UKBB | 274,247 | 56.56±8.08 | | | 147,994/54% | | |
| | Genetics | | UKBB | 311,476 | 56.51±8.07 | | | 167,811/54% | | |
| Summary-level | Genetics | Summary statistics | FinnGen | 521[c] | NA | | | NA | | |
| | | | PGC | 4[c] | NA | | | NA | | |

[a]We re-trained the brain MRIBAG[12] using 119 gray matter volumes using the same cross-validation procedure in this study.
[b]CN was defined as participants with no recorded disease diagnoses following their enrollment in the UK Biobank study.

**eTable 2: Biological age prediction performance before age bias correction**

**a) Lasso and SVR**

In the main manuscript, we present the results before age bias correction; **bolded results** are used for downstream ProWAS, MetWAS, GWAS, and prediction analyses. The models were implemented in MLNI v0.1.2.

| Organ-Dataset-ML | MAE | *r* | p |
|---|---|---|---|
| **Adipose-Train-Lasso** | 4.944396 | 0.539851 | 1.7E-269 |
| **Adipose-Ind.test-Lasso** | 5.738238 | 0.494719 | 3.17E-32 |
| **Brain-Train-Lasso** | 4.869818 | 0.772594 | 0 |
| **Brain-Ind.test-Lasso** | 4.982525 | 0.777984 | 1.19E-73 |
| Heart-Train-Lasso | 4.139974 | 0.695673 | 0 |
| Heart-Ind.test-Lasso | 5.086422 | 0.664608 | 5E-65 |
| **Kidney-Train-Lasso** | 5.6641 | 0.277304 | 1.15E-90 |
| **Kidney-Ind.test-Lasso** | 6.465428 | 0.242908 | 3.79E-08 |
| Liver-Train-Lasso | 5.616313 | 0.277039 | 1.2E-64 |
| Liver-Ind.test-Lasso | 5.545405 | 0.3545 | 2.98E-16 |
| Pancreas-Train-Lasso | 5.680515 | 0.268304 | 7.22E-69 |
| Pancreas-Ind.test-Lasso | 5.861792 | 0.278355 | 2.39E-10 |
| Spleen-Train-Lasso | 5.817169 | 0.187967 | 8.44E-33 |
| Spleen-Ind.test-Lasso | 5.877528 | 0.22724 | 2.81E-07 |
| Adipose-Train-SVR | 4.94216 | 0.537175 | 2.5E-266 |
| Adipose-Ind.test-SVR | 6.198629 | 0.488766 | 2.19E-31 |
| Brain-Train-SVR | 4.888453 | 0.799565 | 0 |
| Brain-Ind.test-SVR | 5.123755 | 0.770636 | 4.21E-72 |
| **Heart-Train-SVR** | 4.118961 | 0.692264 | 0 |
| **Heart-Ind.test-SVR** | 4.590152 | 0.662421 | 1.82E-64 |
| Kidney-Train-SVR | 5.647089 | 0.276444 | 4.3E-90 |
| Kidney-Ind.test-SVR | 6.868699 | 0.238618 | 6.64E-08 |
| **Liver-Train-SVR** | 5.596128 | 0.277354 | 8.53E-65 |
| **Liver-Ind.test-SVR** | 5.552195 | 0.35448 | 3E-16 |
| **Pancreas-Train-SVR** | 5.682083 | 0.268333 | 6.97E-69 |
| **Pancreas-Ind.test-SVR** | 5.850628 | 0.279958 | 1.86E-10 |
| **Spleen-Train-SVR** | 5.816762 | 0.187035 | 1.73E-32 |
| **Spleen-Ind.test-SVR** | 5.858165 | 0.231024 | 1.75E-07 |

**b) Elastic and NN**

We performed additional sensitivity check analyses by applying the data to Elastic Net regression to better handle overfitting by balancing the L1 and L2 regularization terms, and neural network (NN) for modeling non-linearity, implemented in MLNI v0.1.5.1.

| Organ-Dataset-ML | MAE | *r* | p |
|---|---|---|---|
| Adipose-Train-Elastic | 4.949839 | 0.539057 | 1.5E-268 |
| Adipose-Ind.test-Elastic | 6.128813 | 0.493694 | 4.44E-32 |
| Brain-Train-Elastic | 4.918353 | 0.773361 | 0 |
| Brain-Ind.test-Elastic | 4.259515 | 0.696478 | 8.55E-74 |
| Heart-Train-Elastic | 4.141086 | 0.695663 | 0 |
| Heart-Ind.test-Elastic | 4.89329 | 0.665485 | 2.97E-65 |
| Kidney-Train-Elastic | 5.678625 | 0.276802 | 2.49E-90 |
| Kidney-Ind.test-Elastic | 6.184931 | 0.248603 | 1.76E-08 |
| Liver-Train-Elastic | 5.605487 | 0.277559 | 6.83E-65 |
| Liver-Ind.test-Elastic | 5.530138 | 0.3571 | 1.75E-16 |
| Pancreas-Train-Elastic | 5.675544 | 0.269325 | 2.13E-69 |
| Pancreas-Ind.test-Elastic | 5.874641 | 0.278146 | 2.46E-10 |
| Spleen-Train-Elastic | 5.8172 | 0.187967 | 8.44E-33 |
| Spleen-Ind.test-Elastic | 5.877513 | 0.227291 | 2.79E-07 |

| | | | |
|---|---|---|---|
| Adipose-Train-NN | 4.984978 | 0.53263 | 4.7E-261 |
| Adipose-Ind.test-NN | 5.495602 | 0.498159 | 1.02E-32 |
| Brain-Train-NN | 4.295656 | 0.799216 | 0 |
| Brain-Ind.test-NN | 4.936704 | 0.661322 | 3.48E-64 |
| Heart-Train-NN | 3.890294 | 0.718948 | 0 |
| Heart-Ind.test-NN | 5.281431 | 0.600198 | 2.96E-50 |
| Kidney-Train-NN | 5.64669 | 0.280833 | 4.92E-93 |
| Kidney-Ind.test-NN | 6.377033 | 0.234878 | 1.08E-07 |
| Liver-Train-NN | 5.555967 | 0.294589 | 3.08E-73 |
| Liver-Ind.test-NN | 5.472893 | 0.376666 | 2.67E-18 |
| Pancreas-Train-NN | 5.645217 | 0.27979 | 5.67E-75 |
| Pancreas-Ind.test-NN | 5.959289 | 0.273484 | 5.01E-10 |
| Spleen-Train-NN | 5.777958 | 0.210128 | 9.68E-41 |
| Spleen-Ind.test-NN | 5.833968 | 0.237369 | 7.81E-08 |

**c) Elastic net comparison with all available feature set (All) and non-correlated feature set (Uncorr)**

Using the kidney MRIBAG for which we found highly correlated features (shown in **eFigure 3**), we performed additional sensitivity check analyses to compare the results using i) the full set of imaging features (with correlated features) versus ii) a curated set of imaging features (without correlated features).

| **Feature set** | **MAE** | **r** | **p** |
|---|---|---|---|
| Kidney-Train-kidney-Uncorr | 5.678625 | 0.276802 | 2.49E-90 |
| Kidney-Ind.test-kidney_Uncorr | 6.184931 | 0.248603 | 1.76E-08 |
| Kidney-Train-kidney-All | 5.601778 | 0.309668 | 1E-113 |
| Kidney-Ind.test-kidney-All | 7.254153 | 0.269119 | 9.61E-10 |

992 **eTable 3: The description of the 327 metabolites from the UKBB NMR platform**

| Biomarker | Description | Units | Group | Sub-group | Type | UKB Field ID | QC Flag Field ID |
|---|---|---|---|---|---|---|---|
| Total_C | Total Cholesterol | mmol/L | Cholesterol | NA | Composite | 23400 | 23700 |
| non_HDL_C | Total Cholesterol Minus HDL-C | mmol/L | Cholesterol | NA | Composite | 23401 | 23701 |
| Remnant_C | Remnant Cholesterol (Non-HDL, Non-LDL -Cholesterol) | mmol/L | Cholesterol | NA | Composite | 23402 | 23702 |
| VLDL_C | VLDL Cholesterol | mmol/L | Cholesterol | NA | Composite | 23403 | 23703 |
| Clinical_LDL_C | Clinical LDL Cholesterol | mmol/L | Cholesterol | NA | Non-derived | 23404 | 23704 |
| LDL_C | LDL Cholesterol | mmol/L | Cholesterol | NA | Composite | 23405 | 23705 |
| HDL_C | HDL Cholesterol | mmol/L | Cholesterol | NA | Composite | 23406 | 23706 |
| Total_TG | Total Triglycerides | mmol/L | Triglycerides | NA | Composite | 23407 | 23707 |
| VLDL_TG | Triglycerides in VLDL | mmol/L | Triglycerides | NA | Composite | 23408 | 23708 |
| LDL_TG | Triglycerides in LDL | mmol/L | Triglycerides | NA | Composite | 23409 | 23709 |
| HDL_TG | Triglycerides in HDL | mmol/L | Triglycerides | NA | Composite | 23410 | 23710 |
| Total_PL | Total Phospholipids in Lipoprotein Particles | mmol/L | Phospholipids | NA | Composite | 23411 | 23711 |
| VLDL_PL | Phospholipids in VLDL | mmol/L | Phospholipids | NA | Composite | 23412 | 23712 |
| LDL_PL | Phospholipids in LDL | mmol/L | Phospholipids | NA | Composite | 23413 | 23713 |
| HDL_PL | Phospholipids in HDL | mmol/L | Phospholipids | NA | Composite | 23414 | 23714 |
| Total_CE | Total Esterified Cholesterol | mmol/L | Cholesteryl esters | NA | Composite | 23415 | 23715 |
| VLDL_CE | Cholesteryl Esters in VLDL | mmol/L | Cholesteryl esters | NA | Composite | 23416 | 23716 |
| LDL_CE | Cholesteryl Esters in LDL | mmol/L | Cholesteryl esters | NA | Composite | 23417 | 23717 |
| HDL_CE | Cholesteryl Esters in HDL | mmol/L | Cholesteryl esters | NA | Composite | 23418 | 23718 |
| Total_FC | Total Free Cholesterol | mmol/L | Free cholesterol | NA | Composite | 23419 | 23719 |
| VLDL_FC | Free Cholesterol in VLDL | mmol/L | Free cholesterol | NA | Composite | 23420 | 23720 |
| LDL_FC | Free Cholesterol in LDL | mmol/L | Free cholesterol | NA | Composite | 23421 | 23721 |
| HDL_FC | Free Cholesterol in HDL | mmol/L | Free cholesterol | NA | Composite | 23422 | 23722 |
| Total_L | Total Lipids in Lipoprotein Particles | mmol/L | Total lipids | NA | Composite | 23423 | 23723 |
| VLDL_L | Total Lipids in VLDL | mmol/L | Total lipids | NA | Composite | 23424 | 23724 |
| LDL_L | Total Lipids in LDL | mmol/L | Total lipids | NA | Composite | 23425 | 23725 |
| HDL_L | Total Lipids in HDL | mmol/L | Total lipids | NA | Composite | 23426 | 23726 |
| Total_P | Total Concentration of Lipoprotein Particles | mmol/L | Lipoprotein particle concentrations | NA | Composite | 23427 | 23727 |
| VLDL_P | Concentration of VLDL Particles | mmol/L | Lipoprotein particle concentrations | NA | Composite | 23428 | 23728 |
| LDL_P | Concentration of LDL Particles | mmol/L | Lipoprotein particle concentrations | NA | Composite | 23429 | 23729 |
| HDL_P | Concentration of HDL Particles | mmol/L | Lipoprotein particle concentrations | NA | Composite | 23430 | 23730 |
| VLDL_size | Average Diameter for VLDL Particles | nm | Lipoprotein particle sizes | NA | Non-derived | 23431 | 23731 |
| LDL_size | Average Diameter for LDL Particles | nm | Lipoprotein particle sizes | NA | Non-derived | 23432 | 23732 |
| HDL_size | Average Diameter for HDL Particles | nm | Lipoprotein particle sizes | NA | Non-derived | 23433 | 23733 |

| | | | | | | | |
|---|---|---|---|---|---|---|---|
| Phosphoglyc | Phosphoglycerides | mmol/L | Other lipids | NA | Non-derived | 23434 | 23734 |
| TG_by_PG | Triglycerides to Phosphoglycerides ratio | ratio | Other lipids | NA | Ratio | 23435 | 23735 |
| Cholines | Total Cholines | mmol/L | Other lipids | NA | Non-derived | 23436 | 23736 |
| Phosphatidylc | Phosphatidylcholines | mmol/L | Other lipids | NA | Non-derived | 23437 | 23737 |
| Sphingomyelins | Sphingomyelins | mmol/L | Other lipids | NA | Non-derived | 23438 | 23738 |
| ApoB | Apolipoprotein B | g/l | Apolipoproteins | NA | Non-derived | 23439 | 23739 |
| ApoA1 | Apolipoprotein A1 | g/l | Apolipoproteins | NA | Non-derived | 23440 | 23740 |
| ApoB_by_ApoA1 | Apolipoprotein B to Apolipoprotein A1 ratio | ratio | Apolipoproteins | NA | Ratio | 23441 | 23741 |
| Total_FA | Total Fatty Acids | mmol/L | Fatty acids | NA | Composite | 23442 | 23742 |
| Unsaturation | Degree of Unsaturation | degree | Fatty acids | NA | Non-derived | 23443 | 23743 |
| Omega_3 | Omega-3 Fatty Acids | mmol/L | Fatty acids | NA | Non-derived | 23444 | 23744 |
| Omega_6 | Omega-6 Fatty Acids | mmol/L | Fatty acids | NA | Non-derived | 23445 | 23745 |
| PUFA | Polyunsaturated Fatty Acids | mmol/L | Fatty acids | NA | Composite | 23446 | 23746 |
| MUFA | Monounsaturated Fatty Acids | mmol/L | Fatty acids | NA | Non-derived | 23447 | 23747 |
| SFA | Saturated Fatty Acids | mmol/L | Fatty acids | NA | Non-derived | 23448 | 23748 |
| LA | Linoleic Acid | mmol/L | Fatty acids | NA | Non-derived | 23449 | 23749 |
| DHA | Docosahexaenoic Acid | mmol/L | Fatty acids | NA | Non-derived | 23450 | 23750 |
| Omega_3_pct | Omega-3 Fatty Acids to Total Fatty Acids percentage | % | Fatty acids | Fatty acid ratios | Percentage | 23451 | 23751 |
| Omega_6_pct | Omega-6 Fatty Acids to Total Fatty Acids percentage | % | Fatty acids | Fatty acid ratios | Percentage | 23452 | 23752 |
| PUFA_pct | Polyunsaturated Fatty Acids to Total Fatty Acids percentage | % | Fatty acids | Fatty acid ratios | Percentage | 23453 | 23753 |
| MUFA_pct | Monounsaturated Fatty Acids to Total Fatty Acids percentage | % | Fatty acids | Fatty acid ratios | Percentage | 23454 | 23754 |
| SFA_pct | Saturated Fatty Acids to Total Fatty Acids percentage | % | Fatty acids | Fatty acid ratios | Percentage | 23455 | 23755 |
| LA_pct | Linoleic Acid to Total Fatty Acids percentage | % | Fatty acids | Fatty acid ratios | Percentage | 23456 | 23756 |
| DHA_pct | Docosahexaenoic Acid to Total Fatty Acids percentage | % | Fatty acids | Fatty acid ratios | Percentage | 23457 | 23757 |
| PUFA_by_MUFA | Polyunsaturated Fatty Acids to Monounsaturated Fatty Acids ratio | ratio | Fatty acids | Fatty acid ratios | Ratio | 23458 | 23758 |
| Omega_6_by_Omega_3 | Omega-6 Fatty Acids to Omega-3 Fatty Acids ratio | ratio | Fatty acids | Fatty acid ratios | Ratio | 23459 | 23759 |
| Ala | Alanine | mmol/L | Amino acids | NA | Non-derived | 23460 | 23760 |
| Gln | Glutamine | mmol/L | Amino acids | NA | Non-derived | 23461 | 23761 |

| | | | | | | | |
|---|---|---|---|---|---|---|---|
| Gly | Glycine | mmol/L | Amino acids | NA | Non-derived | 23462 | 23762 |
| His | Histidine | mmol/L | Amino acids | NA | Non-derived | 23463 | 23763 |
| Total_BCAA | Total Concentration of Branched-Chain Amino Acids (Leucine + Isoleucine + Valine) | mmol/L | Amino acids | Branched-chain amino acids | Composite | 23464 | 23764 |
| Ile | Isoleucine | mmol/L | Amino acids | Branched-chain amino acids | Non-derived | 23465 | 23765 |
| Leu | Leucine | mmol/L | Amino acids | Branched-chain amino acids | Non-derived | 23466 | 23766 |
| Val | Valine | mmol/L | Amino acids | Branched-chain amino acids | Non-derived | 23467 | 23767 |
| Phe | Phenylalanine | mmol/L | Amino acids | Aromatic amino acids | Non-derived | 23468 | 23768 |
| Tyr | Tyrosine | mmol/L | Amino acids | Aromatic amino acids | Non-derived | 23469 | 23769 |
| Glucose | Glucose | mmol/L | Glycolysis related metabolites | NA | Non-derived | 23470 | 23770 |
| Lactate | Lactate | mmol/L | Glycolysis related metabolites | NA | Non-derived | 23471 | 23771 |
| Pyruvate | Pyruvate | mmol/L | Glycolysis related metabolites | NA | Non-derived | 23472 | 23772 |
| Citrate | Citrate | mmol/L | Glycolysis related metabolites | NA | Non-derived | 23473 | 23773 |
| bOHbutyrate | 3-Hydroxybutyrate | mmol/L | Ketone bodies | NA | Non-derived | 23474 | 23774 |
| Acetate | Acetate | mmol/L | Ketone bodies | NA | Non-derived | 23475 | 23775 |
| Acetoacetate | Acetoacetate | mmol/L | Ketone bodies | NA | Non-derived | 23476 | 23776 |
| Acetone | Acetone | mmol/L | Ketone bodies | NA | Non-derived | 23477 | 23777 |
| Creatinine | Creatinine | mmol/L | Fluid balance | NA | Non-derived | 23478 | 23778 |
| Albumin | Albumin | g/l | Fluid balance | NA | Non-derived | 23479 | 23779 |
| GlycA | Glycoprotein Acetyls | mmol/L | Inflammation | NA | Non-derived | 23480 | 23780 |
| XXL_VLDL_P | Concentration of Chylomicrons and Extremely Large VLDL Particles | mmol/L | Lipoprotein subclasses | Chylomicrons and extremely large VLDL (particle diameters from 75 nm upwards) | Non-derived | 23481 | 23781 |
| XXL_VLDL_L | Total Lipids in Chylomicrons and Extremely Large VLDL | mmol/L | Lipoprotein subclasses | Chylomicrons and extremely large VLDL (particle diameters from 75 nm upwards) | Composite | 23482 | 23782 |
| XXL_VLDL_PL | Phospholipids in Chylomicrons and Extremely Large VLDL | mmol/L | Lipoprotein subclasses | Chylomicrons and extremely large VLDL (particle diameters from 75 nm upwards) | Non-derived | 23483 | 23783 |

| | | | | | | | |
|---|---|---|---|---|---|---|---|
| XXL_VLDL_C | Cholesterol in Chylomicrons and Extremely Large VLDL | mmol/L | Lipoprotein subclasses | Chylomicrons and extremely large VLDL (particle diameters from 75 nm upwards) | Composite | 23484 | 23784 |
| XXL_VLDL_CE | Cholesteryl Esters in Chylomicrons and Extremely Large VLDL | mmol/L | Lipoprotein subclasses | Chylomicrons and extremely large VLDL (particle diameters from 75 nm upwards) | Non-derived | 23485 | 23785 |
| XXL_VLDL_FC | Free Cholesterol in Chylomicrons and Extremely Large VLDL | mmol/L | Lipoprotein subclasses | Chylomicrons and extremely large VLDL (particle diameters from 75 nm upwards) | Non-derived | 23486 | 23786 |
| XXL_VLDL_TG | Triglycerides in Chylomicrons and Extremely Large VLDL | mmol/L | Lipoprotein subclasses | Chylomicrons and extremely large VLDL (particle diameters from 75 nm upwards) | Non-derived | 23487 | 23787 |
| XL_VLDL_P | Concentration of Very Large VLDL Particles | mmol/L | Lipoprotein subclasses | Very large VLDL (average diameter 64 nm) | Non-derived | 23488 | 23788 |
| XL_VLDL_L | Total Lipids in Very Large VLDL | mmol/L | Lipoprotein subclasses | Very large VLDL (average diameter 64 nm) | Composite | 23489 | 23789 |
| XL_VLDL_PL | Phospholipids in Very Large VLDL | mmol/L | Lipoprotein subclasses | Very large VLDL (average diameter 64 nm) | Non-derived | 23490 | 23790 |
| XL_VLDL_C | Cholesterol in Very Large VLDL | mmol/L | Lipoprotein subclasses | Very large VLDL (average diameter 64 nm) | Composite | 23491 | 23791 |
| XL_VLDL_CE | Cholesteryl Esters in Very Large VLDL | mmol/L | Lipoprotein subclasses | Very large VLDL (average diameter 64 nm) | Non-derived | 23492 | 23792 |
| XL_VLDL_FC | Free Cholesterol in Very Large VLDL | mmol/L | Lipoprotein subclasses | Very large VLDL (average diameter 64 nm) | Non-derived | 23493 | 23793 |
| XL_VLDL_TG | Triglycerides in Very Large VLDL | mmol/L | Lipoprotein subclasses | Very large VLDL (average diameter 64 nm) | Non-derived | 23494 | 23794 |
| L_VLDL_P | Concentration of Large VLDL Particles | mmol/L | Lipoprotein subclasses | Large VLDL (average diameter 53.6 nm) | Non-derived | 23495 | 23795 |
| L_VLDL_L | Total Lipids in Large VLDL | mmol/L | Lipoprotein subclasses | Large VLDL (average | Composite | 23496 | 23796 |

| | | | | | | | |
|---|---|---|---|---|---|---|---|
| | | | | diameter 53.6 nm) | | | |
| L_VLDL_PL | Phospholipids in Large VLDL | mmol/L | Lipoprotein subclasses | Large VLDL (average diameter 53.6 nm) | Non-derived | 23497 | 23797 |
| L_VLDL_C | Cholesterol in Large VLDL | mmol/L | Lipoprotein subclasses | Large VLDL (average diameter 53.6 nm) | Composite | 23498 | 23798 |
| L_VLDL_CE | Cholesteryl Esters in Large VLDL | mmol/L | Lipoprotein subclasses | Large VLDL (average diameter 53.6 nm) | Non-derived | 23499 | 23799 |
| L_VLDL_FC | Free Cholesterol in Large VLDL | mmol/L | Lipoprotein subclasses | Large VLDL (average diameter 53.6 nm) | Non-derived | 23500 | 23800 |
| L_VLDL_TG | Triglycerides in Large VLDL | mmol/L | Lipoprotein subclasses | Large VLDL (average diameter 53.6 nm) | Non-derived | 23501 | 23801 |
| M_VLDL_P | Concentration of Medium VLDL Particles | mmol/L | Lipoprotein subclasses | Medium VLDL (average diameter 44.5 nm) | Non-derived | 23502 | 23802 |
| M_VLDL_L | Total Lipids in Medium VLDL | mmol/L | Lipoprotein subclasses | Medium VLDL (average diameter 44.5 nm) | Composite | 23503 | 23803 |
| M_VLDL_PL | Phospholipids in Medium VLDL | mmol/L | Lipoprotein subclasses | Medium VLDL (average diameter 44.5 nm) | Non-derived | 23504 | 23804 |
| M_VLDL_C | Cholesterol in Medium VLDL | mmol/L | Lipoprotein subclasses | Medium VLDL (average diameter 44.5 nm) | Composite | 23505 | 23805 |
| M_VLDL_CE | Cholesteryl Esters in Medium VLDL | mmol/L | Lipoprotein subclasses | Medium VLDL (average diameter 44.5 nm) | Non-derived | 23506 | 23806 |
| M_VLDL_FC | Free Cholesterol in Medium VLDL | mmol/L | Lipoprotein subclasses | Medium VLDL (average diameter 44.5 nm) | Non-derived | 23507 | 23807 |
| M_VLDL_TG | Triglycerides in Medium VLDL | mmol/L | Lipoprotein subclasses | Medium VLDL (average diameter 44.5 nm) | Non-derived | 23508 | 23808 |
| S_VLDL_P | Concentration of Small VLDL Particles | mmol/L | Lipoprotein subclasses | Small VLDL (average diameter 36.8 nm) | Non-derived | 23509 | 23809 |
| S_VLDL_L | Total Lipids in Small VLDL | mmol/L | Lipoprotein subclasses | Small VLDL (average diameter 36.8 nm) | Composite | 23510 | 23810 |
| S_VLDL_PL | Phospholipids in Small VLDL | mmol/L | Lipoprotein subclasses | Small VLDL (average diameter 36.8 nm) | Non-derived | 23511 | 23811 |

| | | | | | | | |
|---|---|---|---|---|---|---|---|
| S_VLDL_C | Cholesterol in Small VLDL | mmol/L | Lipoprotein subclasses | Small VLDL (average diameter 36.8 nm) | Composite | 23512 | 23812 |
| S_VLDL_CE | Cholesteryl Esters in Small VLDL | mmol/L | Lipoprotein subclasses | Small VLDL (average diameter 36.8 nm) | Non-derived | 23513 | 23813 |
| S_VLDL_FC | Free Cholesterol in Small VLDL | mmol/L | Lipoprotein subclasses | Small VLDL (average diameter 36.8 nm) | Non-derived | 23514 | 23814 |
| S_VLDL_TG | Triglycerides in Small VLDL | mmol/L | Lipoprotein subclasses | Small VLDL (average diameter 36.8 nm) | Non-derived | 23515 | 23815 |
| XS_VLDL_P | Concentration of Very Small VLDL Particles | mmol/L | Lipoprotein subclasses | Very small VLDL (average diameter 31.3 nm) | Non-derived | 23516 | 23816 |
| XS_VLDL_L | Total Lipids in Very Small VLDL | mmol/L | Lipoprotein subclasses | Very small VLDL (average diameter 31.3 nm) | Composite | 23517 | 23817 |
| XS_VLDL_PL | Phospholipids in Very Small VLDL | mmol/L | Lipoprotein subclasses | Very small VLDL (average diameter 31.3 nm) | Non-derived | 23518 | 23818 |
| XS_VLDL_C | Cholesterol in Very Small VLDL | mmol/L | Lipoprotein subclasses | Very small VLDL (average diameter 31.3 nm) | Composite | 23519 | 23819 |
| XS_VLDL_CE | Cholesteryl Esters in Very Small VLDL | mmol/L | Lipoprotein subclasses | Very small VLDL (average diameter 31.3 nm) | Non-derived | 23520 | 23820 |
| XS_VLDL_FC | Free Cholesterol in Very Small VLDL | mmol/L | Lipoprotein subclasses | Very small VLDL (average diameter 31.3 nm) | Non-derived | 23521 | 23821 |
| XS_VLDL_TG | Triglycerides in Very Small VLDL | mmol/L | Lipoprotein subclasses | Very small VLDL (average diameter 31.3 nm) | Non-derived | 23522 | 23822 |
| IDL_P | Concentration of IDL Particles | mmol/L | Lipoprotein subclasses | IDL (average diameter 28.6 nm) | Non-derived | 23523 | 23823 |
| IDL_L | Total Lipids in IDL | mmol/L | Lipoprotein subclasses | IDL (average diameter 28.6 nm) | Composite | 23524 | 23824 |
| IDL_PL | Phospholipids in IDL | mmol/L | Lipoprotein subclasses | IDL (average diameter 28.6 nm) | Non-derived | 23525 | 23825 |
| IDL_C | Cholesterol in IDL | mmol/L | Lipoprotein subclasses | IDL (average diameter 28.6 nm) | Composite | 23526 | 23826 |
| IDL_CE | Cholesteryl Esters in IDL | mmol/L | Lipoprotein subclasses | IDL (average diameter 28.6 nm) | Non-derived | 23527 | 23827 |
| IDL_FC | Free Cholesterol in IDL | mmol/L | Lipoprotein subclasses | IDL (average diameter 28.6 nm) | Non-derived | 23528 | 23828 |

| | | | | | | | |
|---|---|---|---|---|---|---|---|
| IDL_TG | Triglycerides in IDL | mmol/L | Lipoprotein subclasses | IDL (average diameter 28.6 nm) | Non-derived | 23529 | 23829 |
| L_LDL_P | Concentration of Large LDL Particles | mmol/L | Lipoprotein subclasses | Large LDL (average diameter 25.5 nm) | Non-derived | 23530 | 23830 |
| L_LDL_L | Total Lipids in Large LDL | mmol/L | Lipoprotein subclasses | Large LDL (average diameter 25.5 nm) | Composite | 23531 | 23831 |
| L_LDL_PL | Phospholipids in Large LDL | mmol/L | Lipoprotein subclasses | Large LDL (average diameter 25.5 nm) | Non-derived | 23532 | 23832 |
| L_LDL_C | Cholesterol in Large LDL | mmol/L | Lipoprotein subclasses | Large LDL (average diameter 25.5 nm) | Composite | 23533 | 23833 |
| L_LDL_CE | Cholesteryl Esters in Large LDL | mmol/L | Lipoprotein subclasses | Large LDL (average diameter 25.5 nm) | Non-derived | 23534 | 23834 |
| L_LDL_FC | Free Cholesterol in Large LDL | mmol/L | Lipoprotein subclasses | Large LDL (average diameter 25.5 nm) | Non-derived | 23535 | 23835 |
| L_LDL_TG | Triglycerides in Large LDL | mmol/L | Lipoprotein subclasses | Large LDL (average diameter 25.5 nm) | Non-derived | 23536 | 23836 |
| M_LDL_P | Concentration of Medium LDL Particles | mmol/L | Lipoprotein subclasses | Medium LDL (average diameter 23 nm) | Non-derived | 23537 | 23837 |
| M_LDL_L | Total Lipids in Medium LDL | mmol/L | Lipoprotein subclasses | Medium LDL (average diameter 23 nm) | Composite | 23538 | 23838 |
| M_LDL_PL | Phospholipids in Medium LDL | mmol/L | Lipoprotein subclasses | Medium LDL (average diameter 23 nm) | Non-derived | 23539 | 23839 |
| M_LDL_C | Cholesterol in Medium LDL | mmol/L | Lipoprotein subclasses | Medium LDL (average diameter 23 nm) | Composite | 23540 | 23840 |
| M_LDL_CE | Cholesteryl Esters in Medium LDL | mmol/L | Lipoprotein subclasses | Medium LDL (average diameter 23 nm) | Non-derived | 23541 | 23841 |
| M_LDL_FC | Free Cholesterol in Medium LDL | mmol/L | Lipoprotein subclasses | Medium LDL (average diameter 23 nm) | Non-derived | 23542 | 23842 |
| M_LDL_TG | Triglycerides in Medium LDL | mmol/L | Lipoprotein subclasses | Medium LDL (average diameter 23 nm) | Non-derived | 23543 | 23843 |
| S_LDL_P | Concentration of Small LDL Particles | mmol/L | Lipoprotein subclasses | Small LDL (average diameter 18.7 nm) | Non-derived | 23544 | 23844 |
| S_LDL_L | Total Lipids in Small LDL | mmol/L | Lipoprotein subclasses | Small LDL (average diameter 18.7 nm) | Composite | 23545 | 23845 |
| S_LDL_PL | Phospholipids in Small LDL | mmol/L | Lipoprotein subclasses | Small LDL (average | Non-derived | 23546 | 23846 |

| | | | | | | | |
|---|---|---|---|---|---|---|---|
| | | | | diameter 18.7 nm) | | | |
| S_LDL_C | Cholesterol in Small LDL | mmol/L | Lipoprotein subclasses | Small LDL (average diameter 18.7 nm) | Composite | 23547 | 23847 |
| S_LDL_CE | Cholesteryl Esters in Small LDL | mmol/L | Lipoprotein subclasses | Small LDL (average diameter 18.7 nm) | Non-derived | 23548 | 23848 |
| S_LDL_FC | Free Cholesterol in Small LDL | mmol/L | Lipoprotein subclasses | Small LDL (average diameter 18.7 nm) | Non-derived | 23549 | 23849 |
| S_LDL_TG | Triglycerides in Small LDL | mmol/L | Lipoprotein subclasses | Small LDL (average diameter 18.7 nm) | Non-derived | 23550 | 23850 |
| XL_HDL_P | Concentration of Very Large HDL Particles | mmol/L | Lipoprotein subclasses | Very large HDL (average diameter 14.3 nm) | Non-derived | 23551 | 23851 |
| XL_HDL_L | Total Lipids in Very Large HDL | mmol/L | Lipoprotein subclasses | Very large HDL (average diameter 14.3 nm) | Composite | 23552 | 23852 |
| XL_HDL_PL | Phospholipids in Very Large HDL | mmol/L | Lipoprotein subclasses | Very large HDL (average diameter 14.3 nm) | Non-derived | 23553 | 23853 |
| XL_HDL_C | Cholesterol in Very Large HDL | mmol/L | Lipoprotein subclasses | Very large HDL (average diameter 14.3 nm) | Composite | 23554 | 23854 |
| XL_HDL_CE | Cholesteryl Esters in Very Large HDL | mmol/L | Lipoprotein subclasses | Very large HDL (average diameter 14.3 nm) | Non-derived | 23555 | 23855 |
| XL_HDL_FC | Free Cholesterol in Very Large HDL | mmol/L | Lipoprotein subclasses | Very large HDL (average diameter 14.3 nm) | Non-derived | 23556 | 23856 |
| XL_HDL_TG | Triglycerides in Very Large HDL | mmol/L | Lipoprotein subclasses | Very large HDL (average diameter 14.3 nm) | Non-derived | 23557 | 23857 |
| L_HDL_P | Concentration of Large HDL Particles | mmol/L | Lipoprotein subclasses | Large HDL (average diameter 12.1 nm) | Non-derived | 23558 | 23858 |
| L_HDL_L | Total Lipids in Large HDL | mmol/L | Lipoprotein subclasses | Large HDL (average diameter 12.1 nm) | Composite | 23559 | 23859 |
| L_HDL_PL | Phospholipids in Large HDL | mmol/L | Lipoprotein subclasses | Large HDL (average diameter 12.1 nm) | Non-derived | 23560 | 23860 |
| L_HDL_C | Cholesterol in Large HDL | mmol/L | Lipoprotein subclasses | Large HDL (average diameter 12.1 nm) | Composite | 23561 | 23861 |
| L_HDL_CE | Cholesteryl Esters in Large HDL | mmol/L | Lipoprotein subclasses | Large HDL (average diameter 12.1 nm) | Non-derived | 23562 | 23862 |
| L_HDL_FC | Free Cholesterol in Large HDL | mmol/L | Lipoprotein subclasses | Large HDL (average diameter 12.1 nm) | Non-derived | 23563 | 23863 |

| | | | | | | | |
|---|---|---|---|---|---|---|---|
| L_HDL_TG | Triglycerides in Large HDL | mmol/L | Lipoprotein subclasses | Large HDL (average diameter 12.1 nm) | Non-derived | 23564 | 23864 |
| M_HDL_P | Concentration of Medium HDL Particles | mmol/L | Lipoprotein subclasses | Medium HDL (average diameter 10.9 nm) | Non-derived | 23565 | 23865 |
| M_HDL_L | Total Lipids in Medium HDL | mmol/L | Lipoprotein subclasses | Medium HDL (average diameter 10.9 nm) | Composite | 23566 | 23866 |
| M_HDL_PL | Phospholipids in Medium HDL | mmol/L | Lipoprotein subclasses | Medium HDL (average diameter 10.9 nm) | Non-derived | 23567 | 23867 |
| M_HDL_C | Cholesterol in Medium HDL | mmol/L | Lipoprotein subclasses | Medium HDL (average diameter 10.9 nm) | Composite | 23568 | 23868 |
| M_HDL_CE | Cholesteryl Esters in Medium HDL | mmol/L | Lipoprotein subclasses | Medium HDL (average diameter 10.9 nm) | Non-derived | 23569 | 23869 |
| M_HDL_FC | Free Cholesterol in Medium HDL | mmol/L | Lipoprotein subclasses | Medium HDL (average diameter 10.9 nm) | Non-derived | 23570 | 23870 |
| M_HDL_TG | Triglycerides in Medium HDL | mmol/L | Lipoprotein subclasses | Medium HDL (average diameter 10.9 nm) | Non-derived | 23571 | 23871 |
| S_HDL_P | Concentration of Small HDL Particles | mmol/L | Lipoprotein subclasses | Small HDL (average diameter 8.7 nm) | Non-derived | 23572 | 23872 |
| S_HDL_L | Total Lipids in Small HDL | mmol/L | Lipoprotein subclasses | Small HDL (average diameter 8.7 nm) | Composite | 23573 | 23873 |
| S_HDL_PL | Phospholipids in Small HDL | mmol/L | Lipoprotein subclasses | Small HDL (average diameter 8.7 nm) | Non-derived | 23574 | 23874 |
| S_HDL_C | Cholesterol in Small HDL | mmol/L | Lipoprotein subclasses | Small HDL (average diameter 8.7 nm) | Composite | 23575 | 23875 |
| S_HDL_CE | Cholesteryl Esters in Small HDL | mmol/L | Lipoprotein subclasses | Small HDL (average diameter 8.7 nm) | Non-derived | 23576 | 23876 |
| S_HDL_FC | Free Cholesterol in Small HDL | mmol/L | Lipoprotein subclasses | Small HDL (average diameter 8.7 nm) | Non-derived | 23577 | 23877 |
| S_HDL_TG | Triglycerides in Small HDL | mmol/L | Lipoprotein subclasses | Small HDL (average diameter 8.7 nm) | Non-derived | 23578 | 23878 |
| XXL_VLDL_PL_pct | Phospholipids to Total Lipids in Chylomicrons and Extremely Large VLDL percentage | % | Relative lipoprotein lipid concentrations | Chylomicrons and extremely large VLDL ratios | Percentage | 23579 | 23879 |
| XXL_VLDL_C_pct | Cholesterol to Total Lipids in Chylomicrons and Extremely Large VLDL percentage | % | Relative lipoprotein lipid concentrations | Chylomicrons and extremely large VLDL ratios | Percentage | 23580 | 23880 |

| | | | | | | | |
|---|---|---|---|---|---|---|---|
| XXL_VLDL_CE_pct | Cholesteryl Esters to Total Lipids in Chylomicrons and Extremely Large VLDL percentage | % | Relative lipoprotein lipid concentrations | Chylomicrons and extremely large VLDL ratios | Percentage | 23581 | 23881 |
| XXL_VLDL_FC_pct | Free Cholesterol to Total Lipids in Chylomicrons and Extremely Large VLDL percentage | % | Relative lipoprotein lipid concentrations | Chylomicrons and extremely large VLDL ratios | Percentage | 23582 | 23882 |
| XXL_VLDL_TG_pct | Triglycerides to Total Lipids in Chylomicrons and Extremely Large VLDL percentage | % | Relative lipoprotein lipid concentrations | Chylomicrons and extremely large VLDL ratios | Percentage | 23583 | 23883 |
| XL_VLDL_PL_pct | Phospholipids to Total Lipids in Very Large VLDL percentage | % | Relative lipoprotein lipid concentrations | Very large VLDL ratios | Percentage | 23584 | 23884 |
| XL_VLDL_C_pct | Cholesterol to Total Lipids in Very Large VLDL percentage | % | Relative lipoprotein lipid concentrations | Very large VLDL ratios | Percentage | 23585 | 23885 |
| XL_VLDL_CE_pct | Cholesteryl Esters to Total Lipids in Very Large VLDL percentage | % | Relative lipoprotein lipid concentrations | Very large VLDL ratios | Percentage | 23586 | 23886 |
| XL_VLDL_FC_pct | Free Cholesterol to Total Lipids in Very Large VLDL percentage | % | Relative lipoprotein lipid concentrations | Very large VLDL ratios | Percentage | 23587 | 23887 |
| XL_VLDL_TG_pct | Triglycerides to Total Lipids in Very Large VLDL percentage | % | Relative lipoprotein lipid concentrations | Very large VLDL ratios | Percentage | 23588 | 23888 |
| L_VLDL_PL_pct | Phospholipids to Total Lipids in Large VLDL percentage | % | Relative lipoprotein lipid concentrations | Large VLDL ratios | Percentage | 23589 | 23889 |
| L_VLDL_C_pct | Cholesterol to Total Lipids in Large VLDL percentage | % | Relative lipoprotein lipid concentrations | Large VLDL ratios | Percentage | 23590 | 23890 |
| L_VLDL_CE_pct | Cholesteryl Esters to Total Lipids in Large VLDL percentage | % | Relative lipoprotein lipid concentrations | Large VLDL ratios | Percentage | 23591 | 23891 |
| L_VLDL_FC_pct | Free Cholesterol to Total Lipids in Large VLDL percentage | % | Relative lipoprotein lipid concentrations | Large VLDL ratios | Percentage | 23592 | 23892 |
| L_VLDL_TG_pct | Triglycerides to Total Lipids in Large VLDL percentage | % | Relative lipoprotein lipid concentrations | Large VLDL ratios | Percentage | 23593 | 23893 |
| M_VLDL_PL_pct | Phospholipids to Total Lipids in Medium VLDL percentage | % | Relative lipoprotein lipid concentrations | Medium VLDL ratios | Percentage | 23594 | 23894 |
| M_VLDL_C_pct | Cholesterol to Total Lipids in Medium VLDL percentage | % | Relative lipoprotein lipid concentrations | Medium VLDL ratios | Percentage | 23595 | 23895 |
| M_VLDL_CE_pct | Cholesteryl Esters to Total Lipids in Medium VLDL percentage | % | Relative lipoprotein lipid concentrations | Medium VLDL ratios | Percentage | 23596 | 23896 |
| M_VLDL_FC_pct | Free Cholesterol to Total Lipids in Medium VLDL percentage | % | Relative lipoprotein lipid concentrations | Medium VLDL ratios | Percentage | 23597 | 23897 |
| M_VLDL_TG_pct | Triglycerides to Total Lipids in Medium VLDL percentage | % | Relative lipoprotein lipid concentrations | Medium VLDL ratios | Percentage | 23598 | 23898 |

| | | | | | | | |
|---|---|---|---|---|---|---|---|
| S_VLDL_PL_pct | Phospholipids to Total Lipids in Small VLDL percentage | % | Relative lipoprotein lipid concentrations | Small VLDL ratios | Percentage | 23599 | 23899 |
| S_VLDL_C_pct | Cholesterol to Total Lipids in Small VLDL percentage | % | Relative lipoprotein lipid concentrations | Small VLDL ratios | Percentage | 23600 | 23900 |
| S_VLDL_CE_pct | Cholesteryl Esters to Total Lipids in Small VLDL percentage | % | Relative lipoprotein lipid concentrations | Small VLDL ratios | Percentage | 23601 | 23901 |
| S_VLDL_FC_pct | Free Cholesterol to Total Lipids in Small VLDL percentage | % | Relative lipoprotein lipid concentrations | Small VLDL ratios | Percentage | 23602 | 23902 |
| S_VLDL_TG_pct | Triglycerides to Total Lipids in Small VLDL percentage | % | Relative lipoprotein lipid concentrations | Small VLDL ratios | Percentage | 23603 | 23903 |
| XS_VLDL_PL_pct | Phospholipids to Total Lipids in Very Small VLDL percentage | % | Relative lipoprotein lipid concentrations | Very small VLDL ratios | Percentage | 23604 | 23904 |
| XS_VLDL_C_pct | Cholesterol to Total Lipids in Very Small VLDL percentage | % | Relative lipoprotein lipid concentrations | Very small VLDL ratios | Percentage | 23605 | 23905 |
| XS_VLDL_CE_pct | Cholesteryl Esters to Total Lipids in Very Small VLDL percentage | % | Relative lipoprotein lipid concentrations | Very small VLDL ratios | Percentage | 23606 | 23906 |
| XS_VLDL_FC_pct | Free Cholesterol to Total Lipids in Very Small VLDL percentage | % | Relative lipoprotein lipid concentrations | Very small VLDL ratios | Percentage | 23607 | 23907 |
| XS_VLDL_TG_pct | Triglycerides to Total Lipids in Very Small VLDL percentage | % | Relative lipoprotein lipid concentrations | Very small VLDL ratios | Percentage | 23608 | 23908 |
| IDL_PL_pct | Phospholipids to Total Lipids in IDL percentage | % | Relative lipoprotein lipid concentrations | IDL ratios | Percentage | 23609 | 23909 |
| IDL_C_pct | Cholesterol to Total Lipids in IDL percentage | % | Relative lipoprotein lipid concentrations | IDL ratios | Percentage | 23610 | 23910 |
| IDL_CE_pct | Cholesteryl Esters to Total Lipids in IDL percentage | % | Relative lipoprotein lipid concentrations | IDL ratios | Percentage | 23611 | 23911 |
| IDL_FC_pct | Free Cholesterol to Total Lipids in IDL percentage | % | Relative lipoprotein lipid concentrations | IDL ratios | Percentage | 23612 | 23912 |
| IDL_TG_pct | Triglycerides to Total Lipids in IDL percentage | % | Relative lipoprotein lipid concentrations | IDL ratios | Percentage | 23613 | 23913 |
| L_LDL_PL_pct | Phospholipids to Total Lipids in Large LDL percentage | % | Relative lipoprotein lipid concentrations | Large LDL ratios | Percentage | 23614 | 23914 |
| L_LDL_C_pct | Cholesterol to Total Lipids in Large LDL percentage | % | Relative lipoprotein lipid concentrations | Large LDL ratios | Percentage | 23615 | 23915 |
| L_LDL_CE_pct | Cholesteryl Esters to Total Lipids in Large LDL percentage | % | Relative lipoprotein lipid concentrations | Large LDL ratios | Percentage | 23616 | 23916 |
| L_LDL_FC_pct | Free Cholesterol to Total Lipids in Large LDL percentage | % | Relative lipoprotein lipid concentrations | Large LDL ratios | Percentage | 23617 | 23917 |
| L_LDL_TG_pct | Triglycerides to Total Lipids in Large LDL percentage | % | Relative lipoprotein lipid concentrations | Large LDL ratios | Percentage | 23618 | 23918 |
| M_LDL_PL_pct | Phospholipids to Total Lipids in Medium LDL percentage | % | Relative lipoprotein lipid concentrations | Medium LDL ratios | Percentage | 23619 | 23919 |

| | | | | | | | |
|---|---|---|---|---|---|---|---|
| M_LDL_C_pct | Cholesterol to Total Lipids in Medium LDL percentage | % | Relative lipoprotein lipid concentrations | Medium LDL ratios | Percentage | 23620 | 23920 |
| M_LDL_CE_pct | Cholesteryl Esters to Total Lipids in Medium LDL percentage | % | Relative lipoprotein lipid concentrations | Medium LDL ratios | Percentage | 23621 | 23921 |
| M_LDL_FC_pct | Free Cholesterol to Total Lipids in Medium LDL percentage | % | Relative lipoprotein lipid concentrations | Medium LDL ratios | Percentage | 23622 | 23922 |
| M_LDL_TG_pct | Triglycerides to Total Lipids in Medium LDL percentage | % | Relative lipoprotein lipid concentrations | Medium LDL ratios | Percentage | 23623 | 23923 |
| S_LDL_PL_pct | Phospholipids to Total Lipids in Small LDL percentage | % | Relative lipoprotein lipid concentrations | Small LDL ratios | Percentage | 23624 | 23924 |
| S_LDL_C_pct | Cholesterol to Total Lipids in Small LDL percentage | % | Relative lipoprotein lipid concentrations | Small LDL ratios | Percentage | 23625 | 23925 |
| S_LDL_CE_pct | Cholesteryl Esters to Total Lipids in Small LDL percentage | % | Relative lipoprotein lipid concentrations | Small LDL ratios | Percentage | 23626 | 23926 |
| S_LDL_FC_pct | Free Cholesterol to Total Lipids in Small LDL percentage | % | Relative lipoprotein lipid concentrations | Small LDL ratios | Percentage | 23627 | 23927 |
| S_LDL_TG_pct | Triglycerides to Total Lipids in Small LDL percentage | % | Relative lipoprotein lipid concentrations | Small LDL ratios | Percentage | 23628 | 23928 |
| XL_HDL_PL_pct | Phospholipids to Total Lipids in Very Large HDL percentage | % | Relative lipoprotein lipid concentrations | Very large HDL ratios | Percentage | 23629 | 23929 |
| XL_HDL_C_pct | Cholesterol to Total Lipids in Very Large HDL percentage | % | Relative lipoprotein lipid concentrations | Very large HDL ratios | Percentage | 23630 | 23930 |
| XL_HDL_CE_pct | Cholesteryl Esters to Total Lipids in Very Large HDL percentage | % | Relative lipoprotein lipid concentrations | Very large HDL ratios | Percentage | 23631 | 23931 |
| XL_HDL_FC_pct | Free Cholesterol to Total Lipids in Very Large HDL percentage | % | Relative lipoprotein lipid concentrations | Very large HDL ratios | Percentage | 23632 | 23932 |
| XL_HDL_TG_pct | Triglycerides to Total Lipids in Very Large HDL percentage | % | Relative lipoprotein lipid concentrations | Very large HDL ratios | Percentage | 23633 | 23933 |
| L_HDL_PL_pct | Phospholipids to Total Lipids in Large HDL percentage | % | Relative lipoprotein lipid concentrations | Large HDL ratios | Percentage | 23634 | 23934 |
| L_HDL_C_pct | Cholesterol to Total Lipids in Large HDL percentage | % | Relative lipoprotein lipid concentrations | Large HDL ratios | Percentage | 23635 | 23935 |
| L_HDL_CE_pct | Cholesteryl Esters to Total Lipids in Large HDL percentage | % | Relative lipoprotein lipid concentrations | Large HDL ratios | Percentage | 23636 | 23936 |
| L_HDL_FC_pct | Free Cholesterol to Total Lipids in Large HDL percentage | % | Relative lipoprotein lipid concentrations | Large HDL ratios | Percentage | 23637 | 23937 |
| L_HDL_TG_pct | Triglycerides to Total Lipids in Large HDL percentage | % | Relative lipoprotein lipid concentrations | Large HDL ratios | Percentage | 23638 | 23938 |
| M_HDL_PL_pct | Phospholipids to Total Lipids in Medium HDL percentage | % | Relative lipoprotein lipid concentrations | Medium HDL ratios | Percentage | 23639 | 23939 |

| | | | | | | | |
|---|---|---|---|---|---|---|---|
| M_HDL_C_pct | Cholesterol to Total Lipids in Medium HDL percentage | % | Relative lipoprotein lipid concentrations | Medium HDL ratios | Percentage | 23640 | 23940 |
| M_HDL_CE_pct | Cholesteryl Esters to Total Lipids in Medium HDL percentage | % | Relative lipoprotein lipid concentrations | Medium HDL ratios | Percentage | 23641 | 23941 |
| M_HDL_FC_pct | Free Cholesterol to Total Lipids in Medium HDL percentage | % | Relative lipoprotein lipid concentrations | Medium HDL ratios | Percentage | 23642 | 23942 |
| M_HDL_TG_pct | Triglycerides to Total Lipids in Medium HDL percentage | % | Relative lipoprotein lipid concentrations | Medium HDL ratios | Percentage | 23643 | 23943 |
| S_HDL_PL_pct | Phospholipids to Total Lipids in Small HDL percentage | % | Relative lipoprotein lipid concentrations | Small HDL ratios | Percentage | 23644 | 23944 |
| S_HDL_C_pct | Cholesterol to Total Lipids in Small HDL percentage | % | Relative lipoprotein lipid concentrations | Small HDL ratios | Percentage | 23645 | 23945 |
| S_HDL_CE_pct | Cholesteryl Esters to Total Lipids in Small HDL percentage | % | Relative lipoprotein lipid concentrations | Small HDL ratios | Percentage | 23646 | 23946 |
| S_HDL_FC_pct | Free Cholesterol to Total Lipids in Small HDL percentage | % | Relative lipoprotein lipid concentrations | Small HDL ratios | Percentage | 23647 | 23947 |
| S_HDL_TG_pct | Triglycerides to Total Lipids in Small HDL percentage | % | Relative lipoprotein lipid concentrations | Small HDL ratios | Percentage | 23648 | 23948 |
| HDL_C_pct | Cholesterol to Total Lipids in HDL percentage | % | Relative lipid concentrations | NA | Percentage | | |
| HDL_CE_pct | Cholesteryl Esters to Total Lipids in HDL percentage | % | Relative lipid concentrations | NA | Percentage | | |
| HDL_CE_pct_C | Cholesteryl Esters to Cholesterol in HDL percentage | % | Relative cholesterol concentrations | NA | Percentage | | |
| HDL_FC_by_CE | Free Cholesterol to Cholesteryl Esters in HDL percentage | ratio | Relative lipid concentrations | NA | Ratio | | |
| HDL_FC_pct | Free Cholesterol to Total Lipids in HDL percentage | % | Relative lipid concentrations | NA | Percentage | | |
| HDL_FC_pct_C | Free Cholesterol to Cholesterol in HDL percentage | % | Relative cholesterol concentrations | NA | Percentage | | |
| HDL_PL_pct | Phospholipids to Total Lipids in HDL percentage | % | Relative lipid concentrations | NA | Percentage | | |
| HDL_TG_pct | Triglycerides to Total Lipids in HDL percentage | % | Relative lipid concentrations | NA | Percentage | | |
| IDL_CE_pct_C | Cholesteryl Esters to Cholesterol in IDL percentage | % | Relative lipoprotein cholesterol concentrations | IDL ratios | Percentage | | |
| IDL_FC_by_CE | Free Cholesterol to Cholesteryl Esters in IDL percentage | ratio | Relative lipoprotein lipid concentrations | IDL ratios | Ratio | | |
| IDL_FC_pct_C | Free Cholesterol to Cholesterol in IDL percentage | % | Relative lipoprotein cholesterol concentrations | IDL ratios | Percentage | | |
| L_HDL_CE_pct_C | Cholesteryl Esters to Cholesterol in Large HDL percentage | % | Relative lipoprotein cholesterol concentrations | Large HDL ratios | Percentage | | |

| | | | | | |
|---|---|---|---|---|---|
| L_HDL_FC_by_CE | Free Cholesterol to Cholesteryl Esters in Large HDL ratio | ratio | Relative lipoprotein lipid concentrations | Large HDL ratios | Ratio |
| L_HDL_FC_pct_C | Free Cholesterol to Cholesterol in Large HDL percentage | % | Relative lipoprotein cholesterol concentrations | Large HDL ratios | Percentage |
| L_LDL_CE_pct_C | Cholesteryl Esters to Cholesterol in Large LDL percentage | % | Relative lipoprotein cholesterol concentrations | Large LDL ratios | Percentage |
| L_LDL_FC_by_CE | Free Cholesterol to Cholesteryl Esters in Large LDL ratio | ratio | Relative lipoprotein lipid concentrations | Large LDL ratios | Ratio |
| L_LDL_FC_pct_C | Free Cholesterol to Cholesterol in Large LDL percentage | % | Relative lipoprotein cholesterol concentrations | Large LDL ratios | Percentage |
| L_VLDL_CE_pct_C | Cholesteryl Esters to Cholesterol in Large VLDL percentage | % | Relative lipoprotein cholesterol concentrations | Large VLDL ratios | Percentage |
| L_VLDL_FC_by_CE | Free Cholesterol to Cholesteryl Esters in Large VLDL ratio | ratio | Relative lipoprotein lipid concentrations | Large VLDL ratios | Ratio |
| L_VLDL_FC_pct_C | Free Cholesterol to Cholesterol in Large VLDL percentage | % | Relative lipoprotein cholesterol concentrations | Large VLDL ratios | Percentage |
| LDL_C_pct | Cholesterol to Total Lipids in LDL percentage | % | Relative lipid concentrations | NA | Percentage |
| LDL_CE_pct | Cholesteryl Esters to Total Lipids in LDL percentage | % | Relative lipid concentrations | NA | Percentage |
| LDL_CE_pct_C | Cholesteryl Esters to Cholesterol in LDL percentage | % | Relative cholesterol concentrations | NA | Percentage |
| LDL_FC_by_CE | Free Cholesterol to Cholesteryl Esters in LDL percentage | ratio | Relative lipid concentrations | NA | Ratio |
| LDL_FC_pct | Free Cholesterol to Total Lipids in LDL percentage | % | Relative lipid concentrations | NA | Percentage |
| LDL_FC_pct_C | Free Cholesterol to Cholesterol in LDL percentage | % | Relative cholesterol concentrations | NA | Percentage |
| LDL_PL_pct | Phospholipids to Total Lipids in LDL percentage | % | Relative lipid concentrations | NA | Percentage |
| LDL_TG_pct | Triglycerides to Total Lipids in LDL percentage | % | Relative lipid concentrations | NA | Percentage |
| M_HDL_CE_pct_C | Cholesteryl Esters to Cholesterol in Medium HDL percentage | % | Relative lipoprotein cholesterol concentrations | Medium HDL ratios | Percentage |
| M_HDL_FC_by_CE | Free Cholesterol to Cholesteryl Esters in Medium HDL ratio | ratio | Relative lipoprotein lipid concentrations | Medium HDL ratios | Ratio |
| M_HDL_FC_pct_C | Free Cholesterol to Cholesterol in Medium HDL percentage | % | Relative lipoprotein cholesterol concentrations | Medium HDL ratios | Percentage |
| M_LDL_CE_pct_C | Cholesteryl Esters to Cholesterol in Medium LDL percentage | % | Relative lipoprotein cholesterol concentrations | Medium LDL ratios | Percentage |

| | | | | | |
|---|---|---|---|---|---|
| M_LDL_FC_by_CE | Free Cholesterol to Cholesteryl Esters in Medium LDL ratio | ratio | Relative lipoprotein lipid concentrations | Medium LDL ratios | Ratio |
| M_LDL_FC_pct_C | Free Cholesterol to Cholesterol in Medium LDL percentage | % | Relative lipoprotein cholesterol concentrations | Medium LDL ratios | Percentage |
| M_VLDL_CE_pct_C | Cholesteryl Esters to Cholesterol in Medium VLDL percentage | % | Relative lipoprotein cholesterol concentrations | Medium VLDL ratios | Percentage |
| M_VLDL_FC_by_CE | Free Cholesterol to Cholesteryl Esters in Medium VLDL ratio | ratio | Relative lipoprotein lipid concentrations | Medium VLDL ratios | Ratio |
| M_VLDL_FC_pct_C | Free Cholesterol to Cholesterol in Medium VLDL percentage | % | Relative lipoprotein cholesterol concentrations | Medium VLDL ratios | Percentage |
| Omega_3_pct_PUFA | Omega-3 Fatty Acids to Polyunsaturated Fatty Acids percentage | % | Fatty acids | Fatty acid ratios | Percentage |
| Omega_6_pct_PUFA | Omega-6 Fatty Acids to Polyunsaturated Fatty Acids percentage | % | Fatty acids | Fatty acid ratios | Percentage |
| S_HDL_CE_pct_C | Cholesteryl Esters to Cholesterol in Small HDL percentage | % | Relative lipoprotein cholesterol concentrations | Small HDL ratios | Percentage |
| S_HDL_FC_by_CE | Free Cholesterol to Cholesteryl Esters in Small HDL ratio | ratio | Relative lipoprotein lipid concentrations | Small HDL ratios | Ratio |
| S_HDL_FC_pct_C | Free Cholesterol to Cholesterol in Small HDL percentage | % | Relative lipoprotein cholesterol concentrations | Small HDL ratios | Percentage |
| S_LDL_CE_pct_C | Cholesteryl Esters to Cholesterol in Small LDL percentage | % | Relative lipoprotein cholesterol concentrations | Small LDL ratios | Percentage |
| S_LDL_FC_by_CE | Free Cholesterol to Cholesteryl Esters in Small LDL ratio | ratio | Relative lipoprotein lipid concentrations | Small LDL ratios | Ratio |
| S_LDL_FC_pct_C | Free Cholesterol to Cholesterol in Small LDL percentage | % | Relative lipoprotein cholesterol concentrations | Small LDL ratios | Percentage |
| S_VLDL_CE_pct_C | Cholesteryl Esters to Cholesterol in Small VLDL percentage | % | Relative lipoprotein cholesterol concentrations | Small VLDL ratios | Percentage |
| S_VLDL_FC_by_CE | Free Cholesterol to Cholesteryl Esters in Small VLDL ratio | ratio | Relative lipoprotein lipid concentrations | Small VLDL ratios | Ratio |
| S_VLDL_FC_pct_C | Free Cholesterol to Cholesterol in Small VLDL percentage | % | Relative lipoprotein cholesterol concentrations | Small VLDL ratios | Percentage |
| Total_C_pct | Total Cholesterol to Total Lipids percentage | % | Relative lipid concentrations | NA | Percentage |
| Total_CE_pct | Total Esterified Cholesterol to Total Lipids percentage | % | Relative lipid concentrations | NA | Percentage |
| Total_CE_pct_C | Total Esterified Cholesterol to Total | % | Relative cholesterol concentrations | NA | Percentage |

| | | | | | |
|---|---|---|---|---|---|
| | Cholesterol percentage | | | | |
| Total_FC_by_CE | Total Free Cholesterol to Total Esterified Cholesterol ratio | ratio | Relative lipid concentrations | NA | Ratio |
| Total_FC_pct | Total Free Cholesterol to Total Lipids percentage | % | Relative lipid concentrations | NA | Percentage |
| Total_FC_pct_C | Total Free Cholesterol to Total Cholesterol percentage | % | Relative cholesterol concentrations | NA | Percentage |
| Total_PL_pct | Total Phospholipids to Total Lipids percentage | % | Relative lipid concentrations | NA | Percentage |
| Total_TG_pct | Total Triglycerides to Total Lipids percentage | % | Relative lipid concentrations | NA | Percentage |
| VLDL_C_pct | Cholesterol to Total Lipids in VLDL percentage | % | Relative lipid concentrations | NA | Percentage |
| VLDL_CE_pct | Cholesteryl Esters to Total Lipids in VLDL percentage | % | Relative lipid concentrations | NA | Percentage |
| VLDL_CE_pct_C | Cholesteryl Esters to Cholesterol in VLDL percentage | % | Relative cholesterol concentrations | NA | Percentage |
| VLDL_FC_by_CE | Free Cholesterol to Cholesteryl Esters in VLDL percentage | ratio | Relative lipid concentrations | NA | Ratio |
| VLDL_FC_pct | Free Cholesterol to Total Lipids in VLDL percentage | % | Relative lipid concentrations | NA | Percentage |
| VLDL_FC_pct_C | Free Cholesterol to Cholesterol in VLDL percentage | % | Relative cholesterol concentrations | NA | Percentage |
| VLDL_PL_pct | Phospholipids to Total Lipids in VLDL percentage | % | Relative lipid concentrations | NA | Percentage |
| VLDL_TG_pct | Triglycerides to Total Lipids in VLDL percentage | % | Relative lipid concentrations | NA | Percentage |
| XL_HDL_CE_pct_C | Cholesteryl Esters to Cholesterol in Very Large HDL percentage | % | Relative lipoprotein cholesterol concentrations | Very large HDL ratios | Percentage |
| XL_HDL_FC_by_CE | Free Cholesterol to Cholesteryl Esters in Very Large HDL ratio | ratio | Relative lipoprotein lipid concentrations | Very large HDL ratios | Ratio |
| XL_HDL_FC_pct_C | Free Cholesterol to Cholesterol in Very Large HDL percentage | % | Relative lipoprotein cholesterol concentrations | Very large HDL ratios | Percentage |
| XL_VLDL_CE_pct_C | Cholesteryl Esters to Cholesterol in Very Large VLDL percentage | % | Relative lipoprotein cholesterol concentrations | Very large VLDL ratios | Percentage |
| XL_VLDL_FC_by_CE | Free Cholesterol to Cholesteryl Esters in Very Large VLDL ratio | ratio | Relative lipoprotein lipid concentrations | Very large VLDL ratios | Ratio |
| XL_VLDL_FC_pct_C | Free Cholesterol to Cholesterol in Very Large VLDL percentage | % | Relative lipoprotein cholesterol concentrations | Very large VLDL ratios | Percentage |
| XS_VLDL_CE_pct_C | Cholesteryl Esters to Cholesterol in Very | % | Relative lipoprotein | Very small VLDL ratios | Percentage |

| | | | | | |
|---|---|---|---|---|---|
| | Small VLDL percentage | | cholesterol concentrations | | |
| XS_VLDL_FC_by_CE | Free Cholesterol to Cholesteryl Esters in Very Small VLDL ratio | ratio | Relative lipoprotein lipid concentrations | Very small VLDL ratios | Ratio |
| XS_VLDL_FC_pct_C | Free Cholesterol to Cholesterol in Very Small VLDL percentage | % | Relative lipoprotein cholesterol concentrations | Very small VLDL ratios | Percentage |
| XXL_VLDL_CE_pct_C | Cholesteryl Esters to Cholesterol in Chylomicrons and Extremely Large VLDL percentage | % | Relative lipoprotein cholesterol concentrations | Chylomicrons and extremely large VLDL ratios | Percentage |
| XXL_VLDL_FC_by_CE | Free Cholesterol to Cholesteryl Esters in Chylomicrons and Extremely Large VLDL ratio | ratio | Relative lipoprotein lipid concentrations | Chylomicrons and extremely large VLDL ratios | Ratio |
| XXL_VLDL_FC_pct_C | Free Cholesterol to Cholesterol in Chylomicrons and Extremely Large VLDL percentage | % | Relative lipoprotein cholesterol concentrations | Chylomicrons and extremely large VLDL ratios | Percentage |

**eTable 4: Genomic loci of the 7 MRIBAG GWASs**

| TopLeadSNP | Chromosome | Position | P-value | MRIBAG | Cytogenetic region |
|---|---|---|---|---|---|
| rs61732315 | 1 | 203028371 | 1.63E-08 | brain | 1q32.1 |
| rs1452628 | 1 | 215139887 | 3.04E-14 | brain | 1q41 |
| rs186399184 | 2 | 203728984 | 1.05E-08 | brain | 2q33.2 |
| rs10933668 | 3 | 194470632 | 9.41E-09 | brain | 3q29 |
| rs34051980 | 8 | 119948046 | 2.02E-08 | brain | 8q24.12 |
| rs534115641 | 17 | 43667537 | 8.45E-23 | brain | 17q21.31 |
| rs4852828 | 2 | 72192369 | 1.35E-08 | adipose | 2p13.2 |
| rs4805881 | 19 | 33896432 | 7.64E-21 | adipose | 19q13.11 |
| rs2009371 | 1 | 16348083 | 2.83E-08 | heart | 1p36.13 |
| rs2562845 | 2 | 179514433 | 6.72E-09 | heart | 2q31.2 |
| rs7714279 | 5 | 95527383 | 2.29E-12 | heart | 5q15 |
| rs11768878 | 7 | 73431169 | 5.87E-16 | heart | 7q11.23 |
| rs914279 | 10 | 30170487 | 1.32E-08 | heart | 10p11.23 |
| rs17875585 | 15 | 81601745 | 8.12E-09 | heart | 15q25.1 |
| rs4649024 | 1 | 23701583 | 1.74E-08 | kidney | 1p36.12 |
| rs2345962 | 1 | 163741788 | 4.09E-10 | kidney | 1q23.3 |
| rs954244 | 2 | 121309231 | 1.98E-08 | kidney | 2q14.2 |
| rs17036160 | 3 | 12329783 | 4.52E-08 | kidney | 3p25.2 |
| rs1007984 | 5 | 52636896 | 2.53E-11 | kidney | 5q11.2 |
| 5:55860907_GC_G | 5 | 55860907 | 4.42E-21 | kidney | 5q11.2 |
| rs11992444 | 8 | 25464690 | 1.10E-13 | kidney | 8p21.2 |
| rs1485748 | 8 | 25870252 | 3.21E-08 | kidney | 8p21.2 |
| rs11614913 | 12 | 54385599 | 4.91E-08 | kidney | 12q13.13 |
| rs77924615 | 16 | 20392332 | 3.32E-14 | kidney | 16p12.3 |
| rs1260326 | 2 | 27730940 | 4.67E-12 | liver | 2p23.3 |
| rs6858148 | 4 | 100065917 | 7.59E-11 | liver | 4q23 |
| rs13107325 | 4 | 103188709 | 1.16E-19 | liver | 4q24 |
| rs199922514 | 8 | 9183102 | 4.80E-37 | liver | 8p23.1 |
| rs7924036 | 10 | 65191645 | 2.25E-18 | liver | 10q21.3 |
| rs1539042 | 10 | 93624017 | 7.30E-09 | liver | 10q23.32 |
| rs141866277 | 15 | 43950699 | 4.72E-11 | liver | 15q15.3 |
| 19:19432290_AG_A | 19 | 19432290 | 1.32E-13 | liver | 19p13.11 |
| rs2294915 | 22 | 44340904 | 7.81E-32 | liver | 22q13.31 |
| rs2453728 | 1 | 51460955 | 2.98E-13 | pancreas | 1p32.3 |
| rs11589479 | 1 | 155033308 | 1.63E-08 | pancreas | 1q22 |
| rs2800734 | 6 | 127417035 | 2.68E-12 | pancreas | 6q22.33 |
| rs1016431 | 7 | 51020336 | 6.58E-10 | pancreas | 7p12.1 |
| rs1561928 | 8 | 129568061 | 1.44E-14 | pancreas | 8q24.21 |
| 9:136138765_GCGCCCACCACTA_G | 9 | 136138765 | 8.91E-15 | pancreas | 9q34.2 |
| rs769008 | 10 | 49390308 | 3.48E-12 | pancreas | 10q11.22 |
| rs7299428 | 12 | 42309242 | 7.71E-14 | pancreas | 12q12 |
| rs3742397 | 14 | 101319686 | 4.39E-08 | pancreas | 14q32.2 |
| 16:75247337_GT_G | 16 | 75247337 | 7.65E-11 | pancreas | 16q23.1 |
| rs7405380 | 16 | 88975910 | 5.86E-12 | pancreas | 16q24.3 |
| rs62090594 | 18 | 42383005 | 3.28E-08 | pancreas | 18q12.3 |
| rs922048 | 18 | 56876386 | 6.74E-10 | pancreas | 18q21.32 |
| rs3786898 | 19 | 33894043 | 1.86E-13 | pancreas | 19q13.11 |
| rs6020369 | 20 | 48835972 | 1.80E-13 | pancreas | 20q13.13 |
| rs9330813 | 22 | 46364161 | 1.09E-11 | pancreas | 22q13.31 |

| | | | | | |
|---|---|---|---|---|---|
| rs7874405 | 9 | 21980944 | 4.39E-08 | spleen | 9p21.3 |
| 9:91424602_GCTGTGCCCAGA_G | 9 | 91424602 | 4.04E-08 | spleen | 9q22.1 |
| rs233721 | 12 | 113031543 | 7.46E-12 | spleen | 12q24.13 |
| 13:108960380_TGCTG_T | 13 | 108960380 | 2.61E-09 | spleen | 13q33.3 |

**eTable 5: The detailed statistics of SNP-based heritability estimates via GCTA**

| MRIBAG | Heritability | SE | P-value |
|---|---|---|---|
| brain | 0.474981 | 0.020076 | 9.79E-149 |
| adipose | 0.377424 | 0.033602 | 5.05E-31 |
| heart | 0.285144 | 0.02309 | 1.31E-38 |
| kidney | 0.428789 | 0.024107 | 1.48E-76 |
| liver | 0.391912 | 0.033382 | 6.51E-36 |
| pancreas | 0.4513 | 0.029413 | 7.02E-59 |
| spleen | 0.328378 | 0.030221 | 4.19E-32 |

**eTable 6: The incremental R2 explained by the MRIBAG-PRS in split2 GWAS**

| $R^2$ | P | BETA | SE | MRIBAG |
|---|---|---|---|---|
| 0.02175592 | 7.63E-77 | 0.40350256 | 0.02162817 | brain |
| 0.01018814 | 1.30E-23 | 0.47424644 | 0.04722673 | adipose |
| 0.00826441 | 7.40E-28 | 0.44990947 | 0.04103825 | heart |
| 0.02025861 | 3.17E-64 | 0.71601925 | 0.04209912 | kidney |
| 0.02036249 | 5.93E-46 | 0.73145762 | 0.05113558 | liver |
| 0.01725355 | 1.69E-44 | 0.5510532 | 0.03920491 | pancreas |
| 0.0069537 | 3.85E-18 | 0.40733878 | 0.04682929 | spleen |

**eTable 7: Colocalization analyses between the 7 MRIBAGs and 11 ProtBAGs and 5 MetBAGs**

| Chromosome | Top Lead SNP | MRI BAG | OmicsBAG | Causal SNP | lABF_MRIBAG | lABF_OmicsBAG | SNP.PP.H4 | nsnps | PP.H0.abf | PP.H1.abf | PP.H2.abf | PP.H3.abf | PP.H4.abf |
|---|---|---|---|---|---|---|---|---|---|---|---|---|---|
| 15 | rs141866277 | liver | ProtBAG-Eye | rs150844304 | 18.9444365 | 7.71467629 | 0.29169602 | 1302 | 7.29E-08 | 0.00791804 | 4.35E-07 | 0.0463297 | 0.94575175 |
| 15 | rs141866277 | liver | ProtBAG-Hepatic | rs139974673 | 18.6136173 | 27.8055165 | 0.55175261 | 1302 | 3.78E-16 | 4.11E-11 | 9.87E-08 | 0.00972832 | 0.99027158 |
| 9 | 9:136138765_GCGCCCACCACTA_G | pancreas | ProtBAG-Heart | 9:136138765_GCGCCCACCACTA_G | 27.0481336 | 14.1400343 | 0.66681535 | 150 | 8.56E-14 | 7.81E-06 | 1.06E-10 | 0.00863656 | 0.99135563 |
| 19 | rs3786898 | pancreas | ProtBAG-Pulmonary | rs3786898 | 23.8468151 | 5.5174027 | 0.27209724 | 902 | 4.54E-09 | 0.04217096 | 1.49E-09 | 0.0128594 | 0.94496963 |
| 9 | 9:136138765_GCGCCCACCACTA_G | pancreas | ProtBAG-Hepatic | 9:136138765_GCGCCCACCACTA_G | 27.0481336 | 6.56697304 | 0.68218038 | 150 | 1.67E-10 | 0.01519139 | 1.83E-10 | 0.01568729 | 0.96912132 |
| 9 | 9:136138765_GCGCCCACCACTA_G | pancreas | ProtBAG-Renal | 9:136138765_GCGCCCACCACTA_G | 27.0481336 | 7.43273098 | 0.49781228 | 150 | 4.80E-11 | 0.00438228 | 9.42E-10 | 0.08505924 | 0.91055848 |
| 9 | 9:136138765_GCGCCCACCACTA_G | pancreas | ProtBAG-Endocrine | 9:136138765_GCGCCCACCACTA_G | 27.0481336 | 75.3650493 | 0.54638504 | 150 | 1.81E-40 | 1.65E-32 | 8.44E-11 | 0.00671151 | 0.99328849 |
| 10 | rs769008 | pancreas | ProtBAG-Endocrine | rs769009 | 20.1687362 | 8.71405827 | 0.35997507 | 606 | 1.02E-08 | 0.00426862 | 1.35E-08 | 0.00464861 | 0.99108274 |
| 9 | rs7874405 | spleen | ProtBAG-Reproductive_female | rs7874405 | 12.2777338 | 6.53802789 | 0.4391779 | 72 | 0.00028694 | 0.01458559 | 0.0003124 | 0.01491012 | 0.96990495 |
| 12 | rs233721 | spleen | ProtBAG-Pulmonary | rs2189272 | 17.6143407 | 9.14268483 | 0.29429228 | 1694 | 6.22E-08 | 0.01059956 | 6.37E-07 | 0.10765369 | 0.88174605 |
| 17 | rs534115641 | brain | MetBAG-Digestive | 17:44204299_AC_A | 41.8750679 | 23.9809439 | 0.41285654 | 1223 | 8.73E-25 | 2.30E-09 | 5.97E-17 | 0.15656923 | 0.84343077 |
| 17 | rs534115641 | brain | MetBAG-Hepatic | 17:44204299_AC_A | 41.8750679 | 6.99747245 | 0.17712171 | 1223 | 8.55E-18 | 0.02254579 | 6.37E-17 | 0.16728064 | 0.81017357 |
| 2 | rs954244 | kidney | MetBAG-Endocrine | rs7578604 | 12.4554355 | 4.24495643 | 0.59847533 | 26 | 0.00282183 | 0.14932955 | 8.60E-05 | 0.00370533 | 0.84405733 |
| 5 | 5:55860907_GC_G | kidney | MetBAG-Endocrine | 5:55860907_GC_G | 41.1409878 | 14.1341173 | 0.78343196 | 213 | 7.67E-20 | 7.24E-06 | 9.38E-17 | 0.00786133 | 0.99213143 |
| 8 | rs199922514 | liver | MetBAG-Endocrine | rs199922514 | 77.7462019 | 39.6383573 | 0.30794634 | 368 | 3.21E-47 | 7.07E-17 | 3.22E-33 | 0.00609843 | 0.99390157 |
| 10 | rs1539042 | liver | MetBAG-Endocrine | rs34370527 | 13.2453949 | 16.8670316 | 0.07843994 | 467 | 6.30E-10 | 1.34E-06 | 1.90E-05 | 0.03932045 | 0.9606592 |
| 19 | 19:19432290_AG_A | liver | MetBAG-Endocrine | rs58542926 | 20.8017853 | 242.104193 | 0.99918391 | 687 | 6.18E-110 | 4.86E-103 | 8.63E-09 | 0.06692839 | 0.9330716 |
| 10 | rs7924036 | liver | MetBAG-Digestive | rs2393977 | 32.41689 | 136.141194 | 0.03181364 | 965 | 1.77E-70 | 2.26E-58 | 8.61E-14 | 0.10918077 | 0.89081923 |
| 19 | 19:19432290_AG_A | liver | MetBAG-Digestive | 19:19432290_AG_A | 24.5910944 | 6.83425574 | 0.56622759 | 687 | 1.24E-09 | 0.0097499 | 2.24E-09 | 0.01661495 | 0.97363514 |

| | | | | | | | | | | | | | |
|---|---|---|---|---|---|---|---|---|---|---|---|---|---|
| 22 | rs2294915 | liver | MetBAG-Digestive | rs738408 | 65.5120216 | 39.360908 | 0.50692161 | 276 | 1.43E-41 | 1.21E-16 | 8.13E-28 | 0.00588036 | 0.99411964 |
| 8 | rs199922514 | liver | MetBAG-Hepatic | rs9987289 | 77.6849368 | 141.801131 | 0.49274256 | 368 | 2.34E-91 | 5.16E-61 | 1.92E-33 | 0.0032325 | 0.9967675 |
| 10 | rs7924036 | liver | MetBAG-Hepatic | 10:65265705_CA_C | 32.8771087 | 47.9628259 | 0.11124792 | 965 | 7.74E-32 | 9.90E-20 | 8.42E-14 | 0.10672159 | 0.89327841 |
| 10 | rs1539042 | liver | MetBAG-Hepatic | rs11186722 | 13.1952373 | 7.00025472 | 0.05277111 | 467 | 8.40E-06 | 0.01780158 | 2.07E-05 | 0.04292797 | 0.93924134 |
| 15 | rs141866277 | liver | MetBAG-Hepatic | rs190543502 | 17.9019029 | 5.7536386 | 0.2232899 | 1302 | 1.03E-06 | 0.11184494 | 2.14E-07 | 0.02238354 | 0.86577027 |
| 19 | 19:19432290_AG_A | liver | MetBAG-Hepatic | 19:19432290_AG_A | 24.5910944 | 106.001754 | 0.79085606 | 687 | 1.51E-52 | 1.19E-45 | 7.15E-10 | 0.00462569 | 0.99537431 |
| 22 | rs2294915 | liver | MetBAG-Hepatic | rs738408 | 65.5120216 | 47.0809328 | 0.43867459 | 276 | 5.51E-45 | 4.66E-20 | 8.19E-28 | 0.00592658 | 0.99407342 |
| 8 | rs199922514 | liver | MetBAG-Immune | rs9987289 | 77.6849368 | 7.42945574 | 0.31350821 | 368 | 3.06E-33 | 0.00674588 | 4.26E-32 | 0.09289334 | 0.90036078 |
| 10 | rs1539042 | liver | MetBAG-Immune | rs11186722 | 13.1952373 | 15.367587 | 0.04702575 | 467 | 1.77E-09 | 3.76E-06 | 2.06E-05 | 0.04265903 | 0.95731662 |
| 15 | rs141866277 | liver | MetBAG-Immune | rs139974673 | 18.6136173 | 22.0903628 | 0.68825331 | 1302 | 1.42E-13 | 1.54E-08 | 1.93E-07 | 0.01994341 | 0.98005638 |
| 19 | 19:19432290_AG_A | liver | MetBAG-Immune | rs739846 | 21.6743432 | 28.8674216 | 0.55288871 | 687 | 6.00E-18 | 4.72E-11 | 4.20E-09 | 0.03205063 | 0.96794937 |
| 8 | rs199922514 | liver | MetBAG-Metabolic | rs4240624 | 77.7110369 | 12.0363929 | 0.28531389 | 368 | 2.44E-35 | 5.38E-05 | 8.59E-32 | 0.18846751 | 0.81147868 |
| 10 | rs7924036 | liver | MetBAG-Metabolic | rs3841602 | 33.9859384 | 49.4685813 | 0.0935051 | 965 | 4.78E-33 | 6.10E-21 | 8.24E-14 | 0.10441372 | 0.89558628 |
| 15 | rs141866277 | liver | MetBAG-Metabolic | rs150844304 | 18.9444365 | 43.2004974 | 0.75467329 | 1302 | 7.69E-23 | 8.35E-18 | 7.01E-08 | 0.00661804 | 0.99338189 |
| 9 | 9:136138765_GCGCCCACCACTA_G | pancreas | MetBAG-Endocrine | 9:136138765_GCGCCCACCACTA_G | 27.0481336 | 16.0584876 | 0.75881125 | 150 | 1.44E-14 | 1.31E-06 | 6.55E-11 | 0.00498487 | 0.99501382 |
| 9 | 9:136138765_GCGCCCACCACTA_G | pancreas | MetBAG-Digestive | 9:136138765_GCGCCCACCACTA_G | 27.0481336 | 103.284089 | 0.91883027 | 150 | 2.29E-52 | 2.09E-44 | 3.21E-11 | 0.00192782 | 0.99807218 |
| 9 | 9:136138765_GCGCCCACCACTA_G | pancreas | MetBAG-Hepatic | 9:136138765_GCGCCCACCACTA_G | 27.0481336 | 58.8795848 | 0.65782026 | 150 | 3.14E-33 | 2.87E-25 | 7.60E-11 | 0.00594191 | 0.99405809 |
| 9 | 9:136138765_GCGCCCACCACTA_G | pancreas | MetBAG-Immune | 9:136138765_GCGCCCACCACTA_G | 27.0481336 | 68.8826485 | 0.71498073 | 150 | 1.55E-37 | 1.41E-29 | 7.50E-11 | 0.00585283 | 0.99414717 |
| 12 | rs233721 | spleen | MetBAG-Hepatic | rs233721 | 20.6644086 | 11.9726126 | 0.9638791 | 1694 | 6.37E-10 | 0.00010859 | 8.26E-08 | 0.01308565 | 0.98680569 |
| 12 | rs233721 | spleen | MetBAG-Immune | rs233721 | 20.6644086 | 10.9841428 | 0.98766015 | 1694 | 1.76E-09 | 0.0003005 | 5.21E-08 | 0.00788351 | 0.99181594 |
| 13 | 13:108960380_TGCTG_T | spleen | MetBAG-Metabolic | 13:108960380_TGCTG_T | 15.1623154 | 3.73744072 | 0.70014672 | 100 | 0.00034961 | 0.19124305 | 5.45E-06 | 0.00217358 | 0.80622831 |

**eTable 8: Results for the survival analyses for predicting all-cause mortality**

**a): all-cause mortality considering all diseases**

| hazard_ratio | CI_lower_bound | CI_upper_bound | p_value | n_case | n_noncase | MRIBAG |
|---|---|---|---|---|---|---|
| 1.2574258 | 1.16252642 | 1.36007202 | 1.06E-08 | 660 | 35628 | brain |
| 1.18834656 | 1.06994228 | 1.31985395 | 0.00127122 | 351 | 23470 | adipose |
| 0.9792014 | 0.91014343 | 1.0534992 | 0.57325498 | 665 | 33031 | kidney |
| 0.98443848 | 0.91045137 | 1.0644381 | 0.69399466 | 646 | 34064 | heart |
| 0.77343491 | 0.69153875 | 0.86502969 | 6.83E-06 | 339 | 23678 | liver |
| 1.11203907 | 1.02741891 | 1.20362871 | 0.00854289 | 453 | 26800 | pancreas |
| 0.80160917 | 0.73190279 | 0.87795439 | 1.90E-06 | 412 | 25880 | spleen |
| 0.88101546 | 0.78395862 | 0.99008828 | 0.03340068 | 292 | 15481 | brain_PRS |
| 1.0783669 | 0.99458905 | 1.16920167 | 0.06747849 | 769 | 38244 | adipose_PRS |
| 1.05402661 | 0.98240942 | 1.13086466 | 0.14274919 | 769 | 38244 | heart_PRS |
| 0.99732727 | 0.93109545 | 1.06827037 | 0.93915321 | 769 | 38244 | kidney_PRS |
| 0.92799112 | 0.85571928 | 1.00636684 | 0.07083335 | 769 | 38244 | liver_PRS |
| 0.9875368 | 0.91359769 | 1.06745993 | 0.75211259 | 769 | 38244 | pancreas_PRS |
| 0.93948043 | 0.86670028 | 1.01837221 | 0.12915472 | 769 | 38244 | spleen_PRS |

**b): all-cause mortality for disease-free analyses (sample size decreases dramatically)**

| hazard_ratio | CI_lower_bound | CI_upper_bound | p_value | n_case | n_noncase | MRIBAG |
|---|---|---|---|---|---|---|
| 1.04503211 | 0.69671899 | 1.56747859 | 0.831371 | 26 | 5912 | brain |
| 1.38042997 | 0.88563344 | 2.15166549 | 0.15454805 | 18 | 3952 | adipose |
| 0.81466962 | 0.56956071 | 1.16526048 | 0.26167743 | 29 | 5439 | kidney |
| 0.90168135 | 0.61018937 | 1.33242121 | 0.60344035 | 28 | 5627 | heart |
| 0.79547207 | 0.45476837 | 1.39142443 | 0.42250978 | 16 | 3987 | liver |
| 1.15139379 | 0.75302839 | 1.76050156 | 0.51524238 | 21 | 4489 | pancreas |
| 0.6251624 | 0.39100093 | 0.99955779 | 0.04978447 | 16 | 4322 | spleen |
| 1.44270661 | 0.77329294 | 2.69160917 | 0.2493477 | 10 | 2566 | brain_PRS |
| 1.39702858 | 0.95284687 | 2.04827127 | 0.08679331 | 32 | 6251 | adipose_PRS |
| 0.70839849 | 0.50092059 | 1.00181232 | 0.05120906 | 32 | 6251 | heart_PRS |
| 1.00975236 | 0.71248642 | 1.43104459 | 0.9564967 | 32 | 6251 | kidney_PRS |
| 1.36243856 | 0.91797344 | 2.02210516 | 0.12474934 | 32 | 6251 | liver_PRS |
| 0.93432187 | 0.62182529 | 1.40386274 | 0.74365518 | 32 | 6251 | pancreas_PRS |
| 0.80490151 | 0.5410609 | 1.19740022 | 0.28417729 | 32 | 6251 | spleen_PRS |

**c): all-cause mortality with liver MRIBAG features**

| hazard_ratio | CI_lower_bound | CI_upper_bound | p_value | n_case | n_noncase | MRIBAG features |
|---|---|---|---|---|---|---|
| 0.77343491 | 0.69153875 | 0.86502969 | 6.83E-06 | 339 | 23678 | Liver MRIBAG |
| 1.27045745 | 1.14975563 | 1.4038306 | 2.60E-06 | 351 | 26701 | Liver_volume_21080-2.0 |
| 1.06419206 | 0.964235 | 1.17451113 | 0.21635818 | 351 | 26701 | Liver_PDFF_(fat_fraction)_21088-2.0 |
| 0.91944003 | 0.82409339 | 1.02581815 | 0.132678 | 351 | 26701 | Liver_iron_21089-2.0 |
| 1.32215362 | 1.20287701 | 1.45325763 | 7.07E-09 | 351 | 26701 | Liver_iron_corrected_T1_(ct1)_40062-2.0 |

**d): all-cause mortality with spleen MRIBAG features**

| hazard_ratio | CI_lower_bound | CI_upper_bound | p_value | n_case | n_noncase | MRIBAG features |
|---|---|---|---|---|---|---|
| 0.80160917 | 0.73190279 | 0.87795439 | 1.90E-06 | 412 | 25880 | Spleen MRIBAG |

| | | | | | | |
|---|---|---|---|---|---|---|
| 1.33574257 | 1.23401729 | 1.44585349 | 7.90E-13 | 424 | 29236 | Spleen_volume_21083-2.0 |
| 0.97341503 | 0.89501445 | 1.05868327 | 0.52940239 | 424 | 29236 | Spleen_iron_-_IDEAL_21170-2.0 |
| 0.98107229 | 0.88936051 | 1.08224148 | 0.70274546 | 424 | 29236 | Spleen_iron_-_protocol_normalised_21173-2.0 |

**e): all-cause mortality with spleen MRIBAG features, including the spleen imaging features as an additional covariate**

| **hazard_ratio** | **CI_lower_bound** | **CI_upper_bound** | **p_value** | **n_case** | **n_noncase** | **MRIBAG features** |
|---|---|---|---|---|---|---|
| 0.80160917 | 0.73190279 | 0.87795439 | 1.90E-06 | 412 | 25880 | Spleen MRIBAG |
| 1.01535846 | 0.8921064 | 1.1556 | 0.81 | 424 | 29236 | Spleen_volume_21083-2.0 |
| 0.7944144 | 0.723313 | 0.8725 | 1.50E-06 | 424 | 29236 | Spleen_iron_-_IDEAL_21170-2.0 |
| 0.7071428 | 0.632836 | 0.79017 | 9.50E-10 | 424 | 29236 | Spleen_iron_-_protocol_normalised_21173-2.0 |

**eTable 9: Results for the mediation analysis between brain ProtBAG, brain MRIBAG, and LLD subtypes**

a1 represents the effect of the independent variable (e.g., Brain ProtBAG) on the mediator (e.g., Brain MRIBAG), while c1 is the effect of the mediator on the outcome (e.g., LLD1). c2 reflects the direct effect of the independent variable on the outcome, bypassing the mediator. The indirect effect is the product of a1 and c1, quantifying the mediated pathway; total effect is the sum of direct (c2) and indirect effects; and proportion mediated (prop_mediated) indicates the fraction of the total effect that is explained by the mediator.

**a): brain ProtBAG -- brain MRIBAG -- LLD subtypes**

| Outcome | pathway | EST | SE | P-value | CI.lower | CI.upper | STD.all |
|---|---|---|---|---|---|---|---|
| LLD_1 | c1 | -0.0846693 | 0.01048422 | 6.66E-16 | -0.1050047 | -0.06347 | -0.1352149 |
| LLD_1 | c2 | 0.01012126 | 0.01070334 | 0.34434374 | -0.0133969 | 0.02898602 | 0.01548619 |
| LLD_1 | a1 | 0.09110585 | 0.01651184 | 3.44E-08 | 0.05721678 | 0.12632933 | 0.08728859 |
| LLD_1 | indirect | -0.0077139 | NA | NA | NA | NA | NA |
| LLD_1 | total | 0.0024074 | NA | NA | NA | NA | NA |
| LLD_1 | prop_mediated | -3.2042368 | NA | NA | NA | NA | NA |
| LLD_2 | c1 | 0.09875053 | 0.00763764 | 0 | 0.08398177 | 0.11410959 | 0.20831517 |
| LLD_2 | c2 | 0.00247252 | 0.00764037 | 0.7462308 | -0.0126207 | 0.01805624 | 0.00499728 |
| LLD_2 | a1 | 0.09110585 | 0.01635658 | 2.55E-08 | 0.05664501 | 0.12503391 | 0.08728859 |
| LLD_2 | indirect | 0.00899675 | NA | NA | NA | NA | NA |
| LLD_2 | total | 0.01146927 | NA | NA | NA | NA | NA |
| LLD_2 | prop_mediated | 0.78442192 | NA | NA | NA | NA | NA |

**b): brain MRIBAG -- brain ProtBAG -- LLD subtypes**

| Outcome | pathway | EST | SE | P-value | CI.lower | CI.upper | STD.all |
|---|---|---|---|---|---|---|---|
| LLD_1 | c1 | 0.01012126 | 0.01109639 | 0.36170475 | -0.0129749 | 0.03224862 | 0.01548619 |
| LLD_1 | c2 | -0.0846693 | 0.0100737 | 0 | -0.1041383 | -0.0639579 | -0.1352149 |
| LLD_1 | a1 | 0.08442812 | 0.01538936 | 4.11E-08 | 0.05331431 | 0.11317458 | 0.08812029 |
| LLD_1 | indirect | 8.55E-04 | NA | NA | NA | NA | NA |
| LLD_1 | total | -0.0838147 | NA | NA | NA | NA | NA |
| LLD_1 | prop_mediated | -0.0101953 | NA | NA | NA | NA | NA |
| LLD_2 | c1 | 0.00247252 | 0.0076545 | 0.7466833 | -0.0130348 | 0.01700746 | 0.00499728 |
| LLD_2 | c2 | 0.09875053 | 0.00791065 | 0 | 0.08262821 | 0.11372114 | 0.20831517 |
| LLD_2 | a1 | 0.08442812 | 0.01552085 | 5.34E-08 | 0.05160248 | 0.11337959 | 0.08812029 |
| LLD_2 | indirect | 2.09E-04 | NA | NA | NA | NA | NA |
| LLD_2 | total | 0.09895928 | NA | NA | NA | NA | NA |
| LLD_2 | prop_mediated | 0.00210946 | NA | NA | NA | NA | NA |

**eTable 10: Concordant hits for the sex-stratified ProWAS analyses**

| MRIBAG | Protein | Female | Male | logP_female | logP_male | P-thres | is_sig_female | is_sig_male | Direction |
|---|---|---|---|---|---|---|---|---|---|
| brain | NCAN | -0.0993783 | -0.1413699 | 4.95368916 | 8.36066762 | 5.61195685 | FALSE | TRUE | Concordant |
| brain | MOG | -0.0556266 | -0.1209231 | 1.77901255 | 6.2358944 | 5.61195685 | FALSE | TRUE | Concordant |
| brain | PTPRS | -0.0707744 | -0.1187677 | 2.9619931 | 6.4363529 | 5.61195685 | FALSE | TRUE | Concordant |
| brain | GDF15 | 0.09037905 | 0.12839726 | 3.78079678 | 5.62439011 | 5.61195685 | FALSE | TRUE | Concordant |
| brain | BCAN | -0.128619 | -0.1532513 | 7.97581306 | 9.56023308 | 5.61195685 | TRUE | TRUE | Concordant |
| brain | ADAM22 | -0.0688142 | -0.1302121 | 2.77260098 | 7.44434457 | 5.61195685 | FALSE | TRUE | Concordant |
| brain | SLITRK1 | -0.1427178 | -0.0758738 | 6.61051062 | 1.94358531 | 5.61195685 | TRUE | FALSE | Concordant |
| adipose | PRCP | 0.11579073 | 0.16174279 | 3.53403366 | 5.95741824 | 5.61195685 | FALSE | TRUE | Concordant |
| adipose | PLAT | 0.14824126 | 0.18799169 | 5.09720283 | 7.27267428 | 5.61195685 | FALSE | TRUE | Concordant |
| adipose | HGF | 0.12533277 | 0.17869253 | 4.0928511 | 7.10121124 | 5.61195685 | FALSE | TRUE | Concordant |
| adipose | NPPC | 0.1690675 | 0.06064528 | 7.64666103 | 1.24411418 | 5.61195685 | TRUE | FALSE | Concordant |
| adipose | SLC39A5 | 0.16391211 | 0.08879256 | 7.00174255 | 2.19570688 | 5.61195685 | TRUE | FALSE | Concordant |
| adipose | RARRES2 | 0.13143159 | 0.19183595 | 4.03940963 | 7.91521173 | 5.61195685 | FALSE | TRUE | Concordant |
| adipose | SELE | 0.05999701 | 0.17700268 | 1.2850476 | 7.03767162 | 5.61195685 | FALSE | TRUE | Concordant |
| adipose | CPM | 0.09881901 | 0.17763509 | 2.55045762 | 6.86635861 | 5.61195685 | FALSE | TRUE | Concordant |
| adipose | GDF15 | 0.08574032 | 0.2108657 | 2.02900882 | 8.22189677 | 5.61195685 | FALSE | TRUE | Concordant |
| adipose | CA14 | -0.0950296 | -0.1971973 | 2.64054296 | 8.24227277 | 5.61195685 | FALSE | TRUE | Concordant |
| adipose | CCL27 | 0.10326169 | 0.16385399 | 2.84636731 | 6.61251842 | 5.61195685 | FALSE | TRUE | Concordant |
| adipose | CCL16 | 0.09247839 | 0.15589177 | 2.47793421 | 5.75940335 | 5.61195685 | FALSE | TRUE | Concordant |
| adipose | ASGR1 | 0.17047195 | 0.13445986 | 6.82718948 | 4.20515137 | 5.61195685 | TRUE | FALSE | Concordant |
| adipose | PRAP1 | 0.10325074 | 0.18332291 | 2.3806422 | 6.13565687 | 5.61195685 | FALSE | TRUE | Concordant |
| adipose | CFI | 0.08436283 | 0.17847718 | 1.61105756 | 5.79889618 | 5.61195685 | FALSE | TRUE | Concordant |
| adipose | IGSF9 | 0.18160397 | 0.15166596 | 6.24756333 | 3.8547573 | 5.61195685 | TRUE | FALSE | Concordant |
| kidney | PIK3IP1 | 0.20272857 | 0.24082247 | 13.4630726 | 17.5083379 | 5.61195685 | TRUE | TRUE | Concordant |
| kidney | PGLYRP1 | 0.12698682 | 0.10017296 | 6.16351907 | 3.74077693 | 5.61195685 | TRUE | FALSE | Concordant |
| kidney | PI3 | 0.12228029 | 0.17047297 | 5.59201535 | 9.572968 | 5.61195685 | FALSE | TRUE | Concordant |
| kidney | OGN | 0.16884759 | 0.2132129 | 10.132706 | 12.2744347 | 5.61195685 | TRUE | TRUE | Concordant |
| kidney | PODXL2 | 0.14100112 | 0.16257086 | 7.55936698 | 8.4420061 | 5.61195685 | TRUE | TRUE | Concordant |
| kidney | PLAUR | 0.13674731 | 0.07101545 | 7.00432892 | 1.97289823 | 5.61195685 | TRUE | FALSE | Concordant |
| kidney | PRSS2 | 0.15654879 | 0.10877993 | 9.03317449 | 4.39268476 | 5.61195685 | TRUE | FALSE | Concordant |
| kidney | IL18BP | 0.1124843 | 0.12940237 | 4.73531766 | 5.78678125 | 5.61195685 | FALSE | TRUE | Concordant |
| kidney | KLK11 | 0.12084613 | 0.16678978 | 5.6736551 | 9.3968975 | 5.61195685 | TRUE | TRUE | Concordant |
| kidney | KLK6 | 0.17753175 | 0.22232029 | 11.4216129 | 16.1963186 | 5.61195685 | TRUE | TRUE | Concordant |

| | | | | | | | | | |
|---|---|---|---|---|---|---|---|---|---|
| kidney | KLK8 | 0.1023798 | 0.15254302 | 4.10640764 | 7.79396352 | 5.61195685 | FALSE | TRUE | Concordant |
| kidney | JAM2 | 0.17161235 | 0.17749055 | 10.0378298 | 10.5303264 | 5.61195685 | TRUE | TRUE | Concordant |
| kidney | HAVCR2 | 0.15208355 | 0.09235006 | 8.4414139 | 3.04267599 | 5.61195685 | TRUE | FALSE | Concordant |
| kidney | GUCA2A | 0.11017795 | 0.15444187 | 4.42429546 | 7.75910859 | 5.61195685 | FALSE | TRUE | Concordant |
| kidney | IFNGR1 | 0.19895447 | 0.15786159 | 13.9700768 | 8.6181671 | 5.61195685 | TRUE | TRUE | Concordant |
| kidney | IGF1R | 0.12707061 | 0.11532817 | 5.83284298 | 4.79500743 | 5.61195685 | TRUE | FALSE | Concordant |
| kidney | IGFBP4 | 0.23310678 | 0.24761175 | 17.1878709 | 16.9111765 | 5.61195685 | TRUE | TRUE | Concordant |
| kidney | IGFBP6 | 0.24430878 | 0.2808135 | 19.9834059 | 24.311588 | 5.61195685 | TRUE | TRUE | Concordant |
| kidney | IL10RB | 0.17794556 | 0.11984672 | 11.3580264 | 4.97035933 | 5.61195685 | TRUE | FALSE | Concordant |
| kidney | HSPG2 | 0.23323386 | 0.21950504 | 16.5546052 | 13.9657512 | 5.61195685 | TRUE | TRUE | Concordant |
| kidney | HYOU1 | 0.15926143 | 0.12929457 | 9.31457509 | 5.99479416 | 5.61195685 | TRUE | TRUE | Concordant |
| kidney | MSMB | 0.07396977 | 0.14387065 | 2.39654153 | 7.10114991 | 5.61195685 | FALSE | TRUE | Concordant |
| kidney | NBL1 | 0.19917961 | 0.17285691 | 14.2530982 | 9.68698086 | 5.61195685 | TRUE | TRUE | Concordant |
| kidney | NCAN | 0.06868163 | 0.19227398 | 2.02968326 | 11.9424272 | 5.61195685 | FALSE | TRUE | Concordant |
| kidney | MIA | 0.13074704 | 0.18929818 | 6.65058118 | 12.0868242 | 5.61195685 | TRUE | TRUE | Concordant |
| kidney | MMP3 | 0.09229534 | 0.14463531 | 3.53037801 | 7.21851482 | 5.61195685 | FALSE | TRUE | Concordant |
| kidney | MOG | 0.17998222 | 0.2549413 | 10.6399231 | 20.3214684 | 5.61195685 | TRUE | TRUE | Concordant |
| kidney | MFAP5 | 0.1561947 | 0.1996726 | 8.27537905 | 12.4062925 | 5.61195685 | TRUE | TRUE | Concordant |
| kidney | NPPC | 0.18226519 | 0.22459514 | 12.0386146 | 16.496532 | 5.61195685 | TRUE | TRUE | Concordant |
| kidney | NPTXR | 0.08335991 | 0.15593788 | 2.86610091 | 8.07402856 | 5.61195685 | FALSE | TRUE | Concordant |
| kidney | NPDC1 | 0.28510612 | 0.27583041 | 27.1252962 | 24.2574236 | 5.61195685 | TRUE | TRUE | Concordant |
| kidney | NCS1 | 0.0419493 | 0.12776823 | 0.87763896 | 5.6957419 | 5.61195685 | FALSE | TRUE | Concordant |
| kidney | NECTIN2 | 0.14148121 | 0.13398884 | 7.41402644 | 6.24174922 | 5.61195685 | TRUE | TRUE | Concordant |
| kidney | NECTIN4 | 0.1958603 | 0.2268272 | 14.1041864 | 17.1204706 | 5.61195685 | TRUE | TRUE | Concordant |
| kidney | NCR1 | 0.12965439 | 0.10171637 | 6.48676153 | 3.85769723 | 5.61195685 | TRUE | FALSE | Concordant |
| kidney | LGALS9 | 0.12225788 | 0.14054021 | 4.84226323 | 6.18381359 | 5.61195685 | FALSE | TRUE | Concordant |
| kidney | LRP11 | 0.13662155 | 0.12474978 | 6.68309971 | 5.43286324 | 5.61195685 | TRUE | FALSE | Concordant |
| kidney | LGALS7_LGALS7B | 0.10808371 | 0.12660981 | 4.63980663 | 5.68438839 | 5.61195685 | FALSE | TRUE | Concordant |
| kidney | LAIR1 | 0.16794012 | 0.13420287 | 9.1363934 | 5.74562953 | 5.61195685 | TRUE | TRUE | Concordant |
| kidney | LAMA4 | 0.1242649 | 0.02549115 | 6.05054949 | 0.46302294 | 5.61195685 | TRUE | FALSE | Concordant |
| kidney | LGALS3 | 0.1447169 | 0.11050959 | 7.02309602 | 4.45472221 | 5.61195685 | TRUE | FALSE | Concordant |
| kidney | LAYN | 0.1984077 | 0.19376597 | 13.2017565 | 11.7313888 | 5.61195685 | TRUE | TRUE | Concordant |
| kidney | LCN2 | 0.14736187 | 0.11286757 | 7.86766301 | 4.34011441 | 5.61195685 | TRUE | FALSE | Concordant |
| kidney | LGALS1 | 0.14567937 | 0.09900267 | 6.8039482 | 3.4049534 | 5.61195685 | TRUE | FALSE | Concordant |
| kidney | MEPE | 0.13250977 | 0.1147517 | 6.69708351 | 4.71634018 | 5.61195685 | TRUE | FALSE | Concordant |

| | | | | | | | | | |
|---|---|---|---|---|---|---|---|---|---|
| kidney | LTBR | 0.18383508 | 0.11337492 | 12.3495848 | 4.50411882 | 5.61195685 | TRUE | FALSE | Concordant |
| kidney | LY6D | 0.19870481 | 0.20365414 | 13.6807115 | 13.5114899 | 5.61195685 | TRUE | TRUE | Concordant |
| kidney | MANSC1 | 0.15710748 | 0.12032782 | 9.3624133 | 5.24589579 | 5.61195685 | TRUE | FALSE | Concordant |
| kidney | SLC39A5 | 0.11805122 | 0.13194097 | 5.12469021 | 5.91900762 | 5.61195685 | FALSE | TRUE | Concordant |
| kidney | SEZ6L | 0.13393529 | 0.20918469 | 6.31372889 | 14.001656 | 5.61195685 | TRUE | TRUE | Concordant |
| kidney | SPP1 | 0.08473196 | 0.12758643 | 2.83940679 | 5.8286093 | 5.61195685 | FALSE | TRUE | Concordant |
| kidney | SMOC2 | 0.17341424 | 0.12730738 | 10.5743874 | 5.46275027 | 5.61195685 | TRUE | FALSE | Concordant |
| kidney | SORCS2 | 0.18626865 | 0.17110887 | 11.6424297 | 9.03790647 | 5.61195685 | TRUE | TRUE | Concordant |
| kidney | SPINK1 | 0.18273953 | 0.1890391 | 12.0253805 | 11.7908153 | 5.61195685 | TRUE | TRUE | Concordant |
| kidney | SPINK5 | 0.10692939 | 0.21753043 | 4.483641 | 15.8449476 | 5.61195685 | FALSE | TRUE | Concordant |
| kidney | QPCT | 0.12569051 | 0.10593602 | 6.0672124 | 4.16986725 | 5.61195685 | TRUE | FALSE | Concordant |
| kidney | RELT | 0.24857173 | 0.2342687 | 20.2789403 | 17.668301 | 5.61195685 | TRUE | TRUE | Concordant |
| kidney | PTGDS | 0.19351976 | 0.22238154 | 13.1696309 | 15.6844774 | 5.61195685 | TRUE | TRUE | Concordant |
| kidney | PTPRF | 0.06438565 | 0.14119597 | 1.77976201 | 7.08974009 | 5.61195685 | FALSE | TRUE | Concordant |
| kidney | PTPRN2 | 0.20990698 | 0.25244033 | 15.5381594 | 20.6545006 | 5.61195685 | TRUE | TRUE | Concordant |
| kidney | PTPRS | 0.10491261 | 0.14051986 | 4.51077979 | 7.04829709 | 5.61195685 | FALSE | TRUE | Concordant |
| kidney | SCARB2 | 0.18870763 | 0.20267101 | 10.5383131 | 11.9492128 | 5.61195685 | TRUE | TRUE | Concordant |
| kidney | SCARF2 | 0.17629655 | 0.1046559 | 8.73628914 | 3.57210411 | 5.61195685 | TRUE | FALSE | Concordant |
| kidney | SCG2 | 0.19051229 | 0.21087129 | 13.0936608 | 14.892798 | 5.61195685 | TRUE | TRUE | Concordant |
| kidney | SCG3 | 0.0825407 | 0.18317738 | 2.84180535 | 10.8453649 | 5.61195685 | FALSE | TRUE | Concordant |
| kidney | SCGB1A1 | 0.08420468 | 0.14572864 | 2.90906649 | 7.19347868 | 5.61195685 | FALSE | TRUE | Concordant |
| kidney | SEMA3F | 0.1888033 | 0.16380051 | 11.303563 | 8.68752278 | 5.61195685 | TRUE | TRUE | Concordant |
| kidney | ROR1 | 0.14892889 | 0.13988878 | 7.82124259 | 6.70119657 | 5.61195685 | TRUE | TRUE | Concordant |
| kidney | RETN | 0.16230114 | 0.11999616 | 9.61532748 | 5.11629334 | 5.61195685 | TRUE | FALSE | Concordant |
| kidney | RGMA | 0.13572099 | 0.10775422 | 6.87854867 | 4.31471188 | 5.61195685 | TRUE | FALSE | Concordant |
| kidney | RGMB | 0.21360248 | 0.18131726 | 16.4441823 | 11.3492317 | 5.61195685 | TRUE | TRUE | Concordant |
| kidney | RSPO3 | 0.18120724 | 0.19125338 | 11.2022535 | 11.0980696 | 5.61195685 | TRUE | TRUE | Concordant |
| kidney | RTBDN | 0.13899169 | 0.20350329 | 6.87598167 | 13.5899637 | 5.61195685 | TRUE | TRUE | Concordant |
| kidney | VSIG4 | 0.14077975 | 0.17556013 | 6.75388314 | 9.25440211 | 5.61195685 | TRUE | TRUE | Concordant |
| kidney | VWC2 | 0.16926357 | 0.14091146 | 9.99542164 | 6.4845636 | 5.61195685 | TRUE | TRUE | Concordant |
| kidney | XG | 0.18216817 | 0.21234816 | 10.5037841 | 13.9533626 | 5.61195685 | TRUE | TRUE | Concordant |
| kidney | WFIKKN2 | 0.12086886 | 0.13980677 | 5.11061623 | 6.1414476 | 5.61195685 | FALSE | TRUE | Concordant |
| kidney | WNT9A | 0.11501883 | 0.13787885 | 4.33346661 | 5.83170936 | 5.61195685 | FALSE | TRUE | Concordant |
| kidney | VASN | 0.09105177 | 0.1475944 | 3.25926089 | 7.5201306 | 5.61195685 | FALSE | TRUE | Concordant |
| kidney | TFF3 | 0.11720059 | 0.20708918 | 5.39990022 | 13.0161066 | 5.61195685 | FALSE | TRUE | Concordant |

| | | | | | | | | | |
|---|---|---|---|---|---|---|---|---|---|
| kidney | TGFA | 0.13416673 | 0.10499806 | 6.64014813 | 3.89878433 | 5.61195685 | TRUE | FALSE | Concordant |
| kidney | TGFBR2 | 0.22470355 | 0.18511873 | 16.0015202 | 10.6067444 | 5.61195685 | TRUE | TRUE | Concordant |
| kidney | TGFBR3 | 0.12603165 | 0.19077023 | 6.18350596 | 12.3251786 | 5.61195685 | TRUE | TRUE | Concordant |
| kidney | THBD | 0.15035709 | 0.12477934 | 8.10827568 | 5.60042736 | 5.61195685 | TRUE | FALSE | Concordant |
| kidney | TIMP4 | 0.12225097 | 0.16051568 | 5.74360834 | 8.62455778 | 5.61195685 | TRUE | TRUE | Concordant |
| kidney | THY1 | 0.15471894 | 0.16315421 | 8.57391719 | 8.50858164 | 5.61195685 | TRUE | TRUE | Concordant |
| kidney | TAFA5 | 0.2385726 | 0.26895722 | 19.6005811 | 22.8716788 | 5.61195685 | TRUE | TRUE | Concordant |
| kidney | TREML2 | 0.12039111 | 0.0384828 | 5.63769756 | 0.82568201 | 5.61195685 | TRUE | FALSE | Concordant |
| kidney | TNFRSF11A | 0.1646295 | 0.15950544 | 9.15893735 | 8.34124167 | 5.61195685 | TRUE | TRUE | Concordant |
| kidney | TNFRSF12A | 0.15308511 | 0.12714133 | 7.67735178 | 5.51734558 | 5.61195685 | TRUE | FALSE | Concordant |
| kidney | TNFRSF13B | 0.14557959 | 0.08149191 | 8.00789392 | 2.57674966 | 5.61195685 | TRUE | FALSE | Concordant |
| kidney | TNFRSF14 | 0.14464949 | 0.10027051 | 7.61720317 | 3.69759659 | 5.61195685 | TRUE | FALSE | Concordant |
| kidney | TNFRSF19 | 0.22896631 | 0.20618824 | 17.8956738 | 13.7587667 | 5.61195685 | TRUE | TRUE | Concordant |
| kidney | TNFRSF1A | 0.23511409 | 0.22333678 | 17.3619596 | 14.7832653 | 5.61195685 | TRUE | TRUE | Concordant |
| kidney | TNFRSF1B | 0.16214758 | 0.15448461 | 9.27190358 | 7.65850969 | 5.61195685 | TRUE | TRUE | Concordant |
| kidney | TNFRSF21 | 0.17342946 | 0.15761083 | 10.9943446 | 8.7019867 | 5.61195685 | TRUE | TRUE | Concordant |
| kidney | TNFRSF4 | 0.13613702 | 0.13475978 | 6.93198919 | 6.23283102 | 5.61195685 | TRUE | TRUE | Concordant |
| kidney | COL18A1 | 0.18056113 | 0.17648401 | 10.9728816 | 10.0261297 | 5.61195685 | TRUE | TRUE | Concordant |
| kidney | COL6A3 | 0.21738053 | 0.21662139 | 15.2615936 | 13.3460248 | 5.61195685 | TRUE | TRUE | Concordant |
| kidney | COLEC12 | 0.24205201 | 0.2294775 | 16.9537711 | 14.6891798 | 5.61195685 | TRUE | TRUE | Concordant |
| kidney | CLEC4D | 0.14941598 | 0.06966417 | 8.18773964 | 2.03755397 | 5.61195685 | TRUE | FALSE | Concordant |
| kidney | CLMP | 0.20141053 | 0.28681157 | 11.6030465 | 23.4306821 | 5.61195685 | TRUE | TRUE | Concordant |
| kidney | CLEC1A | 0.14188898 | 0.12205516 | 7.54285797 | 5.29691251 | 5.61195685 | TRUE | FALSE | Concordant |
| kidney | CX3CL1 | 0.12260358 | 0.09669735 | 5.83183294 | 3.51547539 | 5.61195685 | TRUE | FALSE | Concordant |
| kidney | CSF1 | 0.1389105 | 0.13848426 | 7.09709682 | 6.47292492 | 5.61195685 | TRUE | TRUE | Concordant |
| kidney | CST3 | 0.20146205 | 0.25340814 | 12.6013321 | 18.7145434 | 5.61195685 | TRUE | TRUE | Concordant |
| kidney | CST6 | 0.12702586 | 0.16960374 | 5.7774316 | 9.41615473 | 5.61195685 | TRUE | TRUE | Concordant |
| kidney | CD46 | 0.14094995 | 0.09838816 | 7.17162964 | 3.6231318 | 5.61195685 | TRUE | FALSE | Concordant |
| kidney | CD55 | 0.14762461 | 0.13264202 | 7.71362628 | 5.98886645 | 5.61195685 | TRUE | TRUE | Concordant |
| kidney | CD59 | 0.22580017 | 0.23794313 | 15.8551967 | 17.0483132 | 5.61195685 | TRUE | TRUE | Concordant |
| kidney | CD74 | 0.13333255 | 0.11972247 | 6.48430687 | 4.89369155 | 5.61195685 | TRUE | FALSE | Concordant |
| kidney | CD79B | 0.1393541 | 0.12358303 | 7.36313696 | 5.45288301 | 5.61195685 | TRUE | FALSE | Concordant |
| kidney | CD83 | 0.13020511 | 0.10799163 | 6.25578996 | 4.19943685 | 5.61195685 | TRUE | FALSE | Concordant |
| kidney | CD93 | 0.13630384 | 0.09134472 | 6.77584543 | 3.10079083 | 5.61195685 | TRUE | FALSE | Concordant |
| kidney | CD99 | 0.14155151 | 0.10312471 | 6.8514823 | 3.82566225 | 5.61195685 | TRUE | FALSE | Concordant |

| | | | | | | | | | |
|---|---|---|---|---|---|---|---|---|---|
| kidney | CD99L2 | 0.15055718 | 0.14397565 | 7.54311665 | 7.03107985 | 5.61195685 | TRUE | TRUE | Concordant |
| kidney | CD38 | 0.16913488 | 0.18086544 | 9.98068123 | 10.6853612 | 5.61195685 | TRUE | TRUE | Concordant |
| kidney | CD200 | 0.10905122 | 0.15264938 | 4.502954 | 7.77763005 | 5.61195685 | FALSE | TRUE | Concordant |
| kidney | CD27 | 0.12307179 | 0.12766715 | 5.90118075 | 5.64074723 | 5.61195685 | TRUE | TRUE | Concordant |
| kidney | CD300C | 0.12184315 | 0.07706656 | 5.858066 | 2.40607482 | 5.61195685 | TRUE | FALSE | Concordant |
| kidney | CD300E | 0.15855225 | 0.12204519 | 8.55729282 | 4.94203909 | 5.61195685 | TRUE | FALSE | Concordant |
| kidney | CD300LG | 0.18315146 | 0.13700472 | 12.0277463 | 6.60272363 | 5.61195685 | TRUE | TRUE | Concordant |
| kidney | CD302 | 0.15867038 | 0.18726193 | 8.38282413 | 10.9574335 | 5.61195685 | TRUE | TRUE | Concordant |
| kidney | CFC1 | 0.04903506 | 0.14477392 | 1.25633125 | 6.93776782 | 5.61195685 | FALSE | TRUE | Concordant |
| kidney | CHGB | 0.19718363 | 0.20864035 | 13.358905 | 13.9474788 | 5.61195685 | TRUE | TRUE | Concordant |
| kidney | CLEC14A | 0.16851729 | 0.16716904 | 10.1229871 | 9.5105706 | 5.61195685 | TRUE | TRUE | Concordant |
| kidney | CHRDL1 | 0.17539029 | 0.18664488 | 9.52088659 | 10.5823815 | 5.61195685 | TRUE | TRUE | Concordant |
| kidney | CKAP4 | 0.17098302 | 0.17191675 | 10.8208596 | 9.50798386 | 5.61195685 | TRUE | TRUE | Concordant |
| kidney | CDNF | 0.13902311 | 0.13524076 | 7.08485542 | 6.38231355 | 5.61195685 | TRUE | TRUE | Concordant |
| kidney | CDH3 | 0.15699168 | 0.14974498 | 9.08261778 | 7.88782619 | 5.61195685 | TRUE | TRUE | Concordant |
| kidney | CDSN | 0.11079658 | 0.14341916 | 4.88242508 | 7.24138001 | 5.61195685 | FALSE | TRUE | Concordant |
| kidney | FAM3C | 0.20975437 | 0.19816411 | 14.9640478 | 12.6772952 | 5.61195685 | TRUE | TRUE | Concordant |
| kidney | F3 | 0.11186499 | 0.14083324 | 4.35166573 | 6.69484214 | 5.61195685 | FALSE | TRUE | Concordant |
| kidney | FABP4 | 0.21280673 | 0.22300385 | 10.7817267 | 12.0060588 | 5.61195685 | TRUE | TRUE | Concordant |
| kidney | FAM3B | 0.13912275 | 0.11215936 | 7.21672837 | 4.56891933 | 5.61195685 | TRUE | FALSE | Concordant |
| kidney | GFRA1 | 0.15632679 | 0.18319263 | 7.83523768 | 10.9566924 | 5.61195685 | TRUE | TRUE | Concordant |
| kidney | FOLR1 | 0.13251397 | 0.11915595 | 6.48362737 | 4.90589239 | 5.61195685 | TRUE | FALSE | Concordant |
| kidney | FSTL3 | 0.25653744 | 0.22954446 | 19.8073027 | 14.6084759 | 5.61195685 | TRUE | TRUE | Concordant |
| kidney | DLL1 | 0.1772025 | 0.14557991 | 11.2618015 | 7.36086872 | 5.61195685 | TRUE | TRUE | Concordant |
| kidney | DSG3 | 0.11909035 | 0.16673528 | 5.47294356 | 9.43344253 | 5.61195685 | FALSE | TRUE | Concordant |
| kidney | DPT | 0.11243239 | 0.15630346 | 3.85807535 | 7.65595639 | 5.61195685 | FALSE | TRUE | Concordant |
| kidney | DSC2 | 0.16679018 | 0.14825314 | 9.94400765 | 7.33356761 | 5.61195685 | TRUE | TRUE | Concordant |
| kidney | DKK3 | 0.07976642 | 0.16304662 | 2.48942176 | 8.43473767 | 5.61195685 | FALSE | TRUE | Concordant |
| kidney | EPHA1 | 0.10508269 | 0.14405821 | 4.13875747 | 6.8548315 | 5.61195685 | FALSE | TRUE | Concordant |
| kidney | EPHA2 | 0.14685632 | 0.13236819 | 7.37938478 | 5.75402635 | 5.61195685 | TRUE | TRUE | Concordant |
| kidney | EPHB4 | 0.17682956 | 0.19805501 | 11.8240072 | 13.2796432 | 5.61195685 | TRUE | TRUE | Concordant |
| kidney | EPHB6 | 0.16285043 | 0.22221479 | 9.91026595 | 16.5578084 | 5.61195685 | TRUE | TRUE | Concordant |
| kidney | ESAM | 0.14080815 | 0.09556006 | 7.56629608 | 3.51084222 | 5.61195685 | TRUE | FALSE | Concordant |
| kidney | EDA2R | 0.23402753 | 0.15913767 | 13.7187755 | 5.87156499 | 5.61195685 | TRUE | TRUE | Concordant |
| kidney | EFEMP1 | 0.131188 | 0.1623308 | 6.05939657 | 7.78877505 | 5.61195685 | TRUE | TRUE | Concordant |

| | | | | | | | | | |
|---|---|---|---|---|---|---|---|---|---|
| kidney | EFNA1 | 0.17868611 | 0.17697025 | 11.1208136 | 10.3121304 | 5.61195685 | TRUE | TRUE | Concordant |
| kidney | EFNA4 | 0.19252485 | 0.17103784 | 12.2839619 | 9.43470609 | 5.61195685 | TRUE | TRUE | Concordant |
| kidney | DTX3 | 0.12030219 | 0.14274871 | 4.67939279 | 6.67714867 | 5.61195685 | FALSE | TRUE | Concordant |
| kidney | BSG | 0.13207811 | 0.16522147 | 6.34691446 | 9.35290632 | 5.61195685 | TRUE | TRUE | Concordant |
| kidney | BTN2A1 | 0.20301086 | 0.13950335 | 14.3909653 | 6.72806918 | 5.61195685 | TRUE | TRUE | Concordant |
| kidney | CA4 | 0.14328306 | 0.12902597 | 7.85566524 | 5.94486811 | 5.61195685 | TRUE | TRUE | Concordant |
| kidney | CA14 | 0.14124755 | 0.09829806 | 7.11087216 | 3.32821444 | 5.61195685 | TRUE | FALSE | Concordant |
| kidney | B4GAT1 | 0.06435177 | 0.1315338 | 1.76549948 | 5.93855693 | 5.61195685 | FALSE | TRUE | Concordant |
| kidney | BCAN | 0.07034224 | 0.17322289 | 2.1028063 | 9.4576389 | 5.61195685 | FALSE | TRUE | Concordant |
| kidney | CCL23 | 0.12840351 | 0.13101766 | 6.19342761 | 6.08329309 | 5.61195685 | TRUE | TRUE | Concordant |
| kidney | CCL27 | 0.05333548 | 0.14080393 | 1.28938969 | 6.85230974 | 5.61195685 | FALSE | TRUE | Concordant |
| kidney | CCN5 | 0.13393414 | 0.1188951 | 6.47424017 | 4.2686812 | 5.61195685 | TRUE | FALSE | Concordant |
| kidney | CALCA | 0.08851235 | 0.13093745 | 3.15360564 | 6.01456741 | 5.61195685 | FALSE | TRUE | Concordant |
| kidney | CCL14 | 0.17769787 | 0.10942744 | 11.7085465 | 4.35105792 | 5.61195685 | TRUE | FALSE | Concordant |
| kidney | ADM | 0.2041232 | 0.27344846 | 11.2869427 | 19.4765294 | 5.61195685 | TRUE | TRUE | Concordant |
| kidney | AGER | 0.12732271 | 0.09795135 | 5.9883809 | 3.45170227 | 5.61195685 | TRUE | FALSE | Concordant |
| kidney | AGRN | 0.1668 | 0.1360614 | 9.64258962 | 6.07479789 | 5.61195685 | TRUE | TRUE | Concordant |
| kidney | ACVRL1 | 0.19444028 | 0.17161075 | 12.5123221 | 9.34853754 | 5.61195685 | TRUE | TRUE | Concordant |
| kidney | ADAM22 | 0.15289756 | 0.14903293 | 8.73416159 | 7.66644581 | 5.61195685 | TRUE | TRUE | Concordant |
| kidney | AMBP | 0.14371864 | 0.15661837 | 6.71916411 | 8.00386579 | 5.61195685 | TRUE | TRUE | Concordant |
| kidney | ART3 | 0.20716181 | 0.22189193 | 15.2621784 | 15.8092126 | 5.61195685 | TRUE | TRUE | Concordant |
| kidney | ASGR1 | 0.15620384 | 0.0922602 | 7.9176941 | 2.94027676 | 5.61195685 | TRUE | FALSE | Concordant |
| kidney | AMY2A | 0.10566755 | 0.1568349 | 4.19523213 | 8.05547946 | 5.61195685 | FALSE | TRUE | Concordant |
| kidney | AMY2B | 0.10697643 | 0.16765454 | 4.23172641 | 8.75692852 | 5.61195685 | FALSE | TRUE | Concordant |
| kidney | ANG | 0.02916439 | 0.1343481 | 0.58018492 | 6.24721184 | 5.61195685 | FALSE | TRUE | Concordant |
| kidney | ANGPTL4 | 0.13566243 | 0.10161041 | 6.9312375 | 3.71847764 | 5.61195685 | TRUE | FALSE | Concordant |
| kidney | VGF | 0.21257704 | 0.18968304 | 13.9935321 | 10.1996696 | 5.61195685 | TRUE | TRUE | Concordant |
| kidney | WFDC2 | 0.14992636 | 0.22974144 | 7.54360246 | 13.5144538 | 5.61195685 | TRUE | TRUE | Concordant |
| kidney | VWC2L | 0.13617173 | 0.15116005 | 6.21568794 | 6.69437203 | 5.61195685 | TRUE | TRUE | Concordant |
| kidney | PTPRR | 0.20844095 | 0.22087957 | 13.4525144 | 13.2607669 | 5.61195685 | TRUE | TRUE | Concordant |
| kidney | PSAPL1 | 0.08853689 | 0.14064364 | 2.90551546 | 6.03784227 | 5.61195685 | FALSE | TRUE | Concordant |
| kidney | PRRT3 | 0.2518736 | 0.2326699 | 18.4821527 | 14.389342 | 5.61195685 | TRUE | TRUE | Concordant |
| kidney | YAP1 | 0.22198946 | 0.15054831 | 12.1869318 | 5.91932221 | 5.61195685 | TRUE | TRUE | Concordant |
| kidney | TGOLN2 | 0.13540078 | 0.11629126 | 6.15526011 | 4.2091861 | 5.61195685 | TRUE | FALSE | Concordant |
| kidney | SUSD4 | 0.07460968 | 0.13961275 | 2.19127921 | 5.91731324 | 5.61195685 | FALSE | TRUE | Concordant |

| | | | | | | | | | |
|---|---|---|---|---|---|---|---|---|---|
| kidney | SYT1 | 0.15087738 | 0.14254059 | 7.2452335 | 5.99950175 | 5.61195685 | TRUE | TRUE | Concordant |
| kidney | TNFRSF17 | 0.1251125 | 0.13767424 | 5.27256861 | 5.65744718 | 5.61195685 | FALSE | TRUE | Concordant |
| kidney | NPC2 | 0.17850346 | 0.1702677 | 9.2682635 | 7.47949496 | 5.61195685 | TRUE | TRUE | Concordant |
| kidney | NXPH3 | 0.14884001 | 0.11550042 | 6.56545109 | 3.99081839 | 5.61195685 | TRUE | FALSE | Concordant |
| kidney | NT5C1A | 0.14171199 | 0.11247349 | 5.68661013 | 3.72840485 | 5.61195685 | TRUE | FALSE | Concordant |
| kidney | PBXIP1 | 0.13707295 | 0.1301011 | 6.16927123 | 5.15303081 | 5.61195685 | TRUE | FALSE | Concordant |
| kidney | PALM2 | 0.14463115 | 0.07382684 | 5.83183256 | 1.77949214 | 5.61195685 | TRUE | FALSE | Concordant |
| kidney | PALM | 0.24099019 | 0.21246223 | 14.95801 | 11.7597684 | 5.61195685 | TRUE | TRUE | Concordant |
| kidney | PENK | 0.23151887 | 0.21710993 | 16.1461899 | 12.9317478 | 5.61195685 | TRUE | TRUE | Concordant |
| kidney | B2M | 0.15589614 | 0.15951154 | 7.03159591 | 7.17337633 | 5.61195685 | TRUE | TRUE | Concordant |
| kidney | ASGR2 | 0.13848471 | 0.09164508 | 6.35599117 | 2.82000879 | 5.61195685 | TRUE | FALSE | Concordant |
| kidney | AMY1A_AMY1B_AMY1C | 0.10108066 | 0.15469904 | 3.3028868 | 6.62200105 | 5.61195685 | FALSE | TRUE | Concordant |
| kidney | CD300A | 0.15089669 | 0.06305275 | 7.32366799 | 1.49190941 | 5.61195685 | TRUE | FALSE | Concordant |
| kidney | CEACAM19 | 0.12028199 | 0.1605172 | 4.87402562 | 7.59638019 | 5.61195685 | FALSE | TRUE | Concordant |
| kidney | CALCB | 0.17792404 | 0.15508996 | 10.0457475 | 7.05207827 | 5.61195685 | TRUE | TRUE | Concordant |
| kidney | CCER2 | 0.12874772 | 0.15828459 | 5.42337785 | 7.26341403 | 5.61195685 | FALSE | TRUE | Concordant |
| kidney | SHISA5 | 0.23620551 | 0.21677413 | 15.3361458 | 11.3791062 | 5.61195685 | TRUE | TRUE | Concordant |
| kidney | SERPINF1 | 0.21816004 | 0.15292691 | 12.0081199 | 6.43101341 | 5.61195685 | TRUE | TRUE | Concordant |
| kidney | SPRR3 | 0.13158327 | 0.09205779 | 5.73282548 | 2.82961154 | 5.61195685 | TRUE | FALSE | Concordant |
| kidney | SLURP1 | 0.10398924 | 0.14083899 | 3.69460529 | 5.83770105 | 5.61195685 | FALSE | TRUE | Concordant |
| kidney | SLITRK1 | 0.18046465 | 0.13507466 | 9.52122369 | 5.04345177 | 5.61195685 | TRUE | FALSE | Concordant |
| kidney | SCRG1 | 0.22176131 | 0.17061573 | 12.3054425 | 7.64657972 | 5.61195685 | TRUE | TRUE | Concordant |
| kidney | RBFOX3 | 0.16428692 | 0.10955092 | 7.92844085 | 3.48385954 | 5.61195685 | TRUE | FALSE | Concordant |
| kidney | AHNAK | 0.15676831 | 0.12134877 | 6.65180897 | 4.17521584 | 5.61195685 | TRUE | FALSE | Concordant |
| kidney | SCN4B | 0.19706528 | 0.17443857 | 11.7425811 | 8.71226815 | 5.61195685 | TRUE | TRUE | Concordant |
| kidney | SBSN | 0.14701663 | 0.12237122 | 6.97993101 | 4.6616033 | 5.61195685 | TRUE | FALSE | Concordant |
| kidney | RNASE1 | 0.18715625 | 0.19896995 | 9.68130618 | 9.75146858 | 5.61195685 | TRUE | TRUE | Concordant |
| kidney | RNASE4 | 0.17433804 | 0.19213849 | 8.13262765 | 9.56228162 | 5.61195685 | TRUE | TRUE | Concordant |
| kidney | RNASE6 | 0.16819068 | 0.16941557 | 8.29674576 | 7.75029161 | 5.61195685 | TRUE | TRUE | Concordant |
| kidney | RNF149 | 0.16652004 | 0.14720186 | 8.56743588 | 6.24191602 | 5.61195685 | TRUE | TRUE | Concordant |
| kidney | FABP3 | 0.16956942 | 0.07861631 | 8.19390133 | 1.94898261 | 5.61195685 | TRUE | FALSE | Concordant |
| kidney | FBLN2 | 0.14149521 | 0.0994742 | 6.42898343 | 3.18315265 | 5.61195685 | TRUE | FALSE | Concordant |
| kidney | EDN1 | 0.19803859 | 0.1062085 | 11.4784656 | 3.34967098 | 5.61195685 | TRUE | FALSE | Concordant |
| kidney | EFCAB14 | 0.18601599 | 0.16988236 | 10.3437005 | 8.26360546 | 5.61195685 | TRUE | TRUE | Concordant |

| | | | | | | | | | |
|---|---|---|---|---|---|---|---|---|---|
| kidney | EFHD1 | 0.14987593 | 0.14346955 | 6.85985565 | 5.80723886 | 5.61195685 | TRUE | TRUE | Concordant |
| kidney | EPHA4 | 0.1644741 | 0.15427696 | 8.79046457 | 7.00922455 | 5.61195685 | TRUE | TRUE | Concordant |
| kidney | ELN | 0.19252804 | 0.0816757 | 6.70694785 | 1.69572785 | 5.61195685 | TRUE | FALSE | Concordant |
| kidney | GPR158 | 0.14689125 | 0.11566454 | 6.58937383 | 3.77219377 | 5.61195685 | TRUE | FALSE | Concordant |
| kidney | GM2A | 0.16621525 | 0.19716359 | 8.38544003 | 10.2481392 | 5.61195685 | TRUE | TRUE | Concordant |
| kidney | COL15A1 | 0.27581347 | 0.21624404 | 20.5904401 | 12.6210796 | 5.61195685 | TRUE | TRUE | Concordant |
| kidney | CPXM2 | 0.16339062 | 0.05214252 | 8.18815181 | 1.10416362 | 5.61195685 | TRUE | FALSE | Concordant |
| kidney | CPA4 | 0.07905121 | 0.15836052 | 2.33334734 | 7.17068444 | 5.61195685 | FALSE | TRUE | Concordant |
| kidney | CFD | 0.27406339 | 0.17148135 | 20.3783343 | 8.01144071 | 5.61195685 | TRUE | TRUE | Concordant |
| kidney | CELSR2 | 0.16392006 | 0.19179051 | 8.61190152 | 10.3464791 | 5.61195685 | TRUE | TRUE | Concordant |
| kidney | CHCHD10 | 0.16935991 | 0.08498272 | 8.26468379 | 2.36012072 | 5.61195685 | TRUE | FALSE | Concordant |
| kidney | CYTL1 | 0.20949986 | 0.15034388 | 13.7782172 | 6.70283075 | 5.61195685 | TRUE | TRUE | Concordant |
| kidney | KIAA0319 | 0.16691121 | 0.18650202 | 8.58914071 | 9.77706604 | 5.61195685 | TRUE | TRUE | Concordant |
| kidney | HS6ST2 | 0.16396656 | 0.16176204 | 8.54327644 | 7.52983996 | 5.61195685 | TRUE | TRUE | Concordant |
| kidney | GRP | 0.13512654 | 0.14916812 | 5.80456119 | 6.54758053 | 5.61195685 | TRUE | TRUE | Concordant |
| kidney | LMOD1 | 0.13637739 | 0.16845832 | 5.81195977 | 7.52416298 | 5.61195685 | TRUE | TRUE | Concordant |
| liver | PRCP | 0.13972711 | 0.15267648 | 5.07368549 | 5.80755985 | 5.61195685 | FALSE | TRUE | Concordant |
| liver | PLAT | 0.1799673 | 0.19423598 | 7.37038764 | 8.24562866 | 5.61195685 | TRUE | TRUE | Concordant |
| liver | IL1RN | 0.18699147 | 0.08604236 | 7.9086203 | 1.98946867 | 5.61195685 | TRUE | FALSE | Concordant |
| liver | GUSB | 0.18295737 | 0.09649928 | 8.41576817 | 2.5559757 | 5.61195685 | TRUE | FALSE | Concordant |
| liver | GPNMB | 0.16073547 | 0.01996229 | 6.97544671 | 0.28400545 | 5.61195685 | TRUE | FALSE | Concordant |
| liver | NADK | 0.15785127 | 0.08896896 | 6.85211596 | 2.39643263 | 5.61195685 | TRUE | FALSE | Concordant |
| liver | NCAN | 0.21406038 | 0.16746164 | 11.6075258 | 7.24169986 | 5.61195685 | TRUE | TRUE | Concordant |
| liver | MIA | 0.14042217 | 0.0695502 | 5.76690882 | 1.65348075 | 5.61195685 | TRUE | FALSE | Concordant |
| liver | NPPC | 0.1784032 | 0.13715162 | 8.60681259 | 5.20651498 | 5.61195685 | TRUE | FALSE | Concordant |
| liver | NRCAM | 0.16452872 | 0.13493121 | 7.54833083 | 5.03387752 | 5.61195685 | TRUE | FALSE | Concordant |
| liver | NPDC1 | 0.18707389 | 0.11178324 | 8.88158657 | 3.51735568 | 5.61195685 | TRUE | FALSE | Concordant |
| liver | NELL2 | 0.1736247 | 0.1901349 | 8.26665313 | 9.55124047 | 5.61195685 | TRUE | TRUE | Concordant |
| liver | LEP | 0.21758552 | 0.24718283 | 6.88632558 | 10.040464 | 5.61195685 | TRUE | TRUE | Concordant |
| liver | SEZ6L | 0.20178546 | 0.15985719 | 10.3423868 | 6.65795795 | 5.61195685 | TRUE | TRUE | Concordant |
| liver | SCG3 | 0.16840505 | 0.08864408 | 7.67746388 | 2.3493268 | 5.61195685 | TRUE | FALSE | Concordant |
| liver | SDC1 | 0.14833314 | 0.11874416 | 6.32663607 | 4.05748086 | 5.61195685 | TRUE | FALSE | Concordant |
| liver | RTBDN | 0.12032347 | 0.14499029 | 4.09255082 | 5.62747109 | 5.61195685 | FALSE | TRUE | Concordant |
| liver | CDHR2 | 0.15994024 | 0.17890875 | 5.48739725 | 7.17370987 | 5.61195685 | FALSE | TRUE | Concordant |
| liver | FABP4 | 0.2005396 | 0.15476167 | 7.15744985 | 4.77620249 | 5.61195685 | TRUE | FALSE | Concordant |

| | | | | | | | | | |
|---|---|---|---|---|---|---|---|---|---|
| liver | DPP6 | 0.08926838 | 0.1452849 | 2.42490595 | 5.64083183 | 5.61195685 | FALSE | TRUE | Concordant |
| liver | DCXR | 0.12151678 | 0.14903375 | 4.03847559 | 5.75600062 | 5.61195685 | FALSE | TRUE | Concordant |
| liver | ERBB2 | 0.15464523 | 0.14878582 | 6.14471926 | 5.54616974 | 5.61195685 | TRUE | FALSE | Concordant |
| liver | ENPP2 | 0.01484861 | 0.14962164 | 0.20828392 | 6.12829803 | 5.61195685 | FALSE | TRUE | Concordant |
| liver | B4GAT1 | 0.16846593 | 0.03549196 | 7.05421412 | 0.59840854 | 5.61195685 | TRUE | FALSE | Concordant |
| liver | AGRN | 0.15072166 | 0.08919951 | 6.07002626 | 2.28117314 | 5.61195685 | TRUE | FALSE | Concordant |
| liver | ACP5 | 0.13492654 | 0.19408854 | 4.76346915 | 9.65365845 | 5.61195685 | FALSE | TRUE | Concordant |
| liver | ARSA | 0.14307597 | 0.11537914 | 5.81184393 | 3.66912757 | 5.61195685 | TRUE | FALSE | Concordant |
| liver | VSNL1 | 0.18412082 | 0.12410175 | 8.10242972 | 3.73659501 | 5.61195685 | TRUE | FALSE | Concordant |
| liver | VWC2L | 0.09540867 | 0.16058773 | 2.59418411 | 5.98850812 | 5.61195685 | FALSE | TRUE | Concordant |
| liver | MYL3 | -0.1617001 | -0.0831 | 5.89133389 | 1.82049729 | 5.61195685 | TRUE | FALSE | Concordant |
| liver | MYOM3 | -0.1594869 | -0.1009689 | 5.91170418 | 2.61487761 | 5.61195685 | TRUE | FALSE | Concordant |
| liver | AFM | 0.16028337 | 0.14717904 | 5.70746398 | 4.49629167 | 5.61195685 | TRUE | FALSE | Concordant |
| liver | GPD1 | 0.17305681 | 0.07804466 | 6.3863202 | 1.50981071 | 5.61195685 | TRUE | FALSE | Concordant |
| liver | FUOM | 0.16526283 | 0.13632947 | 5.97263505 | 4.04951039 | 5.61195685 | TRUE | FALSE | Concordant |
| liver | CFH | 0.20244983 | 0.10592988 | 7.06945562 | 2.26212961 | 5.61195685 | TRUE | FALSE | Concordant |
| liver | CELSR2 | 0.15718276 | 0.09463165 | 6.15992115 | 2.35990157 | 5.61195685 | TRUE | FALSE | Concordant |
| liver | IGSF9 | 0.18106559 | 0.14535024 | 6.45509959 | 3.95326697 | 5.61195685 | TRUE | FALSE | Concordant |
| liver | LRTM2 | 0.11525245 | 0.16518448 | 3.40610297 | 6.03753216 | 5.61195685 | FALSE | TRUE | Concordant |
| pancreas | PLA2G1B | -0.3533744 | -0.3975538 | 34.5754925 | 42.8132445 | 5.61195685 | TRUE | TRUE | Concordant |
| pancreas | PRSS2 | -0.2341719 | -0.2705804 | 15.4699582 | 20.4772265 | 5.61195685 | TRUE | TRUE | Concordant |
| pancreas | KIRREL2 | -0.1639084 | -0.1932673 | 7.68553049 | 10.1734507 | 5.61195685 | TRUE | TRUE | Concordant |
| pancreas | GP2 | -0.1462892 | -0.1016404 | 6.13384109 | 3.17616187 | 5.61195685 | TRUE | FALSE | Concordant |
| pancreas | LGMN | 0.14013427 | 0.02488252 | 6.07412069 | 0.39898617 | 5.61195685 | TRUE | FALSE | Concordant |
| pancreas | CPA1 | -0.2147835 | -0.2292154 | 13.4432158 | 14.9312676 | 5.61195685 | TRUE | TRUE | Concordant |
| pancreas | CPB1 | -0.2147901 | -0.2337532 | 13.4714212 | 15.491057 | 5.61195685 | TRUE | TRUE | Concordant |
| pancreas | CLPS | -0.2101694 | -0.2360438 | 12.5936171 | 15.5208812 | 5.61195685 | TRUE | TRUE | Concordant |
| pancreas | CTRC | -0.2186701 | -0.2666684 | 13.8371833 | 19.6467791 | 5.61195685 | TRUE | TRUE | Concordant |
| pancreas | CTRB1 | -0.2392862 | -0.2725035 | 16.3604007 | 20.7152098 | 5.61195685 | TRUE | TRUE | Concordant |
| pancreas | CELA3A | -0.2515168 | -0.24831 | 17.7766253 | 17.2665978 | 5.61195685 | TRUE | TRUE | Concordant |
| pancreas | AMY2A | -0.1528421 | -0.1743472 | 6.50592844 | 8.21348028 | 5.61195685 | TRUE | TRUE | Concordant |
| pancreas | AMY2B | -0.1592622 | -0.1796362 | 6.86387974 | 8.35479652 | 5.61195685 | TRUE | TRUE | Concordant |
| pancreas | OCLN | 0.15292161 | 0.00796487 | 5.72899732 | 0.09249086 | 5.61195685 | TRUE | FALSE | Concordant |
| pancreas | PNLIP | -0.2483207 | -0.2113542 | 15.329272 | 10.9141363 | 5.61195685 | TRUE | TRUE | Concordant |
| pancreas | PNLIPRP1 | -0.1585742 | -0.1877223 | 6.65348063 | 8.64180039 | 5.61195685 | TRUE | TRUE | Concordant |

| | | | | | | | | | |
|---|---|---|---|---|---|---|---|---|---|
| pancreas | SEMA3G | -0.2090778 | -0.2011166 | 10.7945047 | 9.77728398 | 5.61195685 | TRUE | TRUE | Concordant |
| pancreas | CELA2A | -0.2999522 | -0.3182386 | 22.3351613 | 24.4157361 | 5.61195685 | TRUE | TRUE | Concordant |
| pancreas | CTRL | -0.2298464 | -0.2628571 | 12.8577078 | 16.1966443 | 5.61195685 | TRUE | TRUE | Concordant |
| spleen | PDCD1 | -0.0841383 | -0.1643057 | 2.37793442 | 7.41544274 | 5.61195685 | FALSE | TRUE | Concordant |
| spleen | PIK3IP1 | -0.0921294 | -0.1688138 | 2.51932368 | 7.21920789 | 5.61195685 | FALSE | TRUE | Concordant |
| spleen | PCDH17 | -0.1445217 | -0.1266705 | 6.17823172 | 4.64077102 | 5.61195685 | TRUE | FALSE | Concordant |
| spleen | IL18BP | -0.1773329 | -0.2750671 | 8.29555967 | 19.3419526 | 5.61195685 | TRUE | TRUE | Concordant |
| spleen | KLRB1 | -0.2033982 | -0.2202066 | 11.6497523 | 12.9902893 | 5.61195685 | TRUE | TRUE | Concordant |
| spleen | KLRD1 | -0.1027898 | -0.1428652 | 3.14686504 | 5.63804575 | 5.61195685 | FALSE | TRUE | Concordant |
| spleen | KAZALD1 | -0.2070881 | -0.2251867 | 12.1737013 | 13.264548 | 5.61195685 | TRUE | TRUE | Concordant |
| spleen | HAVCR2 | -0.0966837 | -0.1594676 | 2.92318561 | 6.53158225 | 5.61195685 | FALSE | TRUE | Concordant |
| spleen | HLA-E | -0.0939632 | -0.1576368 | 2.72004448 | 6.78093543 | 5.61195685 | FALSE | TRUE | Concordant |
| spleen | IL12B | -0.1303777 | -0.1627832 | 5.16883171 | 7.26770868 | 5.61195685 | FALSE | TRUE | Concordant |
| spleen | IL12A_IL12B | -0.1392435 | -0.1727137 | 5.7269544 | 7.94392282 | 5.61195685 | TRUE | TRUE | Concordant |
| spleen | ICAM2 | -0.1062108 | -0.1644642 | 3.62191953 | 7.38078033 | 5.61195685 | FALSE | TRUE | Concordant |
| spleen | MFGE8 | 0.14102971 | 0.0812056 | 5.78030092 | 2.14324551 | 5.61195685 | TRUE | FALSE | Concordant |
| spleen | LAG3 | -0.1136975 | -0.1578595 | 3.83259472 | 6.76626098 | 5.61195685 | FALSE | TRUE | Concordant |
| spleen | LAIR1 | -0.0827578 | -0.1573885 | 2.08198902 | 6.28376242 | 5.61195685 | FALSE | TRUE | Concordant |
| spleen | LCN2 | -0.0527787 | -0.1521262 | 1.09626966 | 6.01414934 | 5.61195685 | FALSE | TRUE | Concordant |
| spleen | LTA | -0.0932107 | -0.1902193 | 2.84693384 | 9.85592747 | 5.61195685 | FALSE | TRUE | Concordant |
| spleen | SIGLEC10 | -0.1801655 | -0.2573304 | 9.13085022 | 17.0961512 | 5.61195685 | TRUE | TRUE | Concordant |
| spleen | SIGLEC6 | -0.1062055 | -0.1455758 | 3.64555501 | 6.04709665 | 5.61195685 | FALSE | TRUE | Concordant |
| spleen | SIRPB1 | -0.0867946 | -0.1603359 | 2.52055592 | 6.99805213 | 5.61195685 | FALSE | TRUE | Concordant |
| spleen | SLITRK2 | -0.1442892 | -0.2470465 | 6.10362829 | 15.6964001 | 5.61195685 | TRUE | TRUE | Concordant |
| spleen | SEMA7A | -0.1728749 | -0.2688508 | 8.61456842 | 19.0428442 | 5.61195685 | TRUE | TRUE | Concordant |
| spleen | STC2 | -0.0573357 | -0.2342588 | 1.21341652 | 13.715172 | 5.61195685 | FALSE | TRUE | Concordant |
| spleen | RELT | -0.0325643 | -0.1437345 | 0.5320354 | 5.68646008 | 5.61195685 | FALSE | TRUE | Concordant |
| spleen | PTX3 | -0.066368 | -0.1405232 | 1.6107218 | 5.6158991 | 5.61195685 | FALSE | TRUE | Concordant |
| spleen | SDC4 | 0.16077714 | 0.11088122 | 7.48205899 | 3.63052913 | 5.61195685 | TRUE | FALSE | Concordant |
| spleen | SEMA3F | -0.1378975 | -0.2411206 | 4.89230247 | 14.5137353 | 5.61195685 | FALSE | TRUE | Concordant |
| spleen | RNASET2 | -0.0409957 | -0.1497348 | 0.77573536 | 6.10535691 | 5.61195685 | FALSE | TRUE | Concordant |
| spleen | WFIKKN1 | -0.175382 | -0.2500056 | 8.64657444 | 15.9235904 | 5.61195685 | TRUE | TRUE | Concordant |
| spleen | VCAM1 | -0.2696011 | -0.3605629 | 19.7035996 | 34.1122643 | 5.61195685 | TRUE | TRUE | Concordant |
| spleen | TFRC | -0.1408233 | -0.1538394 | 5.96049252 | 6.55623953 | 5.61195685 | TRUE | TRUE | Concordant |
| spleen | TFF1 | -0.0668328 | -0.1535438 | 1.59190622 | 6.14867821 | 5.61195685 | FALSE | TRUE | Concordant |

| | | | | | | | | | |
|---|---|---|---|---|---|---|---|---|---|
| spleen | TREML2 | -0.1525216 | -0.2536249 | 6.63377021 | 16.9558913 | 5.61195685 | TRUE | TRUE | Concordant |
| spleen | TNFRSF9 | -0.1283603 | -0.2319165 | 4.52530123 | 13.9931979 | 5.61195685 | FALSE | TRUE | Concordant |
| spleen | TNF | -0.0455301 | -0.1549954 | 0.87637825 | 6.53354518 | 5.61195685 | FALSE | TRUE | Concordant |
| spleen | TNFRSF13C | -0.0702959 | -0.15057 | 1.73377554 | 6.15882565 | 5.61195685 | FALSE | TRUE | Concordant |
| spleen | TNFRSF1A | -0.0588674 | -0.1684518 | 1.19645512 | 6.99156967 | 5.61195685 | FALSE | TRUE | Concordant |
| spleen | TNFRSF1B | -0.1197926 | -0.2218631 | 4.13501371 | 12.1871986 | 5.61195685 | FALSE | TRUE | Concordant |
| spleen | TNFRSF4 | -0.1339797 | -0.2277209 | 5.25596989 | 13.4520499 | 5.61195685 | FALSE | TRUE | Concordant |
| spleen | CR2 | -0.1535368 | -0.224653 | 6.74645012 | 12.5057488 | 5.61195685 | TRUE | TRUE | Concordant |
| spleen | CLEC7A | -0.0441842 | -0.1428686 | 0.8609199 | 5.69603827 | 5.61195685 | FALSE | TRUE | Concordant |
| spleen | CTSL | -0.1013064 | -0.1894696 | 3.08933821 | 9.59914072 | 5.61195685 | FALSE | TRUE | Concordant |
| spleen | CXCL6 | -0.0239801 | -0.1460827 | 0.37929524 | 6.01198542 | 5.61195685 | FALSE | TRUE | Concordant |
| spleen | CRTAM | -0.1170418 | -0.1865088 | 4.03639728 | 9.33290386 | 5.61195685 | FALSE | TRUE | Concordant |
| spleen | CRLF1 | -0.0882473 | -0.2324504 | 2.38039286 | 13.3479422 | 5.61195685 | FALSE | TRUE | Concordant |
| spleen | CSF1 | -0.1098489 | -0.1638634 | 3.61303429 | 7.04389861 | 5.61195685 | FALSE | TRUE | Concordant |
| spleen | CD4 | -0.0652044 | -0.1428513 | 1.47410006 | 5.63509772 | 5.61195685 | FALSE | TRUE | Concordant |
| spleen | CD48 | -0.1323179 | -0.1802023 | 5.15017309 | 8.87130444 | 5.61195685 | FALSE | TRUE | Concordant |
| spleen | CD5 | -0.0545002 | -0.1583312 | 1.17286841 | 6.94721822 | 5.61195685 | FALSE | TRUE | Concordant |
| spleen | CD74 | -0.1152033 | -0.1902711 | 3.8884831 | 9.2655798 | 5.61195685 | FALSE | TRUE | Concordant |
| spleen | CD79B | -0.1464723 | -0.1954385 | 6.22847374 | 10.2711113 | 5.61195685 | TRUE | TRUE | Concordant |
| spleen | CD83 | -0.1033553 | -0.1494977 | 3.21808057 | 6.1031213 | 5.61195685 | FALSE | TRUE | Concordant |
| spleen | CD8A | -0.1124765 | -0.2285257 | 3.82930572 | 13.4435328 | 5.61195685 | FALSE | TRUE | Concordant |
| spleen | CD244 | -0.1470447 | -0.1635212 | 6.43344448 | 7.28327494 | 5.61195685 | TRUE | TRUE | Concordant |
| spleen | CD160 | -0.164806 | -0.2184222 | 7.90127468 | 12.5794439 | 5.61195685 | TRUE | TRUE | Concordant |
| spleen | CD200R1 | -0.0950819 | -0.1565368 | 2.92098321 | 6.67642472 | 5.61195685 | FALSE | TRUE | Concordant |
| spleen | CD22 | -0.1243348 | -0.1829165 | 4.49427844 | 9.09318789 | 5.61195685 | FALSE | TRUE | Concordant |
| spleen | CD300C | -0.0605785 | -0.1707523 | 1.43481409 | 7.90894414 | 5.61195685 | FALSE | TRUE | Concordant |
| spleen | CDH3 | -0.1008914 | -0.2446307 | 3.23316215 | 16.1492643 | 5.61195685 | FALSE | TRUE | Concordant |
| spleen | FGF23 | -0.1373642 | -0.1276231 | 5.66592854 | 4.63188912 | 5.61195685 | TRUE | FALSE | Concordant |
| spleen | FCRL3 | -0.0399355 | -0.2062361 | 0.67238744 | 9.77488371 | 5.61195685 | FALSE | TRUE | Concordant |
| spleen | FCRL2 | -0.1409998 | -0.1796473 | 5.77470721 | 8.7923752 | 5.61195685 | TRUE | TRUE | Concordant |
| spleen | FCRL1 | -0.1843582 | -0.2382916 | 9.47569966 | 15.1180015 | 5.61195685 | TRUE | TRUE | Concordant |
| spleen | FASLG | -0.1536037 | -0.1279248 | 6.88072963 | 4.40096595 | 5.61195685 | TRUE | FALSE | Concordant |
| spleen | FCER2 | -0.144276 | -0.2274679 | 5.80903755 | 13.6112995 | 5.61195685 | TRUE | TRUE | Concordant |
| spleen | GFRA2 | -0.0852179 | -0.1501277 | 2.46308018 | 6.27242233 | 5.61195685 | FALSE | TRUE | Concordant |
| spleen | GCNT1 | -0.1522829 | -0.2421465 | 6.57396754 | 15.1888968 | 5.61195685 | TRUE | TRUE | Concordant |

| | | | | | | | | | |
|---|---|---|---|---|---|---|---|---|---|
| spleen | FRZB | -0.059497 | -0.1602477 | 1.35669716 | 7.07474333 | 5.61195685 | FALSE | TRUE | Concordant |
| spleen | GALNT3 | -0.1268124 | -0.2230708 | 4.76679518 | 12.582992 | 5.61195685 | FALSE | TRUE | Concordant |
| spleen | GALNT2 | -0.0711423 | -0.1602402 | 1.82341529 | 7.16334456 | 5.61195685 | FALSE | TRUE | Concordant |
| spleen | FLT3LG | 0.12064458 | 0.1541284 | 4.0659492 | 6.44210909 | 5.61195685 | FALSE | TRUE | Concordant |
| spleen | DLL1 | -0.1017543 | -0.1820331 | 3.19569912 | 8.93747649 | 5.61195685 | FALSE | TRUE | Concordant |
| spleen | DSC2 | -0.1155809 | -0.225694 | 3.97750177 | 13.0538467 | 5.61195685 | FALSE | TRUE | Concordant |
| spleen | DEFA1_DEFA1B | -0.0683335 | -0.1671373 | 1.65406831 | 7.2955404 | 5.61195685 | FALSE | TRUE | Concordant |
| spleen | EBI3_IL27 | -0.131609 | -0.1901731 | 5.19186777 | 9.37328603 | 5.61195685 | FALSE | TRUE | Concordant |
| spleen | EFNA4 | -0.052 | -0.1976456 | 1.03010947 | 9.99297379 | 5.61195685 | FALSE | TRUE | Concordant |
| spleen | BTN2A1 | -0.1088547 | -0.1582036 | 3.55332243 | 6.86269719 | 5.61195685 | FALSE | TRUE | Concordant |
| spleen | C1QA | -0.1931533 | -0.2402766 | 10.5063949 | 15.1374725 | 5.61195685 | TRUE | TRUE | Concordant |
| spleen | AXL | -0.1642973 | -0.2238822 | 7.34197388 | 13.3500913 | 5.61195685 | TRUE | TRUE | Concordant |
| spleen | ADGRE5 | -0.1651476 | -0.158845 | 7.42382208 | 6.77844844 | 5.61195685 | TRUE | TRUE | Concordant |
| spleen | ADAM8 | -0.0573681 | -0.2083048 | 1.28276371 | 11.6050052 | 5.61195685 | FALSE | TRUE | Concordant |
| spleen | ADGRE2 | -0.1387304 | -0.1702657 | 5.40830027 | 7.67810098 | 5.61195685 | FALSE | TRUE | Concordant |
| spleen | PTPRC | -0.0946997 | -0.1588253 | 2.4107791 | 5.72343564 | 5.61195685 | FALSE | TRUE | Concordant |
| spleen | PTPRH | -0.2256563 | -0.3237677 | 12.2673589 | 22.8911865 | 5.61195685 | TRUE | TRUE | Concordant |
| spleen | TCOF1 | -0.0564184 | -0.1745966 | 0.99987712 | 6.31498788 | 5.61195685 | FALSE | TRUE | Concordant |
| spleen | NCR3LG1 | -0.1104213 | -0.1675485 | 3.2237333 | 6.33401142 | 5.61195685 | FALSE | TRUE | Concordant |
| spleen | CD72 | -0.1770245 | -0.2626118 | 7.14707398 | 14.6002239 | 5.61195685 | TRUE | TRUE | Concordant |
| spleen | CD80 | -0.1436503 | -0.1559053 | 4.99626233 | 5.69889457 | 5.61195685 | FALSE | TRUE | Concordant |
| spleen | CD5L | -0.0885899 | -0.155385 | 2.30272788 | 5.69532443 | 5.61195685 | FALSE | TRUE | Concordant |
| spleen | SIGLEC8 | -0.0871953 | -0.2017991 | 2.13461456 | 8.68236683 | 5.61195685 | FALSE | TRUE | Concordant |
| spleen | SELL | -0.1258129 | -0.1551378 | 4.14326159 | 5.64376766 | 5.61195685 | FALSE | TRUE | Concordant |
| spleen | SDK2 | -0.0927442 | -0.1695315 | 2.33203677 | 6.58062138 | 5.61195685 | FALSE | TRUE | Concordant |
| spleen | ADGRE1 | -0.1837285 | -0.1706356 | 8.33509275 | 6.70060335 | 5.61195685 | TRUE | TRUE | Concordant |
| spleen | COCH | 0.15730285 | 0.07039701 | 6.08815203 | 1.43141379 | 5.61195685 | TRUE | FALSE | Concordant |
| spleen | KLRK1 | -0.101526 | -0.1801518 | 2.88282861 | 7.62221213 | 5.61195685 | FALSE | TRUE | Concordant |
| spleen | MAMDC2 | -0.0799131 | -0.1590905 | 1.8577805 | 5.70442897 | 5.61195685 | FALSE | TRUE | Concordant |

#### eTable 11: Discordant hits for the sex-stratified MetWAS analyses

| MRIBAG | Metabolite | Female | Male | logP_female | logP_male | P-thres | is_sig_female | is_sig_male | Direction |
|---|---|---|---|---|---|---|---|---|---|
| adipose | S_VLDL_CE_pct | 0.00316858 | -0.0549583 | 0.11229255 | 5.18382872 | 4.66200188 | FALSE | TRUE | Discordant |
| heart | HDL_size | -0.0538197 | 0.00339254 | 7.26041873 | 0.13400566 | 4.66200188 | TRUE | FALSE | Discordant |
| heart | L_HDL_CE | -0.0512198 | 0.00414015 | 6.6632602 | 0.16730746 | 4.66200188 | TRUE | FALSE | Discordant |
| heart | L_HDL_P | -0.044393 | 0.00946454 | 5.17784663 | 0.46463506 | 4.66200188 | TRUE | FALSE | Discordant |
| heart | XL_HDL_P | -0.0471436 | 0.00108435 | 5.81218802 | 0.03986185 | 4.66200188 | TRUE | FALSE | Discordant |
| heart | L_HDL_FC | -0.0455897 | 0.00705694 | 5.47784984 | 0.32132292 | 4.66200188 | TRUE | FALSE | Discordant |
| heart | L_HDL_C | -0.0499459 | 0.00478055 | 6.38213226 | 0.19832071 | 4.66200188 | TRUE | FALSE | Discordant |
| heart | L_HDL_L | -0.0422024 | 0.01046135 | 4.73424813 | 0.53155814 | 4.66200188 | TRUE | FALSE | Discordant |
| heart | XXL_VLDL_PL_pct | 0.04231368 | -0.0003035 | 5.27031838 | 0.01137948 | 4.66200188 | TRUE | FALSE | Discordant |
| heart | L_LDL_FC_pct | -0.0605895 | 0.00237835 | 9.44558961 | 0.09192672 | 4.66200188 | TRUE | FALSE | Discordant |
| heart | IDL_TG_pct | 0.05009348 | -0.001071 | 6.84305813 | 0.03940502 | 4.66200188 | TRUE | FALSE | Discordant |
| heart | XL_HDL_FC_pct | 0.04387245 | -0.0083926 | 4.96504061 | 0.40295601 | 4.66200188 | TRUE | FALSE | Discordant |
| heart | S_VLDL_FC_pct_C | -0.0419487 | 0.00638715 | 4.93355639 | 0.28808104 | 4.66200188 | TRUE | FALSE | Discordant |
| heart | XS_VLDL_FC_pct_C | 0.05397505 | -0.0061008 | 7.62390234 | 0.27075557 | 4.66200188 | TRUE | FALSE | Discordant |
| heart | L_LDL_FC_pct_C | -0.0557573 | 0.00226058 | 8.31902473 | 0.08819121 | 4.66200188 | TRUE | FALSE | Discordant |
| heart | XL_HDL_FC_pct_C | 0.04806246 | -0.0085977 | 5.91298896 | 0.40936942 | 4.66200188 | TRUE | FALSE | Discordant |
| heart | S_VLDL_CE_pct_C | 0.04194873 | -0.0063871 | 4.93355639 | 0.28808104 | 4.66200188 | TRUE | FALSE | Discordant |
| heart | XS_VLDL_CE_pct_C | -0.053975 | 0.0061008 | 7.62390234 | 0.27075557 | 4.66200188 | TRUE | FALSE | Discordant |
| heart | L_LDL_CE_pct_C | 0.05575726 | -0.0022606 | 8.31902473 | 0.08819121 | 4.66200188 | TRUE | FALSE | Discordant |
| heart | XL_HDL_CE_pct_C | -0.0480625 | 0.00859774 | 5.91298896 | 0.40936942 | 4.66200188 | TRUE | FALSE | Discordant |
| heart | XS_VLDL_FC_by_CE | 0.05229915 | -0.0059549 | 7.20021402 | 0.26454966 | 4.66200188 | TRUE | FALSE | Discordant |
| heart | L_LDL_FC_by_CE | -0.0559464 | 0.00285932 | 8.37530056 | 0.11399084 | 4.66200188 | TRUE | FALSE | Discordant |
| kidney | S_LDL_C_pct | -0.0159419 | 0.05636523 | 1.05808221 | 7.93350869 | 4.66200188 | FALSE | TRUE | Discordant |

| | | | | | | | | | |
|---|---|---|---|---|---|---|---|---|---|
| kidney | S_HDL_CE_pct | -0.0008721 | 0.04217683 | 0.03308757 | 4.75384754 | 4.66200188 | FALSE | TRUE | Discordant |
| kidney | S_HDL_PL_pct | 0.00238169 | -0.0447756 | 0.09740311 | 5.36704599 | 4.66200188 | FALSE | TRUE | Discordant |
| liver | VLDL_FC_by_CE | 0.00369812 | -0.053579 | 0.11562018 | 4.79114544 | 4.66200188 | FALSE | TRUE | Discordant |
| liver | LDL_FC_by_CE | -0.0005944 | 0.06320996 | 0.01736784 | 6.64098353 | 4.66200188 | FALSE | TRUE | Discordant |
| liver | Total_FC_pct | -0.0030699 | 0.0538765 | 0.09990292 | 5.07033502 | 4.66200188 | FALSE | TRUE | Discordant |

### References


1. Doshi, J. *et al.* MUSE: MUlti-atlas region Segmentation utilizing Ensembles of registration algorithms and parameters, and locally optimal atlas selection. *Neuroimage* **127**, 186–195 (2016).
2. Bai, W. *et al.* A population-based phenome-wide association study of cardiac and aortic structure and function. *Nat Med* **26**, 1654–1662 (2020).
3. Horvath, S. DNA methylation age of human tissues and cell types. *Genome Biology* **14**, 3156 (2013).
4. Moguilner, S. *et al.* Brain clocks capture diversity and disparities in aging and dementia across geographically diverse populations. *Nat Med* **30**, 3646–3657 (2024).
5. Argentieri, M. A. *et al.* Proteomic aging clock predicts mortality and risk of common age-related diseases in diverse populations. *Nat Med* **30**, 2450–2460 (2024).
6. Eldjarn, G. H. *et al.* Large-scale plasma proteomics comparisons through genetics and disease associations. *Nature* 1–11 (2023) doi:10.1038/s41586-023-06563-x.
7. Wen, J. Refining the generation, interpretation, and application of multi-organ, multi-omics biological aging clocks. *Nature Aging (in press)* 2025.02.06.25321803 (2025) doi:10.1101/2025.02.06.25321803.
8. Wishart, D. S. *et al.* HMDB 5.0: the Human Metabolome Database for 2022. *Nucleic Acids Research* **50**, D622–D631 (2022).
9. Pang, Z. *et al.* MetaboAnalystR 4.0: a unified LC-MS workflow for global metabolomics. *Nat Commun* **15**, 3675 (2024).
10. Regitz-Zagrosek, V. & Gebhard, C. Gender medicine: effects of sex and gender on cardiovascular disease manifestation and outcomes. *Nat Rev Cardiol* **20**, 236–247 (2023).

11. Bulik-Sullivan, B. K. *et al.* LD Score regression distinguishes confounding from polygenicity in genome-wide association studies. *Nat Genet* **47**, 291–295 (2015).

12. Wen, J. *et al.* The genetic architecture of multimodal human brain age. *Nat Commun* **15**, 2604 (2024).

13. Wen, J. *et al.* The genetic architecture of biological age in nine human organ systems. *Nat Aging* 1–18 (2024) doi:10.1038/s43587-024-00662-8.

14. Oh, H. S.-H. *et al.* Plasma proteomics in the UK Biobank reveals youthful brains and immune systems promote healthspan and longevity. 2024.06.07.597771 Preprint at https://doi.org/10.1101/2024.06.07.597771 (2024).

15. Anagnostakis, F. *et al.* Multi-organ metabolome biological age implicates cardiometabolic conditions and mortality risk. *Nat Commun* **16**, 4871 (2025).

16. Ying, K. *et al.* Causality-enriched epigenetic age uncouples damage and adaptation. *Nat Aging* **4**, 231–246 (2024).

17. Ferrucci, L., Barzilai, N., Belsky, D. W. & Gladyshev, V. N. How to measure biological aging in humans. *Nat Med* 1–1 (2025) doi:10.1038/s41591-025-03550-9.

18. Lee, J. *et al.* Deep learning-based brain age prediction in normal aging and dementia. *Nat Aging* **2**, 412–424 (2022).

19. Le Goallec, A. *et al.* Using deep learning to predict abdominal age from liver and pancreas magnetic resonance images. *Nat Commun* **13**, 1979 (2022).

20. Bashyam, V. M. *et al.* MRI signatures of brain age and disease over the lifespan based on a deep brain network and 14 468 individuals worldwide. *Brain* **143**, 2312–2324 (2020).

21. Chen, T., Kornblith, S., Norouzi, M. & Hinton, G. A Simple Framework for Contrastive Learning of Visual Representations. in *Proceedings of the 37th International Conference on Machine Learning* 1597–1607 (PMLR, 2020).

22. He, K., Fan, H., Wu, Y., Xie, S. & Girshick, R. Momentum Contrast for Unsupervised Visual Representation Learning. Preprint at https://doi.org/10.48550/arXiv.1911.05722 (2020).

23. Grill, J.-B. *et al.* Bootstrap your own latent: A new approach to self-supervised Learning. Preprint at https://doi.org/10.48550/arXiv.2006.07733 (2020).

24. Beheshti, I., Nugent, S., Potvin, O. & Duchesne, S. Bias-adjustment in neuroimaging-based brain age frameworks: A robust scheme. *NeuroImage: Clinical* **24**, 102063 (2019).

25. Pomponio, R. *et al.* Harmonization of large MRI datasets for the analysis of brain imaging patterns throughout the lifespan. *Neuroimage* **208**, 116450 (2020).

26. Kendler, K. & Neale, M. Endophenotype: a conceptual analysis. *Mol Psychiatry* **15**, 789–797 (2010).

27. Cannon, T. D. & Keller, M. C. Endophenotypes in the Genetic Analyses of Mental Disorders. *Annual Review of Clinical Psychology* **2**, 267–290 (2006).

28. Gottesman, I. I. & Gould, T. D. The endophenotype concept in psychiatry: etymology and strategic intentions. *Am J Psychiatry* **160**, 636–645 (2003).
